## Supplementary Tables for "Type 2 diabetes and fracture risk: deciphering the complex relationship with both genetic and observational evidence"

**Supplementary Table 1.** Characteristics of the 404 single nucleotide polymorphisms associated with type 2 diabetes.

**Supplementary Table 2.** Multivariable MR analysis of the direct effect of BMI on fracture risk and BMD.

**Supplementary Table 3.** The information of 298 lead SNPs for type 2 diabetes with the risk of fracture.

**Supplementary Table 4.** The information of 389 SNPs for type 2 diabetes with BMD.

**Supplementary Table 5.** Summary of pariwise genetic correlation using linkage disequilibrium score regeression(LDSC). Abbreviations: p1 = trait 1, p2 = trait 2, rg = genetic correlation, se = standard error of rg, p = p-value for rg, gcov_int, gcov_int_se = cross-trait LD Score regression intercept and standard error.

**Supplementary Table 6.** The results of cross-trait analysis with MiXeR model for type 2 diabetes, fracture and BMD. Abbreviations: nc1@p9, nc2@p9, and nc12@p9 = the number of causal variants for trait1, trait2 and both, respectively; nc@p9 = number of trait-influencing variants specific to type 2 diabetes; rho_beta = the correlation of effect sizes within the shared polygenic component; rg = genetic correlation; concordant_fraction = the proportion of shared variants with concordant direction of effect in both traits on all shared variants; pi1@p9, pi2@p9, and pi12@p9 = polygenicity of trait1, trait2 and both, respectively; The best_vs_min_AIC and the best_vs_max_AIC indicates whether MiXeR can accurately distinguish the reported overlap from the minimum and maximum possible overlap allowed, respectively.

**Supplementary Table 7.** Distinct genomic loci shared between type 2 diabetes and fracture at conjFDR<0.05.

**Supplementary Table 8.** The results of SMR analysis for the expression of RSPO3 (ENSG00000146374) with type 2 diabetes, fracture, BMI, WC, WHR and VAT in adipose subcutaneous.

**Supplementary Table 9.** Distinct genomic loci shared between type 2 diabetes and BMD at conjFDR<0.05.

**Supplementary Table 10.** The characteristics of participants and comparison between individuals with and without type 2 diabetes.

**Supplementary Table 11.** The regression between type 2 diabetes, fracture and BMD.

**Supplementary Table 12.** Assessment of the mediators (BMI) for the association between type 2 diabetes and fracture.

**Supplementary Table 13.** Baseline characteristics of participants with different Numbers of Risk Factors.

**Supplementary Table 14.** Detailed information on the field ID and codes for participants included in UK Biobank. Abbreviations: ICD-9, the International Classification of Diseases, 9th Revision; ICD-10, the International Classification of Diseases, 10th Revision.

**Supplementary Table 15.** The International Classification of Diseases (ICD) codes and self-reported codes for excluded diseases.

**Supplementary Table 16.** Detailed information on the field ID and codes for specific fracture sites included in UK Biobank.

**Supplementary Table 17.** Detailed information for instrumental variables of T2D and BMI on fracture and BMD.

**Supplementary Table 1.** Characteristics of the 404 single nucleotide polymorphisms associated with type 2 diabetes.

| chr (b37) | position(b37) | | chrposID | rsID | Effect  allele | Other  allele | | Effect allele  frequency | Effects  beta | Effects  SE | P-value |
| --- | --- | --- | --- | --- | --- | --- | --- | --- | --- | --- | --- |
| 1 | 40035928 | chr1:40035928 | | rs3768321 | t | | g | 0.1993 | 0.084 | 0.008 | 4.753E-26 |
| 1 | 51256091 | chr1:51256091 | | rs58432198 | t | | c | 0.1172 | -0.0636 | 0.0103 | 5.707E-10 |
| 1 | 62579891 | chr1:62579891 | | rs12140153 | t | | g | 0.0953 | -0.0645 | 0.0113 | 1.17E-08 |
| 1 | 117532790 | chr1:117532790 | | rs1127215 | t | | c | 0.4153 | -0.0491 | 0.0065 | 3.918E-14 |
| 1 | 118143517 | chr1:118143517 | | rs320369 | a | | g | 0.3232 | 0.0372 | 0.0068 | 4.602E-08 |
| 1 | 120526982 | chr1:120526982 | | rs1493694 | t | | c | 0.1096 | 0.08 | 0.0102 | 3.353E-15 |
| 1 | 177820861 | chr1:177820861 | | rs1359939 | a | | g | 0.3069 | -0.0399 | 0.0069 | 7.744E-09 |
| 1 | 177873841 | chr1:177873841 | | rs490689 | a | | g | 0.2002 | 0.0543 | 0.008 | 8.863E-12 |
| 1 | 177931532 | chr1:177931532 | | rs4650985 | a | | c | 0.2894 | -0.0383 | 0.007 | 4.77E-08 |
| 1 | 205099959 | chr1:205099959 | | rs3862948 | a | | c | 0.235 | 0.0414 | 0.0075 | 3.98E-08 |
| 1 | 206593900 | chr1:206593900 | | rs9430095 | c | | g | 0.4934 | 0.0356 | 0.0065 | 4.157E-08 |
| 1 | 214150821 | chr1:214150821 | | rs79687284 | c | | g | 0.0343 | 0.1875 | 0.0186 | 8.248E-24 |
| 1 | 214157546 | chr1:214157546 | | rs3738430 | a | | g | 0.0248 | -0.1177 | 0.0211 | 2.621E-08 |
| 1 | 214159256 | chr1:214159256 | | rs340874 | t | | c | 0.449 | -0.0678 | 0.0065 | 1.555E-25 |
| 1 | 219748818 | chr1:219748818 | | rs2820446 | c | | g | 0.7076 | 0.0593 | 0.007 | 2.833E-17 |
| 1 | 219754739 | chr1:219754739 | | rs2785962 | a | | g | 0.1909 | 0.0456 | 0.0081 | 1.548E-08 |
| 1 | 229672955 | chr1:229672955 | | rs348330 | a | | g | 0.6399 | -0.0492 | 0.0067 | 2.1E-13 |
| 2 | 422144 | chr2:422144 | | rs62107261 | t | | c | 0.9541 | 0.1019 | 0.0161 | 2.621E-10 |
| 2 | 653195 | chr2:653195 | | rs13396935 | a | | g | 0.1714 | -0.0513 | 0.0085 | 1.458E-09 |
| 2 | 16574669 | chr2:16574669 | | rs11680058 | a | | g | 0.8648 | 0.0581 | 0.0104 | 2.081E-08 |
| 2 | 25535543 | chr2:25535543 | | rs34048824 | t | | c | 0.5692 | 0.0364 | 0.0065 | 2.055E-08 |
| 2 | 25655391 | chr2:25655391 | | rs11126052 | c | | g | 0.734 | 0.0401 | 0.0072 | 2.847E-08 |
| 2 | 27730940 | chr2:27730940 | | rs1260326 | t | | c | 0.3931 | -0.0644 | 0.0066 | 1.621E-22 |
| 2 | 43429058 | chr2:43429058 | | rs10171620 | t | | g | 0.4643 | 0.0399 | 0.0065 | 7.923E-10 |
| 2 | 43687879 | chr2:43687879 | | rs17030845 | t | | c | 0.1007 | -0.1191 | 0.0108 | 2.349E-28 |
| 2 | 43994026 | chr2:43994026 | | rs142564661 | t | | c | 0.0153 | -0.1632 | 0.0288 | 1.445E-08 |
| 2 | 58990485 | chr2:58990485 | | rs2862874 | t | | g | 0.6061 | 0.036 | 0.0066 | 4.825E-08 |
| 2 | 59307725 | chr2:59307725 | | rs6545714 | a | | g | 0.6081 | -0.0363 | 0.0065 | 2.246E-08 |
| 2 | 60585806 | chr2:60585806 | | rs243019 | t | | c | 0.5359 | -0.0588 | 0.0064 | 3.372E-20 |
| 2 | 65289825 | chr2:65289825 | | rs2540945 | a | | g | 0.6333 | 0.0539 | 0.0066 | 3.051E-16 |
| 2 | 65577885 | chr2:65577885 | | rs61330787 | c | | g | 0.9724 | 0.1129 | 0.0202 | 2.31E-08 |
| 2 | 65655012 | chr2:65655012 | | rs2028150 | c | | g | 0.5982 | 0.0537 | 0.0065 | 1.314E-16 |
| 2 | 121347612 | chr2:121347612 | | rs11688682 | c | | g | 0.2721 | -0.0581 | 0.0076 | 2.928E-14 |
| 2 | 158329237 | chr2:158329237 | | rs16841827 | a | | g | 0.063 | -0.0822 | 0.0133 | 6.339E-10 |
| 2 | 161136656 | chr2:161136656 | | rs7572970 | a | | g | 0.2659 | -0.0447 | 0.0072 | 6.12E-10 |
| 2 | 165528876 | chr2:165528876 | | rs13389219 | t | | c | 0.4014 | -0.0605 | 0.0065 | 1.165E-20 |
| 2 | 165572081 | chr2:165572081 | | rs1869525 | t | | c | 0.5631 | 0.0403 | 0.0065 | 5.364E-10 |
| 2 | 226952104 | chr2:226952104 | | rs4675045 | c | | g | 0.7922 | -0.0543 | 0.0081 | 1.637E-11 |
| 2 | 227019416 | chr2:227019416 | | rs72979712 | t | | g | 0.0525 | -0.0928 | 0.0148 | 3.258E-10 |
| 2 | 227101411 | chr2:227101411 | | rs2972144 | a | | g | 0.3618 | -0.0911 | 0.0066 | 2.192E-43 |
| 2 | 227131174 | chr2:227131174 | | rs41504645 | t | | c | 0.8771 | -0.0689 | 0.0097 | 1.485E-12 |
| 2 | 227228251 | chr2:227228251 | | rs76367336 | t | | c | 0.8863 | 0.0584 | 0.0103 | 1.259E-08 |
| 3 | 12310773 | chr3:12310773 | | rs145268310 | c | | g | 0.1171 | 0.0606 | 0.0098 | 7.401E-10 |
| 3 | 12336507 | chr3:12336507 | | rs11709077 | a | | g | 0.1264 | -0.104 | 0.0097 | 1.255E-26 |
| 3 | 12377344 | chr3:12377344 | | rs4518111 | a | | c | 0.4338 | 0.0468 | 0.0067 | 2.867E-12 |
| 3 | 23454790 | chr3:23454790 | | rs1496653 | a | | g | 0.7887 | 0.0665 | 0.0079 | 2.487E-17 |
| 3 | 23618935 | chr3:23618935 | | rs74486672 | t | | c | 0.0357 | 0.1156 | 0.0175 | 3.81E-11 |
| 3 | 49980596 | chr3:49980596 | | rs4688760 | t | | c | 0.6828 | 0.042 | 0.007 | 2.136E-09 |
| 3 | 53123273 | chr3:53123273 | | rs891368 | a | | g | 0.4335 | -0.0378 | 0.0064 | 3.251E-09 |
| 3 | 54828827 | chr3:54828827 | | rs76263492 | t | | g | 0.0452 | 0.0897 | 0.0156 | 8.937E-09 |
| 3 | 63974258 | chr3:63974258 | | rs3774725 | t | | g | 0.8489 | 0.0662 | 0.009 | 1.953E-13 |
| 3 | 64701387 | chr3:64701387 | | rs4368494 | a | | g | 0.2976 | -0.054 | 0.007 | 1.385E-14 |
| 3 | 123065778 | chr3:123065778 | | rs11708067 | a | | g | 0.775 | 0.0882 | 0.0077 | 5.046E-30 |
| 3 | 124925934 | chr3:124925934 | | rs569255 | a | | g | 0.5393 | -0.0396 | 0.0064 | 5.638E-10 |
| 3 | 150066540 | chr3:150066540 | | rs62271373 | a | | t | 0.0547 | 0.0882 | 0.0144 | 1.033E-09 |
| 3 | 152392877 | chr3:152392877 | | rs9828639 | t | | g | 0.3266 | 0.0383 | 0.0068 | 1.827E-08 |
| 3 | 152456697 | chr3:152456697 | | rs78569745 | t | | g | 0.9578 | 0.0988 | 0.0165 | 2.339E-09 |
| 3 | 168223132 | chr3:168223132 | | rs7642311 | a | | g | 0.8723 | 0.0545 | 0.0096 | 1.533E-08 |
| 3 | 170724883 | chr3:170724883 | | rs8192675 | t | | c | 0.7121 | 0.0659 | 0.007 | 5.772E-21 |
| 3 | 185256242 | chr3:185256242 | | rs62290256 | a | | g | 0.9364 | -0.0768 | 0.0134 | 1.001E-08 |
| 3 | 185354394 | chr3:185354394 | | rs7616221 | t | | g | 0.0351 | 0.1017 | 0.0175 | 6.016E-09 |
| 3 | 185488509 | chr3:185488509 | | rs75974105 | c | | g | 0.0344 | 0.1005 | 0.0177 | 1.35E-08 |
| 3 | 185510613 | chr3:185510613 | | rs7633675 | t | | g | 0.6837 | -0.1085 | 0.0068 | 3.206E-57 |
| 3 | 185540817 | chr3:185540817 | | rs113672528 | a | | c | 0.9753 | -0.128 | 0.0209 | 9.804E-10 |
| 3 | 185548663 | chr3:185548663 | | rs2194411 | a | | g | 0.1291 | -0.0746 | 0.0097 | 1.841E-14 |
| 3 | 186665645 | chr3:186665645 | | rs3887925 | t | | c | 0.545 | 0.0539 | 0.0065 | 1.014E-16 |
| 3 | 187740523 | chr3:187740523 | | rs6808574 | t | | c | 0.3885 | -0.061 | 0.0066 | 2.294E-20 |
| 4 | 738275 | chr4:738275 | | rs56187241 | t | | c | 0.0451 | 0.1014 | 0.0157 | 1.072E-10 |
| 4 | 1784403 | chr4:1784403 | | rs56337234 | t | | c | 0.4978 | -0.0548 | 0.0066 | 9.753E-17 |
| 4 | 3241845 | chr4:3241845 | | rs362307 | t | | c | 0.0776 | 0.0704 | 0.0121 | 6.776E-09 |
| 4 | 6257188 | chr4:6257188 | | rs4689381 | t | | c | 0.4437 | 0.038 | 0.0066 | 8.369E-09 |
| 4 | 6306763 | chr4:6306763 | | rs10937721 | c | | g | 0.5893 | 0.085 | 0.0066 | 5.39E-38 |
| 4 | 17811914 | chr4:17811914 | | rs7667864 | a | | c | 0.2838 | -0.0402 | 0.0071 | 1.64E-08 |
| 4 | 45175691 | chr4:45175691 | | rs13130484 | t | | c | 0.4296 | 0.0435 | 0.0067 | 8.492E-11 |
| 4 | 83578271 | chr4:83578271 | | rs79920718 | a | | g | 0.3385 | 0.0389 | 0.0067 | 6.431E-09 |
| 4 | 89740894 | chr4:89740894 | | rs1903002 | c | | g | 0.4944 | -0.0362 | 0.0064 | 1.446E-08 |
| 4 | 95091911 | chr4:95091911 | | rs6821438 | a | | g | 0.5342 | 0.0397 | 0.0064 | 5.103E-10 |
| 4 | 103900090 | chr4:103900090 | | rs10516495 | a | | t | 0.3128 | -0.0418 | 0.0069 | 1.458E-09 |
| 4 | 153513369 | chr4:153513369 | | rs7669833 | a | | t | 0.2951 | -0.0572 | 0.007 | 3.524E-16 |
| 4 | 157652753 | chr4:157652753 | | rs28819812 | a | | c | 0.3218 | -0.0396 | 0.0072 | 4.22E-08 |
| 4 | 185718132 | chr4:185718132 | | rs745805 | a | | t | 0.82 | 0.0632 | 0.0084 | 4.516E-14 |
| 5 | 14753745 | chr5:14753745 | | rs17250977 | a | | g | 0.9628 | -0.1178 | 0.0178 | 3.636E-11 |
| 5 | 14755919 | chr5:14755919 | | rs147581833 | t | | c | 0.0062 | -0.3713 | 0.0489 | 3.11E-14 |
| 5 | 14768092 | chr5:14768092 | | rs6885132 | c | | g | 0.9036 | 0.077 | 0.011 | 2.488E-12 |
| 5 | 14784165 | chr5:14784165 | | rs143306347 | a | | c | 0.0124 | -0.1747 | 0.0314 | 2.67E-08 |
| 5 | 14794868 | chr5:14794868 | | rs879253 | t | | c | 0.5646 | -0.0372 | 0.0065 | 1.001E-08 |
| 5 | 44682589 | chr5:44682589 | | rs6884702 | a | | g | 0.6055 | -0.0399 | 0.0065 | 7.923E-10 |
| 5 | 51791225 | chr5:51791225 | | rs17261179 | t | | c | 0.5154 | 0.0355 | 0.0064 | 2.724E-08 |
| 5 | 52100489 | chr5:52100489 | | rs3811978 | a | | g | 0.834 | -0.0492 | 0.0085 | 6.581E-09 |
| 5 | 53271420 | chr5:53271420 | | rs702634 | a | | g | 0.689 | 0.0503 | 0.0069 | 3.365E-13 |
| 5 | 53321956 | chr5:53321956 | | rs255761 | t | | g | 0.202 | 0.0541 | 0.008 | 1.056E-11 |
| 5 | 53463117 | chr5:53463117 | | rs154021 | t | | c | 0.7537 | 0.0417 | 0.0074 | 2.03E-08 |
| 5 | 55806751 | chr5:55806751 | | rs459193 | a | | g | 0.2594 | -0.0722 | 0.0073 | 6.783E-23 |
| 5 | 55861894 | chr5:55861894 | | rs9687846 | a | | g | 0.1902 | 0.0701 | 0.0081 | 3.468E-18 |
| 5 | 75003678 | chr5:75003678 | | rs2307111 | t | | c | 0.6039 | 0.0502 | 0.0065 | 1.049E-14 |
| 5 | 76435004 | chr5:76435004 | | rs7732130 | a | | g | 0.6992 | -0.0606 | 0.007 | 5.693E-18 |
| 5 | 78430607 | chr5:78430607 | | rs1316776 | a | | c | 0.3517 | -0.0433 | 0.0067 | 1.035E-10 |
| 5 | 101250990 | chr5:101250990 | | rs77372998 | a | | g | 0.9917 | -0.2834 | 0.0411 | 5.685E-12 |
| 5 | 101273694 | chr5:101273694 | | rs145510090 | a | | t | 0.053 | 0.0976 | 0.0147 | 2.77E-11 |
| 5 | 101631592 | chr5:101631592 | | rs3114659 | a | | g | 0.3163 | 0.0414 | 0.0069 | 2.085E-09 |
| 5 | 101870140 | chr5:101870140 | | rs145762933 | a | | g | 0.0497 | 0.1453 | 0.0151 | 5.574E-22 |
| 5 | 102338739 | chr5:102338739 | | rs78408340 | c | | g | 0.9918 | -0.3788 | 0.0389 | 2.348E-22 |
| 5 | 102360747 | chr5:102360747 | | rs17296280 | a | | c | 0.6731 | -0.0456 | 0.007 | 8.016E-11 |
| 5 | 102422968 | chr5:102422968 | | rs115505614 | t | | c | 0.0502 | 0.1657 | 0.0149 | 7.58E-29 |
| 5 | 102704259 | chr5:102704259 | | rs10068434 | t | | c | 0.7236 | -0.0393 | 0.0071 | 3.393E-08 |
| 5 | 102973337 | chr5:102973337 | | rs116407196 | a | | g | 0.9591 | -0.0979 | 0.0169 | 6.336E-09 |
| 5 | 133864599 | chr5:133864599 | | rs329122 | a | | g | 0.4292 | 0.0366 | 0.0065 | 1.719E-08 |
| 6 | 7203714 | chr6:7203714 | | rs6921580 | c | | g | 0.4158 | 0.0487 | 0.0067 | 3.659E-13 |
| 6 | 7231843 | chr6:7231843 | | rs9379084 | a | | g | 0.1121 | -0.0994 | 0.0106 | 5.478E-21 |
| 6 | 7245458 | chr6:7245458 | | rs1815311 | a | | g | 0.652 | -0.0511 | 0.0067 | 2.426E-14 |
| 6 | 7275941 | chr6:7275941 | | rs11243150 | t | | c | 0.6015 | 0.0512 | 0.0065 | 3.091E-15 |
| 6 | 20518450 | chr6:20518450 | | rs6903706 | a | | g | 0.181 | -0.062 | 0.0084 | 1.342E-13 |
| 6 | 20541166 | chr6:20541166 | | rs58761454 | t | | c | 0.2537 | 0.05 | 0.0073 | 8.975E-12 |
| 6 | 20553878 | chr6:20553878 | | rs76393458 | a | | g | 0.018 | 0.1507 | 0.0249 | 1.471E-09 |
| 6 | 20633973 | chr6:20633973 | | rs78421627 | a | | t | 0.9731 | -0.1251 | 0.0203 | 7.328E-10 |
| 6 | 20683900 | chr6:20683900 | | rs72830693 | c | | g | 0.9648 | -0.1421 | 0.0176 | 6.559E-16 |
| 6 | 20686996 | chr6:20686996 | | rs9368222 | a | | c | 0.2717 | 0.1379 | 0.0071 | 1.42E-83 |
| 6 | 20725940 | chr6:20725940 | | rs6915155 | t | | g | 0.5574 | -0.0493 | 0.0064 | 1.173E-14 |
| 6 | 20894610 | chr6:20894610 | | rs77786848 | a | | g | 0.9474 | -0.1102 | 0.0148 | 8.356E-14 |
| 6 | 20931156 | chr6:20931156 | | rs7775995 | a | | g | 0.6891 | 0.0487 | 0.0069 | 1.823E-12 |
| 6 | 30993958 | chr6:30993958 | | rs2523897 | a | | g | 0.1632 | -0.0487 | 0.0088 | 3.073E-08 |
| 6 | 31115441 | chr6:31115441 | | rs3131012 | t | | c | 0.4605 | -0.0471 | 0.0064 | 1.649E-13 |
| 6 | 31209288 | chr6:31209288 | | rs3095248 | t | | c | 0.4153 | -0.0376 | 0.0065 | 6.949E-09 |
| 6 | 31249267 | chr6:31249267 | | rs35840219 | c | | g | 0.0957 | 0.0628 | 0.0112 | 2.076E-08 |
| 6 | 31280079 | chr6:31280079 | | rs9264981 | t | | c | 0.5725 | 0.0487 | 0.0068 | 8.318E-13 |
| 6 | 31440014 | chr6:31440014 | | rs14597 | a | | c | 0.4136 | 0.0359 | 0.0065 | 3.197E-08 |
| 6 | 31555657 | chr6:31555657 | | rs3135041 | a | | g | 0.6768 | -0.0405 | 0.0068 | 2.666E-09 |
| 6 | 31577825 | chr6:31577825 | | rs2857609 | a | | g | 0.1631 | -0.0705 | 0.0088 | 1.093E-15 |
| 6 | 31864538 | chr6:31864538 | | rs115884658 | a | | g | 0.0288 | 0.126 | 0.0207 | 1.218E-09 |
| 6 | 32080146 | chr6:32080146 | | rs3130342 | a | | c | 0.1528 | -0.0709 | 0.009 | 3.437E-15 |
| 6 | 32190390 | chr6:32190390 | | rs915894 | t | | g | 0.6351 | -0.0406 | 0.0067 | 1.371E-09 |
| 6 | 32373378 | chr6:32373378 | | rs3806155 | a | | t | 0.9643 | -0.1691 | 0.0183 | 2.735E-20 |
| 6 | 32573415 | chr6:32573415 | | rs601945 | a | | g | 0.8259 | -0.0849 | 0.0086 | 4.662E-23 |
| 6 | 32612397 | chr6:32612397 | | rs6927022 | a | | g | 0.5427 | -0.0585 | 0.0069 | 2.549E-17 |
| 6 | 32632832 | chr6:32632832 | | rs9274407 | a | | t | 0.1956 | -0.073 | 0.0087 | 4.458E-17 |
| 6 | 32814447 | chr6:32814447 | | rs6457684 | t | | c | 0.5743 | -0.0369 | 0.0065 | 1.313E-08 |
| 6 | 40409243 | chr6:40409243 | | rs34298980 | t | | c | 0.4994 | 0.0379 | 0.0067 | 1.55E-08 |
| 6 | 43760327 | chr6:43760327 | | rs11967262 | c | | g | 0.5142 | -0.0378 | 0.0065 | 5.782E-09 |
| 6 | 43815364 | chr6:43815364 | | rs6937438 | a | | g | 0.7112 | -0.0514 | 0.007 | 2.352E-13 |
| 6 | 50788778 | chr6:50788778 | | rs3798519 | a | | c | 0.811 | -0.0616 | 0.0082 | 4.602E-14 |
| 6 | 107433400 | chr6:107433400 | | rs1665901 | a | | t | 0.6576 | 0.0398 | 0.0068 | 4.973E-09 |
| 6 | 126792095 | chr6:126792095 | | rs11759026 | a | | g | 0.768 | -0.066 | 0.0075 | 2.042E-18 |
| 6 | 127409882 | chr6:127409882 | | rs4580892 | t | | c | 0.2933 | 0.0404 | 0.007 | 8.459E-09 |
| 6 | 127414838 | chr6:127414838 | | rs719727 | a | | g | 0.7582 | 0.0592 | 0.0074 | 1.672E-15 |
| 6 | 137302159 | chr6:137302159 | | rs1573090 | t | | g | 0.5328 | 0.0458 | 0.0065 | 1.722E-12 |
| 6 | 153433701 | chr6:153433701 | | rs6557267 | t | | c | 0.4201 | 0.036 | 0.0065 | 2.928E-08 |
| 6 | 160770312 | chr6:160770312 | | rs474513 | a | | g | 0.5162 | 0.0399 | 0.0064 | 4.178E-10 |
| 6 | 164133001 | chr6:164133001 | | rs4709746 | t | | c | 0.1313 | -0.0567 | 0.0096 | 3.951E-09 |
| 7 | 14898282 | chr7:14898282 | | rs17168486 | t | | c | 0.1802 | 0.0672 | 0.0083 | 4.5E-16 |
| 7 | 15062983 | chr7:15062983 | | rs2215383 | t | | c | 0.4625 | -0.0641 | 0.0064 | 1.056E-23 |
| 7 | 27977174 | chr7:27977174 | | rs62451130 | t | | c | 0.0918 | -0.0654 | 0.0112 | 5.295E-09 |
| 7 | 28191672 | chr7:28191672 | | rs10225433 | a | | g | 0.0668 | -0.0767 | 0.0138 | 2.863E-08 |
| 7 | 28198677 | chr7:28198677 | | rs1708302 | t | | c | 0.4885 | -0.0909 | 0.0064 | 5.769E-46 |
| 7 | 28227024 | chr7:28227024 | | rs73300665 | a | | g | 0.1066 | 0.0587 | 0.0103 | 1.06E-08 |
| 7 | 30728452 | chr7:30728452 | | rs917195 | t | | c | 0.2287 | -0.0473 | 0.0077 | 1.029E-09 |
| 7 | 44189321 | chr7:44189321 | | rs2268574 | a | | g | 0.5095 | -0.0383 | 0.0064 | 2.014E-09 |
| 7 | 44255643 | chr7:44255643 | | rs878521 | a | | g | 0.2434 | 0.0618 | 0.0074 | 9.308E-17 |
| 7 | 102086552 | chr7:102086552 | | rs77655131 | t | | c | 0.1301 | 0.0554 | 0.0097 | 1.274E-08 |
| 7 | 102486254 | chr7:102486254 | | rs11496066 | t | | c | 0.8173 | 0.0508 | 0.0083 | 8.171E-10 |
| 7 | 130457914 | chr7:130457914 | | rs1562396 | a | | g | 0.6796 | -0.0555 | 0.0069 | 9.635E-16 |
| 7 | 150537635 | chr7:150537635 | | rs62492368 | a | | g | 0.3123 | 0.044 | 0.0069 | 1.924E-10 |
| 7 | 156975136 | chr7:156975136 | | rs6459737 | a | | g | 0.338 | -0.0583 | 0.0067 | 3.311E-18 |
| 8 | 8304502 | chr8:8304502 | | rs2921077 | a | | g | 0.4619 | 0.0373 | 0.0065 | 9.142E-09 |
| 8 | 9188762 | chr8:9188762 | | rs11774915 | t | | c | 0.3331 | 0.0436 | 0.0068 | 1.49E-10 |
| 8 | 9265105 | chr8:9265105 | | rs17662402 | t | | c | 0.9435 | 0.0835 | 0.0142 | 4.52E-09 |
| 8 | 9996389 | chr8:9996389 | | rs34990153 | a | | g | 0.5601 | 0.0476 | 0.0065 | 2.257E-13 |
| 8 | 10009087 | chr8:10009087 | | rs56408109 | t | | g | 0.0514 | -0.0864 | 0.0157 | 3.77E-08 |
| 8 | 10903475 | chr8:10903475 | | rs2001433 | a | | t | 0.526 | -0.0442 | 0.0065 | 9.838E-12 |
| 8 | 11450422 | chr8:11450422 | | rs2244648 | a | | g | 0.5372 | -0.0363 | 0.0065 | 2.246E-08 |
| 8 | 12618225 | chr8:12618225 | | rs12680692 | a | | t | 0.3206 | 0.0413 | 0.0071 | 6.602E-09 |
| 8 | 19844415 | chr8:19844415 | | rs7819706 | a | | g | 0.8835 | 0.0669 | 0.0101 | 2.818E-11 |
| 8 | 41435225 | chr8:41435225 | | rs1060731 | t | | c | 0.2915 | 0.0414 | 0.007 | 3.6E-09 |
| 8 | 41500861 | chr8:41500861 | | rs59191643 | t | | c | 0.628 | 0.0496 | 0.0066 | 5.502E-14 |
| 8 | 41522991 | chr8:41522991 | | rs508419 | a | | g | 0.2356 | -0.0811 | 0.0075 | 5.433E-27 |
| 8 | 41529789 | chr8:41529789 | | rs80105613 | t | | c | 0.0366 | 0.1074 | 0.0174 | 6.438E-10 |
| 8 | 95739642 | chr8:95739642 | | rs67763258 | t | | g | 0.3228 | -0.0387 | 0.0068 | 1.297E-08 |
| 8 | 95961626 | chr8:95961626 | | rs10097617 | t | | c | 0.485 | 0.0487 | 0.0064 | 2.439E-14 |
| 8 | 96128981 | chr8:96128981 | | rs76471882 | t | | c | 0.0468 | 0.0875 | 0.0154 | 1.308E-08 |
| 8 | 116482423 | chr8:116482423 | | rs3808415 | a | | g | 0.5575 | -0.0349 | 0.0064 | 4.646E-08 |
| 8 | 118185025 | chr8:118185025 | | rs3802177 | a | | g | 0.3154 | -0.1077 | 0.0069 | 9.163E-55 |
| 8 | 118190620 | chr8:118190620 | | rs16889471 | a | | g | 0.2135 | 0.0448 | 0.0077 | 7.375E-09 |
| 8 | 128711742 | chr8:128711742 | | rs17772814 | a | | g | 0.0857 | -0.0746 | 0.0125 | 2.133E-09 |
| 8 | 129568078 | chr8:129568078 | | rs1561927 | t | | c | 0.7328 | -0.042 | 0.0072 | 6.115E-09 |
| 8 | 145507304 | chr8:145507304 | | rs4977213 | t | | c | 0.6281 | -0.0507 | 0.0069 | 2.187E-13 |
| 8 | 145639726 | chr8:145639726 | | rs2272662 | t | | c | 0.4123 | -0.0402 | 0.0072 | 2.63E-08 |
| 8 | 145879883 | chr8:145879883 | | rs12719778 | t | | c | 0.5365 | 0.0381 | 0.0065 | 4.38E-09 |
| 9 | 3273781 | chr9:3273781 | | rs672271 | t | | c | 0.9062 | -0.06 | 0.011 | 4.825E-08 |
| 9 | 4291928 | chr9:4291928 | | rs10974438 | a | | c | 0.6416 | -0.0514 | 0.0067 | 1.713E-14 |
| 9 | 19065825 | chr9:19065825 | | rs62563593 | a | | g | 0.6063 | -0.0394 | 0.0065 | 1.284E-09 |
| 9 | 21954653 | chr9:21954653 | | rs10965199 | t | | c | 0.0361 | -0.1139 | 0.019 | 1.853E-09 |
| 9 | 22003367 | chr9:22003367 | | rs1063192 | a | | g | 0.5617 | 0.0575 | 0.0065 | 8.17E-19 |
| 9 | 22051295 | chr9:22051295 | | rs17694555 | a | | g | 0.9076 | -0.0614 | 0.011 | 2.337E-08 |
| 9 | 22133773 | chr9:22133773 | | rs76011118 | a | | g | 0.0335 | 0.1871 | 0.0195 | 7.443E-22 |
| 9 | 22134068 | chr9:22134068 | | rs10811660 | a | | g | 0.1704 | -0.1598 | 0.0086 | 2.541E-77 |
| 9 | 22136440 | chr9:22136440 | | rs12555274 | c | | g | 0.2646 | 0.1068 | 0.0072 | 1.883E-49 |
| 9 | 22139684 | chr9:22139684 | | rs72655474 | c | | g | 0.0217 | -0.1908 | 0.025 | 2.446E-14 |
| 9 | 22140224 | chr9:22140224 | | rs2065501 | a | | c | 0.319 | 0.0443 | 0.0073 | 1.5E-09 |
| 9 | 28410683 | chr9:28410683 | | rs1412234 | t | | c | 0.6739 | -0.0393 | 0.0068 | 7.713E-09 |
| 9 | 34074476 | chr9:34074476 | | rs12001437 | t | | c | 0.6309 | -0.0402 | 0.0066 | 1.099E-09 |
| 9 | 81905590 | chr9:81905590 | | rs17791513 | a | | g | 0.9323 | 0.1016 | 0.0132 | 1.346E-14 |
| 9 | 84308948 | chr9:84308948 | | rs2796441 | a | | g | 0.406 | -0.0674 | 0.0065 | 2.972E-25 |
| 9 | 97001682 | chr9:97001682 | | rs55653563 | a | | c | 0.7333 | 0.0435 | 0.0072 | 1.731E-09 |
| 9 | 136149229 | chr9:136149229 | | rs505922 | t | | c | 0.6634 | -0.0473 | 0.0068 | 3.65E-12 |
| 9 | 139241828 | chr9:139241828 | | rs28533815 | t | | c | 0.2507 | -0.0748 | 0.008 | 5.452E-21 |
| 9 | 139335599 | chr9:139335599 | | rs1127152 | a | | g | 0.5683 | 0.0358 | 0.0065 | 3.491E-08 |
| 10 | 12307894 | chr10:12307894 | | rs11257655 | t | | c | 0.2169 | 0.0859 | 0.0077 | 1.455E-28 |
| 10 | 71317835 | chr10:71317835 | | rs2616055 | a | | g | 0.9402 | 0.0799 | 0.0137 | 5.701E-09 |
| 10 | 71321279 | chr10:71321279 | | rs177045 | a | | g | 0.6832 | -0.05 | 0.0069 | 4.638E-13 |
| 10 | 71332301 | chr10:71332301 | | rs41277236 | t | | c | 0.0426 | 0.0999 | 0.0168 | 2.471E-09 |
| 10 | 71465359 | chr10:71465359 | | rs2812539 | t | | g | 0.7021 | 0.051 | 0.007 | 3.593E-13 |
| 10 | 80907740 | chr10:80907740 | | rs11591689 | t | | c | 0.233 | 0.0486 | 0.0077 | 3.551E-10 |
| 10 | 80952826 | chr10:80952826 | | rs703972 | c | | g | 0.4676 | -0.0698 | 0.0065 | 5.771E-27 |
| 10 | 80973926 | chr10:80973926 | | rs76087804 | a | | g | 0.1736 | -0.0605 | 0.0086 | 1.832E-12 |
| 10 | 80992381 | chr10:80992381 | | rs1574190 | t | | c | 0.392 | 0.0476 | 0.0067 | 1.217E-12 |
| 10 | 93749380 | chr10:93749380 | | rs41287646 | t | | g | 0.9139 | -0.0767 | 0.0116 | 4.123E-11 |
| 10 | 94138239 | chr10:94138239 | | rs7903767 | a | | g | 0.54 | -0.055 | 0.0064 | 7.214E-18 |
| 10 | 94233120 | chr10:94233120 | | rs2249960 | a | | g | 0.8658 | -0.0583 | 0.0095 | 9.425E-10 |
| 10 | 94274809 | chr10:94274809 | | rs17875327 | a | | g | 0.8822 | -0.0592 | 0.0101 | 3.869E-09 |
| 10 | 94307157 | chr10:94307157 | | rs77014180 | a | | g | 0.9396 | -0.0863 | 0.0135 | 1.665E-10 |
| 10 | 94460650 | chr10:94460650 | | rs10882099 | t | | c | 0.5879 | 0.1095 | 0.0065 | 7.726E-64 |
| 10 | 94466064 | chr10:94466064 | | rs146935743 | t | | c | 0.0503 | -0.1046 | 0.0159 | 4.947E-11 |
| 10 | 94500111 | chr10:94500111 | | rs11187152 | a | | g | 0.0786 | -0.0695 | 0.0124 | 1.852E-08 |
| 10 | 94501355 | chr10:94501355 | | rs12357047 | a | | c | 0.361 | -0.0384 | 0.0068 | 1.677E-08 |
| 10 | 114578143 | chr10:114578143 | | rs145220772 | t | | c | 0.0213 | 0.1315 | 0.0237 | 2.74E-08 |
| 10 | 114593614 | chr10:114593614 | | rs11814823 | a | | t | 0.8075 | -0.0523 | 0.0088 | 2.738E-09 |
| 10 | 114597109 | chr10:114597109 | | rs545572 | a | | g | 0.3929 | -0.0762 | 0.0068 | 4.238E-29 |
| 10 | 114648499 | chr10:114648499 | | rs1885283 | a | | c | 0.1303 | 0.0727 | 0.0102 | 8.172E-13 |
| 10 | 114668724 | chr10:114668724 | | rs139688524 | t | | c | 0.0226 | 0.1739 | 0.0216 | 7.468E-16 |
| 10 | 114671172 | chr10:114671172 | | rs11196148 | t | | c | 0.1755 | -0.0602 | 0.0087 | 4.291E-12 |
| 10 | 114681965 | chr10:114681965 | | rs116425039 | a | | g | 0.012 | -0.272 | 0.0348 | 5.084E-15 |
| 10 | 114711883 | chr10:114711883 | | rs10885397 | a | | g | 0.1539 | -0.0736 | 0.0091 | 6.486E-16 |
| 10 | 114721404 | chr10:114721404 | | rs7895657 | a | | g | 0.328 | -0.0487 | 0.007 | 3.86E-12 |
| 10 | 114721568 | chr10:114721568 | | rs720785 | c | | g | 0.2402 | 0.1093 | 0.0077 | 3.443E-45 |
| 10 | 114725079 | chr10:114725079 | | rs61875108 | a | | g | 0.0488 | -0.093 | 0.0155 | 1.954E-09 |
| 10 | 114735110 | chr10:114735110 | | rs117736037 | a | | g | 0.0145 | 0.2514 | 0.0282 | 4.417E-19 |
| 10 | 114738287 | chr10:114738287 | | rs146262265 | t | | c | 0.0092 | 0.2255 | 0.0399 | 1.578E-08 |
| 10 | 114752410 | chr10:114752410 | | rs116859590 | t | | c | 0.0263 | 0.229 | 0.021 | 1.415E-27 |
| 10 | 114754071 | chr10:114754071 | | rs34872471 | t | | c | 0.708 | -0.313 | 0.007 | 9.06e-435 |
| 10 | 114756459 | chr10:114756459 | | rs117987174 | a | | g | 0.0135 | -0.1723 | 0.0305 | 1.558E-08 |
| 10 | 114757956 | chr10:114757956 | | rs78025551 | c | | g | 0.8468 | 0.1493 | 0.0091 | 2.247E-60 |
| 10 | 114765390 | chr10:114765390 | | rs61872774 | a | | g | 0.0124 | 0.288 | 0.0298 | 4.848E-22 |
| 10 | 114770644 | chr10:114770644 | | rs140908036 | a | | g | 0.0153 | 0.2887 | 0.0285 | 3.775E-24 |
| 10 | 114770860 | chr10:114770860 | | rs141356057 | t | | c | 0.0105 | -0.1907 | 0.0336 | 1.395E-08 |
| 10 | 114773068 | chr10:114773068 | | rs10885404 | t | | g | 0.1767 | -0.1071 | 0.0088 | 4.103E-34 |
| 10 | 114775551 | chr10:114775551 | | rs141241414 | a | | g | 0.9857 | 0.1947 | 0.0284 | 6.798E-12 |
| 10 | 114784926 | chr10:114784926 | | rs180726800 | t | | c | 0.0194 | 0.2518 | 0.0247 | 2.191E-24 |
| 10 | 114787948 | chr10:114787948 | | rs140820620 | a | | g | 0.0224 | 0.2841 | 0.0227 | 7.084E-36 |
| 10 | 114793572 | chr10:114793572 | | rs116369954 | t | | c | 0.9667 | -0.2919 | 0.0182 | 8.911E-58 |
| 10 | 114794451 | chr10:114794451 | | rs185025714 | t | | c | 0.9951 | -0.2643 | 0.0477 | 3.099E-08 |
| 10 | 114803307 | chr10:114803307 | | rs11196201 | a | | t | 0.9222 | -0.1851 | 0.012 | 2.493E-53 |
| 10 | 114815466 | chr10:114815466 | | rs72826101 | t | | g | 0.9354 | 0.1085 | 0.0143 | 3.908E-14 |
| 10 | 114824473 | chr10:114824473 | | rs10885410 | a | | g | 0.2863 | -0.0858 | 0.0074 | 8.124E-31 |
| 10 | 114830306 | chr10:114830306 | | rs4918791 | a | | g | 0.379 | -0.0729 | 0.0071 | 1.325E-24 |
| 10 | 114861304 | chr10:114861304 | | rs10885414 | a | | g | 0.6855 | 0.0949 | 0.007 | 1.068E-41 |
| 10 | 114865489 | chr10:114865489 | | rs10749128 | t | | c | 0.7481 | -0.046 | 0.0074 | 6.097E-10 |
| 10 | 114886341 | chr10:114886341 | | rs189966089 | t | | c | 0.029 | -0.1284 | 0.0221 | 6.173E-09 |
| 10 | 114887722 | chr10:114887722 | | rs11196236 | t | | c | 0.7903 | -0.0739 | 0.0079 | 4.924E-21 |
| 10 | 114915214 | chr10:114915214 | | rs290483 | t | | g | 0.6019 | 0.0701 | 0.0067 | 1.302E-25 |
| 10 | 114942869 | chr10:114942869 | | rs72828149 | a | | g | 0.018 | 0.148 | 0.0265 | 2.309E-08 |
| 10 | 124193181 | chr10:124193181 | | rs2280141 | t | | g | 0.5118 | 0.0489 | 0.0064 | 1.913E-14 |
| 11 | 2073182 | chr11:2073182 | | rs76074250 | a | | g | 0.7971 | -0.0463 | 0.0081 | 9.305E-09 |
| 11 | 2181338 | chr11:2181338 | | rs3842749 | a | | g | 0.0592 | 0.0858 | 0.0142 | 1.686E-09 |
| 11 | 2197286 | chr11:2197286 | | rs4929965 | a | | g | 0.3813 | 0.0668 | 0.0068 | 9.669E-23 |
| 11 | 2632430 | chr11:2632430 | | rs11023461 | c | | g | 0.9242 | -0.0751 | 0.0129 | 5.494E-09 |
| 11 | 2692249 | chr11:2692249 | | rs231360 | t | | c | 0.3963 | 0.0576 | 0.0067 | 8.269E-18 |
| 11 | 2752130 | chr11:2752130 | | rs231907 | a | | t | 0.6711 | -0.0389 | 0.007 | 2.936E-08 |
| 11 | 2835757 | chr11:2835757 | | rs2237890 | t | | c | 0.0659 | -0.0797 | 0.0139 | 1.044E-08 |
| 11 | 2857194 | chr11:2857194 | | rs2237895 | a | | c | 0.573 | -0.0892 | 0.0066 | 1.147E-41 |
| 11 | 2858546 | chr11:2858546 | | rs2237897 | t | | c | 0.0456 | -0.1924 | 0.0166 | 6.791E-31 |
| 11 | 14763828 | chr11:14763828 | | rs141521721 | a | | c | 0.0233 | 0.1212 | 0.0214 | 1.392E-08 |
| 11 | 17408630 | chr11:17408630 | | rs5215 | t | | c | 0.6272 | -0.0706 | 0.0066 | 9.848E-27 |
| 11 | 32927778 | chr11:32927778 | | rs145678014 | t | | g | 0.0426 | -0.1048 | 0.0163 | 1.397E-10 |
| 11 | 34782972 | chr11:34782972 | | rs10047488 | c | | g | 0.0822 | 0.0635 | 0.0116 | 4.661E-08 |
| 11 | 34982148 | chr11:34982148 | | rs2767036 | a | | c | 0.7086 | -0.0389 | 0.007 | 2.936E-08 |
| 11 | 43877934 | chr11:43877934 | | rs1061810 | a | | c | 0.2867 | 0.0502 | 0.007 | 8.305E-13 |
| 11 | 45858584 | chr11:45858584 | | rs12419690 | a | | g | 0.5524 | -0.0351 | 0.0064 | 3.892E-08 |
| 11 | 47529947 | chr11:47529947 | | rs7124681 | a | | c | 0.4106 | 0.0362 | 0.0065 | 2.454E-08 |
| 11 | 65294799 | chr11:65294799 | | rs1783541 | t | | c | 0.201 | 0.0608 | 0.0081 | 4.647E-14 |
| 11 | 69456000 | chr11:69456000 | | rs55911137 | c | | g | 0.0259 | -0.1454 | 0.0214 | 9.942E-12 |
| 11 | 72460398 | chr11:72460398 | | rs77464186 | a | | c | 0.8357 | 0.1002 | 0.0087 | 9.304E-31 |
| 11 | 92707806 | chr11:92707806 | | rs191142560 | t | | c | 0.0176 | 0.1432 | 0.0258 | 2.702E-08 |
| 11 | 92708710 | chr11:92708710 | | rs10830963 | c | | g | 0.7208 | -0.101 | 0.0071 | 1.121E-45 |
| 11 | 92756551 | chr11:92756551 | | rs9971402 | a | | c | 0.1437 | 0.0528 | 0.0091 | 6.776E-09 |
| 11 | 93012957 | chr11:93012957 | | rs10444213 | a | | t | 0.539 | 0.0419 | 0.0065 | 1.085E-10 |
| 11 | 128041582 | chr11:128041582 | | rs7933438 | a | | g | 0.1472 | -0.058 | 0.0093 | 4.841E-10 |
| 11 | 128234144 | chr11:128234144 | | rs10750397 | a | | g | 0.2829 | 0.0394 | 0.0071 | 3.132E-08 |
| 11 | 128398938 | chr11:128398938 | | rs67232546 | t | | c | 0.2047 | 0.0531 | 0.008 | 2.505E-11 |
| 12 | 4031104 | chr12:4031104 | | rs10848958 | t | | c | 0.1966 | -0.0451 | 0.0083 | 4.968E-08 |
| 12 | 4200707 | chr12:4200707 | | rs141892016 | a | | g | 0.9921 | 0.2347 | 0.0401 | 4.836E-09 |
| 12 | 4301301 | chr12:4301301 | | rs11063029 | t | | c | 0.0581 | 0.0858 | 0.0138 | 5.364E-10 |
| 12 | 4374373 | chr12:4374373 | | rs11063069 | a | | g | 0.7868 | -0.056 | 0.0079 | 9.941E-13 |
| 12 | 4376091 | chr12:4376091 | | rs12818766 | a | | g | 0.1682 | -0.0538 | 0.0089 | 1.493E-09 |
| 12 | 4384696 | chr12:4384696 | | rs3217792 | t | | c | 0.0866 | -0.1169 | 0.0121 | 6.272E-22 |
| 12 | 4384844 | chr12:4384844 | | rs76895963 | t | | g | 0.9801 | 0.4826 | 0.0275 | 9.124E-69 |
| 12 | 4399050 | chr12:4399050 | | rs3217860 | a | | g | 0.7378 | -0.0536 | 0.0074 | 5.585E-13 |
| 12 | 4406281 | chr12:4406281 | | rs12299509 | a | | g | 0.5304 | -0.0496 | 0.0068 | 3.145E-13 |
| 12 | 12871099 | chr12:12871099 | | rs2066827 | t | | g | 0.7675 | -0.0473 | 0.0081 | 4.438E-09 |
| 12 | 26457190 | chr12:26457190 | | rs1872992 | a | | g | 0.7414 | -0.0457 | 0.0072 | 2.52E-10 |
| 12 | 27826780 | chr12:27826780 | | rs2052673 | t | | g | 0.1702 | -0.0557 | 0.0089 | 3.884E-10 |
| 12 | 27905210 | chr12:27905210 | | rs7969720 | a | | g | 0.6841 | -0.0383 | 0.0069 | 2.983E-08 |
| 12 | 27962934 | chr12:27962934 | | rs7966976 | a | | g | 0.1964 | -0.0751 | 0.0081 | 1.217E-20 |
| 12 | 66221060 | chr12:66221060 | | rs2258238 | a | | t | 0.8963 | -0.102 | 0.0106 | 5.139E-22 |
| 12 | 66371880 | chr12:66371880 | | rs7968682 | t | | g | 0.5072 | 0.0538 | 0.0064 | 3.652E-17 |
| 12 | 71523043 | chr12:71523043 | | rs1705263 | a | | c | 0.4367 | -0.0484 | 0.0064 | 3.507E-14 |
| 12 | 95921998 | chr12:95921998 | | rs61939481 | t | | c | 0.9318 | -0.0714 | 0.0127 | 1.742E-08 |
| 12 | 97848775 | chr12:97848775 | | rs77864822 | a | | g | 0.9295 | 0.0753 | 0.0129 | 5.005E-09 |
| 12 | 108629780 | chr12:108629780 | | rs1426371 | a | | g | 0.2659 | -0.0517 | 0.0073 | 1.738E-12 |
| 12 | 118394008 | chr12:118394008 | | rs7313918 | t | | c | 0.8635 | -0.0559 | 0.0094 | 2.99E-09 |
| 12 | 121105264 | chr12:121105264 | | rs73222800 | a | | g | 0.9289 | 0.078 | 0.0133 | 4.467E-09 |
| 12 | 121278266 | chr12:121278266 | | rs625228 | a | | g | 0.4419 | -0.0359 | 0.0064 | 1.899E-08 |
| 12 | 121297815 | chr12:121297815 | | rs11065299 | a | | g | 0.0757 | 0.0788 | 0.012 | 5.972E-11 |
| 12 | 121380541 | chr12:121380541 | | rs73226260 | a | | g | 0.0328 | -0.121 | 0.0188 | 1.36E-10 |
| 12 | 121416864 | chr12:121416864 | | rs1800574 | t | | c | 0.0294 | 0.1514 | 0.0192 | 2.752E-15 |
| 12 | 121432117 | chr12:121432117 | | rs56348580 | c | | g | 0.3082 | -0.0617 | 0.0069 | 4.313E-19 |
| 12 | 121462139 | chr12:121462139 | | rs2259883 | c | | g | 0.5389 | 0.0417 | 0.0064 | 6.619E-11 |
| 12 | 121501461 | chr12:121501461 | | rs28638142 | a | | c | 0.0464 | 0.0919 | 0.0157 | 4.871E-09 |
| 12 | 121602760 | chr12:121602760 | | rs208302 | a | | g | 0.2278 | 0.0414 | 0.0075 | 3.98E-08 |
| 12 | 123450765 | chr12:123450765 | | rs4148856 | c | | g | 0.7828 | 0.047 | 0.0077 | 1.311E-09 |
| 12 | 124510391 | chr12:124510391 | | rs10773051 | t | | c | 0.235 | -0.0411 | 0.0075 | 4.982E-08 |
| 12 | 133068484 | chr12:133068484 | | rs35318451 | a | | g | 0.331 | 0.0478 | 0.007 | 9.498E-12 |
| 13 | 26776999 | chr13:26776999 | | rs34584161 | a | | g | 0.7609 | 0.0468 | 0.0075 | 5.364E-10 |
| 13 | 31042452 | chr13:31042452 | | rs11842871 | t | | g | 0.2669 | -0.04 | 0.0073 | 4.825E-08 |
| 13 | 33554302 | chr13:33554302 | | rs576674 | a | | g | 0.8313 | -0.0538 | 0.0086 | 3.697E-10 |
| 13 | 51094114 | chr13:51094114 | | rs9316500 | t | | g | 0.7121 | 0.0387 | 0.007 | 3.454E-08 |
| 13 | 58666728 | chr13:58666728 | | rs35559811 | a | | g | 0.7641 | 0.0425 | 0.0077 | 4.128E-08 |
| 13 | 59077406 | chr13:59077406 | | rs9563615 | a | | t | 0.7092 | 0.0408 | 0.0072 | 1.628E-08 |
| 13 | 80717156 | chr13:80717156 | | rs1359790 | a | | g | 0.2791 | -0.0817 | 0.0071 | 1.76E-30 |
| 13 | 108797836 | chr13:108797836 | | rs7325671 | t | | c | 0.1264 | 0.0545 | 0.0096 | 1.533E-08 |
| 14 | 23288935 | chr14:23288935 | | rs17122772 | c | | g | 0.7736 | -0.0428 | 0.0077 | 3.313E-08 |
| 14 | 33302882 | chr14:33302882 | | rs17522122 | t | | g | 0.474 | 0.0356 | 0.0064 | 2.49E-08 |
| 14 | 79944099 | chr14:79944099 | | rs8008910 | a | | g | 0.2192 | 0.0554 | 0.0076 | 4.223E-13 |
| 14 | 91968313 | chr14:91968313 | | rs2896177 | a | | g | 0.5413 | -0.0367 | 0.0064 | 9.128E-09 |
| 14 | 103858673 | chr14:103858673 | | rs3783394 | a | | g | 0.3477 | -0.038 | 0.0067 | 1.421E-08 |
| 15 | 38834033 | chr15:38834033 | | rs8032939 | t | | c | 0.7529 | -0.0418 | 0.0074 | 1.878E-08 |
| 15 | 38873115 | chr15:38873115 | | rs34715063 | t | | c | 0.8778 | -0.0772 | 0.0099 | 8.403E-15 |
| 15 | 41801512 | chr15:41801512 | | rs2289739 | t | | g | 0.3556 | 0.0469 | 0.0067 | 2.578E-12 |
| 15 | 53091553 | chr15:53091553 | | rs2456530 | t | | c | 0.126 | 0.054 | 0.0096 | 2.071E-08 |
| 15 | 57469927 | chr15:57469927 | | rs144801310 | a | | c | 0.9661 | -0.1006 | 0.0183 | 4.01E-08 |
| 15 | 62399093 | chr15:62399093 | | rs11856307 | a | | c | 0.5706 | 0.047 | 0.0065 | 4.481E-13 |
| 15 | 63871292 | chr15:63871292 | | rs7178762 | t | | c | 0.5364 | -0.0387 | 0.0064 | 1.367E-09 |
| 15 | 68080886 | chr15:68080886 | | rs4776970 | a | | t | 0.642 | 0.0378 | 0.0067 | 1.691E-08 |
| 15 | 75932129 | chr15:75932129 | | rs13737 | t | | g | 0.2406 | -0.0481 | 0.0075 | 1.765E-10 |
| 15 | 77508046 | chr15:77508046 | | rs8028830 | a | | g | 0.8542 | -0.0562 | 0.0091 | 6.844E-10 |
| 15 | 77714618 | chr15:77714618 | | rs75145098 | t | | g | 0.9659 | 0.1037 | 0.0184 | 1.829E-08 |
| 15 | 77782335 | chr15:77782335 | | rs12910361 | a | | g | 0.2926 | -0.0814 | 0.007 | 3.951E-31 |
| 15 | 90384045 | chr15:90384045 | | rs2351707 | t | | c | 0.7178 | -0.0644 | 0.007 | 4.301E-20 |
| 15 | 91511260 | chr15:91511260 | | rs12910825 | a | | g | 0.6371 | -0.0525 | 0.0067 | 4.699E-15 |
| 16 | 295795 | chr16:295795 | | rs6600191 | t | | c | 0.8246 | 0.0587 | 0.0085 | 4.471E-12 |
| 16 | 30033633 | chr16:30033633 | | rs12325539 | t | | c | 0.5998 | -0.041 | 0.0065 | 2.686E-10 |
| 16 | 53454855 | chr16:53454855 | | rs62048489 | t | | g | 0.0893 | -0.0711 | 0.0124 | 8.674E-09 |
| 16 | 53769293 | chr16:53769293 | | rs7203521 | a | | g | 0.615 | 0.0424 | 0.0066 | 1.294E-10 |
| 16 | 53792464 | chr16:53792464 | | rs74498370 | t | | c | 0.9367 | -0.1013 | 0.0142 | 1.127E-12 |
| 16 | 53807498 | chr16:53807498 | | rs16952522 | c | | g | 0.9577 | -0.0884 | 0.0161 | 4.195E-08 |
| 16 | 53809123 | chr16:53809123 | | rs55872725 | t | | c | 0.4175 | 0.122 | 0.0065 | 8.509E-79 |
| 16 | 53834684 | chr16:53834684 | | rs12596054 | a | | c | 0.8566 | 0.0586 | 0.0092 | 2.017E-10 |
| 16 | 53843533 | chr16:53843533 | | rs6499646 | t | | c | 0.9215 | 0.0656 | 0.0119 | 3.886E-08 |
| 16 | 53877592 | chr16:53877592 | | rs6499653 | t | | c | 0.2552 | 0.043 | 0.0073 | 4.438E-09 |
| 16 | 69651866 | chr16:69651866 | | rs862320 | t | | c | 0.4231 | -0.0433 | 0.0065 | 2.554E-11 |
| 16 | 75243657 | chr16:75243657 | | rs72802358 | c | | g | 0.1032 | -0.1135 | 0.0107 | 2.215E-26 |
| 16 | 81534790 | chr16:81534790 | | rs2925979 | t | | c | 0.2987 | 0.0546 | 0.007 | 7.068E-15 |
| 16 | 89564055 | chr16:89564055 | | rs12920022 | a | | t | 0.1582 | 0.0528 | 0.0092 | 1.001E-08 |
| 17 | 3828086 | chr17:3828086 | | rs1043246 | c | | g | 0.8445 | -0.0566 | 0.0096 | 4.206E-09 |
| 17 | 3860356 | chr17:3860356 | | rs3826482 | a | | t | 0.5777 | 0.0367 | 0.0066 | 2.64E-08 |
| 17 | 3988451 | chr17:3988451 | | rs8071043 | t | | c | 0.6738 | -0.0523 | 0.0068 | 1.533E-14 |
| 17 | 9793756 | chr17:9793756 | | rs55973554 | a | | g | 0.3215 | 0.039 | 0.0068 | 1.001E-08 |
| 17 | 17661802 | chr17:17661802 | | rs4925109 | a | | g | 0.3177 | 0.0476 | 0.0069 | 5.65E-12 |
| 17 | 36043653 | chr17:36043653 | | rs3094515 | t | | c | 0.3699 | -0.0464 | 0.0072 | 1.339E-10 |
| 17 | 36064897 | chr17:36064897 | | rs2107133 | a | | g | 0.8727 | 0.0643 | 0.0097 | 4.017E-11 |
| 17 | 36099952 | chr17:36099952 | | rs10908278 | a | | t | 0.5174 | -0.0749 | 0.0067 | 5.254E-29 |
| 17 | 40731411 | chr17:40731411 | | rs34855406 | c | | g | 0.2746 | 0.049 | 0.0071 | 5.891E-12 |
| 17 | 47060322 | chr17:47060322 | | rs35895680 | a | | c | 0.323 | -0.0554 | 0.0069 | 1.084E-15 |
| 17 | 62202689 | chr17:62202689 | | rs58642235 | t | | c | 0.8612 | -0.0566 | 0.0094 | 1.896E-09 |
| 17 | 65892507 | chr17:65892507 | | rs61676547 | c | | g | 0.1937 | 0.0525 | 0.0081 | 7.417E-11 |
| 18 | 7070642 | chr18:7070642 | | rs7240767 | t | | c | 0.626 | -0.0372 | 0.0066 | 1.705E-08 |
| 18 | 40087098 | chr18:40087098 | | rs1431841 | t | | g | 0.2113 | 0.0429 | 0.0077 | 3.078E-08 |
| 18 | 53050646 | chr18:53050646 | | rs72926932 | a | | c | 0.9163 | -0.0765 | 0.0114 | 2.04E-11 |
| 18 | 53428347 | chr18:53428347 | | rs17089932 | t | | c | 0.2494 | 0.0407 | 0.0073 | 2.806E-08 |
| 18 | 56876430 | chr18:56876430 | | rs9957320 | t | | g | 0.1719 | -0.0487 | 0.0085 | 9.339E-09 |
| 18 | 57857135 | chr18:57857135 | | rs8097210 | t | | g | 0.7325 | -0.0537 | 0.0072 | 1.061E-13 |
| 18 | 57961249 | chr18:57961249 | | rs111638368 | t | | c | 0.3051 | 0.0391 | 0.0071 | 3.98E-08 |
| 18 | 58050968 | chr18:58050968 | | rs79688165 | t | | c | 0.023 | -0.1383 | 0.0226 | 9.643E-10 |
| 18 | 60845884 | chr18:60845884 | | rs12454712 | t | | c | 0.6129 | 0.0477 | 0.0068 | 2.402E-12 |
| 19 | 4951064 | chr19:4951064 | | rs262549 | c | | g | 0.8067 | -0.0474 | 0.0082 | 6.475E-09 |
| 19 | 7970635 | chr19:7970635 | | rs4804833 | a | | g | 0.3912 | 0.048 | 0.0066 | 3.417E-13 |
| 19 | 13036677 | chr19:13036677 | | rs10419627 | a | | g | 0.591 | 0.0431 | 0.0065 | 3.15E-11 |
| 19 | 19419071 | chr19:19419071 | | rs739846 | a | | g | 0.0761 | 0.0877 | 0.0119 | 2.02E-13 |
| 19 | 33897149 | chr19:33897149 | | rs3786900 | a | | g | 0.7307 | 0.0434 | 0.0072 | 1.886E-09 |
| 19 | 45411941 | chr19:45411941 | | rs429358 | t | | c | 0.8473 | 0.0746 | 0.0091 | 2.621E-16 |
| 19 | 46157019 | chr19:46157019 | | rs10406431 | a | | g | 0.5626 | 0.0603 | 0.0065 | 1.557E-20 |
| 19 | 46178661 | chr19:46178661 | | rs2238689 | t | | c | 0.5787 | -0.0492 | 0.0066 | 8.733E-14 |
| 19 | 47569003 | chr19:47569003 | | rs3810291 | a | | g | 0.6718 | 0.0434 | 0.0069 | 3.376E-10 |
| 20 | 21460433 | chr20:21460433 | | rs17744783 | a | | t | 0.1073 | 0.0577 | 0.0103 | 1.873E-08 |
| 20 | 32381337 | chr20:32381337 | | rs2747567 | a | | g | 0.4263 | -0.0375 | 0.0067 | 2.191E-08 |
| 20 | 32582871 | chr20:32582871 | | rs1007090 | t | | c | 0.3316 | -0.044 | 0.0068 | 1.012E-10 |
| 20 | 43003122 | chr20:43003122 | | rs36112520 | a | | g | 0.1107 | 0.0783 | 0.0104 | 4.225E-14 |
| 20 | 43015281 | chr20:43015281 | | rs6031580 | a | | g | 0.2566 | -0.0429 | 0.0074 | 7.886E-09 |
| 20 | 43042364 | chr20:43042364 | | rs1800961 | t | | c | 0.0349 | 0.1602 | 0.0175 | 5.095E-20 |
| 20 | 45581777 | chr20:45581777 | | rs1999536 | c | | g | 0.5732 | -0.0404 | 0.0065 | 4.863E-10 |
| 20 | 48832135 | chr20:48832135 | | rs11699802 | t | | c | 0.4649 | -0.0443 | 0.0065 | 8.838E-12 |
| 20 | 57397566 | chr20:57397566 | | rs4812034 | t | | g | 0.5465 | 0.0416 | 0.0064 | 7.347E-11 |
| 22 | 30599562 | chr22:30599562 | | rs2023681 | a | | g | 0.087 | -0.0826 | 0.0115 | 7.399E-13 |
| 22 | 41489920 | chr22:41489920 | | rs5758223 | a | | g | 0.7134 | 0.0401 | 0.0071 | 1.779E-08 |
| 22 | 44324727 | chr22:44324727 | | rs738409 | c | | g | 0.7743 | -0.0436 | 0.0076 | 1.168E-08 |
| 22 | 50422348 | chr22:50422348 | | rs36138276 | a | | g | 0.4992 | -0.0431 | 0.0066 | 6.403E-11 |
| 22 | 50604696 | chr22:50604696 | | rs112915006 | a | | g | 0.9489 | -0.0902 | 0.0154 | 4.616E-09 |

| **Supplementary Table 2.** Multivariable MR analysis of the direct effect of BMI on fracture risk and BMD. | | | | | | |
| --- | --- | --- | --- | --- | --- | --- |
| Exposure/Outcome | Adjusted Factors | Multivariate MR Analysis | | | | Mediation Effect(%) |
|  |  | nSNP | OR | 95% CI | P value |  |
| T2D/fracture | None | 289 | 0.965 | (0.943-0.988) | 0.003 |  |
| T2D/fracture | BMI | 390 | 0.974 | (0.953,0.995) | 0.017 | 9.03% |
| T2D/BMD | None | 380 | 0.041 | (0.027-0.054) | 8.14E-09 |  |
| T2D/BMD | BMI | 379 | 0.042 | (0.026,0.057) | 1.92E-07 | 3.60% |

**Supplementary Table 3.** The information of 298 lead SNPs for type 2 diabetes with the risk of fracture.

| CHR | BP | SNP | type 2 diabetes | | | | | fracture | | | | | | | |
| --- | --- | --- | --- | --- | --- | --- | --- | --- | --- | --- | --- | --- | --- | --- | --- |
|  |  |  | A1 | A2 | beta | p | se | A1 | A2 | beta | lb | ub | p | se | padj |
| 1 | 117532790 | rs1127215 | t | c | 0.049 | 3.92E-14 | 0.007 | t | c | -0.010 | -0.026 | 0.007 | 0.2483 | 0.008 | 0.658 |
| 1 | 118143517 | rs320369 | a | g | 0.037 | 4.60E-08 | 0.007 | a | g | 0.001 | -0.016 | 0.018 | 0.8878 | 0.009 | 0.997 |
| 1 | 120526982 | rs1493694 | t | c | 0.080 | 3.35E-15 | 0.010 | t | c | -0.011 | -0.037 | 0.015 | 0.4088 | 0.013 | 0.771 |
| 1 | 177820861 | rs1359939 | a | g | 0.040 | 7.74E-09 | 0.007 | a | g | -0.004 | -0.021 | 0.013 | 0.6559 | 0.009 | 0.944 |
| 1 | 177881651 | rs693232 | c | g | 0.054 | 9.67E-12 | 0.008 | c | g | -0.012 | -0.032 | 0.008 | 0.2226 | 0.010 | 0.655 |
| 1 | 177931532 | rs4650985 | a | c | 0.038 | 4.77E-08 | 0.007 | a | c | 0.005 | -0.012 | 0.023 | 0.5485 | 0.009 | 0.876 |
| 1 | 205099959 | rs3862948 | a | c | 0.041 | 3.98E-08 | 0.008 | a | c | -0.022 | -0.041 | -0.003 | 0.02068 | 0.010 | 0.280 |
| 1 | 206595069 | rs2244510 | t | c | 0.035 | 4.95E-08 | 0.007 | t | c | 0.007 | -0.010 | 0.023 | 0.4306 | 0.008 | 0.790 |
| 1 | 214150445 | rs17712208 | a | t | 0.185 | 1.46E-23 | 0.019 | a | t | -0.018 | -0.071 | 0.035 | 0.5109 | 0.027 | 0.841 |
| 1 | 214159256 | rs340874 | t | c | 0.068 | 1.56E-25 | 0.007 | t | c | -0.004 | -0.020 | 0.012 | 0.6199 | 0.008 | 0.919 |
| 1 | 219748818 | rs2820446 | c | g | 0.059 | 2.83E-17 | 0.007 | c | g | 0.002 | -0.015 | 0.019 | 0.8248 | 0.009 | 0.997 |
| 1 | 219756559 | rs11118352 | t | g | 0.046 | 1.55E-08 | 0.008 | t | g | -0.028 | -0.049 | -0.008 | 0.006434 | 0.010 | 0.157 |
| 1 | 39814960 | rs3754346 | t | c | 0.080 | 2.45E-25 | 0.008 | t | c | -0.026 | -0.046 | -0.007 | 0.007661 | 0.010 | 0.157 |
| 1 | 51266474 | rs3789587 | t | c | 0.067 | 1.79E-09 | 0.011 | t | c | -0.032 | -0.060 | -0.004 | 0.02319 | 0.014 | 0.300 |
| 10 | 114597109 | rs545572 | a | g | 0.076 | 4.24E-29 | 0.007 | a | g | 0.005 | -0.012 | 0.022 | 0.584 | 0.009 | 0.901 |
| 10 | 114648499 | rs1885283 | a | c | 0.073 | 8.17E-13 | 0.010 | a | c | 0.004 | -0.020 | 0.029 | 0.7343 | 0.012 | 0.985 |
| 10 | 114667539 | rs2009075 | t | c | 0.171 | 2.49E-15 | 0.022 | t | c | 0.007 | -0.048 | 0.062 | 0.8027 | 0.028 | 0.997 |
| 10 | 114704781 | rs17746916 | t | c | 0.149 | 3.53E-29 | 0.013 | t | c | 0.011 | -0.024 | 0.045 | 0.5495 | 0.018 | 0.876 |
| 10 | 114754088 | rs7901695 | t | c | 0.285 | 0.00E+00 | 0.007 | t | c | 0.019 | 0.002 | 0.037 | 0.03007 | 0.009 | 0.312 |
| 10 | 114764959 | rs12266632 | c | g | 0.226 | 9.57E-70 | 0.013 | c | g | 0.034 | 0.001 | 0.066 | 0.0411 | 0.016 | 0.350 |
| 10 | 114766717 | rs7896811 | t | c | 0.147 | 3.37E-58 | 0.009 | t | c | 0.013 | -0.010 | 0.036 | 0.255 | 0.012 | 0.658 |
| 10 | 114772429 | rs12354626 | a | g | 0.109 | 1.28E-08 | 0.019 | a | g | -0.015 | -0.067 | 0.038 | 0.5866 | 0.027 | 0.901 |
| 10 | 114797471 | rs17685538 | c | g | 0.095 | 4.15E-19 | 0.011 | c | g | -0.005 | -0.032 | 0.023 | 0.7485 | 0.014 | 0.987 |
| 10 | 114824473 | rs10885410 | a | g | 0.086 | 8.12E-31 | 0.007 | a | g | 0.010 | -0.009 | 0.030 | 0.3035 | 0.010 | 0.680 |
| 10 | 114861304 | rs10885414 | a | g | 0.095 | 1.07E-41 | 0.007 | a | g | -0.001 | -0.019 | 0.017 | 0.9267 | 0.009 | 0.997 |
| 10 | 114865489 | rs10749128 | t | c | 0.046 | 6.10E-10 | 0.007 | t | c | 0.009 | -0.010 | 0.027 | 0.3523 | 0.009 | 0.734 |
| 10 | 114887722 | rs11196236 | t | c | 0.074 | 4.92E-21 | 0.008 | t | c | -0.002 | -0.021 | 0.018 | 0.8712 | 0.010 | 0.997 |
| 10 | 114915214 | rs290483 | t | g | 0.070 | 1.30E-25 | 0.007 | t | g | -0.007 | -0.023 | 0.010 | 0.4391 | 0.008 | 0.790 |
| 10 | 12307894 | rs11257655 | t | c | 0.086 | 1.46E-28 | 0.008 | t | c | -0.018 | -0.038 | 0.002 | 0.07173 | 0.010 | 0.411 |
| 10 | 124193181 | rs2280141 | t | g | 0.049 | 1.91E-14 | 0.006 | t | g | -0.020 | -0.036 | -0.004 | 0.01238 | 0.008 | 0.217 |
| 10 | 71317835 | rs2616055 | a | g | 0.080 | 5.70E-09 | 0.014 | a | g | -0.008 | -0.043 | 0.027 | 0.6513 | 0.018 | 0.942 |
| 10 | 71321279 | rs177045 | a | g | 0.050 | 4.64E-13 | 0.007 | a | g | 0.010 | -0.007 | 0.028 | 0.247 | 0.009 | 0.658 |
| 10 | 71466578 | rs2642588 | t | g | 0.051 | 3.59E-13 | 0.007 | t | g | 0.003 | -0.015 | 0.020 | 0.78 | 0.009 | 0.997 |
| 10 | 80907740 | rs11591689 | t | c | 0.049 | 3.55E-10 | 0.008 | t | c | 0.008 | -0.011 | 0.027 | 0.3839 | 0.010 | 0.766 |
| 10 | 80952826 | rs703972 | c | g | 0.070 | 5.77E-27 | 0.007 | c | g | -0.001 | -0.017 | 0.015 | 0.87 | 0.008 | 0.997 |
| 10 | 80959517 | rs10509406 | t | c | 0.056 | 5.06E-11 | 0.009 | t | c | -0.018 | -0.039 | 0.003 | 0.09193 | 0.011 | 0.460 |
| 10 | 80992381 | rs1574190 | t | c | 0.048 | 1.22E-12 | 0.007 | t | c | 0.009 | -0.007 | 0.025 | 0.2926 | 0.008 | 0.661 |
| 10 | 94138239 | rs7903767 | a | g | 0.055 | 7.21E-18 | 0.006 | a | g | 0.009 | -0.007 | 0.025 | 0.2852 | 0.008 | 0.658 |
| 10 | 94465559 | rs5015480 | t | c | 0.109 | 2.19E-63 | 0.007 | t | c | 0.002 | -0.014 | 0.018 | 0.8397 | 0.008 | 0.997 |
| 10 | 94480391 | rs2497313 | t | c | 0.061 | 8.96E-09 | 0.011 | t | c | 0.010 | -0.017 | 0.036 | 0.4719 | 0.014 | 0.815 |
| 10 | 94500111 | rs11187152 | a | g | 0.070 | 1.85E-08 | 0.012 | a | g | 0.000 | -0.032 | 0.032 | 0.9868 | 0.016 | 0.999 |
| 11 | 128041582 | rs7933438 | a | g | 0.058 | 4.84E-10 | 0.009 | a | g | 0.008 | -0.014 | 0.031 | 0.4624 | 0.012 | 0.811 |
| 11 | 128234144 | rs10750397 | a | g | 0.039 | 3.13E-08 | 0.007 | a | g | 0.001 | -0.017 | 0.019 | 0.8819 | 0.009 | 0.997 |
| 11 | 128389391 | rs11819995 | t | c | 0.050 | 1.30E-10 | 0.008 | t | c | -0.009 | -0.028 | 0.010 | 0.3626 | 0.010 | 0.750 |
| 11 | 17408630 | rs5215 | t | c | 0.071 | 9.85E-27 | 0.007 | t | c | 0.004 | -0.012 | 0.021 | 0.6195 | 0.008 | 0.919 |
| 11 | 2193840 | rs10770141 | a | g | 0.066 | 4.11E-22 | 0.007 | a | g | -0.016 | -0.033 | 0.001 | 0.06104 | 0.009 | 0.396 |
| 11 | 2632430 | rs11023461 | c | g | 0.075 | 5.49E-09 | 0.013 | c | g | 0.006 | -0.029 | 0.041 | 0.7459 | 0.018 | 0.987 |
| 11 | 2663891 | rs756852 | a | g | 0.043 | 2.41E-10 | 0.007 | a | g | 0.005 | -0.016 | 0.026 | 0.6465 | 0.011 | 0.940 |
| 11 | 2691500 | rs231361 | a | g | 0.061 | 3.21E-16 | 0.007 | a | g | -0.005 | -0.024 | 0.014 | 0.6083 | 0.010 | 0.912 |
| 11 | 2752130 | rs231907 | a | t | 0.039 | 2.94E-08 | 0.007 | a | t | 0.007 | -0.011 | 0.025 | 0.4406 | 0.009 | 0.790 |
| 11 | 2847069 | rs163184 | t | g | 0.078 | 2.90E-32 | 0.007 | t | g | -0.018 | -0.034 | -0.002 | 0.02807 | 0.008 | 0.312 |
| 11 | 2858546 | rs2237897 | t | c | 0.192 | 6.79E-31 | 0.017 | t | c | -0.036 | -0.077 | 0.005 | 0.08167 | 0.021 | 0.443 |
| 11 | 34982148 | rs2767036 | a | c | 0.039 | 2.94E-08 | 0.007 | a | c | -0.003 | -0.021 | 0.015 | 0.7344 | 0.009 | 0.985 |
| 11 | 43877934 | rs1061810 | a | c | 0.050 | 8.31E-13 | 0.007 | a | c | -0.006 | -0.024 | 0.012 | 0.5252 | 0.009 | 0.860 |
| 11 | 45870177 | rs10838524 | a | g | 0.035 | 4.25E-08 | 0.006 | a | g | -0.010 | -0.026 | 0.006 | 0.2227 | 0.008 | 0.655 |
| 11 | 47529947 | rs7124681 | a | c | 0.036 | 2.45E-08 | 0.007 | a | c | -0.009 | -0.025 | 0.007 | 0.2858 | 0.008 | 0.658 |
| 11 | 65302893 | rs947791 | a | g | 0.061 | 5.62E-14 | 0.008 | a | g | -0.017 | -0.036 | 0.003 | 0.09873 | 0.010 | 0.467 |
| 11 | 69463273 | rs3918298 | a | g | 0.135 | 1.18E-10 | 0.021 | a | g | -0.045 | -0.106 | 0.015 | 0.139 | 0.031 | 0.560 |
| 11 | 72433098 | rs1552224 | a | c | 0.098 | 1.50E-29 | 0.009 | a | c | -0.002 | -0.024 | 0.020 | 0.8456 | 0.011 | 0.997 |
| 11 | 92708710 | rs10830963 | c | g | 0.101 | 1.12E-45 | 0.007 | c | g | 0.014 | -0.005 | 0.033 | 0.1409 | 0.010 | 0.560 |
| 11 | 92756551 | rs9971402 | a | c | 0.053 | 6.78E-09 | 0.009 | a | c | 0.000 | -0.024 | 0.023 | 0.9704 | 0.012 | 0.997 |
| 11 | 93012957 | rs10444213 | a | t | 0.042 | 1.09E-10 | 0.007 | a | t | 0.016 | -0.001 | 0.033 | 0.06263 | 0.009 | 0.396 |
| 12 | 108618630 | rs3764002 | t | c | 0.051 | 3.12E-12 | 0.007 | t | c | -0.004 | -0.022 | 0.014 | 0.6795 | 0.009 | 0.951 |
| 12 | 121405126 | rs2251468 | a | c | 0.047 | 1.51E-12 | 0.007 | a | c | -0.003 | -0.020 | 0.014 | 0.7374 | 0.009 | 0.985 |
| 12 | 121460686 | rs7957197 | a | t | 0.067 | 3.04E-17 | 0.008 | a | t | -0.002 | -0.022 | 0.018 | 0.8494 | 0.010 | 0.997 |
| 12 | 121602760 | rs208302 | a | g | 0.041 | 3.98E-08 | 0.008 | a | g | 0.010 | -0.009 | 0.029 | 0.2835 | 0.010 | 0.658 |
| 12 | 123593382 | rs7132277 | t | c | 0.048 | 6.02E-09 | 0.008 | t | c | -0.012 | -0.033 | 0.009 | 0.2532 | 0.011 | 0.658 |
| 12 | 12871099 | rs2066827 | t | g | 0.047 | 4.44E-09 | 0.008 | t | g | -0.006 | -0.027 | 0.015 | 0.5602 | 0.011 | 0.883 |
| 12 | 133069134 | rs1882297 | a | g | 0.047 | 1.41E-11 | 0.007 | a | g | -0.008 | -0.025 | 0.010 | 0.3934 | 0.009 | 0.766 |
| 12 | 26457190 | rs1872992 | a | g | 0.046 | 2.52E-10 | 0.007 | a | g | -0.008 | -0.027 | 0.010 | 0.371 | 0.009 | 0.762 |
| 12 | 27826780 | rs2052673 | t | g | 0.056 | 3.88E-10 | 0.009 | t | g | -0.016 | -0.037 | 0.006 | 0.1495 | 0.011 | 0.579 |
| 12 | 27905210 | rs7969720 | a | g | 0.038 | 2.98E-08 | 0.007 | a | g | -0.001 | -0.019 | 0.016 | 0.8795 | 0.009 | 0.997 |
| 12 | 27963676 | rs3751239 | c | g | 0.075 | 1.54E-20 | 0.008 | c | g | 0.001 | -0.019 | 0.021 | 0.945 | 0.010 | 0.997 |
| 12 | 4031104 | rs10848958 | t | c | 0.045 | 4.97E-08 | 0.008 | t | c | -0.014 | -0.034 | 0.006 | 0.1628 | 0.010 | 0.608 |
| 12 | 4374373 | rs11063069 | a | g | 0.056 | 9.94E-13 | 0.008 | a | g | -0.004 | -0.024 | 0.017 | 0.7407 | 0.011 | 0.985 |
| 12 | 4386064 | rs3217795 | a | g | 0.107 | 4.56E-19 | 0.012 | a | g | 0.002 | -0.027 | 0.031 | 0.8939 | 0.015 | 0.997 |
| 12 | 4406281 | rs12299509 | a | g | 0.050 | 3.15E-13 | 0.007 | a | g | 0.001 | -0.016 | 0.018 | 0.9263 | 0.009 | 0.997 |
| 12 | 66215292 | rs2612069 | c | g | 0.103 | 2.19E-21 | 0.011 | c | g | 0.015 | -0.012 | 0.043 | 0.2714 | 0.014 | 0.658 |
| 12 | 66371880 | rs7968682 | t | g | 0.054 | 3.65E-17 | 0.006 | t | g | -0.008 | -0.024 | 0.008 | 0.3357 | 0.008 | 0.720 |
| 12 | 71522953 | rs1796330 | c | g | 0.049 | 6.28E-14 | 0.007 | c | g | 0.001 | -0.015 | 0.017 | 0.9342 | 0.008 | 0.997 |
| 12 | 95928113 | rs11108094 | a | c | 0.071 | 2.28E-08 | 0.013 | a | c | 0.034 | 0.003 | 0.066 | 0.03346 | 0.016 | 0.318 |
| 13 | 108797836 | rs7325671 | t | c | 0.055 | 1.53E-08 | 0.010 | t | c | -0.012 | -0.037 | 0.013 | 0.3465 | 0.013 | 0.734 |
| 13 | 26783692 | rs12865499 | c | g | 0.042 | 2.01E-08 | 0.008 | c | g | 0.005 | -0.014 | 0.024 | 0.6093 | 0.010 | 0.912 |
| 13 | 33554302 | rs576674 | a | g | 0.054 | 3.70E-10 | 0.009 | a | g | -0.003 | -0.024 | 0.019 | 0.8213 | 0.011 | 0.997 |
| 13 | 51094114 | rs9316500 | t | g | 0.039 | 3.45E-08 | 0.007 | t | g | 0.006 | -0.011 | 0.024 | 0.4826 | 0.009 | 0.817 |
| 13 | 58663457 | rs12583517 | t | g | 0.042 | 4.78E-08 | 0.008 | t | g | -0.007 | -0.025 | 0.012 | 0.4948 | 0.010 | 0.826 |
| 13 | 59077406 | rs9563615 | a | t | 0.041 | 1.63E-08 | 0.007 | a | t | 0.008 | -0.010 | 0.025 | 0.4073 | 0.009 | 0.771 |
| 13 | 80717156 | rs1359790 | a | g | 0.082 | 1.76E-30 | 0.007 | a | g | 0.014 | -0.004 | 0.031 | 0.1279 | 0.009 | 0.544 |
| 14 | 103858673 | rs3783394 | a | g | 0.038 | 1.42E-08 | 0.007 | a | g | -0.025 | -0.042 | -0.008 | 0.003517 | 0.009 | 0.118 |
| 14 | 33302882 | rs17522122 | t | g | 0.036 | 2.49E-08 | 0.006 | t | g | 0.000 | -0.016 | 0.016 | 0.9939 | 0.008 | 0.999 |
| 14 | 79910119 | rs17109221 | t | c | 0.056 | 7.18E-13 | 0.008 | t | c | -0.002 | -0.021 | 0.018 | 0.8489 | 0.010 | 0.997 |
| 14 | 91971265 | rs7143394 | t | c | 0.036 | 1.20E-08 | 0.006 | t | c | -0.009 | -0.025 | 0.007 | 0.2829 | 0.008 | 0.658 |
| 15 | 38834033 | rs8032939 | t | c | 0.042 | 1.88E-08 | 0.007 | t | c | -0.001 | -0.019 | 0.018 | 0.956 | 0.010 | 0.997 |
| 15 | 41801512 | rs2289739 | t | g | 0.047 | 2.58E-12 | 0.007 | t | g | 0.009 | -0.008 | 0.026 | 0.3222 | 0.009 | 0.696 |
| 15 | 53091553 | rs2456530 | t | c | 0.054 | 2.07E-08 | 0.010 | t | c | 0.005 | -0.019 | 0.029 | 0.6963 | 0.012 | 0.958 |
| 15 | 62394264 | rs8037894 | c | g | 0.047 | 6.29E-13 | 0.007 | c | g | 0.004 | -0.012 | 0.020 | 0.6339 | 0.008 | 0.935 |
| 15 | 63871292 | rs7178762 | t | c | 0.039 | 1.37E-09 | 0.006 | t | c | -0.004 | -0.020 | 0.012 | 0.6079 | 0.008 | 0.912 |
| 15 | 68080886 | rs4776970 | a | t | 0.038 | 1.69E-08 | 0.007 | a | t | 0.008 | -0.009 | 0.024 | 0.3744 | 0.009 | 0.764 |
| 15 | 75932129 | rs13737 | t | g | 0.048 | 1.77E-10 | 0.008 | t | g | -0.015 | -0.034 | 0.004 | 0.1238 | 0.010 | 0.535 |
| 15 | 77508046 | rs8028830 | a | g | 0.056 | 6.84E-10 | 0.009 | a | g | -0.017 | -0.040 | 0.006 | 0.1396 | 0.012 | 0.560 |
| 15 | 77782335 | rs12910361 | a | g | 0.081 | 3.95E-31 | 0.007 | a | g | -0.022 | -0.039 | -0.004 | 0.01516 | 0.009 | 0.238 |
| 15 | 90423293 | rs4932265 | t | c | 0.065 | 1.59E-19 | 0.007 | t | c | -0.025 | -0.043 | -0.006 | 0.007663 | 0.009 | 0.157 |
| 15 | 91511260 | rs12910825 | a | g | 0.053 | 4.70E-15 | 0.007 | a | g | 0.028 | 0.012 | 0.045 | 0.0008483 | 0.009 | 0.073 |
| 16 | 295795 | rs6600191 | t | c | 0.059 | 4.47E-12 | 0.009 | t | c | 0.015 | -0.006 | 0.035 | 0.1713 | 0.011 | 0.611 |
| 16 | 30033633 | rs12325539 | t | c | 0.041 | 2.69E-10 | 0.007 | t | c | 0.009 | -0.007 | 0.026 | 0.262 | 0.008 | 0.658 |
| 16 | 53769293 | rs7203521 | a | g | 0.042 | 1.29E-10 | 0.007 | a | g | -0.007 | -0.024 | 0.009 | 0.3767 | 0.008 | 0.764 |
| 16 | 53800954 | rs1421085 | t | c | 0.122 | 1.52E-78 | 0.007 | t | c | -0.024 | -0.041 | -0.007 | 0.006019 | 0.009 | 0.157 |
| 16 | 53807498 | rs16952522 | c | g | 0.088 | 4.20E-08 | 0.016 | c | g | 0.003 | -0.050 | 0.055 | 0.9274 | 0.027 | 0.997 |
| 16 | 53843533 | rs6499646 | t | c | 0.066 | 3.89E-08 | 0.012 | t | c | -0.007 | -0.037 | 0.023 | 0.6468 | 0.015 | 0.940 |
| 16 | 53844579 | rs17218700 | a | g | 0.062 | 2.97E-09 | 0.010 | a | g | -0.007 | -0.032 | 0.018 | 0.5789 | 0.013 | 0.899 |
| 16 | 53877592 | rs6499653 | t | c | 0.043 | 4.44E-09 | 0.007 | t | c | -0.010 | -0.029 | 0.009 | 0.2922 | 0.010 | 0.661 |
| 16 | 69622762 | rs244418 | a | g | 0.043 | 3.88E-11 | 0.007 | a | g | 0.009 | -0.007 | 0.025 | 0.2863 | 0.008 | 0.658 |
| 16 | 75242012 | rs889512 | c | g | 0.111 | 2.40E-25 | 0.011 | c | g | 0.009 | -0.017 | 0.036 | 0.4961 | 0.014 | 0.826 |
| 16 | 81534790 | rs2925979 | t | c | 0.055 | 7.07E-15 | 0.007 | t | c | -0.016 | -0.034 | 0.001 | 0.0654 | 0.009 | 0.398 |
| 17 | 17661802 | rs4925109 | a | g | 0.048 | 5.65E-12 | 0.007 | a | g | -0.003 | -0.020 | 0.014 | 0.7298 | 0.009 | 0.985 |
| 17 | 36043653 | rs3094515 | t | c | 0.046 | 1.34E-10 | 0.007 | t | c | 0.013 | -0.007 | 0.032 | 0.1937 | 0.010 | 0.654 |
| 17 | 36064897 | rs2107133 | a | g | 0.064 | 4.02E-11 | 0.010 | a | g | -0.019 | -0.043 | 0.005 | 0.1137 | 0.012 | 0.511 |
| 17 | 36098987 | rs4239217 | a | g | 0.068 | 1.03E-24 | 0.007 | a | g | -0.005 | -0.022 | 0.012 | 0.5451 | 0.009 | 0.876 |
| 17 | 3860356 | rs3826482 | a | t | 0.037 | 2.64E-08 | 0.007 | a | t | -0.014 | -0.030 | 0.002 | 0.09081 | 0.008 | 0.460 |
| 17 | 4045440 | rs1377807 | c | g | 0.052 | 3.38E-14 | 0.007 | c | g | -0.013 | -0.031 | 0.004 | 0.1326 | 0.009 | 0.557 |
| 17 | 40731597 | rs6963 | a | t | 0.047 | 4.92E-11 | 0.007 | a | t | -0.036 | -0.053 | -0.018 | 0.00007313 | 0.009 | 0.011 |
| 17 | 46970259 | rs1962412 | t | c | 0.052 | 1.11E-13 | 0.007 | t | c | -0.004 | -0.021 | 0.014 | 0.6828 | 0.009 | 0.951 |
| 17 | 65898809 | rs7216064 | a | g | 0.051 | 1.87E-10 | 0.008 | a | g | 0.016 | -0.004 | 0.036 | 0.1148 | 0.010 | 0.511 |
| 17 | 9787958 | rs7219033 | a | g | 0.039 | 1.30E-08 | 0.007 | a | g | -0.002 | -0.019 | 0.015 | 0.8389 | 0.009 | 0.997 |
| 18 | 40087098 | rs1431841 | t | g | 0.043 | 3.08E-08 | 0.008 | t | g | 0.000 | -0.020 | 0.020 | 0.9936 | 0.010 | 0.999 |
| 18 | 56876430 | rs9957320 | t | g | 0.049 | 9.34E-09 | 0.009 | t | g | -0.013 | -0.035 | 0.008 | 0.2172 | 0.011 | 0.655 |
| 18 | 57858829 | rs8089364 | t | c | 0.054 | 1.31E-13 | 0.007 | t | c | 0.011 | -0.007 | 0.029 | 0.2338 | 0.009 | 0.658 |
| 18 | 58055731 | rs7227255 | a | g | 0.137 | 1.12E-09 | 0.022 | a | g | -0.028 | -0.085 | 0.028 | 0.3213 | 0.029 | 0.696 |
| 18 | 60845884 | rs12454712 | t | c | 0.048 | 2.40E-12 | 0.007 | t | c | -0.002 | -0.020 | 0.016 | 0.8476 | 0.009 | 0.997 |
| 18 | 7070642 | rs7240767 | t | c | 0.037 | 1.71E-08 | 0.007 | t | c | -0.003 | -0.020 | 0.013 | 0.6895 | 0.008 | 0.956 |
| 19 | 13002563 | rs2242517 | t | g | 0.043 | 7.09E-11 | 0.007 | t | g | 0.000 | -0.017 | 0.017 | 0.9843 | 0.009 | 0.999 |
| 19 | 19407718 | rs10401969 | t | c | 0.088 | 3.46E-13 | 0.012 | t | c | 0.033 | 0.002 | 0.065 | 0.03812 | 0.016 | 0.334 |
| 19 | 45392254 | rs6857 | t | c | 0.066 | 1.50E-14 | 0.009 | t | c | -0.001 | -0.022 | 0.020 | 0.9519 | 0.011 | 0.997 |
| 19 | 46157019 | rs10406431 | a | g | 0.060 | 1.56E-20 | 0.007 | a | g | 0.001 | -0.016 | 0.019 | 0.8755 | 0.009 | 0.997 |
| 19 | 46172278 | rs11671664 | a | g | 0.061 | 7.61E-10 | 0.010 | a | g | -0.008 | -0.034 | 0.019 | 0.5717 | 0.013 | 0.897 |
| 19 | 47569003 | rs3810291 | a | g | 0.043 | 3.38E-10 | 0.007 | a | g | 0.001 | -0.017 | 0.018 | 0.9454 | 0.009 | 0.997 |
| 19 | 7970635 | rs4804833 | a | g | 0.048 | 3.42E-13 | 0.007 | a | g | 0.004 | -0.012 | 0.021 | 0.5949 | 0.008 | 0.909 |
| 2 | 121317747 | rs11677557 | a | g | 0.059 | 5.66E-11 | 0.009 | a | g | -0.001 | -0.024 | 0.021 | 0.9182 | 0.012 | 0.997 |
| 2 | 121348224 | rs3860374 | t | c | 0.039 | 3.02E-09 | 0.007 | t | c | -0.001 | -0.017 | 0.016 | 0.9548 | 0.008 | 0.997 |
| 2 | 158329237 | rs16841827 | a | g | 0.082 | 6.34E-10 | 0.013 | a | g | -0.021 | -0.054 | 0.012 | 0.2208 | 0.017 | 0.655 |
| 2 | 161136656 | rs7572970 | a | g | 0.045 | 6.12E-10 | 0.007 | a | g | 0.006 | -0.012 | 0.024 | 0.5038 | 0.009 | 0.834 |
| 2 | 165528876 | rs13389219 | t | c | 0.061 | 1.17E-20 | 0.007 | t | c | 0.000 | -0.016 | 0.016 | 0.9837 | 0.008 | 0.999 |
| 2 | 165566877 | rs3820981 | a | g | 0.040 | 5.92E-10 | 0.007 | a | g | 0.005 | -0.011 | 0.021 | 0.5335 | 0.008 | 0.869 |
| 2 | 227101411 | rs2972144 | a | g | 0.091 | 2.19E-43 | 0.007 | a | g | 0.004 | -0.012 | 0.021 | 0.6006 | 0.009 | 0.912 |
| 2 | 25655391 | rs11126052 | c | g | 0.040 | 2.85E-08 | 0.007 | c | g | 0.028 | 0.010 | 0.046 | 0.002576 | 0.009 | 0.118 |
| 2 | 27730940 | rs1260326 | t | c | 0.064 | 1.62E-22 | 0.007 | t | c | 0.002 | -0.014 | 0.018 | 0.793 | 0.008 | 0.997 |
| 2 | 43429058 | rs10171620 | t | g | 0.040 | 7.92E-10 | 0.007 | t | g | 0.009 | -0.008 | 0.025 | 0.2864 | 0.008 | 0.658 |
| 2 | 43687879 | rs17030845 | t | c | 0.119 | 2.35E-28 | 0.011 | t | c | 0.019 | -0.007 | 0.045 | 0.144 | 0.013 | 0.565 |
| 2 | 58990485 | rs2862874 | t | g | 0.036 | 4.83E-08 | 0.007 | t | g | 0.002 | -0.015 | 0.018 | 0.8536 | 0.008 | 0.997 |
| 2 | 59307725 | rs6545714 | a | g | 0.036 | 2.25E-08 | 0.007 | a | g | -0.012 | -0.028 | 0.005 | 0.1677 | 0.008 | 0.611 |
| 2 | 60585806 | rs243019 | t | c | 0.059 | 3.37E-20 | 0.006 | t | c | -0.001 | -0.017 | 0.015 | 0.8698 | 0.008 | 0.997 |
| 2 | 65296280 | rs2252867 | t | c | 0.054 | 3.05E-16 | 0.007 | t | c | 0.000 | -0.017 | 0.016 | 0.9702 | 0.009 | 0.997 |
| 2 | 653195 | rs13396935 | a | g | 0.051 | 1.46E-09 | 0.009 | a | g | 0.002 | -0.020 | 0.023 | 0.8909 | 0.011 | 0.997 |
| 2 | 65574192 | rs13431750 | t | c | 0.112 | 4.48E-08 | 0.020 | t | c | -0.048 | -0.101 | 0.005 | 0.07743 | 0.027 | 0.435 |
| 2 | 65655012 | rs2028150 | c | g | 0.054 | 1.31E-16 | 0.007 | c | g | -0.011 | -0.027 | 0.005 | 0.1826 | 0.008 | 0.633 |
| 20 | 21451848 | rs4813428 | t | c | 0.060 | 3.78E-08 | 0.011 | t | c | -0.018 | -0.046 | 0.009 | 0.1952 | 0.014 | 0.654 |
| 20 | 32582871 | rs1007090 | t | c | 0.044 | 1.01E-10 | 0.007 | t | c | 0.002 | -0.015 | 0.019 | 0.8149 | 0.009 | 0.997 |
| 20 | 43015281 | rs6031580 | a | g | 0.043 | 7.89E-09 | 0.007 | a | g | -0.007 | -0.026 | 0.011 | 0.4451 | 0.009 | 0.790 |
| 20 | 43042364 | rs1800961 | t | c | 0.160 | 5.10E-20 | 0.018 | t | c | -0.020 | -0.066 | 0.026 | 0.3866 | 0.023 | 0.766 |
| 20 | 45581777 | rs1999536 | c | g | 0.040 | 4.86E-10 | 0.007 | c | g | 0.001 | -0.015 | 0.017 | 0.9044 | 0.008 | 0.997 |
| 20 | 48830265 | rs6012876 | t | c | 0.043 | 4.31E-11 | 0.007 | t | c | 0.025 | 0.009 | 0.041 | 0.002869 | 0.008 | 0.118 |
| 20 | 57397566 | rs4812034 | t | g | 0.042 | 7.35E-11 | 0.006 | t | g | -0.014 | -0.030 | 0.002 | 0.08111 | 0.008 | 0.443 |
| 22 | 30599562 | rs2023681 | a | g | 0.083 | 7.40E-13 | 0.012 | a | g | 0.004 | -0.025 | 0.032 | 0.8005 | 0.014 | 0.997 |
| 22 | 41489920 | rs5758223 | a | g | 0.040 | 1.78E-08 | 0.007 | a | g | 0.010 | -0.007 | 0.028 | 0.2477 | 0.009 | 0.658 |
| 22 | 44324727 | rs738409 | c | g | 0.044 | 1.17E-08 | 0.008 | c | g | 0.021 | 0.002 | 0.041 | 0.03037 | 0.010 | 0.312 |
| 22 | 50435480 | rs5771069 | a | g | 0.041 | 2.97E-10 | 0.007 | a | g | 0.000 | -0.016 | 0.016 | 0.9586 | 0.008 | 0.997 |
| 22 | 50588313 | rs12484907 | a | g | 0.087 | 1.17E-08 | 0.015 | a | g | -0.003 | -0.042 | 0.037 | 0.9027 | 0.020 | 0.997 |
| 3 | 123065778 | rs11708067 | a | g | 0.088 | 5.05E-30 | 0.008 | a | g | -0.024 | -0.043 | -0.005 | 0.01313 | 0.010 | 0.217 |
| 3 | 12315297 | rs2347101 | a | g | 0.060 | 1.09E-09 | 0.010 | a | g | -0.017 | -0.042 | 0.007 | 0.1733 | 0.013 | 0.611 |
| 3 | 12336507 | rs11709077 | a | g | 0.104 | 1.26E-26 | 0.010 | a | g | -0.010 | -0.035 | 0.014 | 0.4072 | 0.013 | 0.771 |
| 3 | 12396588 | rs4135247 | a | g | 0.044 | 1.51E-11 | 0.007 | a | g | 0.000 | -0.016 | 0.017 | 0.9734 | 0.008 | 0.997 |
| 3 | 124925934 | rs569255 | a | g | 0.040 | 5.64E-10 | 0.006 | a | g | -0.002 | -0.018 | 0.014 | 0.8368 | 0.008 | 0.997 |
| 3 | 152392877 | rs9828639 | t | g | 0.038 | 1.83E-08 | 0.007 | t | g | -0.009 | -0.025 | 0.008 | 0.3196 | 0.009 | 0.696 |
| 3 | 152535365 | rs10513432 | t | c | 0.092 | 1.78E-08 | 0.016 | t | c | 0.005 | -0.036 | 0.046 | 0.8032 | 0.021 | 0.997 |
| 3 | 168223132 | rs7642311 | a | g | 0.055 | 1.53E-08 | 0.010 | a | g | 0.015 | -0.009 | 0.039 | 0.2242 | 0.012 | 0.655 |
| 3 | 170724883 | rs8192675 | t | c | 0.066 | 5.77E-21 | 0.007 | t | c | 0.007 | -0.011 | 0.024 | 0.4618 | 0.009 | 0.811 |
| 3 | 185493590 | rs16860219 | t | g | 0.122 | 4.49E-09 | 0.021 | t | g | 0.073 | 0.019 | 0.127 | 0.007895 | 0.028 | 0.157 |
| 3 | 185510613 | rs7633675 | t | g | 0.109 | 3.21E-57 | 0.007 | t | g | -0.021 | -0.038 | -0.004 | 0.01644 | 0.009 | 0.245 |
| 3 | 185531697 | rs4686698 | a | g | 0.095 | 2.86E-08 | 0.017 | a | g | 0.023 | -0.021 | 0.067 | 0.3105 | 0.023 | 0.685 |
| 3 | 186665645 | rs3887925 | t | c | 0.054 | 1.01E-16 | 0.007 | t | c | -0.004 | -0.020 | 0.013 | 0.6673 | 0.008 | 0.945 |
| 3 | 187740523 | rs6808574 | t | c | 0.061 | 2.29E-20 | 0.007 | t | c | 0.014 | -0.002 | 0.030 | 0.09471 | 0.008 | 0.463 |
| 3 | 23454790 | rs1496653 | a | g | 0.067 | 2.49E-17 | 0.008 | a | g | -0.019 | -0.039 | 0.000 | 0.05263 | 0.010 | 0.388 |
| 3 | 23578594 | rs17013433 | a | g | 0.113 | 8.85E-11 | 0.018 | a | g | -0.003 | -0.049 | 0.043 | 0.8979 | 0.024 | 0.997 |
| 3 | 49860854 | rs6446298 | t | c | 0.037 | 2.60E-08 | 0.007 | t | c | -0.014 | -0.031 | 0.003 | 0.1063 | 0.009 | 0.487 |
| 3 | 50174197 | rs2624847 | t | g | 0.041 | 3.48E-08 | 0.007 | t | g | -0.007 | -0.025 | 0.012 | 0.4815 | 0.009 | 0.817 |
| 3 | 53123273 | rs891368 | a | g | 0.038 | 3.25E-09 | 0.006 | a | g | -0.015 | -0.031 | 0.001 | 0.0631 | 0.008 | 0.396 |
| 3 | 63897215 | rs2292662 | t | c | 0.065 | 2.24E-13 | 0.009 | t | c | -0.014 | -0.036 | 0.008 | 0.2142 | 0.011 | 0.655 |
| 3 | 64701146 | rs9860730 | a | g | 0.054 | 1.55E-14 | 0.007 | a | g | -0.006 | -0.024 | 0.011 | 0.4729 | 0.009 | 0.815 |
| 4 | 103900090 | rs10516495 | a | t | 0.042 | 1.46E-09 | 0.007 | a | t | 0.015 | -0.003 | 0.032 | 0.09738 | 0.009 | 0.467 |
| 4 | 153513369 | rs7669833 | a | t | 0.057 | 3.52E-16 | 0.007 | a | t | -0.010 | -0.028 | 0.008 | 0.268 | 0.009 | 0.658 |
| 4 | 17811914 | rs7667864 | a | c | 0.040 | 1.64E-08 | 0.007 | a | c | -0.011 | -0.029 | 0.006 | 0.2054 | 0.009 | 0.654 |
| 4 | 185718132 | rs745805 | a | t | 0.063 | 4.52E-14 | 0.008 | a | t | 0.003 | -0.018 | 0.024 | 0.7979 | 0.011 | 0.997 |
| 4 | 45175691 | rs13130484 | t | c | 0.044 | 8.49E-11 | 0.007 | t | c | -0.016 | -0.032 | 0.001 | 0.06247 | 0.008 | 0.396 |
| 4 | 6257188 | rs4689381 | t | c | 0.038 | 8.37E-09 | 0.007 | t | c | 0.001 | -0.016 | 0.017 | 0.9483 | 0.008 | 0.997 |
| 4 | 6306763 | rs10937721 | c | g | 0.085 | 5.39E-38 | 0.007 | c | g | -0.002 | -0.018 | 0.015 | 0.8427 | 0.008 | 0.997 |
| 4 | 744972 | rs1531583 | t | g | 0.100 | 1.85E-10 | 0.016 | t | g | -0.022 | -0.063 | 0.019 | 0.2871 | 0.021 | 0.658 |
| 4 | 83582546 | rs1848068 | t | c | 0.039 | 7.03E-09 | 0.007 | t | c | -0.012 | -0.029 | 0.005 | 0.1605 | 0.009 | 0.608 |
| 4 | 89740894 | rs1903002 | c | g | 0.036 | 1.45E-08 | 0.006 | c | g | 0.009 | -0.008 | 0.025 | 0.3066 | 0.008 | 0.682 |
| 4 | 95093855 | rs7678054 | a | g | 0.039 | 6.88E-10 | 0.006 | a | g | -0.001 | -0.017 | 0.015 | 0.9213 | 0.008 | 0.997 |
| 5 | 101558769 | rs6891076 | a | c | 0.066 | 1.60E-09 | 0.011 | a | c | -0.021 | -0.049 | 0.007 | 0.1349 | 0.014 | 0.558 |
| 5 | 101711999 | rs7733501 | t | c | 0.045 | 3.75E-10 | 0.007 | t | c | -0.017 | -0.034 | 0.001 | 0.06691 | 0.009 | 0.399 |
| 5 | 102100576 | rs7729395 | t | c | 0.147 | 4.14E-23 | 0.015 | t | c | -0.023 | -0.062 | 0.016 | 0.2401 | 0.020 | 0.658 |
| 5 | 102360747 | rs17296280 | a | c | 0.046 | 8.02E-11 | 0.007 | a | c | -0.017 | -0.034 | 0.000 | 0.04574 | 0.009 | 0.368 |
| 5 | 102704259 | rs10068434 | t | c | 0.039 | 3.39E-08 | 0.007 | t | c | -0.022 | -0.040 | -0.003 | 0.02002 | 0.009 | 0.280 |
| 5 | 102754950 | rs13188193 | t | c | 0.111 | 4.52E-14 | 0.015 | t | c | -0.022 | -0.060 | 0.016 | 0.2514 | 0.019 | 0.658 |
| 5 | 133864599 | rs329122 | a | g | 0.037 | 1.72E-08 | 0.007 | a | g | -0.006 | -0.022 | 0.010 | 0.4726 | 0.008 | 0.815 |
| 5 | 14753745 | rs17250977 | a | g | 0.118 | 3.64E-11 | 0.018 | a | g | 0.001 | -0.051 | 0.053 | 0.9671 | 0.027 | 0.997 |
| 5 | 14777799 | rs9312873 | a | g | 0.068 | 7.14E-10 | 0.011 | a | g | -0.022 | -0.049 | 0.005 | 0.1059 | 0.014 | 0.487 |
| 5 | 14810899 | rs31911 | a | g | 0.037 | 7.58E-09 | 0.006 | a | g | -0.009 | -0.025 | 0.007 | 0.2818 | 0.008 | 0.658 |
| 5 | 44683819 | rs12187196 | a | c | 0.040 | 7.92E-10 | 0.007 | a | c | 0.014 | -0.002 | 0.030 | 0.08843 | 0.008 | 0.460 |
| 5 | 53271420 | rs702634 | a | g | 0.050 | 3.37E-13 | 0.007 | a | g | 0.003 | -0.014 | 0.021 | 0.6973 | 0.009 | 0.958 |
| 5 | 53321956 | rs255761 | t | g | 0.054 | 1.06E-11 | 0.008 | t | g | 0.002 | -0.018 | 0.022 | 0.8628 | 0.010 | 0.997 |
| 5 | 53463117 | rs154021 | t | c | 0.042 | 2.03E-08 | 0.007 | t | c | 0.002 | -0.017 | 0.021 | 0.8451 | 0.010 | 0.997 |
| 5 | 55806751 | rs459193 | a | g | 0.072 | 6.78E-23 | 0.007 | a | g | -0.015 | -0.033 | 0.004 | 0.1172 | 0.009 | 0.514 |
| 5 | 55861894 | rs9687846 | a | g | 0.070 | 3.47E-18 | 0.008 | a | g | -0.013 | -0.033 | 0.007 | 0.2085 | 0.010 | 0.654 |
| 5 | 75003678 | rs2307111 | t | c | 0.050 | 1.05E-14 | 0.007 | t | c | 0.001 | -0.016 | 0.017 | 0.9363 | 0.008 | 0.997 |
| 5 | 76427311 | rs6878122 | a | g | 0.061 | 6.45E-18 | 0.007 | a | g | -0.003 | -0.020 | 0.015 | 0.7633 | 0.009 | 0.996 |
| 5 | 78522895 | rs10078815 | t | c | 0.042 | 1.43E-10 | 0.007 | t | c | 0.017 | 0.001 | 0.034 | 0.0438 | 0.009 | 0.363 |
| 6 | 107433400 | rs1665901 | a | t | 0.040 | 4.97E-09 | 0.007 | a | t | 0.007 | -0.011 | 0.026 | 0.4391 | 0.009 | 0.790 |
| 6 | 126964510 | rs4273712 | a | g | 0.059 | 1.03E-16 | 0.007 | a | g | -0.030 | -0.048 | -0.012 | 0.0009778 | 0.009 | 0.073 |
| 6 | 127409882 | rs4580892 | t | c | 0.040 | 8.46E-09 | 0.007 | t | c | -0.058 | -0.076 | -0.039 | 3.68E-10 | 0.009 | 0.000 |
| 6 | 127414838 | rs719727 | a | g | 0.059 | 1.67E-15 | 0.007 | a | g | 0.019 | 0.000 | 0.037 | 0.05213 | 0.010 | 0.388 |
| 6 | 137301998 | rs11752908 | a | g | 0.046 | 2.15E-12 | 0.007 | a | g | 0.008 | -0.008 | 0.024 | 0.3478 | 0.008 | 0.734 |
| 6 | 153440062 | rs6901126 | t | c | 0.036 | 4.16E-08 | 0.007 | t | c | -0.007 | -0.023 | 0.009 | 0.3837 | 0.008 | 0.766 |
| 6 | 160770312 | rs474513 | a | g | 0.040 | 4.18E-10 | 0.006 | a | g | -0.016 | -0.032 | 0.001 | 0.06372 | 0.009 | 0.396 |
| 6 | 164107529 | rs17630640 | a | g | 0.054 | 2.48E-08 | 0.010 | a | g | 0.001 | -0.023 | 0.024 | 0.9655 | 0.012 | 0.997 |
| 6 | 20429804 | rs12523853 | t | c | 0.097 | 7.83E-09 | 0.017 | t | c | 0.034 | -0.015 | 0.083 | 0.1707 | 0.025 | 0.611 |
| 6 | 20518450 | rs6903706 | a | g | 0.062 | 1.34E-13 | 0.008 | a | g | 0.008 | -0.013 | 0.029 | 0.4437 | 0.011 | 0.790 |
| 6 | 20601915 | rs10946394 | a | c | 0.057 | 2.66E-14 | 0.007 | a | c | 0.011 | -0.008 | 0.031 | 0.2578 | 0.010 | 0.658 |
| 6 | 20686996 | rs9368222 | a | c | 0.138 | 1.42E-83 | 0.007 | a | c | 0.011 | -0.007 | 0.029 | 0.2312 | 0.009 | 0.658 |
| 6 | 20689945 | rs7748720 | a | g | 0.042 | 3.63E-08 | 0.008 | a | g | -0.008 | -0.027 | 0.012 | 0.4297 | 0.010 | 0.790 |
| 6 | 20760177 | rs6909558 | t | c | 0.085 | 1.80E-13 | 0.012 | t | c | 0.018 | -0.011 | 0.048 | 0.2221 | 0.015 | 0.655 |
| 6 | 20876613 | rs4077404 | a | g | 0.057 | 1.88E-13 | 0.008 | a | g | 0.006 | -0.013 | 0.025 | 0.549 | 0.010 | 0.876 |
| 6 | 31122315 | rs130066 | c | g | 0.042 | 3.52E-11 | 0.006 | c | g | 0.001 | -0.015 | 0.017 | 0.8928 | 0.008 | 0.997 |
| 6 | 31282777 | rs1634746 | t | c | 0.047 | 2.58E-12 | 0.007 | t | c | -0.004 | -0.022 | 0.015 | 0.681 | 0.009 | 0.951 |
| 6 | 31321123 | rs2523612 | t | g | 0.069 | 5.42E-14 | 0.009 | a | c | 0.021 | -0.009 | 0.051 | 0.1744 | 0.015 | 0.611 |
| 6 | 31472720 | rs2855812 | t | g | 0.043 | 7.28E-09 | 0.007 | t | g | -0.017 | -0.036 | 0.001 | 0.06911 | 0.010 | 0.404 |
| 6 | 31518169 | rs2844492 | a | g | 0.135 | 1.38E-09 | 0.022 | a | g | -0.029 | -0.100 | 0.042 | 0.4215 | 0.036 | 0.790 |
| 6 | 31628733 | rs805262 | t | c | 0.038 | 2.96E-09 | 0.006 | t | c | -0.018 | -0.034 | -0.002 | 0.03024 | 0.008 | 0.312 |
| 6 | 31845985 | rs9267658 | t | c | 0.072 | 3.83E-15 | 0.009 | t | c | -0.032 | -0.055 | -0.009 | 0.006838 | 0.012 | 0.157 |
| 6 | 32187605 | rs1109771 | a | g | 0.041 | 3.99E-10 | 0.007 | a | g | -0.022 | -0.042 | -0.001 | 0.03803 | 0.011 | 0.334 |
| 6 | 32411846 | rs2239802 | c | g | 0.063 | 2.77E-16 | 0.008 | c | g | -0.009 | -0.028 | 0.011 | 0.3926 | 0.010 | 0.766 |
| 6 | 32441100 | rs9269081 | a | c | 0.057 | 1.87E-15 | 0.007 | a | c | -0.004 | -0.022 | 0.014 | 0.669 | 0.009 | 0.945 |
| 6 | 32591588 | rs9271608 | a | g | 0.085 | 1.91E-22 | 0.009 | a | g | 0.008 | -0.012 | 0.028 | 0.4365 | 0.010 | 0.790 |
| 6 | 32636866 | rs3134996 | a | t | 0.042 | 1.60E-09 | 0.007 | a | t | 0.011 | -0.006 | 0.027 | 0.2074 | 0.009 | 0.654 |
| 6 | 32781112 | rs2071479 | t | c | 0.116 | 1.19E-10 | 0.018 | t | c | 0.011 | -0.035 | 0.057 | 0.6465 | 0.024 | 0.940 |
| 6 | 32814447 | rs6457684 | t | c | 0.037 | 1.31E-08 | 0.007 | t | c | -0.014 | -0.031 | 0.002 | 0.09261 | 0.009 | 0.460 |
| 6 | 43758873 | rs6905288 | a | g | 0.037 | 8.35E-09 | 0.007 | a | g | -0.010 | -0.027 | 0.007 | 0.2346 | 0.009 | 0.658 |
| 6 | 43815364 | rs6937438 | a | g | 0.051 | 2.35E-13 | 0.007 | a | g | -0.011 | -0.028 | 0.007 | 0.2396 | 0.009 | 0.658 |
| 6 | 50803050 | rs987237 | a | g | 0.060 | 3.20E-13 | 0.008 | a | g | -0.012 | -0.033 | 0.008 | 0.2523 | 0.011 | 0.658 |
| 6 | 7153152 | rs4585612 | t | c | 0.041 | 1.80E-10 | 0.007 | t | c | -0.009 | -0.025 | 0.007 | 0.2673 | 0.008 | 0.658 |
| 6 | 7231843 | rs9379084 | a | g | 0.099 | 5.48E-21 | 0.011 | a | g | -0.025 | -0.050 | 0.001 | 0.06309 | 0.013 | 0.396 |
| 6 | 7275941 | rs11243150 | t | c | 0.051 | 3.09E-15 | 0.007 | t | c | -0.001 | -0.018 | 0.015 | 0.8719 | 0.008 | 0.997 |
| 7 | 102486254 | rs11496066 | t | c | 0.051 | 8.17E-10 | 0.008 | t | c | 0.000 | -0.020 | 0.021 | 0.9661 | 0.011 | 0.997 |
| 7 | 130465654 | rs1447059 | a | g | 0.052 | 2.50E-15 | 0.007 | a | g | 0.007 | -0.010 | 0.023 | 0.4319 | 0.009 | 0.790 |
| 7 | 14898282 | rs17168486 | t | c | 0.067 | 4.50E-16 | 0.008 | t | c | 0.001 | -0.020 | 0.022 | 0.9176 | 0.011 | 0.997 |
| 7 | 150508720 | rs6955948 | t | c | 0.040 | 1.79E-08 | 0.007 | t | c | -0.021 | -0.039 | -0.002 | 0.02563 | 0.009 | 0.312 |
| 7 | 15062983 | rs2215383 | t | c | 0.064 | 1.06E-23 | 0.006 | t | c | 0.014 | -0.002 | 0.030 | 0.08832 | 0.008 | 0.460 |
| 7 | 156977794 | rs3802120 | t | c | 0.058 | 3.78E-18 | 0.007 | t | c | 0.025 | 0.008 | 0.042 | 0.003571 | 0.009 | 0.118 |
| 7 | 27978715 | rs6948511 | t | c | 0.064 | 1.06E-08 | 0.011 | t | c | 0.004 | -0.024 | 0.032 | 0.7839 | 0.015 | 0.997 |
| 7 | 28191672 | rs10225433 | a | g | 0.077 | 2.86E-08 | 0.014 | a | g | 0.043 | -0.001 | 0.087 | 0.0537 | 0.022 | 0.388 |
| 7 | 28196413 | rs849135 | a | g | 0.091 | 1.13E-45 | 0.006 | a | g | -0.002 | -0.018 | 0.014 | 0.7811 | 0.008 | 0.997 |
| 7 | 44200884 | rs2041547 | t | c | 0.041 | 1.68E-10 | 0.006 | t | c | 0.010 | -0.006 | 0.026 | 0.2084 | 0.008 | 0.654 |
| 7 | 44231778 | rs2971669 | t | c | 0.060 | 3.87E-14 | 0.008 | t | c | 0.011 | -0.009 | 0.031 | 0.2689 | 0.010 | 0.658 |
| 8 | 10903475 | rs2001433 | a | t | 0.044 | 9.84E-12 | 0.007 | a | t | -0.021 | -0.037 | -0.004 | 0.01311 | 0.008 | 0.217 |
| 8 | 11450422 | rs2244648 | a | g | 0.036 | 2.25E-08 | 0.007 | a | g | -0.016 | -0.032 | 0.000 | 0.0547 | 0.008 | 0.388 |
| 8 | 116482423 | rs3808415 | a | g | 0.035 | 4.65E-08 | 0.006 | a | g | -0.011 | -0.027 | 0.005 | 0.1632 | 0.008 | 0.608 |
| 8 | 118185025 | rs3802177 | a | g | 0.108 | 9.16E-55 | 0.007 | a | g | -0.020 | -0.037 | -0.002 | 0.02647 | 0.009 | 0.312 |
| 8 | 118190620 | rs16889471 | a | g | 0.045 | 7.38E-09 | 0.008 | a | g | -0.004 | -0.024 | 0.017 | 0.727 | 0.010 | 0.985 |
| 8 | 128711742 | rs17772814 | a | g | 0.075 | 2.13E-09 | 0.013 | a | g | 0.016 | -0.021 | 0.053 | 0.3998 | 0.019 | 0.771 |
| 8 | 129568078 | rs1561927 | t | c | 0.042 | 6.12E-09 | 0.007 | t | c | 0.010 | -0.008 | 0.028 | 0.2688 | 0.009 | 0.658 |
| 8 | 145511201 | rs4977218 | a | t | 0.051 | 2.71E-13 | 0.007 | a | t | -0.007 | -0.024 | 0.009 | 0.3881 | 0.009 | 0.766 |
| 8 | 145639726 | rs2272662 | t | c | 0.040 | 2.63E-08 | 0.007 | t | c | 0.020 | 0.001 | 0.038 | 0.03412 | 0.009 | 0.318 |
| 8 | 145903077 | rs997258 | a | c | 0.038 | 6.95E-09 | 0.007 | a | c | 0.007 | -0.009 | 0.023 | 0.4041 | 0.008 | 0.771 |
| 8 | 19844415 | rs7819706 | a | g | 0.067 | 2.82E-11 | 0.010 | a | g | 0.003 | -0.021 | 0.028 | 0.7854 | 0.013 | 0.997 |
| 8 | 41503727 | rs755162 | a | c | 0.040 | 1.00E-08 | 0.007 | a | c | 0.000 | -0.018 | 0.018 | 0.9988 | 0.009 | 0.999 |
| 8 | 41522991 | rs508419 | a | g | 0.081 | 5.43E-27 | 0.008 | a | g | -0.013 | -0.032 | 0.007 | 0.2035 | 0.010 | 0.654 |
| 8 | 41555473 | rs2241896 | t | c | 0.045 | 1.28E-10 | 0.007 | t | c | 0.004 | -0.013 | 0.021 | 0.6677 | 0.009 | 0.945 |
| 8 | 41593309 | rs17603828 | a | g | 0.080 | 1.78E-09 | 0.013 | a | g | -0.021 | -0.054 | 0.012 | 0.2077 | 0.017 | 0.654 |
| 8 | 8304502 | rs2921077 | a | g | 0.037 | 9.14E-09 | 0.007 | a | g | -0.024 | -0.040 | -0.008 | 0.003571 | 0.008 | 0.118 |
| 8 | 9200472 | rs6990912 | a | c | 0.043 | 2.05E-10 | 0.007 | a | c | -0.011 | -0.028 | 0.006 | 0.1867 | 0.009 | 0.640 |
| 8 | 9265105 | rs17662402 | t | c | 0.084 | 4.52E-09 | 0.014 | t | c | 0.005 | -0.031 | 0.040 | 0.8001 | 0.018 | 0.997 |
| 8 | 95965695 | rs13257021 | a | g | 0.049 | 3.11E-14 | 0.006 | a | g | 0.005 | -0.011 | 0.021 | 0.5573 | 0.008 | 0.883 |
| 8 | 9974824 | rs17689007 | a | g | 0.047 | 5.62E-13 | 0.007 | a | g | -0.008 | -0.024 | 0.008 | 0.3498 | 0.008 | 0.734 |
| 9 | 136149229 | rs505922 | t | c | 0.047 | 3.65E-12 | 0.007 | t | c | 0.002 | -0.016 | 0.019 | 0.8611 | 0.009 | 0.997 |
| 9 | 139240630 | rs28539249 | t | c | 0.055 | 3.10E-15 | 0.007 | a | g | 0.004 | -0.017 | 0.024 | 0.7368 | 0.011 | 0.985 |
| 9 | 19065295 | rs7467618 | t | c | 0.039 | 1.41E-09 | 0.007 | t | c | -0.002 | -0.018 | 0.014 | 0.8054 | 0.008 | 0.997 |
| 9 | 22003367 | rs1063192 | a | g | 0.058 | 8.17E-19 | 0.007 | a | g | 0.000 | -0.016 | 0.016 | 0.9979 | 0.008 | 0.999 |
| 9 | 22051295 | rs17694555 | a | g | 0.061 | 2.34E-08 | 0.011 | a | g | 0.010 | -0.018 | 0.038 | 0.477 | 0.014 | 0.817 |
| 9 | 22128600 | rs10811658 | a | g | 0.052 | 2.27E-13 | 0.007 | a | g | -0.003 | -0.023 | 0.017 | 0.7602 | 0.010 | 0.996 |
| 9 | 22128709 | rs12347779 | c | g | 0.079 | 4.22E-08 | 0.014 | c | g | -0.024 | -0.067 | 0.019 | 0.2797 | 0.022 | 0.658 |
| 9 | 22134094 | rs10811661 | t | c | 0.160 | 3.16E-77 | 0.009 | t | c | 0.007 | -0.014 | 0.029 | 0.4942 | 0.011 | 0.826 |
| 9 | 22140224 | rs2065501 | a | c | 0.044 | 1.50E-09 | 0.007 | a | c | -0.003 | -0.022 | 0.016 | 0.7652 | 0.010 | 0.996 |
| 9 | 28410683 | rs1412234 | t | c | 0.039 | 7.71E-09 | 0.007 | t | c | -0.005 | -0.022 | 0.012 | 0.5787 | 0.009 | 0.899 |
| 9 | 4291928 | rs10974438 | a | c | 0.051 | 1.71E-14 | 0.007 | a | c | -0.004 | -0.021 | 0.013 | 0.6673 | 0.009 | 0.945 |
| 9 | 81905590 | rs17791513 | a | g | 0.102 | 1.35E-14 | 0.013 | a | g | -0.036 | -0.068 | -0.003 | 0.03327 | 0.017 | 0.318 |
| 9 | 84308948 | rs2796441 | a | g | 0.067 | 2.97E-25 | 0.007 | a | g | -0.002 | -0.018 | 0.015 | 0.8293 | 0.008 | 0.997 |
| 9 | 96970677 | rs12004367 | a | g | 0.037 | 3.90E-08 | 0.007 | a | g | -0.017 | -0.035 | 0.000 | 0.04866 | 0.009 | 0.382 |

**Supplementary Table** **4.** The information of 389 SNPs for type 2 diabetes with BMD.

| RSID | CHR | BP | type 2 diabetes | | | | | | BMD | | | | | |
| --- | --- | --- | --- | --- | --- | --- | --- | --- | --- | --- | --- | --- | --- | --- |
|  |  |  | A1 | A2 | eaf | BETA | P.value | SE | A1 | A2 | eaf | BETA | SE | P.value |
| rs3768321 | 1 | 40035928 | T | G | 0.199 | 0.084 | 4.75E-26 | 0.008 | G | T | 0.803 | -0.014 | 0.002 | 1.00E-05 |
| rs58432198 | 1 | 51256091 | T | C | 0.117 | -0.064 | 5.71E-10 | 0.010 | C | T | 0.883 | 0.013 | 0.003 | 0.0013 |
| rs12140153 | 1 | 62579891 | T | G | 0.095 | -0.065 | 1.17E-08 | 0.011 | G | T | 0.904 | 0.004 | 0.003 | 0.19 |
| rs1127215 | 1 | 117532790 | T | C | 0.415 | -0.049 | 3.92E-14 | 0.007 | C | T | 0.579 | 0.003 | 0.002 | 0.14 |
| rs320369 | 1 | 118143517 | A | G | 0.323 | 0.037 | 4.60E-08 | 0.007 | A | G | 0.319 | 0.002 | 0.002 | 0.61 |
| rs1493694 | 1 | 120526982 | T | C | 0.110 | 0.080 | 3.35E-15 | 0.010 | C | T | 0.892 | 0.007 | 0.003 | 0.074 |
| rs1359939 | 1 | 177820861 | A | G | 0.307 | -0.040 | 7.74E-09 | 0.007 | G | A | 0.693 | 0.003 | 0.002 | 0.084 |
| rs490689 | 1 | 177873841 | A | G | 0.200 | 0.054 | 8.86E-12 | 0.008 | G | A | 0.789 | -0.011 | 0.002 | 6.20E-06 |
| rs4650985 | 1 | 177931532 | A | C | 0.289 | -0.038 | 4.77E-08 | 0.007 | C | A | 0.717 | 0.005 | 0.002 | 0.0032 |
| rs3862948 | 1 | 205099959 | A | C | 0.235 | 0.041 | 3.98E-08 | 0.008 | C | A | 0.770 | -0.004 | 0.002 | 0.26 |
| rs79687284 | 1 | 214150821 | C | G | 0.034 | 0.188 | 8.25E-24 | 0.019 | G | C | 0.965 | 0.001 | 0.005 | 0.67 |
| rs3738430 | 1 | 214157546 | A | G | 0.025 | -0.118 | 2.62E-08 | 0.021 | G | A | 0.977 | -0.019 | 0.006 | 0.0029 |
| rs340874 | 1 | 214159256 | T | C | 0.449 | -0.068 | 1.56E-25 | 0.007 | T | C | 0.429 | -0.004 | 0.002 | 0.088 |
| rs2820446 | 1 | 219748818 | C | G | 0.708 | 0.059 | 2.83E-17 | 0.007 | C | G | 0.701 | -0.002 | 0.002 | 0.34 |
| rs2785962 | 1 | 219754739 | A | G | 0.191 | 0.046 | 1.55E-08 | 0.008 | A | G | 0.191 | 0.016 | 0.002 | 8.00E-09 |
| rs348330 | 1 | 229672955 | A | G | 0.640 | -0.049 | 2.10E-13 | 0.007 | G | A | 0.366 | 0.007 | 0.002 | 1.30E-05 |
| rs62107261 | 2 | 422144 | T | C | 0.954 | 0.102 | 2.62E-10 | 0.016 | T | C | 0.952 | 0.021 | 0.004 | 1.60E-06 |
| rs13396935 | 2 | 653195 | A | G | 0.171 | -0.051 | 1.46E-09 | 0.009 | G | A | 0.828 | 0.015 | 0.002 | 5.90E-08 |
| rs34048824 | 2 | 25535543 | T | C | 0.569 | 0.036 | 2.06E-08 | 0.007 | T | C | 0.567 | 0.000 | 0.002 | 0.27 |
| rs11126052 | 2 | 25655391 | C | G | 0.734 | 0.040 | 2.85E-08 | 0.007 | C | G | 0.732 | -0.002 | 0.002 | 0.63 |
| rs1260326 | 2 | 27730940 | T | C | 0.393 | -0.064 | 1.62E-22 | 0.007 | T | C | 0.393 | -0.005 | 0.002 | 0.016 |
| rs10171620 | 2 | 43429058 | T | G | 0.464 | 0.040 | 7.92E-10 | 0.007 | G | T | 0.528 | 0.010 | 0.002 | 2.30E-08 |
| rs17030845 | 2 | 43687879 | T | C | 0.101 | -0.119 | 2.35E-28 | 0.011 | C | T | 0.893 | -0.027 | 0.003 | 3.30E-17 |
| rs142564661 | 2 | 43994026 | T | C | 0.015 | -0.163 | 1.45E-08 | 0.029 | C | T | 0.982 | -0.037 | 0.007 | 5.90E-06 |
| rs2862874 | 2 | 58990485 | T | G | 0.606 | 0.036 | 4.83E-08 | 0.007 | G | T | 0.389 | 0.002 | 0.002 | 0.37 |
| rs6545714 | 2 | 59307725 | A | G | 0.608 | -0.036 | 2.25E-08 | 0.007 | G | A | 0.399 | -0.003 | 0.002 | 0.14 |
| rs243019 | 2 | 60585806 | T | C | 0.536 | -0.059 | 3.37E-20 | 0.006 | T | C | 0.545 | 0.002 | 0.002 | 0.56 |
| rs2540945 | 2 | 65289825 | A | G | 0.633 | 0.054 | 3.05E-16 | 0.007 | A | G | 0.638 | 0.003 | 0.002 | 0.32 |
| rs61330787 | 2 | 65577885 | C | G | 0.972 | 0.113 | 2.31E-08 | 0.020 | C | G | 0.976 | 0.011 | 0.006 | 0.14 |
| rs2028150 | 2 | 65655012 | C | G | 0.598 | 0.054 | 1.31E-16 | 0.007 | C | G | 0.591 | 0.010 | 0.002 | 0.00029 |
| rs11688931 | 2 | 121318166 | C | G | 0.850 | 0.059 | 4.53E-11 | 0.009 | C | G | 0.846 | 0.000 | 0.003 | 0.99 |
| rs4849906 | 2 | 121342814 | A | G | 0.423 | 0.039 | 3.02E-09 | 0.007 | A | G | 0.420 | -0.002 | 0.002 | 0.37 |
| rs16841827 | 2 | 158329237 | A | G | 0.063 | -0.082 | 6.34E-10 | 0.013 | G | A | 0.936 | 0.004 | 0.004 | 0.76 |
| rs7572970 | 2 | 161136656 | A | G | 0.266 | -0.045 | 6.12E-10 | 0.007 | A | G | 0.279 | 0.004 | 0.002 | 0.15 |
| rs13389219 | 2 | 165528876 | T | C | 0.401 | -0.061 | 1.17E-20 | 0.007 | C | T | 0.607 | -0.003 | 0.002 | 0.34 |
| rs1869525 | 2 | 165572081 | T | C | 0.563 | 0.040 | 5.36E-10 | 0.007 | T | C | 0.560 | -0.002 | 0.002 | 0.64 |
| rs4675045 | 2 | 226952104 | C | G | 0.792 | -0.054 | 1.64E-11 | 0.008 | C | G | 0.785 | 0.001 | 0.002 | 0.96 |
| rs72979712 | 2 | 227019416 | T | G | 0.053 | -0.093 | 3.26E-10 | 0.015 | G | T | 0.950 | -0.001 | 0.004 | 0.7 |
| rs2972144 | 2 | 227101411 | A | G | 0.362 | -0.091 | 2.19E-43 | 0.007 | A | G | 0.353 | -0.006 | 0.002 | 0.018 |
| rs41504645 | 2 | 227131174 | T | C | 0.877 | -0.069 | 1.49E-12 | 0.010 | T | C | 0.877 | -0.003 | 0.003 | 0.34 |
| rs76367336 | 2 | 227228251 | T | C | 0.886 | 0.058 | 1.26E-08 | 0.010 | T | C | 0.891 | -0.001 | 0.003 | 0.34 |
| rs145268310 | 3 | 12310773 | C | G | 0.117 | 0.061 | 7.40E-10 | 0.010 | G | C | 0.878 | -0.006 | 0.003 | 0.3 |
| rs11709077 | 3 | 12336507 | A | G | 0.126 | -0.104 | 1.26E-26 | 0.010 | G | A | 0.880 | 0.007 | 0.003 | 0.0085 |
| rs4518111 | 3 | 12377344 | A | C | 0.434 | 0.047 | 2.87E-12 | 0.007 | A | C | 0.435 | -0.008 | 0.002 | 1.80E-05 |
| rs1496653 | 3 | 23454790 | A | G | 0.789 | 0.067 | 2.49E-17 | 0.008 | A | G | 0.795 | 0.003 | 0.002 | 0.45 |
| rs74486672 | 3 | 23618935 | T | C | 0.036 | 0.116 | 3.81E-11 | 0.018 | C | T | 0.971 | -0.008 | 0.006 | 0.34 |
| rs4688760 | 3 | 49980596 | T | C | 0.683 | 0.042 | 2.14E-09 | 0.007 | C | T | 0.309 | -0.015 | 0.002 | 4.90E-13 |
| rs891368 | 3 | 53123273 | A | G | 0.434 | -0.038 | 3.25E-09 | 0.006 | A | G | 0.441 | -0.012 | 0.002 | 2.90E-08 |
| rs76263492 | 3 | 54828827 | T | G | 0.045 | 0.090 | 8.94E-09 | 0.016 | G | T | 0.957 | 0.006 | 0.005 | 0.31 |
| rs3774725 | 3 | 63974258 | T | G | 0.849 | 0.066 | 1.95E-13 | 0.009 | T | G | 0.860 | 0.002 | 0.003 | 0.45 |
| rs4368494 | 3 | 64701387 | A | G | 0.298 | -0.054 | 1.39E-14 | 0.007 | G | A | 0.712 | 0.002 | 0.002 | 0.39 |
| rs11708067 | 3 | 123065778 | A | G | 0.775 | 0.088 | 5.05E-30 | 0.008 | A | G | 0.755 | 0.013 | 0.002 | 2.50E-07 |
| rs569255 | 3 | 124925934 | A | G | 0.539 | -0.040 | 5.64E-10 | 0.006 | G | A | 0.467 | 0.000 | 0.002 | 0.63 |
| rs62271373 | 3 | 150066540 | A | T | 0.055 | 0.088 | 1.03E-09 | 0.014 | T | A | 0.940 | -0.028 | 0.004 | 1.50E-12 |
| rs9828639 | 3 | 152392877 | T | G | 0.327 | 0.038 | 1.83E-08 | 0.007 | T | G | 0.330 | 0.001 | 0.002 | 0.53 |
| rs78569745 | 3 | 152456697 | T | G | 0.958 | 0.099 | 2.34E-09 | 0.017 | T | G | 0.964 | -0.006 | 0.005 | 0.037 |
| rs7642311 | 3 | 168223132 | A | G | 0.872 | 0.055 | 1.53E-08 | 0.010 | A | G | 0.877 | -0.003 | 0.003 | 0.23 |
| rs8192675 | 3 | 170724883 | T | C | 0.712 | 0.066 | 5.77E-21 | 0.007 | T | C | 0.713 | -0.003 | 0.002 | 0.11 |
| rs62290256 | 3 | 185256242 | A | G | 0.936 | -0.077 | 1.00E-08 | 0.013 | A | G | 0.941 | -0.010 | 0.004 | 0.049 |
| rs7616221 | 3 | 185354394 | T | G | 0.035 | 0.102 | 6.02E-09 | 0.018 | T | G | 0.036 | 0.010 | 0.005 | 0.071 |
| rs75974105 | 3 | 185488509 | C | G | 0.034 | 0.101 | 1.35E-08 | 0.018 | G | C | 0.969 | -0.008 | 0.005 | 0.23 |
| rs7633675 | 3 | 185510613 | T | G | 0.684 | -0.109 | 3.21E-57 | 0.007 | T | G | 0.685 | -0.015 | 0.002 | 4.90E-08 |
| rs113672528 | 3 | 185540817 | A | C | 0.975 | -0.128 | 9.80E-10 | 0.021 | A | C | 0.976 | -0.025 | 0.006 | 0.0038 |
| rs2194411 | 3 | 185548663 | A | G | 0.129 | -0.075 | 1.84E-14 | 0.010 | G | A | 0.872 | 0.012 | 0.003 | 0.012 |
| rs3887925 | 3 | 186665645 | T | C | 0.545 | 0.054 | 1.01E-16 | 0.007 | C | T | 0.450 | -0.002 | 0.002 | 0.37 |
| rs6808574 | 3 | 187740523 | T | C | 0.389 | -0.061 | 2.29E-20 | 0.007 | T | C | 0.390 | 0.002 | 0.002 | 0.11 |
| rs56187241 | 4 | 738275 | T | C | 0.045 | 0.101 | 1.07E-10 | 0.016 | C | T | 0.961 | -0.007 | 0.005 | 0.13 |
| rs56337234 | 4 | 1784403 | T | C | 0.498 | -0.055 | 9.75E-17 | 0.007 | C | T | 0.506 | 0.005 | 0.002 | 0.0096 |
| rs362307 | 4 | 3241845 | T | C | 0.078 | 0.070 | 6.78E-09 | 0.012 | C | T | 0.925 | 0.005 | 0.004 | 0.14 |
| rs4689381 | 4 | 6257188 | T | C | 0.444 | 0.038 | 8.37E-09 | 0.007 | C | T | 0.562 | -0.002 | 0.002 | 0.42 |
| rs10937721 | 4 | 6306763 | C | G | 0.589 | 0.085 | 5.39E-38 | 0.007 | G | C | 0.410 | 0.000 | 0.002 | 0.82 |
| rs7667864 | 4 | 17811914 | A | C | 0.284 | -0.040 | 1.64E-08 | 0.007 | A | C | 0.287 | -0.014 | 0.002 | 2.10E-08 |
| rs13130484 | 4 | 45175691 | T | C | 0.430 | 0.044 | 8.49E-11 | 0.007 | C | T | 0.566 | -0.008 | 0.002 | 4.90E-05 |
| rs10471048 | 4 | 83587562 | C | G | 0.657 | -0.039 | 6.43E-09 | 0.007 | G | C | 0.336 | 0.003 | 0.002 | 0.074 |
| rs1903002 | 4 | 89740894 | C | G | 0.494 | -0.036 | 1.45E-08 | 0.006 | C | G | 0.507 | 0.002 | 0.002 | 0.44 |
| rs6821438 | 4 | 95091911 | A | G | 0.534 | 0.040 | 5.10E-10 | 0.006 | A | G | 0.530 | -0.018 | 0.002 | 3.90E-17 |
| rs10516495 | 4 | 103900090 | A | T | 0.313 | -0.042 | 1.46E-09 | 0.007 | T | A | 0.693 | -0.008 | 0.002 | 0.00098 |
| rs7669833 | 4 | 153513369 | A | T | 0.295 | -0.057 | 3.52E-16 | 0.007 | T | A | 0.707 | 0.003 | 0.002 | 0.32 |
| rs28819812 | 4 | 157652753 | A | C | 0.322 | -0.040 | 4.22E-08 | 0.007 | C | A | 0.681 | 0.007 | 0.002 | 0.012 |
| rs745805 | 4 | 185718132 | A | T | 0.820 | 0.063 | 4.52E-14 | 0.008 | A | T | 0.825 | -0.009 | 0.002 | 0.00029 |
| rs17250977 | 5 | 14753745 | A | G | 0.963 | -0.118 | 3.64E-11 | 0.018 | A | G | 0.960 | 0.002 | 0.005 | 0.98 |
| rs147581833 | 5 | 14755919 | T | C | 0.006 | -0.371 | 3.11E-14 | 0.049 | C | T | 0.993 | -0.015 | 0.011 | 0.32 |
| rs6885132 | 5 | 14768092 | C | G | 0.904 | 0.077 | 2.49E-12 | 0.011 | C | G | 0.900 | 0.000 | 0.003 | 0.99 |
| rs143306347 | 5 | 14784165 | A | C | 0.012 | -0.175 | 2.67E-08 | 0.031 | C | A | 0.987 | -0.014 | 0.008 | 0.027 |
| rs879253 | 5 | 14794868 | T | C | 0.565 | -0.037 | 1.00E-08 | 0.007 | C | T | 0.449 | 0.007 | 0.002 | 0.0052 |
| rs6884702 | 5 | 44682589 | A | G | 0.606 | -0.040 | 7.92E-10 | 0.007 | A | G | 0.605 | 0.002 | 0.002 | 0.61 |
| rs17261179 | 5 | 51791225 | T | C | 0.515 | 0.036 | 2.72E-08 | 0.006 | T | C | 0.513 | -0.003 | 0.002 | 0.036 |
| rs3811978 | 5 | 52100489 | A | G | 0.834 | -0.049 | 6.58E-09 | 0.009 | A | G | 0.830 | 0.003 | 0.002 | 0.57 |
| rs702634 | 5 | 53271420 | A | G | 0.689 | 0.050 | 3.37E-13 | 0.007 | G | A | 0.307 | -0.006 | 0.002 | 0.00042 |
| rs255761 | 5 | 53321956 | T | G | 0.202 | 0.054 | 1.06E-11 | 0.008 | G | T | 0.795 | 0.002 | 0.002 | 0.81 |
| rs154021 | 5 | 53463117 | T | C | 0.754 | 0.042 | 2.03E-08 | 0.007 | T | C | 0.757 | 0.003 | 0.002 | 0.17 |
| rs459193 | 5 | 55806751 | A | G | 0.259 | -0.072 | 6.78E-23 | 0.007 | A | G | 0.253 | 0.008 | 0.002 | 8.00E-04 |
| rs9687846 | 5 | 55861894 | A | G | 0.190 | 0.070 | 3.47E-18 | 0.008 | G | A | 0.800 | -0.009 | 0.002 | 5.80E-05 |
| rs2307111 | 5 | 75003678 | T | C | 0.604 | 0.050 | 1.05E-14 | 0.007 | T | C | 0.607 | 0.007 | 0.002 | 0.00017 |
| rs7732130 | 5 | 76435004 | A | G | 0.699 | -0.061 | 5.69E-18 | 0.007 | G | A | 0.319 | 0.004 | 0.002 | 0.12 |
| rs1316776 | 5 | 78430607 | A | C | 0.352 | -0.043 | 1.04E-10 | 0.007 | C | A | 0.651 | 0.009 | 0.002 | 7.20E-05 |
| rs77372998 | 5 | 101250990 | A | G | 0.992 | -0.283 | 5.69E-12 | 0.041 | A | G | 0.991 | -0.008 | 0.010 | 0.24 |
| rs145510090 | 5 | 101273694 | A | T | 0.053 | 0.098 | 2.77E-11 | 0.015 | T | A | 0.948 | -0.007 | 0.004 | 0.19 |
| rs3114659 | 5 | 101631592 | A | G | 0.316 | 0.041 | 2.09E-09 | 0.007 | A | G | 0.324 | 0.005 | 0.002 | 0.03 |
| rs145762933 | 5 | 101870140 | A | G | 0.050 | 0.145 | 5.57E-22 | 0.015 | G | A | 0.950 | -0.007 | 0.004 | 0.19 |
| rs17154913 | 5 | 102328490 | C | G | 0.684 | -0.044 | 1.64E-10 | 0.007 | C | G | 0.681 | -0.007 | 0.002 | 0.00037 |
| rs78408340 | 5 | 102338739 | C | G | 0.992 | -0.379 | 2.35E-22 | 0.039 | C | G | 0.990 | -0.011 | 0.009 | 0.013 |
| rs115505614 | 5 | 102422968 | T | C | 0.050 | 0.166 | 7.58E-29 | 0.015 | C | T | 0.948 | -0.005 | 0.004 | 0.52 |
| rs116407196 | 5 | 102973337 | A | G | 0.959 | -0.098 | 6.34E-09 | 0.017 | A | G | 0.957 | -0.011 | 0.005 | 0.1 |
| rs329122 | 5 | 133864599 | A | G | 0.429 | 0.037 | 1.72E-08 | 0.007 | G | A | 0.581 | 0.003 | 0.002 | 0.22 |
| rs6921580 | 6 | 7203714 | C | G | 0.416 | 0.049 | 3.66E-13 | 0.007 | C | G | 0.419 | 0.008 | 0.002 | 1.50E-05 |
| rs9379084 | 6 | 7231843 | A | G | 0.112 | -0.099 | 5.48E-21 | 0.011 | G | A | 0.885 | 0.035 | 0.003 | 5.10E-28 |
| rs1815311 | 6 | 7245458 | A | G | 0.652 | -0.051 | 2.43E-14 | 0.007 | A | G | 0.651 | 0.006 | 0.002 | 0.0032 |
| rs11243150 | 6 | 7275941 | T | C | 0.602 | 0.051 | 3.09E-15 | 0.007 | T | C | 0.598 | -0.007 | 0.002 | 0.0052 |
| rs6903706 | 6 | 20518450 | A | G | 0.181 | -0.062 | 1.34E-13 | 0.008 | G | A | 0.814 | -0.005 | 0.002 | 0.23 |
| rs58761454 | 6 | 20541166 | T | C | 0.254 | 0.050 | 8.98E-12 | 0.007 | C | T | 0.758 | 0.004 | 0.002 | 0.062 |
| rs76393458 | 6 | 20553878 | A | G | 0.018 | 0.151 | 1.47E-09 | 0.025 | G | A | 0.981 | -0.003 | 0.007 | 0.49 |
| rs78421627 | 6 | 20633973 | A | T | 0.973 | -0.125 | 7.33E-10 | 0.020 | A | T | 0.975 | 0.002 | 0.006 | 0.85 |
| rs72830693 | 6 | 20683900 | C | G | 0.965 | -0.142 | 6.56E-16 | 0.018 | C | G | 0.967 | 0.001 | 0.005 | 0.71 |
| rs9368222 | 6 | 20686996 | A | C | 0.272 | 0.138 | 1.42E-83 | 0.007 | C | A | 0.738 | 0.002 | 0.002 | 0.48 |
| rs6915155 | 6 | 20725940 | T | G | 0.557 | -0.049 | 1.17E-14 | 0.006 | G | T | 0.439 | -0.003 | 0.002 | 0.019 |
| rs77786848 | 6 | 20894610 | A | G | 0.947 | -0.110 | 8.36E-14 | 0.015 | A | G | 0.946 | 0.001 | 0.004 | 0.72 |
| rs7775995 | 6 | 20931156 | A | G | 0.689 | 0.049 | 1.82E-12 | 0.007 | A | G | 0.685 | -0.005 | 0.002 | 0.032 |
| rs2523897 | 6 | 30993958 | A | G | 0.163 | -0.049 | 3.07E-08 | 0.009 | A | G | 0.161 | 0.001 | 0.002 | 0.69 |
| rs3131012 | 6 | 31115441 | T | C | 0.461 | -0.047 | 1.65E-13 | 0.006 | T | C | 0.455 | -0.004 | 0.002 | 0.047 |
| rs3095248 | 6 | 31209288 | T | C | 0.415 | -0.038 | 6.95E-09 | 0.007 | T | C | 0.388 | 0.002 | 0.002 | 0.56 |
| rs35840219 | 6 | 31249267 | C | G | 0.096 | 0.063 | 2.08E-08 | 0.011 | G | C | 0.922 | 0.005 | 0.003 | 0.11 |
| rs9266000 | 6 | 31316651 | T | C | 0.479 | -0.039 | 7.59E-10 | 0.006 | T | C | 0.455 | 0.006 | 0.002 | 0.012 |
| rs3135041 | 6 | 31555657 | A | G | 0.677 | -0.041 | 2.67E-09 | 0.007 | A | G | 0.656 | 0.000 | 0.002 | 0.67 |
| rs2857609 | 6 | 31577825 | A | G | 0.163 | -0.071 | 1.09E-15 | 0.009 | A | G | 0.161 | -0.001 | 0.002 | 0.66 |
| rs115884658 | 6 | 31864538 | A | G | 0.029 | 0.126 | 1.22E-09 | 0.021 | G | A | 0.980 | 0.002 | 0.007 | 0.48 |
| rs3130342 | 6 | 32080146 | A | C | 0.153 | -0.071 | 3.44E-15 | 0.009 | A | C | 0.151 | 0.002 | 0.003 | 0.62 |
| rs915894 | 6 | 32190390 | T | G | 0.635 | -0.041 | 1.37E-09 | 0.007 | T | G | 0.603 | -0.002 | 0.002 | 0.71 |
| rs3806155 | 6 | 32373378 | A | T | 0.964 | -0.169 | 2.74E-20 | 0.018 | A | T | 0.965 | 0.006 | 0.005 | 0.26 |
| rs601945 | 6 | 32573415 | A | G | 0.826 | -0.085 | 4.66E-23 | 0.009 | A | G | 0.808 | 0.009 | 0.002 | 0.00036 |
| rs17843580 | 6 | 32615551 | A | G | 0.449 | -0.053 | 1.34E-13 | 0.007 | A | G | 0.398 | 0.001 | 0.002 | 0.91 |
| rs9274407 | 6 | 32632832 | A | T | 0.196 | -0.073 | 4.46E-17 | 0.009 | A | T | 0.170 | 0.001 | 0.002 | 0.57 |
| rs6457684 | 6 | 32814447 | T | C | 0.574 | -0.037 | 1.31E-08 | 0.007 | T | C | 0.563 | 0.000 | 0.002 | 0.66 |
| rs11967262 | 6 | 43760327 | C | G | 0.514 | -0.038 | 5.78E-09 | 0.007 | C | G | 0.512 | -0.013 | 0.002 | 4.70E-10 |
| rs6937438 | 6 | 43815364 | A | G | 0.711 | -0.051 | 2.35E-13 | 0.007 | G | A | 0.292 | 0.001 | 0.002 | 0.37 |
| rs3798519 | 6 | 50788778 | A | C | 0.811 | -0.062 | 4.60E-14 | 0.008 | A | C | 0.820 | 0.000 | 0.002 | 0.59 |
| rs1665901 | 6 | 107433400 | A | T | 0.658 | 0.040 | 4.97E-09 | 0.007 | A | T | 0.662 | 0.000 | 0.002 | 0.99 |
| rs11759026 | 6 | 126792095 | A | G | 0.768 | -0.066 | 2.04E-18 | 0.008 | A | G | 0.773 | -0.005 | 0.002 | 0.058 |
| rs4580892 | 6 | 127409882 | T | C | 0.293 | 0.040 | 8.46E-09 | 0.007 | C | T | 0.720 | -0.048 | 0.002 | 1.50E-96 |
| rs719727 | 6 | 127414838 | A | G | 0.758 | 0.059 | 1.67E-15 | 0.007 | A | G | 0.750 | -0.035 | 0.002 | 1.90E-53 |
| rs6557267 | 6 | 153433701 | T | C | 0.420 | 0.036 | 2.93E-08 | 0.007 | C | T | 0.599 | -0.003 | 0.002 | 0.13 |
| rs474513 | 6 | 160770312 | A | G | 0.516 | 0.040 | 4.18E-10 | 0.006 | A | G | 0.525 | 0.001 | 0.002 | 0.59 |
| rs4709746 | 6 | 164133001 | T | C | 0.131 | -0.057 | 3.95E-09 | 0.010 | C | T | 0.866 | 0.032 | 0.003 | 1.40E-26 |
| rs17168486 | 7 | 14898282 | T | C | 0.180 | 0.067 | 4.50E-16 | 0.008 | C | T | 0.826 | 0.001 | 0.002 | 0.64 |
| rs2215383 | 7 | 15062983 | T | C | 0.463 | -0.064 | 1.06E-23 | 0.006 | T | C | 0.451 | 0.008 | 0.002 | 2.00E-04 |
| rs62451130 | 7 | 27977174 | T | C | 0.092 | -0.065 | 5.30E-09 | 0.011 | C | T | 0.915 | 0.010 | 0.003 | 0.0068 |
| rs10225433 | 7 | 28191672 | A | G | 0.067 | -0.077 | 2.86E-08 | 0.014 | G | A | 0.935 | -0.002 | 0.004 | 0.63 |
| rs1708302 | 7 | 28198677 | T | C | 0.489 | -0.091 | 5.77E-46 | 0.006 | C | T | 0.499 | -0.006 | 0.002 | 0.0062 |
| rs73300665 | 7 | 28227024 | A | G | 0.107 | 0.059 | 1.06E-08 | 0.010 | G | A | 0.890 | 0.009 | 0.003 | 0.0091 |
| rs917195 | 7 | 30728452 | T | C | 0.229 | -0.047 | 1.03E-09 | 0.008 | C | T | 0.769 | 0.003 | 0.002 | 0.1 |
| rs2268574 | 7 | 44189321 | A | G | 0.510 | -0.038 | 2.01E-09 | 0.006 | A | G | 0.516 | 0.000 | 0.002 | 0.91 |
| rs878521 | 7 | 44255643 | A | G | 0.243 | 0.062 | 9.31E-17 | 0.007 | G | A | 0.746 | 0.003 | 0.002 | 0.13 |
| rs77655131 | 7 | 102086552 | T | C | 0.130 | 0.055 | 1.27E-08 | 0.010 | C | T | 0.874 | -0.003 | 0.003 | 0.32 |
| rs11496066 | 7 | 102486254 | T | C | 0.817 | 0.051 | 8.17E-10 | 0.008 | T | C | 0.815 | -0.001 | 0.002 | 0.88 |
| rs1562396 | 7 | 130457914 | A | G | 0.680 | -0.056 | 9.64E-16 | 0.007 | A | G | 0.680 | 0.006 | 0.002 | 0.0044 |
| rs62492368 | 7 | 150537635 | A | G | 0.312 | 0.044 | 1.92E-10 | 0.007 | G | A | 0.697 | -0.007 | 0.002 | 0.0022 |
| rs6459737 | 7 | 156975136 | A | G | 0.338 | -0.058 | 3.31E-18 | 0.007 | G | A | 0.652 | -0.004 | 0.002 | 0.37 |
| rs2921077 | 8 | 8304502 | A | G | 0.462 | 0.037 | 9.14E-09 | 0.007 | G | A | 0.536 | -0.036 | 0.002 | 1.30E-64 |
| rs11774915 | 8 | 9188762 | T | C | 0.333 | 0.044 | 1.49E-10 | 0.007 | C | T | 0.660 | -0.030 | 0.002 | 5.20E-36 |
| rs17662402 | 8 | 9265105 | T | C | 0.944 | 0.084 | 4.52E-09 | 0.014 | T | C | 0.939 | 0.012 | 0.004 | 0.012 |
| rs34990153 | 8 | 9996389 | A | G | 0.560 | 0.048 | 2.26E-13 | 0.007 | A | G | 0.558 | 0.025 | 0.002 | 2.10E-31 |
| rs56408109 | 8 | 10009087 | T | G | 0.051 | -0.086 | 3.77E-08 | 0.016 | G | T | 0.948 | 0.012 | 0.004 | 0.028 |
| rs2001433 | 8 | 10903475 | A | T | 0.526 | -0.044 | 9.84E-12 | 0.007 | T | A | 0.480 | 0.036 | 0.002 | 2.40E-60 |
| rs2244648 | 8 | 11450422 | A | G | 0.537 | -0.036 | 2.25E-08 | 0.007 | A | G | 0.533 | -0.038 | 0.002 | 1.30E-66 |
| rs7819706 | 8 | 19844415 | A | G | 0.884 | 0.067 | 2.82E-11 | 0.010 | A | G | 0.882 | 0.006 | 0.003 | 0.07 |
| rs1060731 | 8 | 41435225 | T | C | 0.292 | 0.041 | 3.60E-09 | 0.007 | C | T | 0.709 | 0.001 | 0.002 | 0.8 |
| rs59191643 | 8 | 41500861 | T | C | 0.628 | 0.050 | 5.50E-14 | 0.007 | C | T | 0.382 | 0.003 | 0.002 | 0.43 |
| rs508419 | 8 | 41522991 | A | G | 0.236 | -0.081 | 5.43E-27 | 0.008 | A | G | 0.241 | -0.001 | 0.002 | 0.49 |
| rs80105613 | 8 | 41529789 | T | C | 0.037 | 0.107 | 6.44E-10 | 0.017 | C | T | 0.964 | 0.005 | 0.005 | 0.29 |
| rs67763258 | 8 | 95739642 | T | G | 0.323 | -0.039 | 1.30E-08 | 0.007 | G | T | 0.658 | -0.001 | 0.002 | 0.81 |
| rs10097617 | 8 | 95961626 | T | C | 0.485 | 0.049 | 2.44E-14 | 0.006 | T | C | 0.470 | -0.002 | 0.002 | 0.49 |
| rs76471882 | 8 | 96128981 | T | C | 0.047 | 0.088 | 1.31E-08 | 0.015 | C | T | 0.961 | -0.007 | 0.005 | 0.18 |
| rs3808415 | 8 | 116482423 | A | G | 0.558 | -0.035 | 4.65E-08 | 0.006 | A | G | 0.549 | -0.010 | 0.002 | 5.20E-06 |
| rs3802177 | 8 | 118185025 | A | G | 0.315 | -0.108 | 9.16E-55 | 0.007 | G | A | 0.689 | 0.004 | 0.002 | 0.27 |
| rs16889471 | 8 | 118190620 | A | G | 0.214 | 0.045 | 7.38E-09 | 0.008 | G | A | 0.780 | -0.002 | 0.002 | 0.35 |
| rs1561927 | 8 | 129568078 | T | C | 0.733 | -0.042 | 6.12E-09 | 0.007 | C | T | 0.268 | 0.001 | 0.002 | 0.87 |
| rs4977213 | 8 | 145507304 | T | C | 0.628 | -0.051 | 2.19E-13 | 0.007 | C | T | 0.381 | 0.008 | 0.002 | 2.30E-06 |
| rs2272662 | 8 | 145639726 | T | C | 0.412 | -0.040 | 2.63E-08 | 0.007 | T | C | 0.422 | -0.005 | 0.002 | 0.035 |
| rs12719778 | 8 | 145879883 | T | C | 0.537 | 0.038 | 4.38E-09 | 0.007 | T | C | 0.536 | 0.000 | 0.002 | 0.9 |
| rs672271 | 9 | 3273781 | T | C | 0.906 | -0.060 | 4.83E-08 | 0.011 | C | T | 0.089 | 0.003 | 0.003 | 0.33 |
| rs10974438 | 9 | 4291928 | A | C | 0.642 | -0.051 | 1.71E-14 | 0.007 | A | C | 0.650 | 0.002 | 0.002 | 0.12 |
| rs62563593 | 9 | 19065825 | A | G | 0.606 | -0.039 | 1.28E-09 | 0.007 | A | G | 0.608 | 0.003 | 0.002 | 0.15 |
| rs10965199 | 9 | 21954653 | T | C | 0.036 | -0.114 | 1.85E-09 | 0.019 | C | T | 0.958 | 0.003 | 0.005 | 0.34 |
| rs1063192 | 9 | 22003367 | A | G | 0.562 | 0.058 | 8.17E-19 | 0.007 | G | A | 0.445 | -0.001 | 0.002 | 0.44 |
| rs17694555 | 9 | 22051295 | A | G | 0.908 | -0.061 | 2.34E-08 | 0.011 | A | G | 0.913 | 0.001 | 0.003 | 0.77 |
| rs76011118 | 9 | 22133773 | A | G | 0.034 | 0.187 | 7.44E-22 | 0.020 | G | A | 0.967 | 0.001 | 0.006 | 0.64 |
| rs10811660 | 9 | 22134068 | A | G | 0.170 | -0.160 | 2.54E-77 | 0.009 | G | A | 0.827 | -0.004 | 0.002 | 0.061 |
| rs12555274 | 9 | 22136440 | C | G | 0.265 | 0.107 | 1.88E-49 | 0.007 | G | C | 0.749 | -0.005 | 0.002 | 0.023 |
| rs72655474 | 9 | 22139684 | C | G | 0.022 | -0.191 | 2.45E-14 | 0.025 | G | C | 0.977 | 0.014 | 0.007 | 0.19 |
| rs1412234 | 9 | 28410683 | T | C | 0.674 | -0.039 | 7.71E-09 | 0.007 | T | C | 0.670 | 0.000 | 0.002 | 0.77 |
| rs12001437 | 9 | 34074476 | T | C | 0.631 | -0.040 | 1.10E-09 | 0.007 | T | C | 0.634 | -0.010 | 0.002 | 1.90E-08 |
| rs17791513 | 9 | 81905590 | A | G | 0.932 | 0.102 | 1.35E-14 | 0.013 | A | G | 0.938 | 0.011 | 0.004 | 0.023 |
| rs2796441 | 9 | 84308948 | A | G | 0.406 | -0.067 | 2.97E-25 | 0.007 | G | A | 0.580 | -0.003 | 0.002 | 0.02 |
| rs55653563 | 9 | 97001682 | A | C | 0.733 | 0.044 | 1.73E-09 | 0.007 | A | C | 0.736 | 0.009 | 0.002 | 4.90E-06 |
| rs505922 | 9 | 136149229 | T | C | 0.663 | -0.047 | 3.65E-12 | 0.007 | T | C | 0.684 | 0.017 | 0.002 | 4.40E-13 |
| rs28533815 | 9 | 139241828 | T | C | 0.251 | -0.075 | 5.45E-21 | 0.008 | T | C | 0.244 | 0.000 | 0.002 | 0.73 |
| rs1127152 | 9 | 139335599 | A | G | 0.568 | 0.036 | 3.49E-08 | 0.007 | A | G | 0.553 | 0.001 | 0.002 | 0.95 |
| rs11257655 | 10 | 12307894 | T | C | 0.217 | 0.086 | 1.46E-28 | 0.008 | C | T | 0.792 | 0.004 | 0.002 | 0.077 |
| rs2616055 | 10 | 71317835 | A | G | 0.940 | 0.080 | 5.70E-09 | 0.014 | G | A | 0.056 | 0.005 | 0.004 | 0.17 |
| rs177045 | 10 | 71321279 | A | G | 0.683 | -0.050 | 4.64E-13 | 0.007 | A | G | 0.687 | 0.004 | 0.002 | 0.34 |
| rs41277236 | 10 | 71332301 | T | C | 0.043 | 0.100 | 2.47E-09 | 0.017 | C | T | 0.957 | 0.003 | 0.005 | 0.27 |
| rs2812539 | 10 | 71465359 | T | G | 0.702 | 0.051 | 3.59E-13 | 0.007 | G | T | 0.302 | 0.001 | 0.002 | 0.34 |
| rs11591689 | 10 | 80907740 | T | C | 0.233 | 0.049 | 3.55E-10 | 0.008 | C | T | 0.762 | -0.002 | 0.002 | 0.56 |
| rs703972 | 10 | 80952826 | C | G | 0.468 | -0.070 | 5.77E-27 | 0.007 | G | C | 0.539 | 0.002 | 0.002 | 0.14 |
| rs76087804 | 10 | 80973926 | A | G | 0.174 | -0.061 | 1.83E-12 | 0.009 | G | A | 0.827 | 0.003 | 0.002 | 0.27 |
| rs1574190 | 10 | 80992381 | T | C | 0.392 | 0.048 | 1.22E-12 | 0.007 | C | T | 0.587 | 0.001 | 0.002 | 0.75 |
| rs41287646 | 10 | 93749380 | T | G | 0.914 | -0.077 | 4.12E-11 | 0.012 | T | G | 0.910 | -0.007 | 0.003 | 0.08 |
| rs7903767 | 10 | 94138239 | A | G | 0.540 | -0.055 | 7.21E-18 | 0.006 | A | G | 0.533 | -0.006 | 0.002 | 0.0037 |
| rs2249960 | 10 | 94233120 | A | G | 0.866 | -0.058 | 9.43E-10 | 0.010 | G | A | 0.136 | 0.003 | 0.003 | 0.65 |
| rs17875327 | 10 | 94274809 | A | G | 0.882 | -0.059 | 3.87E-09 | 0.010 | A | G | 0.894 | -0.005 | 0.003 | 0.1 |
| rs77014180 | 10 | 94307157 | A | G | 0.940 | -0.086 | 1.67E-10 | 0.014 | A | G | 0.938 | -0.006 | 0.004 | 0.13 |
| rs10882099 | 10 | 94460650 | T | C | 0.588 | 0.110 | 7.73E-64 | 0.007 | T | C | 0.591 | 0.001 | 0.002 | 0.76 |
| rs146935743 | 10 | 94466064 | T | C | 0.050 | -0.105 | 4.95E-11 | 0.016 | C | T | 0.947 | 0.005 | 0.004 | 0.27 |
| rs11187152 | 10 | 94500111 | A | G | 0.079 | -0.070 | 1.85E-08 | 0.012 | G | A | 0.923 | -0.001 | 0.004 | 0.59 |
| rs145220772 | 10 | 114578143 | T | C | 0.021 | 0.132 | 2.74E-08 | 0.024 | C | T | 0.977 | -0.010 | 0.007 | 0.27 |
| rs545572 | 10 | 114597109 | A | G | 0.393 | -0.076 | 4.24E-29 | 0.007 | G | A | 0.603 | 0.005 | 0.002 | 0.011 |
| rs1885283 | 10 | 114648499 | A | C | 0.130 | 0.073 | 8.17E-13 | 0.010 | C | A | 0.867 | 0.006 | 0.003 | 0.23 |
| rs139688524 | 10 | 114668724 | T | C | 0.023 | 0.174 | 7.47E-16 | 0.022 | C | T | 0.978 | 0.013 | 0.006 | 0.28 |
| rs11196148 | 10 | 114671172 | T | C | 0.176 | -0.060 | 4.29E-12 | 0.009 | C | T | 0.826 | 0.003 | 0.002 | 0.065 |
| rs116425039 | 10 | 114681965 | A | G | 0.012 | -0.272 | 5.08E-15 | 0.035 | G | A | 0.992 | -0.005 | 0.010 | 0.54 |
| rs10885397 | 10 | 114711883 | A | G | 0.154 | -0.074 | 6.49E-16 | 0.009 | G | A | 0.847 | 0.013 | 0.003 | 7.30E-07 |
| rs2094406 | 10 | 114714725 | T | C | 0.265 | 0.050 | 4.07E-12 | 0.007 | C | T | 0.734 | -0.007 | 0.002 | 0.0029 |
| rs61875108 | 10 | 114725079 | A | G | 0.049 | -0.093 | 1.95E-09 | 0.016 | G | A | 0.949 | 0.020 | 0.004 | 5.30E-05 |
| rs117736037 | 10 | 114735110 | A | G | 0.015 | 0.251 | 4.42E-19 | 0.028 | G | A | 0.985 | 0.033 | 0.008 | 0.00012 |
| rs146262265 | 10 | 114738287 | T | C | 0.009 | 0.226 | 1.58E-08 | 0.040 | C | T | 0.994 | 0.001 | 0.013 | 0.77 |
| rs116859590 | 10 | 114752410 | T | C | 0.026 | 0.229 | 1.42E-27 | 0.021 | C | T | 0.974 | -0.014 | 0.006 | 0.02 |
| rs34872471 | 10 | 114754071 | T | C | 0.708 | -0.313 | 0 | 0.007 | T | C | 0.709 | -0.011 | 0.002 | 1.10E-08 |
| rs117987174 | 10 | 114756459 | A | G | 0.014 | -0.172 | 1.56E-08 | 0.031 | G | A | 0.987 | -0.003 | 0.008 | 0.64 |
| rs78025551 | 10 | 114757956 | C | G | 0.847 | 0.149 | 2.25E-60 | 0.009 | C | G | 0.855 | 0.008 | 0.003 | 0.0032 |
| rs61872774 | 10 | 114765390 | A | G | 0.012 | 0.288 | 4.85E-22 | 0.030 | G | A | 0.987 | -0.006 | 0.008 | 0.19 |
| rs140908036 | 10 | 114770644 | A | G | 0.015 | 0.289 | 3.78E-24 | 0.029 | G | A | 0.986 | -0.015 | 0.008 | 0.17 |
| rs141356057 | 10 | 114770860 | T | C | 0.011 | -0.191 | 1.40E-08 | 0.034 | C | T | 0.989 | 0.026 | 0.009 | 0.062 |
| rs10885404 | 10 | 114773068 | T | G | 0.177 | -0.107 | 4.10E-34 | 0.009 | G | T | 0.824 | 0.007 | 0.002 | 0.0015 |
| rs141241414 | 10 | 114775551 | A | G | 0.986 | 0.195 | 6.80E-12 | 0.028 | A | G | 0.985 | -0.003 | 0.008 | 0.21 |
| rs180726800 | 10 | 114784926 | T | C | 0.019 | 0.252 | 2.19E-24 | 0.025 | C | T | 0.984 | -0.007 | 0.008 | 0.16 |
| rs140820620 | 10 | 114787948 | A | G | 0.022 | 0.284 | 7.08E-36 | 0.023 | G | A | 0.976 | -0.015 | 0.006 | 0.021 |
| rs116369954 | 10 | 114793572 | T | C | 0.967 | -0.292 | 8.91E-58 | 0.018 | T | C | 0.968 | -0.020 | 0.005 | 0.00011 |
| rs185025714 | 10 | 114794451 | T | C | 0.995 | -0.264 | 3.10E-08 | 0.048 | T | C | 0.995 | -0.038 | 0.013 | 0.0072 |
| rs11196201 | 10 | 114803307 | A | T | 0.922 | -0.185 | 2.49E-53 | 0.012 | A | T | 0.921 | -0.002 | 0.003 | 0.39 |
| rs72826101 | 10 | 114815466 | T | G | 0.935 | 0.109 | 3.91E-14 | 0.014 | T | G | 0.938 | 0.000 | 0.004 | 0.6 |
| rs10885414 | 10 | 114861304 | A | G | 0.686 | 0.095 | 1.07E-41 | 0.007 | A | G | 0.688 | 0.007 | 0.002 | 0.005 |
| rs10749128 | 10 | 114865489 | T | C | 0.748 | -0.046 | 6.10E-10 | 0.007 | C | T | 0.261 | 0.005 | 0.002 | 0.0025 |
| rs189966089 | 10 | 114886341 | T | C | 0.029 | -0.128 | 6.17E-09 | 0.022 | C | T | 0.979 | 0.003 | 0.007 | 0.46 |
| rs11196236 | 10 | 114887722 | T | C | 0.790 | -0.074 | 4.92E-21 | 0.008 | T | C | 0.790 | 0.004 | 0.002 | 0.11 |
| rs290483 | 10 | 114915214 | T | G | 0.602 | 0.070 | 1.30E-25 | 0.007 | G | T | 0.402 | -0.006 | 0.002 | 0.01 |
| rs72828149 | 10 | 114942869 | A | G | 0.018 | 0.148 | 2.31E-08 | 0.027 | G | A | 0.981 | -0.002 | 0.007 | 0.43 |
| rs2280141 | 10 | 124193181 | T | G | 0.512 | 0.049 | 1.91E-14 | 0.006 | T | G | 0.529 | 0.004 | 0.002 | 0.1 |
| rs76074250 | 11 | 2073182 | A | G | 0.797 | -0.046 | 9.31E-09 | 0.008 | A | G | 0.797 | -0.008 | 0.002 | 0.0016 |
| rs3842749 | 11 | 2181338 | A | G | 0.059 | 0.086 | 1.69E-09 | 0.014 | G | A | 0.939 | 0.000 | 0.004 | 0.63 |
| rs4929965 | 11 | 2197286 | A | G | 0.381 | 0.067 | 9.67E-23 | 0.007 | A | G | 0.381 | 0.004 | 0.002 | 0.12 |
| rs11023461 | 11 | 2632430 | C | G | 0.924 | -0.075 | 5.49E-09 | 0.013 | C | G | 0.928 | 0.001 | 0.004 | 0.76 |
| rs231360 | 11 | 2692249 | T | C | 0.396 | 0.058 | 8.27E-18 | 0.007 | C | T | 0.607 | 0.001 | 0.002 | 0.5 |
| rs11023937 | 11 | 2755573 | C | G | 0.680 | 0.039 | 3.68E-08 | 0.007 | C | G | 0.691 | 0.000 | 0.002 | 0.55 |
| rs2237890 | 11 | 2835757 | T | C | 0.066 | -0.080 | 1.04E-08 | 0.014 | C | T | 0.931 | 0.005 | 0.004 | 0.12 |
| rs7480855 | 11 | 2836085 | A | G | 0.933 | 0.101 | 3.65E-14 | 0.013 | A | G | 0.934 | -0.003 | 0.004 | 0.57 |
| rs2237895 | 11 | 2857194 | A | C | 0.573 | -0.089 | 1.15E-41 | 0.007 | A | C | 0.584 | 0.000 | 0.002 | 0.8 |
| rs141521721 | 11 | 14763828 | A | C | 0.023 | 0.121 | 1.39E-08 | 0.021 | C | A | 0.976 | -0.008 | 0.006 | 0.33 |
| rs5215 | 11 | 17408630 | T | C | 0.627 | -0.071 | 9.85E-27 | 0.007 | C | T | 0.358 | 0.001 | 0.002 | 0.55 |
| rs145678014 | 11 | 32927778 | T | G | 0.043 | -0.105 | 1.40E-10 | 0.016 | G | T | 0.955 | -0.006 | 0.004 | 0.43 |
| rs10047488 | 11 | 34782972 | C | G | 0.082 | 0.064 | 4.66E-08 | 0.012 | G | C | 0.918 | 0.000 | 0.003 | 0.6 |
| rs2767036 | 11 | 34982148 | A | C | 0.709 | -0.039 | 2.94E-08 | 0.007 | C | A | 0.286 | -0.002 | 0.002 | 0.31 |
| rs1061810 | 11 | 43877934 | A | C | 0.287 | 0.050 | 8.31E-13 | 0.007 | C | A | 0.709 | 0.000 | 0.002 | 0.79 |
| rs12419690 | 11 | 45858584 | A | G | 0.552 | -0.035 | 3.89E-08 | 0.006 | G | A | 0.437 | -0.003 | 0.002 | 0.17 |
| rs7124681 | 11 | 47529947 | A | C | 0.411 | 0.036 | 2.45E-08 | 0.007 | C | A | 0.591 | 0.001 | 0.002 | 0.93 |
| rs1783541 | 11 | 65294799 | T | C | 0.201 | 0.061 | 4.65E-14 | 0.008 | C | T | 0.782 | -0.026 | 0.002 | 2.20E-23 |
| rs55911137 | 11 | 69456000 | C | G | 0.026 | -0.145 | 9.94E-12 | 0.021 | G | C | 0.975 | 0.027 | 0.006 | 7.50E-05 |
| rs77464186 | 11 | 72460398 | A | C | 0.836 | 0.100 | 9.30E-31 | 0.009 | A | C | 0.843 | 0.009 | 0.003 | 0.0063 |
| rs191142560 | 11 | 92707806 | T | C | 0.018 | 0.143 | 2.70E-08 | 0.026 | C | T | 0.981 | -0.007 | 0.007 | 0.31 |
| rs10830963 | 11 | 92708710 | C | G | 0.721 | -0.101 | 1.12E-45 | 0.007 | C | G | 0.725 | -0.009 | 0.002 | 0.00027 |
| rs9971402 | 11 | 92756551 | A | C | 0.144 | 0.053 | 6.78E-09 | 0.009 | C | A | 0.859 | -0.002 | 0.003 | 0.29 |
| rs10444213 | 11 | 93012957 | A | T | 0.539 | 0.042 | 1.09E-10 | 0.007 | T | A | 0.452 | -0.003 | 0.002 | 0.22 |
| rs7933438 | 11 | 128041582 | A | G | 0.147 | -0.058 | 4.84E-10 | 0.009 | G | A | 0.857 | -0.001 | 0.003 | 0.93 |
| rs10750397 | 11 | 128234144 | A | G | 0.283 | 0.039 | 3.13E-08 | 0.007 | A | G | 0.279 | -0.004 | 0.002 | 0.023 |
| rs67232546 | 11 | 128398938 | T | C | 0.205 | 0.053 | 2.51E-11 | 0.008 | C | T | 0.788 | -0.002 | 0.002 | 0.64 |
| rs10848958 | 12 | 4031104 | T | C | 0.197 | -0.045 | 4.97E-08 | 0.008 | C | T | 0.799 | -0.001 | 0.002 | 0.27 |
| rs141892016 | 12 | 4200707 | A | G | 0.992 | 0.235 | 4.84E-09 | 0.040 | A | G | 0.992 | 0.039 | 0.011 | 0.0041 |
| rs11063029 | 12 | 4301301 | T | C | 0.058 | 0.086 | 5.36E-10 | 0.014 | C | T | 0.949 | 0.001 | 0.004 | 0.85 |
| rs11063069 | 12 | 4374373 | A | G | 0.787 | -0.056 | 9.94E-13 | 0.008 | A | G | 0.793 | -0.003 | 0.002 | 0.53 |
| rs12818766 | 12 | 4376091 | A | G | 0.168 | -0.054 | 1.49E-09 | 0.009 | G | A | 0.837 | 0.003 | 0.003 | 0.23 |
| rs3217792 | 12 | 4384696 | T | C | 0.087 | -0.117 | 6.27E-22 | 0.012 | C | T | 0.915 | -0.002 | 0.003 | 0.95 |
| rs76895963 | 12 | 4384844 | T | G | 0.980 | 0.483 | 9.12E-69 | 0.028 | T | G | 0.979 | 0.077 | 0.007 | 9.00E-25 |
| rs3217860 | 12 | 4399050 | A | G | 0.738 | -0.054 | 5.59E-13 | 0.007 | A | G | 0.752 | -0.009 | 0.002 | 9.30E-05 |
| rs12299509 | 12 | 4406281 | A | G | 0.530 | -0.050 | 3.15E-13 | 0.007 | A | G | 0.530 | -0.007 | 0.002 | 0.0019 |
| rs2066827 | 12 | 12871099 | T | G | 0.768 | -0.047 | 4.44E-09 | 0.008 | T | G | 0.768 | -0.004 | 0.002 | 0.065 |
| rs1872992 | 12 | 26457190 | A | G | 0.741 | -0.046 | 2.52E-10 | 0.007 | A | G | 0.757 | -0.028 | 0.002 | 4.80E-28 |
| rs2052673 | 12 | 27826780 | T | G | 0.170 | -0.056 | 3.88E-10 | 0.009 | T | G | 0.174 | -0.005 | 0.002 | 0.013 |
| rs7969720 | 12 | 27905210 | A | G | 0.684 | -0.038 | 2.98E-08 | 0.007 | G | A | 0.315 | -0.002 | 0.002 | 0.29 |
| rs7966976 | 12 | 27962934 | A | G | 0.196 | -0.075 | 1.22E-20 | 0.008 | G | A | 0.803 | 0.004 | 0.002 | 0.11 |
| rs2258238 | 12 | 66221060 | A | T | 0.896 | -0.102 | 5.14E-22 | 0.011 | A | T | 0.895 | -0.002 | 0.003 | 0.53 |
| rs7968682 | 12 | 66371880 | T | G | 0.507 | 0.054 | 3.65E-17 | 0.006 | G | T | 0.485 | -0.014 | 0.002 | 4.10E-11 |
| rs1705263 | 12 | 71523043 | A | C | 0.437 | -0.048 | 3.51E-14 | 0.006 | C | A | 0.559 | 0.004 | 0.002 | 0.17 |
| rs61939481 | 12 | 95921998 | T | C | 0.932 | -0.071 | 1.74E-08 | 0.013 | T | C | 0.929 | 0.009 | 0.004 | 0.0018 |
| rs77864822 | 12 | 97848775 | A | G | 0.930 | 0.075 | 5.01E-09 | 0.013 | A | G | 0.934 | 0.002 | 0.004 | 0.83 |
| rs1426371 | 12 | 108629780 | A | G | 0.266 | -0.052 | 1.74E-12 | 0.007 | G | A | 0.740 | 0.005 | 0.002 | 0.37 |
| rs7313918 | 12 | 118394008 | T | C | 0.864 | -0.056 | 2.99E-09 | 0.009 | T | C | 0.873 | -0.001 | 0.003 | 0.8 |
| rs625228 | 12 | 121278266 | A | G | 0.442 | -0.036 | 1.90E-08 | 0.006 | G | A | 0.565 | -0.003 | 0.002 | 0.17 |
| rs11065299 | 12 | 121297815 | A | G | 0.076 | 0.079 | 5.97E-11 | 0.012 | G | A | 0.925 | 0.007 | 0.004 | 0.048 |
| rs73226260 | 12 | 121380541 | A | G | 0.033 | -0.121 | 1.36E-10 | 0.019 | G | A | 0.966 | -0.004 | 0.005 | 0.49 |
| rs1800574 | 12 | 121416864 | T | C | 0.029 | 0.151 | 2.75E-15 | 0.019 | C | T | 0.971 | -0.007 | 0.005 | 0.35 |
| rs56348580 | 12 | 121432117 | C | G | 0.308 | -0.062 | 4.31E-19 | 0.007 | G | C | 0.692 | 0.005 | 0.002 | 0.038 |
| rs2259883 | 12 | 121462139 | C | G | 0.539 | 0.042 | 6.62E-11 | 0.006 | G | C | 0.452 | -0.001 | 0.002 | 0.52 |
| rs28638142 | 12 | 121501461 | A | C | 0.046 | 0.092 | 4.87E-09 | 0.016 | C | A | 0.959 | -0.005 | 0.005 | 0.52 |
| rs28360442 | 12 | 121570940 | C | G | 0.039 | -0.106 | 5.39E-09 | 0.018 | G | C | 0.965 | 0.001 | 0.005 | 0.82 |
| rs208302 | 12 | 121602760 | A | G | 0.228 | 0.041 | 3.98E-08 | 0.008 | G | A | 0.765 | 0.003 | 0.002 | 0.18 |
| rs4148856 | 12 | 123450765 | C | G | 0.783 | 0.047 | 1.31E-09 | 0.008 | G | C | 0.212 | -0.016 | 0.002 | 4.10E-09 |
| rs10773051 | 12 | 124510391 | T | C | 0.235 | -0.041 | 4.98E-08 | 0.008 | C | T | 0.780 | -0.019 | 0.002 | 6.30E-13 |
| rs35318451 | 12 | 133068484 | A | G | 0.331 | 0.048 | 9.50E-12 | 0.007 | G | A | 0.671 | -0.002 | 0.002 | 0.53 |
| rs34584161 | 13 | 26776999 | A | G | 0.761 | 0.047 | 5.36E-10 | 0.008 | A | G | 0.764 | -0.001 | 0.002 | 0.63 |
| rs11842871 | 13 | 31042452 | T | G | 0.267 | -0.040 | 4.83E-08 | 0.007 | G | T | 0.738 | -0.002 | 0.002 | 0.5 |
| rs576674 | 13 | 33554302 | A | G | 0.831 | -0.054 | 3.70E-10 | 0.009 | G | A | 0.167 | -0.001 | 0.002 | 0.22 |
| rs9316500 | 13 | 51094114 | T | G | 0.712 | 0.039 | 3.45E-08 | 0.007 | T | G | 0.706 | 0.014 | 0.002 | 1.20E-11 |
| rs35559811 | 13 | 58666728 | A | G | 0.764 | 0.043 | 4.13E-08 | 0.008 | A | G | 0.770 | -0.001 | 0.002 | 0.76 |
| rs9563615 | 13 | 59077406 | A | T | 0.709 | 0.041 | 1.63E-08 | 0.007 | A | T | 0.704 | 0.006 | 0.002 | 0.0028 |
| rs1359790 | 13 | 80717156 | A | G | 0.279 | -0.082 | 1.76E-30 | 0.007 | G | A | 0.711 | 0.009 | 0.002 | 2.00E-04 |
| rs7325671 | 13 | 108797836 | T | C | 0.126 | 0.055 | 1.53E-08 | 0.010 | C | T | 0.870 | 0.004 | 0.003 | 0.039 |
| rs17122772 | 14 | 23288935 | C | G | 0.774 | -0.043 | 3.31E-08 | 0.008 | C | G | 0.770 | -0.001 | 0.002 | 0.97 |
| rs17522122 | 14 | 33302882 | T | G | 0.474 | 0.036 | 2.49E-08 | 0.006 | G | T | 0.529 | 0.002 | 0.002 | 0.17 |
| rs8008910 | 14 | 79944099 | A | G | 0.219 | 0.055 | 4.22E-13 | 0.008 | G | A | 0.776 | -0.007 | 0.002 | 0.082 |
| rs2896177 | 14 | 91968313 | A | G | 0.541 | -0.037 | 9.13E-09 | 0.006 | A | G | 0.549 | 0.003 | 0.002 | 0.24 |
| rs3783394 | 14 | 103858673 | A | G | 0.348 | -0.038 | 1.42E-08 | 0.007 | G | A | 0.657 | 0.042 | 0.002 | 9.50E-80 |
| rs8032939 | 15 | 38834033 | T | C | 0.753 | -0.042 | 1.88E-08 | 0.007 | T | C | 0.754 | 0.004 | 0.002 | 0.052 |
| rs34715063 | 15 | 38873115 | T | C | 0.878 | -0.077 | 8.40E-15 | 0.010 | T | C | 0.871 | -0.001 | 0.003 | 0.59 |
| rs2289739 | 15 | 41801512 | T | G | 0.356 | 0.047 | 2.58E-12 | 0.007 | G | T | 0.654 | 0.004 | 0.002 | 0.031 |
| rs2456530 | 15 | 53091553 | T | C | 0.126 | 0.054 | 2.07E-08 | 0.010 | C | T | 0.867 | 0.002 | 0.003 | 0.5 |
| rs144801310 | 15 | 57469927 | A | C | 0.966 | -0.101 | 4.01E-08 | 0.018 | A | C | 0.968 | 0.022 | 0.005 | 0.00063 |
| rs11856307 | 15 | 62399093 | A | C | 0.571 | 0.047 | 4.48E-13 | 0.007 | A | C | 0.578 | -0.004 | 0.002 | 0.024 |
| rs7178762 | 15 | 63871292 | T | C | 0.536 | -0.039 | 1.37E-09 | 0.006 | C | T | 0.449 | 0.009 | 0.002 | 1.00E-05 |
| rs4776970 | 15 | 68080886 | A | T | 0.642 | 0.038 | 1.69E-08 | 0.007 | A | T | 0.644 | 0.000 | 0.002 | 0.37 |
| rs13737 | 15 | 75932129 | T | G | 0.241 | -0.048 | 1.77E-10 | 0.008 | G | T | 0.750 | -0.005 | 0.002 | 0.12 |
| rs12904882 | 15 | 77401143 | A | C | 0.853 | -0.055 | 1.67E-09 | 0.009 | A | C | 0.852 | -0.002 | 0.003 | 0.71 |
| rs75145098 | 15 | 77714618 | T | G | 0.966 | 0.104 | 1.83E-08 | 0.018 | T | G | 0.963 | -0.011 | 0.005 | 0.075 |
| rs12910361 | 15 | 77782335 | A | G | 0.293 | -0.081 | 3.95E-31 | 0.007 | A | G | 0.286 | 0.004 | 0.002 | 0.054 |
| rs2351707 | 15 | 90384045 | T | C | 0.718 | -0.064 | 4.30E-20 | 0.007 | C | T | 0.282 | 0.004 | 0.002 | 0.078 |
| rs12910825 | 15 | 91511260 | A | G | 0.637 | -0.053 | 4.70E-15 | 0.007 | A | G | 0.643 | -0.003 | 0.002 | 0.19 |
| rs6600191 | 16 | 295795 | T | C | 0.825 | 0.059 | 4.47E-12 | 0.009 | T | C | 0.819 | -0.002 | 0.002 | 0.41 |
| rs12325539 | 16 | 30033633 | T | C | 0.600 | -0.041 | 2.69E-10 | 0.007 | T | C | 0.597 | 0.006 | 0.002 | 2.00E-04 |
| rs7203521 | 16 | 53769293 | A | G | 0.615 | 0.042 | 1.29E-10 | 0.007 | G | A | 0.390 | -0.005 | 0.002 | 0.025 |
| rs16952522 | 16 | 53807498 | C | G | 0.958 | -0.088 | 4.20E-08 | 0.016 | C | G | 0.962 | -0.017 | 0.005 | 0.011 |
| rs55872725 | 16 | 53809123 | T | C | 0.418 | 0.122 | 8.51E-79 | 0.007 | C | T | 0.598 | -0.019 | 0.002 | 1.10E-19 |
| rs12596054 | 16 | 53834684 | A | C | 0.857 | 0.059 | 2.02E-10 | 0.009 | A | C | 0.855 | 0.003 | 0.003 | 0.17 |
| rs6499646 | 16 | 53843533 | T | C | 0.922 | 0.066 | 3.89E-08 | 0.012 | T | C | 0.920 | 0.006 | 0.003 | 0.19 |
| rs139891432 | 16 | 53843623 | A | G | 0.029 | 0.134 | 9.58E-11 | 0.021 | G | A | 0.976 | -0.012 | 0.006 | 0.12 |
| rs6499653 | 16 | 53877592 | T | C | 0.255 | 0.043 | 4.44E-09 | 0.007 | T | C | 0.246 | 0.010 | 0.002 | 2.00E-05 |
| rs862320 | 16 | 69651866 | T | C | 0.423 | -0.043 | 2.55E-11 | 0.007 | C | T | 0.590 | -0.003 | 0.002 | 0.055 |
| rs72802358 | 16 | 75243657 | C | G | 0.103 | -0.114 | 2.22E-26 | 0.011 | G | C | 0.898 | 0.007 | 0.003 | 0.03 |
| rs2925979 | 16 | 81534790 | T | C | 0.299 | 0.055 | 7.07E-15 | 0.007 | T | C | 0.301 | -0.004 | 0.002 | 0.028 |
| rs12920022 | 16 | 89564055 | A | T | 0.158 | 0.053 | 1.00E-08 | 0.009 | T | A | 0.839 | -0.009 | 0.003 | 0.0028 |
| rs1043246 | 17 | 3828086 | C | G | 0.845 | -0.057 | 4.21E-09 | 0.010 | C | G | 0.841 | -0.005 | 0.003 | 0.088 |
| rs3826482 | 17 | 3860356 | A | T | 0.578 | 0.037 | 2.64E-08 | 0.007 | A | T | 0.583 | 0.002 | 0.002 | 0.12 |
| rs8071043 | 17 | 3988451 | T | C | 0.674 | -0.052 | 1.53E-14 | 0.007 | T | C | 0.669 | -0.011 | 0.002 | 2.00E-07 |
| rs55973554 | 17 | 9793756 | A | G | 0.322 | 0.039 | 1.00E-08 | 0.007 | G | A | 0.668 | -0.009 | 0.002 | 2.70E-05 |
| rs4925109 | 17 | 17661802 | A | G | 0.318 | 0.048 | 5.65E-12 | 0.007 | A | G | 0.302 | -0.009 | 0.002 | 0.00049 |
| rs2107133 | 17 | 36064897 | A | G | 0.873 | 0.064 | 4.02E-11 | 0.010 | A | G | 0.863 | 0.001 | 0.003 | 0.62 |
| rs10908278 | 17 | 36099952 | A | T | 0.517 | -0.075 | 5.25E-29 | 0.007 | T | A | 0.485 | 0.003 | 0.002 | 0.2 |
| rs34855406 | 17 | 40731411 | C | G | 0.275 | 0.049 | 5.89E-12 | 0.007 | G | C | 0.717 | -0.013 | 0.002 | 1.40E-09 |
| rs35895680 | 17 | 47060322 | A | C | 0.323 | -0.055 | 1.08E-15 | 0.007 | C | A | 0.673 | 0.003 | 0.002 | 0.043 |
| rs58642235 | 17 | 62202689 | T | C | 0.861 | -0.057 | 1.90E-09 | 0.009 | T | C | 0.860 | -0.001 | 0.003 | 0.53 |
| rs61676547 | 17 | 65892507 | C | G | 0.194 | 0.053 | 7.42E-11 | 0.008 | G | C | 0.811 | 0.009 | 0.002 | 0.00036 |
| rs7240767 | 18 | 7070642 | T | C | 0.626 | -0.037 | 1.71E-08 | 0.007 | T | C | 0.612 | 0.002 | 0.002 | 0.42 |
| rs1431841 | 18 | 40087098 | T | G | 0.211 | 0.043 | 3.08E-08 | 0.008 | G | T | 0.796 | -0.007 | 0.002 | 0.052 |
| rs72926982 | 18 | 53087984 | A | T | 0.085 | 0.074 | 8.93E-11 | 0.011 | T | A | 0.919 | -0.008 | 0.003 | 0.017 |
| rs17089932 | 18 | 53428347 | T | C | 0.249 | 0.041 | 2.81E-08 | 0.007 | C | T | 0.747 | 0.008 | 0.002 | 0.00078 |
| rs9957320 | 18 | 56876430 | T | G | 0.172 | -0.049 | 9.34E-09 | 0.009 | G | T | 0.834 | 0.008 | 0.002 | 0.025 |
| rs8097210 | 18 | 57857135 | T | G | 0.733 | -0.054 | 1.06E-13 | 0.007 | T | G | 0.733 | -0.016 | 0.002 | 2.00E-09 |
| rs111638368 | 18 | 57961249 | T | C | 0.305 | 0.039 | 3.98E-08 | 0.007 | C | T | 0.698 | -0.012 | 0.002 | 2.20E-07 |
| rs79688165 | 18 | 58050968 | T | C | 0.023 | -0.138 | 9.64E-10 | 0.023 | C | T | 0.980 | 0.004 | 0.007 | 0.83 |
| rs12454712 | 18 | 60845884 | T | C | 0.613 | 0.048 | 2.40E-12 | 0.007 | T | C | 0.623 | 0.004 | 0.002 | 0.23 |
| rs262549 | 19 | 4951064 | C | G | 0.807 | -0.047 | 6.48E-09 | 0.008 | C | G | 0.803 | -0.004 | 0.002 | 0.24 |
| rs4804833 | 19 | 7970635 | A | G | 0.391 | 0.048 | 3.42E-13 | 0.007 | A | G | 0.381 | -0.002 | 0.002 | 0.21 |
| rs10419627 | 19 | 13036677 | A | G | 0.591 | 0.043 | 3.15E-11 | 0.007 | G | A | 0.415 | -0.006 | 0.002 | 0.002 |
| rs739846 | 19 | 19419071 | A | G | 0.076 | 0.088 | 2.02E-13 | 0.012 | G | A | 0.924 | -0.008 | 0.004 | 0.044 |
| rs3786900 | 19 | 33897149 | A | G | 0.731 | 0.043 | 1.89E-09 | 0.007 | A | G | 0.731 | -0.001 | 0.002 | 0.47 |
| rs429358 | 19 | 45411941 | T | C | 0.847 | 0.075 | 2.62E-16 | 0.009 | T | C | 0.844 | -0.001 | 0.003 | 0.79 |
| rs10406431 | 19 | 46157019 | A | G | 0.563 | 0.060 | 1.56E-20 | 0.007 | A | G | 0.560 | -0.004 | 0.002 | 0.14 |
| rs2238689 | 19 | 46178661 | T | C | 0.579 | -0.049 | 8.73E-14 | 0.007 | T | C | 0.603 | 0.004 | 0.002 | 0.035 |
| rs3810291 | 19 | 47569003 | A | G | 0.672 | 0.043 | 3.38E-10 | 0.007 | G | A | 0.324 | 0.000 | 0.002 | 0.62 |
| rs17744783 | 20 | 21460433 | A | T | 0.107 | 0.058 | 1.87E-08 | 0.010 | T | A | 0.894 | -0.005 | 0.003 | 0.23 |
| rs1007090 | 20 | 32582871 | T | C | 0.332 | -0.044 | 1.01E-10 | 0.007 | T | C | 0.352 | -0.010 | 0.002 | 3.60E-05 |
| rs6031558 | 20 | 42999643 | C | G | 0.323 | -0.039 | 1.19E-08 | 0.007 | C | G | 0.331 | -0.001 | 0.002 | 0.19 |
| rs36112520 | 20 | 43003122 | A | G | 0.111 | 0.078 | 4.23E-14 | 0.010 | G | A | 0.904 | -0.003 | 0.003 | 0.59 |
| rs1800961 | 20 | 43042364 | T | C | 0.035 | 0.160 | 5.10E-20 | 0.018 | C | T | 0.969 | -0.010 | 0.005 | 0.077 |
| rs1999536 | 20 | 45581777 | C | G | 0.573 | -0.040 | 4.86E-10 | 0.007 | G | C | 0.430 | -0.021 | 0.002 | 6.30E-22 |
| rs11699802 | 20 | 48832135 | T | C | 0.465 | -0.044 | 8.84E-12 | 0.007 | C | T | 0.535 | -0.008 | 0.002 | 0.00096 |
| rs4812034 | 20 | 57397566 | T | G | 0.547 | 0.042 | 7.35E-11 | 0.006 | G | T | 0.456 | -0.003 | 0.002 | 0.023 |
| rs2023681 | 22 | 30599562 | A | G | 0.087 | -0.083 | 7.40E-13 | 0.012 | A | G | 0.093 | 0.020 | 0.003 | 1.60E-08 |
| rs5758223 | 22 | 41489920 | A | G | 0.713 | 0.040 | 1.78E-08 | 0.007 | G | A | 0.279 | 0.008 | 0.002 | 0.011 |
| rs738409 | 22 | 44324727 | C | G | 0.774 | -0.044 | 1.17E-08 | 0.008 | C | G | 0.784 | 0.000 | 0.002 | 0.57 |
| rs36138276 | 22 | 50422348 | A | G | 0.499 | -0.043 | 6.40E-11 | 0.007 | G | A | 0.484 | 0.005 | 0.002 | 0.082 |
| rs112915006 | 22 | 50604696 | A | G | 0.949 | -0.090 | 4.62E-09 | 0.015 | A | G | 0.956 | -0.013 | 0.005 | 0.00061 |

| **Supplementary Table** **5.** Summary of pariwise genetic correlation using linkage disequilibrium score regeression (LDSC). | | | | | | | | | |  |  |
| --- | --- | --- | --- | --- | --- | --- | --- | --- | --- | --- | --- |
| p1 | p2 | rg | se | z | p | h2_obs | h2_obs_se | h2_int | h2_int_se | gcov_int | gcov_int_se |
| Type 2 diabetes | Fracture | -0.0114 | 0.0374 | -0.3058 | 0.7598 | 0.063 | 0.0031 | 0.9894 | 0.0229 | -0.0123 | 0.006 |
| Type 2 diabetes | Heel BMD | 0.0923 | 0.0196 | 4.7082 | 2.4996E-06 | 0.064 | 0.0032 | 0.9905 | 0.0215 | 0.0384 | 0.0114 |
| Heel BMD | Fracture | -0.5649 | 0.0413 | -13.6652 | 1.6381E-42 | 0.2937 | 0.0246 | 1.2139 | 0.0543 | -0.0525 | 0.0123 |

p1 = trait 1, p2 = trait 2, rg = genetic correlation, se = standard error of rg, p = p-value for rg, gcov_int, gcov_int_se = cross-trait LD Score regression intercept and standard error

**Supplementary Table** **6.** The results of cross-trait analysis with MiXeR model for type 2 diabetes, fracture and BMD.

| trait1 | trait2 | nc1@p9, mean (std) | nc2@p9, mean (std) |) |) | rho_beta, mean (std) | rg, mean (std) | fraction_concordant_within_shared, mean (std) | π_1_,  mean (std) | π_2_, mean (std) | π_12_, mean (std) | dice, mean (std) | AIC | |
| --- | --- | --- | --- | --- | --- | --- | --- | --- | --- | --- | --- | --- | --- | --- |
|  |  |  |  |  |  |  |  |  |  |  |  |  | best_vs_min | best_vs_max |
| T2D | fracture | 1942 (492) | 627 (173) | 428 (137) | 2370(499) | -0.336(0.134) | -0.086(0.027) | 0.390 (0.046) | 6.09E-04 (1.54E-04) | 1.97E-04 (5.41E-05) | 1.34E-04 (4.29E-05) | 25.25% (7.47%) | 9 | 21 |
| T2D | BMD | 1679(394) | 1022(121) | 691(120) | 2370(499) | 0.354(0.029) | 0.121(0.005) | 0.615(0.010) | 5.26E-04(1.24E-4) | 3.2E-04(3.8E-05) | 2.17E-04(3.77E-05) | 33.67%（2.70%） | 98 | 345 |

nc1@p9, nc2@p9, and nc12@p9 = the number of causal variants for trait1, trait2 and both, respectively; nc@p9 = number of trait-influencing variants specific to type 2 diabetes; rho_beta = the correlation of effect sizes within the shared polygenic component; rg = genetic correlation; concordant_fraction = the proportion of shared variants with concordant direction of effect in both traits on all shared variants; π_1_, π_2_, and π_12_ = polygenicity of trait1, trait2 and both, respectively; The best_vs_min_AIC and the best_vs_max_AIC indicates whether MiXeR can accurately distinguish the reported overlap from the minimum and maximum possible overlap allowed, respectively.

**Supplementary Table 7.** Distinct genomic loci shared between type 2 diabetes and fracture at conjFDR<0.05.

| locusnum | snpid | chrnum | chrpos | A1 | A2 | zscore_  T2D | zscore_  fracture | conjfdr_  T2D_fracture | prune_  T2D_fracture | min_  conjfdr | pval_  T2D | pval_  fracture |
| --- | --- | --- | --- | --- | --- | --- | --- | --- | --- | --- | --- | --- |
| 1 | rs16860216 | 3 | 185488882 | A | G | 11.107 | -4.218 | 0.009 | 1 | 0.009 | 1.16E-28 | 2.47E-05 |
| 2 | rs3093978 | 6 | 31498497 | A | C | -5.463 | 4.078 | 0.016 | 1 | 0.016 | 4.68E-08 | 4.53E-05 |
| 3 | rs12213548 | 6 | 79328352 | T | G | 3.340 | 3.928 | 0.036 | 1 | 0.036 | 8.38E-04 | 8.58E-05 |
| 4 | rs4580892 | 6 | 127409882 | T | C | 5.295 | -6.204 | 0.000 | 1 | 0.000 | 1.19E-07 | 5.51E-10 |
| 5 | rs7824564 | 8 | 11780180 | A | G | -4.001 | 4.108 | 0.014 | 1 | 0.014 | 6.31E-05 | 4.00E-05 |
| 6 | rs9663185 | 10 | 124089514 | A | G | -3.455 | 3.981 | 0.026 | 1 | 0.026 | 5.50E-04 | 6.85E-05 |
| 7 | rs930782 | 11 | 68871794 | T | C | 3.818 | -3.875 | 0.032 | 1 | 0.032 | 1.35E-04 | 1.07E-04 |
| 8 | rs9568867 | 13 | 54107352 | A | G | 4.101 | 4.475 | 0.003 | 1 | 0.003 | 4.11E-05 | 7.62E-06 |
| 9 | rs1494092 | 14 | 46970304 | T | C | -3.266 | -3.799 | 0.045 | 1 | 0.045 | 1.09E-03 | 1.45E-04 |
| 10 | rs676387 | 17 | 40706273 | A | C | 6.031 | -3.980 | 0.023 | 1 | 0.023 | 1.63E-09 | 6.89E-05 |

**Supplementary Table** **8.** The results of SMR analyses for the expression of RSPO3 (ENSG00000146374) with type 2 diabetes, fracture, BMI, WC, WHR and VAT in adipose subcutaneous.

| trait | topSNP | topSNP_pos | A1 | A2 | Freq | b_  GWAS | se_  GWAS | p_  GWAS | b_  eQTL | se_  eQTL | p_  eQTL | b_  SMR | se_  SMR | p_  SMR | p_  HEIDI |
| --- | --- | --- | --- | --- | --- | --- | --- | --- | --- | --- | --- | --- | --- | --- | --- |
| T2D | rs72959041 | chr6:  127454893 | A | G | 0.051 | 0.050 | 0.014 | 5.31E-04 | 0.510 | 0.102 | 6.19E-07 | 0.098 | 0.034 | 0.00436 | 0.001 |
| fracture | rs1936806 | chr6:  127451665 | T | C | 0.410 | -0.061 | 0.008 | 1.86E-13 | 0.167 | 0.043 | 8.78E-05 | -0.364 | 0.105 | 0.00053 | 0.571 |
| BMI | rs1936806 | chr6:  127451665 | T | C | 0.410 | 0.002 | 0.003 | 4.31E-01 | 0.167 | 0.043 | 8.78E-05 | 0.014 | 0.019 | 0.44753 | 0.143 |
| WC | rs1936806 | chr6:  127451665 | T | C | 0.410 | 0.015 | 0.004 | 1.10E-05 | 0.167 | 0.043 | 8.78E-05 | 0.090 | 0.031 | 0.00381 | 0.568 |
| WHR | rs1936806 | chr6:  127451665 | T | C | 0.410 | 0.034 | 0.003 | 1.10E-24 | 0.167 | 0.043 | 8.78E-05 | 0.204 | 0.056 | 0.00025 | 0.481 |
| VAT | rs141783576 | chr6:  127439897 | C | G | 0.071 | 0.092 | 0.014 | 1.60E-11 | 0.464 | 0.089 | 1.68E-07 | 0.199 | 0.049 | 4.36E-05 | 0.870 |

**Supplementary Table** **9.** Distinct genomic loci shared between type 2 diabetes and BMD at conjFDR<0.05.

| locusnum | chrnum | chrpos | zscore_T2D | zscore_BMD | conjfdr_T2D_BMD | prune_T2D_BMD | min_conjfdr | pval_T2D | pval_BMD |
| --- | --- | --- | --- | --- | --- | --- | --- | --- | --- |
| 1 | 1 | 6709911 | 4.969871 | 6.614455 | 7.94E-05 | 1 | 7.94E-05 | 6.70E-07 | 3.73E-11 |
| 2 | 1 | 11128654 | -3.83534 | -6.89051 | 6.95E-03 | 1 | 6.95E-03 | 1.25E-04 | 5.56E-12 |
| 3 | 1 | 16317381 | 3.193884 | 5.007775 | 4.03E-02 | 1 | 4.03E-02 | 1.40E-03 | 5.51E-07 |
| 4 | 1 | 20734368 | 4.047526 | 2.781336 | 4.48E-02 | 1 | 4.48E-02 | 5.18E-05 | 5.41E-03 |
| 5 | 1 | 22287607 | 3.366951 | -3.40682 | 2.61E-02 | 1 | 2.61E-02 | 7.60E-04 | 6.57E-04 |
| 6 | 1 | 22366102 | 3.210808 | -10.1851 | 3.86E-02 | 1 | 3.86E-02 | 1.32E-03 | 2.31E-24 |
| 7 | 1 | 26482556 | 3.134948 | 5.118577 | 4.64E-02 | 1 | 4.64E-02 | 1.72E-03 | 3.08E-07 |
| 8 | 1 | 27284913 | 3.196227 | 3.022638 | 4.52E-02 | 1 | 4.52E-02 | 1.39E-03 | 2.51E-03 |
| 9 | 1 | 31487513 | -4.13586 | 3.204257 | 1.49E-02 | 1 | 1.49E-02 | 3.54E-05 | 1.35E-03 |
| 10 | 1 | 39622588 | 9.16398 | 4.042576 | 9.35E-04 | 1 | 9.35E-04 | 5.00E-20 | 5.29E-05 |
| 10 | 1 | 40157643 | 4.019748 | 3.361517 | 9.44E-03 | 1 | 9.44E-03 | 5.83E-05 | 7.75E-04 |
| 11 | 1 | 51506886 | -5.26205 | -3.73069 | 2.89E-03 | 1 | 2.89E-03 | 1.42E-07 | 1.91E-04 |
| 12 | 1 | 58101851 | 3.470676 | -3.261 | 2.02E-02 | 1 | 2.02E-02 | 5.19E-04 | 1.11E-03 |
| 13 | 1 | 67127242 | 3.385239 | -5.35944 | 2.49E-02 | 1 | 2.49E-02 | 7.11E-04 | 8.35E-08 |
| 14 | 1 | 72647903 | -3.88449 | -3.35647 | 9.57E-03 | 1 | 9.57E-03 | 1.03E-04 | 7.89E-04 |
| 15 | 1 | 75581678 | 3.540634 | 3.448487 | 1.64E-02 | 1 | 1.64E-02 | 3.99E-04 | 5.64E-04 |
| 16 | 1 | 82364429 | 3.306097 | 2.781336 | 4.48E-02 | 1 | 4.48E-02 | 9.46E-04 | 5.41E-03 |
| 17 | 1 | 89363264 | 3.2937 | 6.257267 | 3.14E-02 | 1 | 3.14E-02 | 9.89E-04 | 3.92E-10 |
| 18 | 1 | 92335232 | 4.328733 | 3.715791 | 3.04E-03 | 1 | 3.04E-03 | 1.50E-05 | 2.03E-04 |
| 18 | 1 | 92451018 | -3.16989 | -3.45631 | 4.26E-02 | 1 | 4.26E-02 | 1.52E-03 | 5.48E-04 |
| 18 | 1 | 93247835 | 3.228985 | 3.822015 | 3.69E-02 | 1 | 3.69E-02 | 1.24E-03 | 1.32E-04 |
| 19 | 1 | 100000012 | -3.66317 | 2.95306 | 2.92E-02 | 1 | 2.92E-02 | 2.49E-04 | 3.15E-03 |
| 20 | 1 | 103214274 | 3.852976 | -5.54655 | 6.59E-03 | 1 | 6.59E-03 | 1.17E-04 | 2.91E-08 |
| 20 | 1 | 103415461 | -3.33186 | 3.689017 | 2.86E-02 | 1 | 2.86E-02 | 8.63E-04 | 2.25E-04 |
| 21 | 1 | 110470592 | 3.230908 | -2.91321 | 4.38E-02 | 1 | 4.38E-02 | 1.23E-03 | 3.58E-03 |
| 22 | 1 | 149865483 | -4.24805 | -4.87008 | 1.67E-03 | 1 | 1.67E-03 | 2.16E-05 | 1.12E-06 |
| 23 | 1 | 150650581 | -3.38556 | -3.23084 | 2.56E-02 | 1 | 2.56E-02 | 7.10E-04 | 1.23E-03 |
| 24 | 1 | 154305010 | 3.70662 | 3.204257 | 1.49E-02 | 1 | 1.49E-02 | 2.10E-04 | 1.35E-03 |
| 25 | 1 | 154668217 | -3.2283 | -2.8989 | 4.44E-02 | 1 | 4.44E-02 | 1.25E-03 | 3.74E-03 |
| 26 | 1 | 155148781 | 3.143141 | -4.80659 | 4.55E-02 | 1 | 4.55E-02 | 1.67E-03 | 1.54E-06 |
| 27 | 1 | 156688152 | -3.27442 | -2.8633 | 4.05E-02 | 1 | 4.05E-02 | 1.06E-03 | 4.19E-03 |
| 28 | 1 | 171076966 | 3.502966 | -3.92994 | 1.83E-02 | 1 | 1.83E-02 | 4.60E-04 | 8.50E-05 |
| 29 | 1 | 172349246 | -3.77851 | -4.26208 | 8.27E-03 | 1 | 8.27E-03 | 1.58E-04 | 2.03E-05 |
| 30 | 1 | 177763013 | 4.976923 | 3.91302 | 1.52E-03 | 1 | 1.52E-03 | 6.46E-07 | 9.11E-05 |
| 30 | 1 | 177885762 | 5.560437 | 4.089251 | 7.84E-04 | 1 | 7.84E-04 | 2.69E-08 | 4.33E-05 |
| 31 | 1 | 180603162 | 3.289672 | -2.80427 | 4.23E-02 | 1 | 4.23E-02 | 1.00E-03 | 5.04E-03 |
| 32 | 1 | 191424825 | -3.44161 | -3.05331 | 2.40E-02 | 1 | 2.40E-02 | 5.78E-04 | 2.26E-03 |
| 33 | 1 | 214154547 | -6.86651 | -2.97569 | 2.77E-02 | 1 | 2.77E-02 | 6.58E-12 | 2.92E-03 |
| 34 | 1 | 219685466 | 3.731896 | -3.45631 | 9.53E-03 | 1 | 9.53E-03 | 1.90E-04 | 5.48E-04 |
| 34 | 1 | 219717264 | -6.21117 | -4.1381 | 6.50E-04 | 1 | 6.50E-04 | 5.26E-10 | 3.50E-05 |
| 34 | 1 | 219762070 | 5.344882 | 4.947981 | 2.18E-05 | 1 | 2.18E-05 | 9.05E-08 | 7.50E-07 |
| 35 | 1 | 219966068 | -3.51979 | -3.67688 | 1.74E-02 | 1 | 1.74E-02 | 4.32E-04 | 2.36E-04 |
| 36 | 1 | 229672955 | 7.015201 | 3.715791 | 3.04E-03 | 1 | 3.04E-03 | 2.30E-12 | 2.03E-04 |
| 37 | 2 | 422144 | -6.03817 | -4.08925 | 7.84E-04 | 1 | 7.84E-04 | 1.56E-09 | 4.33E-05 |
| 37 | 2 | 437664 | 3.364993 | 2.872524 | 3.58E-02 | 1 | 3.58E-02 | 7.65E-04 | 4.07E-03 |
| 38 | 2 | 621254 | 3.374008 | 2.909556 | 3.28E-02 | 1 | 3.28E-02 | 7.41E-04 | 3.62E-03 |
| 38 | 2 | 630070 | -5.47529 | -4.87008 | 3.14E-05 | 1 | 3.14E-05 | 4.37E-08 | 1.12E-06 |
| 39 | 2 | 18707873 | -4.09174 | -3.2959 | 1.14E-02 | 1 | 1.14E-02 | 4.28E-05 | 9.81E-04 |
| 40 | 2 | 25345685 | 3.365585 | 3.053307 | 2.92E-02 | 1 | 2.92E-02 | 7.64E-04 | 2.26E-03 |
| 41 | 2 | 26018880 | 3.455079 | 2.975694 | 2.77E-02 | 1 | 2.77E-02 | 5.50E-04 | 2.92E-03 |
| 42 | 2 | 40849925 | -3.73069 | 3.73069 | 9.56E-03 | 1 | 9.56E-03 | 1.91E-04 | 1.91E-04 |
| 42 | 2 | 40974083 | 3.236685 | -5.01118 | 3.62E-02 | 1 | 3.62E-02 | 1.21E-03 | 5.41E-07 |
| 43 | 2 | 43429806 | 5.740224 | -4.79025 | 4.49E-05 | 1 | 4.49E-05 | 9.46E-09 | 1.67E-06 |
| 44 | 2 | 43452183 | -5.13305 | 5.428003 | 3.68E-05 | 1 | 3.68E-05 | 2.85E-07 | 5.70E-08 |
| 44 | 2 | 43584121 | 3.895681 | -3.261 | 1.27E-02 | 1 | 1.27E-02 | 9.79E-05 | 1.11E-03 |
| 44 | 2 | 43671176 | -10.2844 | 7.380633 | 1.02E-11 | 1 | 1.02E-11 | 8.29E-25 | 1.58E-13 |
| 44 | 2 | 43679175 | 4.68716 | -3.51058 | 5.99E-03 | 1 | 5.99E-03 | 2.77E-06 | 4.47E-04 |
| 44 | 2 | 43920357 | -4.59615 | 4.012386 | 1.05E-03 | 1 | 1.05E-03 | 4.30E-06 | 6.01E-05 |
| 44 | 2 | 43994026 | -5.41548 | 3.86053 | 1.83E-03 | 1 | 1.83E-03 | 6.11E-08 | 1.13E-04 |
| 45 | 2 | 48471055 | -3.37499 | -2.76026 | 4.70E-02 | 1 | 4.70E-02 | 7.38E-04 | 5.78E-03 |
| 46 | 2 | 54421356 | 3.289268 | 4.613039 | 3.17E-02 | 1 | 3.17E-02 | 1.00E-03 | 3.97E-06 |
| 47 | 2 | 54986180 | 3.841072 | 2.872524 | 3.58E-02 | 1 | 3.58E-02 | 1.22E-04 | 4.07E-03 |
| 48 | 2 | 55135551 | -3.27285 | 3.104232 | 3.60E-02 | 1 | 3.60E-02 | 1.06E-03 | 1.91E-03 |
| 49 | 2 | 55319643 | 3.395828 | -4.74386 | 2.42E-02 | 1 | 2.42E-02 | 6.84E-04 | 2.10E-06 |
| 50 | 2 | 59314142 | -3.79501 | 3.495632 | 7.89E-03 | 1 | 7.89E-03 | 1.48E-04 | 4.73E-04 |
| 51 | 2 | 59562715 | -3.4595 | -2.83209 | 3.96E-02 | 1 | 3.96E-02 | 5.41E-04 | 4.62E-03 |
| 52 | 2 | 60540393 | -3.63904 | 2.95306 | 2.92E-02 | 1 | 2.92E-02 | 2.74E-04 | 3.15E-03 |
| 53 | 2 | 65390209 | 3.541209 | 3.158919 | 1.74E-02 | 1 | 1.74E-02 | 3.98E-04 | 1.58E-03 |
| 54 | 2 | 65543014 | 3.500593 | 3.510584 | 1.84E-02 | 1 | 1.84E-02 | 4.64E-04 | 4.47E-04 |
| 54 | 2 | 65656969 | 5.225034 | 4.188277 | 5.35E-04 | 1 | 5.35E-04 | 1.74E-07 | 2.81E-05 |
| 55 | 2 | 67906813 | 4.106475 | 3.472879 | 6.72E-03 | 1 | 6.72E-03 | 4.02E-05 | 5.15E-04 |
| 56 | 2 | 100874627 | 3.462975 | 3.129968 | 2.19E-02 | 1 | 2.19E-02 | 5.34E-04 | 1.75E-03 |
| 57 | 2 | 101731397 | -3.17216 | 3.481684 | 4.24E-02 | 1 | 4.24E-02 | 1.51E-03 | 4.98E-04 |
| 58 | 2 | 111831793 | -3.18279 | 3.689017 | 4.14E-02 | 1 | 4.14E-02 | 1.46E-03 | 2.25E-04 |
| 59 | 2 | 111908567 | -4.51708 | -3.82706 | 2.06E-03 | 1 | 2.06E-03 | 6.27E-06 | 1.30E-04 |
| 60 | 2 | 112270857 | 3.389412 | 3.180465 | 2.59E-02 | 1 | 2.59E-02 | 7.00E-04 | 1.47E-03 |
| 61 | 2 | 112776979 | -3.90514 | 3.349114 | 9.79E-03 | 1 | 9.79E-03 | 9.42E-05 | 8.11E-04 |
| 62 | 2 | 119507201 | 3.176338 | 4.542695 | 4.20E-02 | 1 | 4.20E-02 | 1.49E-03 | 5.55E-06 |
| 63 | 2 | 119509170 | -3.38057 | -6.59559 | 2.52E-02 | 1 | 2.52E-02 | 7.23E-04 | 4.24E-11 |
| 64 | 2 | 119585954 | 3.429386 | 17.11948 | 2.22E-02 | 1 | 2.22E-02 | 6.05E-04 | 1.06E-65 |
| 65 | 2 | 145256545 | 3.910895 | 3.500497 | 6.18E-03 | 1 | 6.18E-03 | 9.20E-05 | 4.64E-04 |
| 66 | 2 | 145759511 | -3.49805 | -5.9592 | 1.85E-02 | 1 | 1.85E-02 | 4.69E-04 | 2.53E-09 |
| 67 | 2 | 146380936 | 3.799776 | 4.539567 | 7.78E-03 | 1 | 7.78E-03 | 1.45E-04 | 5.64E-06 |
| 68 | 2 | 148563066 | -3.60272 | -2.95742 | 2.89E-02 | 1 | 2.89E-02 | 3.15E-04 | 3.10E-03 |
| 69 | 2 | 151987931 | 4.069428 | 2.94456 | 2.99E-02 | 1 | 2.99E-02 | 4.71E-05 | 3.23E-03 |
| 69 | 2 | 152143578 | 3.590285 | 3.034438 | 2.37E-02 | 1 | 2.37E-02 | 3.30E-04 | 2.41E-03 |
| 70 | 2 | 161536620 | -3.80017 | -3.51582 | 7.77E-03 | 1 | 7.77E-03 | 1.45E-04 | 4.38E-04 |
| 71 | 2 | 166146520 | 3.350547 | -2.85151 | 3.78E-02 | 1 | 3.78E-02 | 8.07E-04 | 4.35E-03 |
| 72 | 2 | 166603281 | 3.85962 | 2.811564 | 4.16E-02 | 1 | 4.16E-02 | 1.14E-04 | 4.93E-03 |
| 73 | 2 | 172401050 | 3.125209 | 3.625352 | 4.75E-02 | 1 | 4.75E-02 | 1.78E-03 | 2.89E-04 |
| 74 | 2 | 196635762 | 3.591263 | 3.169439 | 1.64E-02 | 1 | 1.64E-02 | 3.29E-04 | 1.53E-03 |
| 75 | 2 | 202962946 | -3.21632 | -3.70195 | 3.81E-02 | 1 | 3.81E-02 | 1.30E-03 | 2.14E-04 |
| 75 | 2 | 203137466 | 3.115296 | 5.687211 | 4.86E-02 | 1 | 4.86E-02 | 1.84E-03 | 1.29E-08 |
| 75 | 2 | 203288774 | 4.147975 | -5.72142 | 2.41E-03 | 1 | 2.41E-03 | 3.35E-05 | 1.06E-08 |
| 76 | 2 | 219184872 | 3.317036 | 3.204257 | 3.08E-02 | 1 | 3.08E-02 | 9.10E-04 | 1.35E-03 |
| 77 | 2 | 227020399 | -6.68113 | -3.67688 | 3.48E-03 | 1 | 3.48E-03 | 2.37E-11 | 2.36E-04 |
| 78 | 2 | 240301377 | 3.387478 | 4.277132 | 2.48E-02 | 1 | 2.48E-02 | 7.05E-04 | 1.89E-05 |
| 79 | 3 | 11487506 | -3.80598 | 7.649836 | 7.63E-03 | 1 | 7.63E-03 | 1.41E-04 | 2.01E-14 |
| 80 | 3 | 12294202 | -6.3142 | 5.594938 | 8.17E-07 | 1 | 8.17E-07 | 2.72E-10 | 2.21E-08 |
| 81 | 3 | 18698928 | -3.47573 | 2.948773 | 2.96E-02 | 1 | 2.96E-02 | 5.09E-04 | 3.19E-03 |
| 82 | 3 | 28753072 | 3.294887 | 3.326251 | 3.13E-02 | 1 | 3.13E-02 | 9.85E-04 | 8.80E-04 |
| 83 | 3 | 29786145 | 3.156499 | 3.14886 | 4.67E-02 | 1 | 4.67E-02 | 1.60E-03 | 1.64E-03 |
| 84 | 3 | 30007227 | 3.862695 | 3.625352 | 6.40E-03 | 1 | 6.40E-03 | 1.12E-04 | 2.89E-04 |
| 85 | 3 | 46716597 | 4.018166 | 3.591958 | 4.61E-03 | 1 | 4.61E-03 | 5.87E-05 | 3.28E-04 |
| 85 | 3 | 47080127 | -4.87639 | -5.38685 | 1.23E-04 | 1 | 1.23E-04 | 1.08E-06 | 7.17E-08 |
| 85 | 3 | 48068610 | 4.84949 | 7.335098 | 1.38E-04 | 1 | 1.38E-04 | 1.24E-06 | 2.22E-13 |
| 85 | 3 | 49931760 | 3.935353 | 4.977541 | 5.05E-03 | 1 | 5.05E-03 | 8.31E-05 | 6.44E-07 |
| 85 | 3 | 49980596 | -5.72041 | -6.15992 | 2.11E-06 | 1 | 2.11E-06 | 1.06E-08 | 7.28E-10 |
| 85 | 3 | 50181135 | 3.280526 | 8.741327 | 3.25E-02 | 1 | 3.25E-02 | 1.04E-03 | 2.30E-18 |
| 85 | 3 | 50589611 | -3.19388 | -3.51058 | 4.03E-02 | 1 | 4.03E-02 | 1.40E-03 | 4.47E-04 |
| 86 | 3 | 52076178 | -3.70997 | -3.21716 | 1.44E-02 | 1 | 1.44E-02 | 2.07E-04 | 1.29E-03 |
| 86 | 3 | 52607511 | 3.409489 | 4.42327 | 2.34E-02 | 1 | 2.34E-02 | 6.51E-04 | 9.72E-06 |
| 86 | 3 | 53131772 | -5.55005 | -4.835 | 3.68E-05 | 1 | 3.68E-05 | 2.86E-08 | 1.33E-06 |
| 87 | 3 | 57893050 | 3.414699 | -5.6524 | 2.30E-02 | 1 | 2.30E-02 | 6.39E-04 | 1.58E-08 |
| 88 | 3 | 66448307 | -3.58004 | 3.346716 | 1.48E-02 | 1 | 1.48E-02 | 3.44E-04 | 8.18E-04 |
| 89 | 3 | 71594837 | -4.20953 | -3.56331 | 5.06E-03 | 1 | 5.06E-03 | 2.56E-05 | 3.66E-04 |
| 90 | 3 | 94015993 | -3.39352 | -2.81404 | 4.13E-02 | 1 | 4.13E-02 | 6.90E-04 | 4.89E-03 |
| 91 | 3 | 123010775 | -3.47359 | -3.09625 | 2.15E-02 | 1 | 2.15E-02 | 5.14E-04 | 1.96E-03 |
| 91 | 3 | 123068744 | -10.838 | -4.42327 | 2.10E-04 | 1 | 2.10E-04 | 2.27E-27 | 9.72E-06 |
| 91 | 3 | 123110776 | 3.447412 | 2.990354 | 2.66E-02 | 1 | 2.66E-02 | 5.66E-04 | 2.79E-03 |
| 92 | 3 | 125082650 | 3.301277 | 3.19205 | 3.22E-02 | 1 | 3.22E-02 | 9.62E-04 | 1.41E-03 |
| 93 | 3 | 125208841 | 3.500396 | -3.47724 | 1.84E-02 | 1 | 1.84E-02 | 4.65E-04 | 5.07E-04 |
| 94 | 3 | 129323322 | 3.76337 | -3.01695 | 2.48E-02 | 1 | 2.48E-02 | 1.68E-04 | 2.55E-03 |
| 95 | 3 | 129336406 | -4.25854 | 4.138096 | 1.60E-03 | 1 | 1.60E-03 | 2.06E-05 | 3.50E-05 |
| 96 | 3 | 138051423 | 4.933468 | -3.62535 | 4.14E-03 | 1 | 4.14E-03 | 8.08E-07 | 2.89E-04 |
| 97 | 3 | 141076084 | -4.56273 | -5.46778 | 4.79E-04 | 1 | 4.79E-04 | 5.05E-06 | 4.56E-08 |
| 98 | 3 | 150047893 | 4.745976 | 4.152764 | 6.13E-04 | 1 | 6.13E-04 | 2.08E-06 | 3.28E-05 |
| 98 | 3 | 150074251 | 4.403006 | 3.158919 | 1.69E-02 | 1 | 1.69E-02 | 1.07E-05 | 1.58E-03 |
| 99 | 3 | 151758998 | -3.27112 | -2.80427 | 4.29E-02 | 1 | 4.29E-02 | 1.07E-03 | 5.04E-03 |
| 100 | 3 | 152022890 | 4.802081 | 4.300745 | 3.45E-04 | 1 | 3.45E-04 | 1.57E-06 | 1.70E-05 |
| 101 | 3 | 152481845 | 4.265614 | -3.13922 | 1.79E-02 | 1 | 1.79E-02 | 1.99E-05 | 1.69E-03 |
| 102 | 3 | 154088411 | -3.5965 | -3.43012 | 1.41E-02 | 1 | 1.41E-02 | 3.23E-04 | 6.03E-04 |
| 103 | 3 | 160303591 | 4.084051 | -3.55036 | 5.27E-03 | 1 | 5.27E-03 | 4.43E-05 | 3.85E-04 |
| 104 | 3 | 170711968 | -5.09834 | 2.760255 | 4.70E-02 | 1 | 4.70E-02 | 3.43E-07 | 5.78E-03 |
| 105 | 3 | 185425976 | -4.54825 | -3.98059 | 1.18E-03 | 1 | 1.18E-03 | 5.41E-06 | 6.87E-05 |
| 105 | 3 | 185530290 | 15.12012 | 4.70831 | 6.40E-05 | 1 | 6.40E-05 | 1.19E-51 | 2.50E-06 |
| 106 | 4 | 797490 | 3.292245 | -4.43913 | 3.15E-02 | 1 | 3.15E-02 | 9.94E-04 | 9.03E-06 |
| 107 | 4 | 1010077 | 3.622034 | -19.2781 | 1.31E-02 | 1 | 1.31E-02 | 2.92E-04 | 8.21E-83 |
| 108 | 4 | 1794188 | 3.898034 | 3.066772 | 2.18E-02 | 1 | 2.18E-02 | 9.70E-05 | 2.16E-03 |
| 109 | 4 | 4890108 | -3.14937 | 3.158919 | 4.74E-02 | 1 | 4.74E-02 | 1.64E-03 | 1.58E-03 |
| 110 | 4 | 18025484 | -5.27969 | -5.63136 | 1.83E-05 | 1 | 1.83E-05 | 1.29E-07 | 1.79E-08 |
| 111 | 4 | 18232082 | 3.385239 | 3.295899 | 2.49E-02 | 1 | 2.49E-02 | 7.11E-04 | 9.81E-04 |
| 112 | 4 | 20265535 | 4.0557 | -3.61647 | 4.25E-03 | 1 | 4.25E-03 | 5.00E-05 | 2.99E-04 |
| 113 | 4 | 25408838 | -3.69876 | -5.51926 | 1.05E-02 | 1 | 1.05E-02 | 2.17E-04 | 3.40E-08 |
| 114 | 4 | 26314032 | 4.171582 | 4.798198 | 2.21E-03 | 1 | 2.21E-03 | 3.02E-05 | 1.60E-06 |
| 115 | 4 | 38383633 | 3.829699 | 6.22438 | 7.08E-03 | 1 | 7.08E-03 | 1.28E-04 | 4.83E-10 |
| 116 | 4 | 38731295 | 3.590145 | 3.490878 | 1.43E-02 | 1 | 1.43E-02 | 3.30E-04 | 4.81E-04 |
| 117 | 4 | 45186139 | 6.034548 | 3.510584 | 5.99E-03 | 1 | 5.99E-03 | 1.59E-09 | 4.47E-04 |
| 118 | 4 | 53322300 | 3.663725 | -3.73069 | 1.16E-02 | 1 | 1.16E-02 | 2.49E-04 | 1.91E-04 |
| 119 | 4 | 89779909 | -4.06085 | 3.689017 | 3.34E-03 | 1 | 3.34E-03 | 4.89E-05 | 2.25E-04 |
| 120 | 4 | 94836997 | 3.183123 | -2.92451 | 4.89E-02 | 1 | 4.89E-02 | 1.46E-03 | 3.45E-03 |
| 120 | 4 | 95091911 | -5.93901 | 7.172212 | 6.41E-07 | 1 | 6.41E-07 | 2.87E-09 | 7.38E-13 |
| 121 | 4 | 95946506 | -3.84978 | -3.34672 | 9.87E-03 | 1 | 9.87E-03 | 1.18E-04 | 8.18E-04 |
| 122 | 4 | 100239319 | -3.20321 | 3.577127 | 3.94E-02 | 1 | 3.94E-02 | 1.36E-03 | 3.47E-04 |
| 123 | 4 | 103146888 | 3.422051 | -2.81404 | 4.13E-02 | 1 | 4.13E-02 | 6.22E-04 | 4.89E-03 |
| 123 | 4 | 103680984 | -5.27565 | 3.958625 | 1.28E-03 | 1 | 1.28E-03 | 1.32E-07 | 7.54E-05 |
| 123 | 4 | 104199922 | 3.823221 | -3.37198 | 9.16E-03 | 1 | 9.16E-03 | 1.32E-04 | 7.46E-04 |
| 124 | 4 | 119490085 | 3.672852 | -3.40369 | 1.13E-02 | 1 | 1.13E-02 | 2.40E-04 | 6.65E-04 |
| 125 | 4 | 123833154 | -3.83515 | -3.6444 | 6.95E-03 | 1 | 6.95E-03 | 1.25E-04 | 2.68E-04 |
| 126 | 4 | 145659064 | 4.667992 | 4.487176 | 3.09E-04 | 1 | 3.09E-04 | 3.04E-06 | 7.22E-06 |
| 127 | 4 | 157814426 | -3.24376 | -3.00064 | 4.07E-02 | 1 | 4.07E-02 | 1.18E-03 | 2.69E-03 |
| 128 | 4 | 174751634 | 3.156499 | 3.394563 | 4.40E-02 | 1 | 4.40E-02 | 1.60E-03 | 6.87E-04 |
| 129 | 4 | 185726914 | -7.03811 | 3.715791 | 3.04E-03 | 1 | 3.04E-03 | 1.95E-12 | 2.03E-04 |
| 129 | 4 | 185766446 | -3.45657 | 3.04685 | 2.31E-02 | 1 | 2.31E-02 | 5.47E-04 | 2.31E-03 |
| 130 | 4 | 186668660 | -3.70907 | 3.616469 | 1.02E-02 | 1 | 1.02E-02 | 2.08E-04 | 2.99E-04 |
| 131 | 5 | 9010282 | 3.379253 | -2.78134 | 4.48E-02 | 1 | 4.48E-02 | 7.27E-04 | 5.41E-03 |
| 132 | 5 | 14712327 | -3.89627 | 4.012386 | 5.73E-03 | 1 | 5.73E-03 | 9.77E-05 | 6.01E-05 |
| 133 | 5 | 14815699 | -5.47529 | -3.83757 | 1.98E-03 | 1 | 1.98E-03 | 4.37E-08 | 1.24E-04 |
| 134 | 5 | 15597234 | 3.362873 | 3.419818 | 2.64E-02 | 1 | 2.64E-02 | 7.71E-04 | 6.27E-04 |
| 135 | 5 | 51787940 | 3.900361 | -3.61647 | 5.66E-03 | 1 | 5.66E-03 | 9.60E-05 | 2.99E-04 |
| 136 | 5 | 52031269 | 3.748438 | 3.430119 | 9.08E-03 | 1 | 9.08E-03 | 1.78E-04 | 6.03E-04 |
| 137 | 5 | 52184268 | 3.405338 | 4.634067 | 2.36E-02 | 1 | 2.36E-02 | 6.61E-04 | 3.59E-06 |
| 138 | 5 | 53275370 | -5.364 | -3.33738 | 1.02E-02 | 1 | 1.02E-02 | 8.14E-08 | 8.46E-04 |
| 139 | 5 | 55812130 | -8.8846 | 3.096247 | 2.01E-02 | 1 | 2.01E-02 | 6.41E-19 | 1.96E-03 |
| 140 | 5 | 55848669 | 3.547149 | 2.851506 | 3.78E-02 | 1 | 3.78E-02 | 3.89E-04 | 4.35E-03 |
| 140 | 5 | 55861464 | 8.028066 | 3.848702 | 1.91E-03 | 1 | 1.91E-03 | 9.90E-16 | 1.19E-04 |
| 141 | 5 | 64219611 | 3.529066 | 3.059944 | 2.22E-02 | 1 | 2.22E-02 | 4.17E-04 | 2.21E-03 |
| 142 | 5 | 66287883 | 3.370547 | 3.440947 | 2.59E-02 | 1 | 2.59E-02 | 7.50E-04 | 5.80E-04 |
| 143 | 5 | 72256434 | 3.580974 | 3.081049 | 2.09E-02 | 1 | 2.09E-02 | 3.42E-04 | 2.06E-03 |
| 144 | 5 | 74909972 | -5.18557 | -3.71579 | 3.04E-03 | 1 | 3.04E-03 | 2.15E-07 | 2.03E-04 |
| 145 | 5 | 76444754 | 4.341935 | 2.760255 | 4.70E-02 | 1 | 4.70E-02 | 1.41E-05 | 5.78E-03 |
| 146 | 5 | 78522359 | -6.00281 | -3.76815 | 2.54E-03 | 1 | 2.54E-03 | 1.94E-09 | 1.64E-04 |
| 147 | 5 | 87986284 | 4.111571 | 3.406817 | 8.24E-03 | 1 | 8.24E-03 | 3.93E-05 | 6.57E-04 |
| 148 | 5 | 99997791 | 4.089791 | 2.829433 | 3.99E-02 | 1 | 3.99E-02 | 4.32E-05 | 4.66E-03 |
| 149 | 5 | 102615675 | 5.873699 | 3.607963 | 4.37E-03 | 1 | 4.37E-03 | 4.26E-09 | 3.09E-04 |
| 150 | 5 | 112263863 | 3.576576 | 4.782688 | 1.49E-02 | 1 | 1.49E-02 | 3.48E-04 | 1.73E-06 |
| 151 | 5 | 127350549 | -4.08764 | -3.74683 | 3.00E-03 | 1 | 3.00E-03 | 4.36E-05 | 1.79E-04 |
| 152 | 5 | 135348376 | -3.56382 | 3.854523 | 1.54E-02 | 1 | 1.54E-02 | 3.65E-04 | 1.16E-04 |
| 153 | 5 | 176597559 | -3.62618 | -3.38581 | 1.30E-02 | 1 | 1.30E-02 | 2.88E-04 | 7.10E-04 |
| 154 | 6 | 7005942 | 3.501579 | 2.970996 | 2.80E-02 | 1 | 2.80E-02 | 4.63E-04 | 2.97E-03 |
| 154 | 6 | 7032944 | 3.267491 | 4.206044 | 3.36E-02 | 1 | 3.36E-02 | 1.09E-03 | 2.60E-05 |
| 154 | 6 | 7039853 | 4.857071 | -4.37177 | 2.59E-04 | 1 | 2.59E-04 | 1.19E-06 | 1.23E-05 |
| 154 | 6 | 7114438 | 3.899021 | -4.2124 | 5.69E-03 | 1 | 5.69E-03 | 9.66E-05 | 2.53E-05 |
| 154 | 6 | 7188633 | 3.295529 | 10.39513 | 3.13E-02 | 1 | 3.13E-02 | 9.82E-04 | 2.61E-25 |
| 154 | 6 | 7196323 | -4.66672 | 4.782688 | 3.10E-04 | 1 | 3.10E-04 | 3.06E-06 | 1.73E-06 |
| 154 | 6 | 7231843 | -8.98087 | -9.35227 | 1.43E-16 | 1 | 1.43E-16 | 2.69E-19 | 8.58E-21 |
| 154 | 6 | 7255015 | 6.730003 | -3.97485 | 1.21E-03 | 1 | 1.21E-03 | 1.70E-11 | 7.04E-05 |
| 155 | 6 | 7307114 | -5.12318 | -3.06677 | 2.18E-02 | 1 | 2.18E-02 | 3.00E-07 | 2.16E-03 |
| 159 | 6 | 31461979 | 4.622562 | 4.476363 | 3.74E-04 | 1 | 3.74E-04 | 3.79E-06 | 7.59E-06 |
| 161 | 6 | 34203893 | 4.387226 | 3.059944 | 2.22E-02 | 1 | 2.22E-02 | 1.15E-05 | 2.21E-03 |
| 162 | 6 | 36645203 | -3.1227 | -3.24542 | 4.87E-02 | 1 | 4.87E-02 | 1.79E-03 | 1.17E-03 |
| 163 | 6 | 37474197 | -3.91798 | -2.76026 | 4.70E-02 | 1 | 4.70E-02 | 8.93E-05 | 5.78E-03 |
| 164 | 6 | 39835848 | -3.57539 | 4.408813 | 1.50E-02 | 1 | 1.50E-02 | 3.50E-04 | 1.04E-05 |
| 165 | 6 | 43345803 | 3.899021 | 3.807636 | 5.69E-03 | 1 | 5.69E-03 | 9.66E-05 | 1.40E-04 |
| 166 | 6 | 43757896 | -5.3428 | -5.39738 | 1.37E-05 | 1 | 1.37E-05 | 9.15E-08 | 6.76E-08 |
| 167 | 6 | 43844072 | -3.40741 | 3.245416 | 2.40E-02 | 1 | 2.40E-02 | 6.56E-04 | 1.17E-03 |
| 168 | 6 | 44683123 | 3.532382 | 13.50799 | 1.68E-02 | 1 | 1.68E-02 | 4.12E-04 | 1.40E-41 |
| 169 | 6 | 55089936 | 3.797346 | 3.19205 | 1.54E-02 | 1 | 1.54E-02 | 1.46E-04 | 1.41E-03 |
| 170 | 6 | 56231723 | -3.45883 | -3.01695 | 2.48E-02 | 1 | 2.48E-02 | 5.43E-04 | 2.55E-03 |
| 171 | 6 | 56280670 | -3.59015 | 4.983107 | 1.43E-02 | 1 | 1.43E-02 | 3.30E-04 | 6.26E-07 |
| 172 | 6 | 80814080 | -3.51558 | -3.92994 | 1.76E-02 | 1 | 1.76E-02 | 4.39E-04 | 8.50E-05 |
| 173 | 6 | 100836022 | 3.275145 | -4.21899 | 3.30E-02 | 1 | 3.30E-02 | 1.06E-03 | 2.45E-05 |
| 174 | 6 | 111903329 | -3.80226 | -3.43367 | 7.72E-03 | 1 | 7.72E-03 | 1.43E-04 | 5.95E-04 |
| 175 | 6 | 113435831 | -3.38722 | 2.924508 | 3.15E-02 | 1 | 3.15E-02 | 7.06E-04 | 3.45E-03 |
| 176 | 6 | 114173162 | -3.62682 | 2.819071 | 4.09E-02 | 1 | 4.09E-02 | 2.87E-04 | 4.82E-03 |
| 177 | 6 | 117874257 | 4.150659 | -3.08853 | 2.05E-02 | 1 | 2.05E-02 | 3.32E-05 | 2.01E-03 |
| 178 | 6 | 126290265 | 3.350995 | 5.193613 | 2.72E-02 | 1 | 2.72E-02 | 8.05E-04 | 2.06E-07 |
| 179 | 6 | 126802598 | -4.57534 | -6.45534 | 4.56E-04 | 1 | 4.56E-04 | 4.75E-06 | 1.08E-10 |
| 179 | 6 | 126836780 | 3.853791 | 3.969278 | 6.58E-03 | 1 | 6.58E-03 | 1.16E-04 | 7.21E-05 |
| 179 | 6 | 127118646 | -5.12824 | 6.257267 | 3.76E-05 | 1 | 3.76E-05 | 2.92E-07 | 3.92E-10 |
| 179 | 6 | 127202364 | 3.726806 | 4.124559 | 9.67E-03 | 1 | 9.67E-03 | 1.94E-04 | 3.71E-05 |
| 179 | 6 | 127399384 | -3.29498 | 5.392014 | 3.13E-02 | 1 | 3.13E-02 | 9.84E-04 | 6.97E-08 |
| 179 | 6 | 127409882 | 5.502504 | 17.76955 | 6.37E-06 | 1 | 6.37E-06 | 3.74E-08 | 1.22E-70 |
| 179 | 6 | 127414838 | -7.60879 | 13.11604 | 1.07E-11 | 1 | 1.07E-11 | 2.77E-14 | 2.66E-39 |
| 179 | 6 | 127497275 | -3.29834 | 4.138096 | 3.11E-02 | 1 | 3.11E-02 | 9.73E-04 | 3.50E-05 |
| 181 | 6 | 127817401 | 4.910341 | 6.080368 | 1.05E-04 | 1 | 1.05E-04 | 9.09E-07 | 1.20E-09 |
| 182 | 6 | 130323447 | 4.184677 | 4.341505 | 2.11E-03 | 1 | 2.11E-03 | 2.86E-05 | 1.42E-05 |
| 182 | 6 | 130397319 | 4.264546 | 3.992649 | 1.56E-03 | 1 | 1.56E-03 | 2.00E-05 | 6.53E-05 |
| 183 | 6 | 131928008 | 3.79237 | 3.339679 | 1.01E-02 | 1 | 1.01E-02 | 1.49E-04 | 8.39E-04 |
| 184 | 6 | 133135974 | 3.219453 | 7.27385 | 3.78E-02 | 1 | 3.78E-02 | 1.28E-03 | 3.49E-13 |
| 185 | 6 | 133631458 | 3.242923 | 5.223953 | 3.57E-02 | 1 | 3.57E-02 | 1.18E-03 | 1.75E-07 |
| 186 | 6 | 136228617 | -3.3355 | 5.104723 | 2.83E-02 | 1 | 2.83E-02 | 8.51E-04 | 3.31E-07 |
| 187 | 6 | 143058408 | 3.312657 | 2.80911 | 4.18E-02 | 1 | 4.18E-02 | 9.24E-04 | 4.97E-03 |
| 188 | 6 | 160567928 | 3.193884 | 4.02685 | 4.03E-02 | 1 | 4.03E-02 | 1.40E-03 | 5.65E-05 |
| 189 | 6 | 164133001 | -5.62402 | -9.0936 | 3.48E-06 | 1 | 3.48E-06 | 1.87E-08 | 9.58E-20 |
| 190 | 7 | 1973970 | -3.8872 | 2.819071 | 4.09E-02 | 1 | 4.09E-02 | 1.01E-04 | 4.82E-03 |
| 191 | 7 | 4683276 | 4.362167 | 3.998993 | 1.10E-03 | 1 | 1.10E-03 | 1.29E-05 | 6.36E-05 |
| 192 | 7 | 7249747 | 4.562729 | -3.2959 | 1.14E-02 | 1 | 1.14E-02 | 5.05E-06 | 9.81E-04 |
| 193 | 7 | 15031507 | 7.584548 | -4.95719 | 2.09E-05 | 1 | 2.09E-05 | 3.34E-14 | 7.15E-07 |
| 194 | 7 | 15424028 | 3.238063 | -3.1125 | 3.91E-02 | 1 | 3.91E-02 | 1.20E-03 | 1.86E-03 |
| 195 | 7 | 15913588 | 3.968831 | 5.0647 | 4.53E-03 | 1 | 4.53E-03 | 7.22E-05 | 4.09E-07 |
| 196 | 7 | 16268502 | 3.241559 | 3.104232 | 3.89E-02 | 1 | 3.89E-02 | 1.19E-03 | 1.91E-03 |
| 197 | 7 | 25930431 | 3.385813 | -4.30441 | 2.49E-02 | 1 | 2.49E-02 | 7.10E-04 | 1.67E-05 |
| 197 | 7 | 25952189 | -3.41152 | 3.081049 | 2.56E-02 | 1 | 2.56E-02 | 6.46E-04 | 2.06E-03 |
| 198 | 7 | 27786969 | -3.58004 | 3.676883 | 1.48E-02 | 1 | 1.48E-02 | 3.44E-04 | 2.36E-04 |
| 199 | 7 | 28009799 | 3.770358 | 3.339679 | 1.01E-02 | 1 | 1.01E-02 | 1.63E-04 | 8.39E-04 |
| 200 | 7 | 28250599 | 5.214525 | -4.10026 | 7.50E-04 | 1 | 7.50E-04 | 1.84E-07 | 4.13E-05 |
| 201 | 7 | 38134467 | 3.300809 | -3.27777 | 3.11E-02 | 1 | 3.11E-02 | 9.64E-04 | 1.05E-03 |
| 202 | 7 | 40911632 | 3.759289 | 3.180465 | 1.59E-02 | 1 | 1.59E-02 | 1.70E-04 | 1.47E-03 |
| 203 | 7 | 44808091 | 3.500593 | 3.12107 | 1.98E-02 | 1 | 1.98E-02 | 4.64E-04 | 1.80E-03 |
| 204 | 7 | 45161303 | 4.033255 | 2.980483 | 2.73E-02 | 1 | 2.73E-02 | 5.50E-05 | 2.88E-03 |
| 205 | 7 | 51059049 | -3.77723 | -3.01695 | 2.48E-02 | 1 | 2.48E-02 | 1.59E-04 | 2.55E-03 |
| 206 | 7 | 55773037 | 3.413989 | 3.369315 | 2.31E-02 | 1 | 2.31E-02 | 6.40E-04 | 7.54E-04 |
| 206 | 7 | 55980652 | 3.831328 | 3.577127 | 7.04E-03 | 1 | 7.04E-03 | 1.27E-04 | 3.47E-04 |
| 207 | 7 | 69789374 | 3.950033 | -3.67688 | 4.81E-03 | 1 | 4.81E-03 | 7.81E-05 | 2.36E-04 |
| 208 | 7 | 74108135 | 5.030076 | -4.71766 | 6.16E-05 | 1 | 6.16E-05 | 4.90E-07 | 2.39E-06 |
| 209 | 7 | 76568671 | 3.454665 | 2.936343 | 3.06E-02 | 1 | 3.06E-02 | 5.51E-04 | 3.32E-03 |
| 210 | 7 | 83794493 | 3.515584 | -3.73069 | 1.76E-02 | 1 | 1.76E-02 | 4.39E-04 | 1.91E-04 |
| 211 | 7 | 92286980 | -3.63668 | -7.5324 | 1.26E-02 | 1 | 1.26E-02 | 2.76E-04 | 4.98E-14 |
| 212 | 7 | 99171449 | -3.61905 | -4.43101 | 1.32E-02 | 1 | 1.32E-02 | 2.96E-04 | 9.38E-06 |
| 213 | 7 | 100198666 | 3.602569 | 2.892044 | 3.42E-02 | 1 | 3.42E-02 | 3.15E-04 | 3.83E-03 |
| 213 | 7 | 100342672 | 3.76203 | 4.917831 | 8.72E-03 | 1 | 8.72E-03 | 1.69E-04 | 8.75E-07 |
| 213 | 7 | 100373832 | -3.39306 | -2.93234 | 3.09E-02 | 1 | 3.09E-02 | 6.91E-04 | 3.36E-03 |
| 213 | 7 | 100414386 | -4.12465 | -5.19361 | 2.63E-03 | 1 | 2.63E-03 | 3.71E-05 | 2.06E-07 |
| 214 | 7 | 102113093 | 3.491501 | -2.92451 | 3.15E-02 | 1 | 3.15E-02 | 4.80E-04 | 3.45E-03 |
| 215 | 7 | 103417134 | 4.592633 | 3.837572 | 1.98E-03 | 1 | 1.98E-03 | 4.38E-06 | 1.24E-04 |
| 216 | 7 | 118861979 | -3.23318 | -3.2959 | 3.65E-02 | 1 | 3.65E-02 | 1.22E-03 | 9.81E-04 |
| 217 | 7 | 130028723 | 3.849783 | -2.92451 | 3.15E-02 | 1 | 3.15E-02 | 1.18E-04 | 3.45E-03 |
| 218 | 7 | 130887687 | 3.145862 | 5.025636 | 4.52E-02 | 1 | 4.52E-02 | 1.66E-03 | 5.02E-07 |
| 219 | 7 | 143166588 | 3.591963 | 3.066772 | 2.18E-02 | 1 | 2.18E-02 | 3.28E-04 | 2.16E-03 |
| 220 | 7 | 150536681 | -3.60082 | -3.16944 | 1.64E-02 | 1 | 1.64E-02 | 3.17E-04 | 1.53E-03 |
| 220 | 7 | 150540196 | 5.587564 | 3.059944 | 2.22E-02 | 1 | 2.22E-02 | 2.30E-08 | 2.21E-03 |
| 221 | 8 | 8171978 | 3.155882 | 3.380172 | 4.40E-02 | 1 | 4.40E-02 | 1.60E-03 | 7.24E-04 |
| 221 | 8 | 8236231 | 3.886732 | 8.7884 | 5.91E-03 | 1 | 5.91E-03 | 1.02E-04 | 1.52E-18 |
| 221 | 8 | 8540189 | 3.604479 | 5.311217 | 1.38E-02 | 1 | 1.38E-02 | 3.13E-04 | 1.09E-07 |
| 221 | 8 | 8570452 | 3.295666 | 3.277765 | 3.14E-02 | 1 | 3.14E-02 | 9.82E-04 | 1.05E-03 |
| 221 | 8 | 8574282 | -4.25053 | -7.7477 | 1.65E-03 | 1 | 1.65E-03 | 2.13E-05 | 9.36E-15 |
| 221 | 8 | 8610267 | 4.696906 | 10.94454 | 2.73E-04 | 1 | 2.73E-04 | 2.64E-06 | 7.06E-28 |
| 221 | 8 | 8912911 | 3.166524 | 4.498765 | 4.30E-02 | 1 | 4.30E-02 | 1.54E-03 | 6.83E-06 |
| 221 | 8 | 8922271 | -3.58205 | -9.03305 | 1.47E-02 | 1 | 1.47E-02 | 3.41E-04 | 1.67E-19 |
| 221 | 8 | 8966606 | -3.12673 | -5.31715 | 4.73E-02 | 1 | 4.73E-02 | 1.77E-03 | 1.05E-07 |
| 221 | 8 | 9030160 | 3.590983 | 6.320768 | 1.43E-02 | 1 | 1.43E-02 | 3.29E-04 | 2.60E-10 |
| 221 | 8 | 9049381 | 5.774907 | 9.724433 | 1.57E-06 | 1 | 1.57E-06 | 7.70E-09 | 2.37E-22 |
| 221 | 8 | 9154394 | 3.826074 | 8.566586 | 7.16E-03 | 1 | 7.16E-03 | 1.30E-04 | 1.07E-17 |
| 221 | 8 | 9181611 | 5.661315 | 10.02906 | 2.87E-06 | 1 | 2.87E-06 | 1.50E-08 | 1.14E-23 |
| 221 | 8 | 9216584 | 4.080414 | 3.584407 | 4.73E-03 | 1 | 4.73E-03 | 4.50E-05 | 3.38E-04 |
| 221 | 8 | 9261601 | 4.040278 | 4.195949 | 3.54E-03 | 1 | 3.54E-03 | 5.34E-05 | 2.72E-05 |
| 221 | 8 | 9526032 | 3.255474 | 3.544187 | 3.46E-02 | 1 | 3.46E-02 | 1.13E-03 | 3.94E-04 |
| 221 | 8 | 9675483 | 4.384925 | 6.11045 | 9.84E-04 | 1 | 9.84E-04 | 1.16E-05 | 9.94E-10 |
| 221 | 8 | 9775120 | 4.790853 | 7.376584 | 1.80E-04 | 1 | 1.80E-04 | 1.66E-06 | 1.62E-13 |
| 221 | 8 | 9828897 | -4.661 | -3.13922 | 1.79E-02 | 1 | 1.79E-02 | 3.15E-06 | 1.69E-03 |
| 221 | 8 | 9879283 | 4.282722 | 8.145607 | 1.46E-03 | 1 | 1.46E-03 | 1.85E-05 | 3.77E-16 |
| 221 | 8 | 9996389 | -7.00598 | -9.93486 | 8.74E-10 | 1 | 8.74E-10 | 2.45E-12 | 2.94E-23 |
| 221 | 8 | 10065961 | 3.255872 | 4.712912 | 3.46E-02 | 1 | 3.46E-02 | 1.13E-03 | 2.44E-06 |
| 221 | 8 | 10091194 | 3.140739 | 5.813582 | 4.57E-02 | 1 | 4.57E-02 | 1.69E-03 | 6.12E-09 |
| 221 | 8 | 10227910 | 3.144296 | 4.286852 | 4.53E-02 | 1 | 4.53E-02 | 1.66E-03 | 1.81E-05 |
| 221 | 8 | 10277978 | -4.7148 | -6.32077 | 2.52E-04 | 1 | 2.52E-04 | 2.42E-06 | 2.60E-10 |
| 221 | 8 | 10345972 | -4.04753 | -4.81548 | 3.45E-03 | 1 | 3.45E-03 | 5.18E-05 | 1.47E-06 |
| 221 | 8 | 10375365 | 3.722317 | 3.261004 | 1.27E-02 | 1 | 1.27E-02 | 1.97E-04 | 1.11E-03 |
| 221 | 8 | 10445702 | 3.946427 | 6.585252 | 4.86E-03 | 1 | 4.86E-03 | 7.93E-05 | 4.54E-11 |
| 221 | 8 | 10697015 | 5.78431 | 10.00556 | 1.50E-06 | 1 | 1.50E-06 | 7.28E-09 | 1.44E-23 |
| 221 | 8 | 10831752 | 3.632031 | 4.545885 | 1.28E-02 | 1 | 1.28E-02 | 2.81E-04 | 5.47E-06 |
| 221 | 8 | 10854864 | -5.31752 | -6.39858 | 1.54E-05 | 1 | 1.54E-05 | 1.05E-07 | 1.57E-10 |
| 221 | 8 | 10876759 | 3.268822 | 5.287316 | 3.35E-02 | 1 | 3.35E-02 | 1.08E-03 | 1.24E-07 |
| 221 | 8 | 10907051 | 3.766981 | 5.069776 | 8.58E-03 | 1 | 8.58E-03 | 1.65E-04 | 3.98E-07 |
| 221 | 8 | 11079437 | 3.210198 | 5.499005 | 3.87E-02 | 1 | 3.87E-02 | 1.33E-03 | 3.82E-08 |
| 221 | 8 | 11136683 | -4.63636 | -9.41015 | 3.54E-04 | 1 | 3.54E-04 | 3.55E-06 | 4.95E-21 |
| 221 | 8 | 11217572 | 3.217919 | 4.389258 | 3.79E-02 | 1 | 3.79E-02 | 1.29E-03 | 1.14E-05 |
| 221 | 8 | 11290059 | -3.55773 | -7.83744 | 1.57E-02 | 1 | 1.57E-02 | 3.74E-04 | 4.60E-15 |
| 221 | 8 | 11347660 | 3.990989 | 8.670233 | 4.19E-03 | 1 | 4.19E-03 | 6.58E-05 | 4.31E-18 |
| 221 | 8 | 11367357 | 4.049995 | 3.481684 | 6.53E-03 | 1 | 6.53E-03 | 5.12E-05 | 4.98E-04 |
| 221 | 8 | 11389783 | 3.373642 | 9.004308 | 2.57E-02 | 1 | 2.57E-02 | 7.42E-04 | 2.17E-19 |
| 221 | 8 | 11441789 | 3.842625 | 4.415882 | 6.80E-03 | 1 | 6.80E-03 | 1.22E-04 | 1.01E-05 |
| 221 | 8 | 11496687 | 3.364226 | 3.277765 | 2.64E-02 | 1 | 2.64E-02 | 7.68E-04 | 1.05E-03 |
| 221 | 8 | 11657716 | 3.537552 | 6.11045 | 1.66E-02 | 1 | 1.66E-02 | 4.04E-04 | 9.94E-10 |
| 221 | 8 | 11752021 | -3.14314 | -6.55903 | 4.55E-02 | 1 | 4.55E-02 | 1.67E-03 | 5.42E-11 |
| 221 | 8 | 11828303 | 4.343255 | 6.209058 | 1.16E-03 | 1 | 1.16E-03 | 1.40E-05 | 5.33E-10 |
| 221 | 8 | 11846083 | -3.53733 | -4.42327 | 1.66E-02 | 1 | 1.66E-02 | 4.04E-04 | 9.72E-06 |
| 222 | 8 | 36846109 | 3.368503 | -2.78134 | 4.48E-02 | 1 | 4.48E-02 | 7.56E-04 | 5.41E-03 |
| 223 | 8 | 106417183 | 3.128759 | 3.245416 | 4.80E-02 | 1 | 4.80E-02 | 1.76E-03 | 1.17E-03 |
| 224 | 8 | 116645056 | 4.924823 | 4.70831 | 9.80E-05 | 1 | 9.80E-05 | 8.44E-07 | 2.50E-06 |
| 224 | 8 | 116679547 | -4.36615 | 5.840834 | 1.06E-03 | 1 | 1.06E-03 | 1.26E-05 | 5.19E-09 |
| 224 | 8 | 116980628 | -3.39583 | 4.304405 | 2.42E-02 | 1 | 2.42E-02 | 6.84E-04 | 1.67E-05 |
| 224 | 8 | 117111562 | 3.110463 | -3.261 | 4.98E-02 | 1 | 4.98E-02 | 1.87E-03 | 1.11E-03 |
| 225 | 8 | 119250802 | -3.2053 | -3.58441 | 3.92E-02 | 1 | 3.92E-02 | 1.35E-03 | 3.38E-04 |
| 226 | 8 | 121061879 | 3.706398 | 4.36633 | 1.03E-02 | 1 | 1.03E-02 | 2.10E-04 | 1.26E-05 |
| 227 | 8 | 129397730 | 3.896267 | -3.46861 | 6.82E-03 | 1 | 6.82E-03 | 9.77E-05 | 5.23E-04 |
| 228 | 8 | 129596278 | 3.390709 | 3.464429 | 2.46E-02 | 1 | 2.46E-02 | 6.97E-04 | 5.31E-04 |
| 229 | 8 | 129627838 | 3.650102 | 4.250919 | 1.21E-02 | 1 | 1.21E-02 | 2.62E-04 | 2.13E-05 |
| 230 | 8 | 131482432 | 3.163488 | 3.771941 | 4.33E-02 | 1 | 4.33E-02 | 1.56E-03 | 1.62E-04 |
| 231 | 8 | 145536056 | 6.496718 | 4.498765 | 1.55E-04 | 1 | 1.55E-04 | 8.21E-11 | 6.83E-06 |
| 232 | 8 | 145666664 | -3.62172 | -3.23084 | 1.38E-02 | 1 | 1.38E-02 | 2.93E-04 | 1.23E-03 |
| 233 | 9 | 13235045 | -3.13994 | -5.58781 | 4.58E-02 | 1 | 4.58E-02 | 1.69E-03 | 2.30E-08 |
| 234 | 9 | 13989084 | 3.116762 | -3.82963 | 4.84E-02 | 1 | 4.84E-02 | 1.83E-03 | 1.28E-04 |
| 235 | 9 | 23352293 | 3.370547 | 2.781336 | 4.48E-02 | 1 | 4.48E-02 | 7.50E-04 | 5.41E-03 |
| 236 | 9 | 34074476 | 5.822847 | 4.790245 | 4.49E-05 | 1 | 4.49E-05 | 5.79E-09 | 1.67E-06 |
| 237 | 9 | 35726821 | 3.733129 | -4.29366 | 9.50E-03 | 1 | 9.50E-03 | 1.89E-04 | 1.76E-05 |
| 238 | 9 | 86570075 | -3.98084 | -4.30441 | 4.35E-03 | 1 | 4.35E-03 | 6.87E-05 | 1.67E-05 |
| 239 | 9 | 96454513 | 4.280907 | 9.901997 | 1.47E-03 | 1 | 1.47E-03 | 1.86E-05 | 4.08E-23 |
| 240 | 9 | 96812138 | 3.750554 | 7.412021 | 9.03E-03 | 1 | 9.03E-03 | 1.76E-04 | 1.24E-13 |
| 240 | 9 | 96892988 | -3.19388 | -5.06978 | 4.03E-02 | 1 | 4.03E-02 | 1.40E-03 | 3.98E-07 |
| 240 | 9 | 96904554 | -4.07705 | -3.32409 | 1.06E-02 | 1 | 1.06E-02 | 4.56E-05 | 8.87E-04 |
| 240 | 9 | 96970677 | 5.250613 | 7.546094 | 2.11E-05 | 1 | 2.11E-05 | 1.52E-07 | 4.49E-14 |
| 240 | 9 | 97415022 | 3.234168 | 4.319893 | 3.64E-02 | 1 | 3.64E-02 | 1.22E-03 | 1.56E-05 |
| 241 | 9 | 97838083 | -3.81121 | -3.02847 | 2.41E-02 | 1 | 2.41E-02 | 1.38E-04 | 2.46E-03 |
| 241 | 9 | 97873957 | 4.871781 | 4.431006 | 2.04E-04 | 1 | 2.04E-04 | 1.11E-06 | 9.38E-06 |
| 241 | 9 | 98143402 | -3.81207 | -5.88511 | 7.49E-03 | 1 | 7.49E-03 | 1.38E-04 | 3.98E-09 |
| 242 | 9 | 110003827 | -3.45533 | 3.437276 | 2.07E-02 | 1 | 2.07E-02 | 5.50E-04 | 5.88E-04 |
| 243 | 9 | 113499263 | 3.167689 | -3.82963 | 4.28E-02 | 1 | 4.28E-02 | 1.54E-03 | 1.28E-04 |
| 244 | 9 | 118272452 | -3.69197 | 4.835004 | 1.07E-02 | 1 | 1.07E-02 | 2.23E-04 | 1.33E-06 |
| 245 | 9 | 124625886 | 3.180061 | 3.073803 | 4.58E-02 | 1 | 4.58E-02 | 1.47E-03 | 2.11E-03 |
| 246 | 9 | 125789160 | -4.32305 | -3.57713 | 4.84E-03 | 1 | 4.84E-03 | 1.54E-05 | 3.47E-04 |
| 247 | 9 | 126316511 | 3.752822 | -3.14886 | 1.74E-02 | 1 | 1.74E-02 | 1.75E-04 | 1.64E-03 |
| 248 | 9 | 126617260 | 3.300342 | -2.78134 | 4.48E-02 | 1 | 4.48E-02 | 9.66E-04 | 5.41E-03 |
| 249 | 9 | 129258837 | 3.369403 | 3.040563 | 2.91E-02 | 1 | 2.91E-02 | 7.53E-04 | 2.36E-03 |
| 250 | 9 | 129408290 | -3.46762 | -3.5998 | 2.01E-02 | 1 | 2.01E-02 | 5.25E-04 | 3.18E-04 |
| 251 | 9 | 130576776 | 3.164468 | 5.38188 | 4.32E-02 | 1 | 4.32E-02 | 1.55E-03 | 7.37E-08 |
| 252 | 9 | 136149500 | 6.528184 | -6.24202 | 2.00E-08 | 1 | 2.00E-08 | 6.66E-11 | 4.32E-10 |
| 252 | 9 | 136191010 | 3.472085 | -4.29716 | 1.98E-02 | 1 | 1.98E-02 | 5.16E-04 | 1.73E-05 |
| 253 | 9 | 139239807 | 3.230544 | -3.53237 | 3.67E-02 | 1 | 3.67E-02 | 1.24E-03 | 4.12E-04 |
| 254 | 10 | 77634234 | -4.65104 | -5.86301 | 3.32E-04 | 1 | 3.32E-04 | 3.30E-06 | 4.55E-09 |
| 255 | 10 | 80923862 | -5.13672 | -2.86634 | 3.64E-02 | 1 | 3.64E-02 | 2.80E-07 | 4.15E-03 |
| 256 | 10 | 81078944 | 3.164468 | -4.49877 | 4.32E-02 | 1 | 4.32E-02 | 1.55E-03 | 6.83E-06 |
| 257 | 10 | 89581975 | -3.13055 | -4.59018 | 4.69E-02 | 1 | 4.69E-02 | 1.74E-03 | 4.43E-06 |
| 258 | 10 | 89774578 | -3.14154 | -4.43913 | 4.57E-02 | 1 | 4.57E-02 | 1.68E-03 | 9.03E-06 |
| 259 | 10 | 99118929 | 4.455365 | 3.21716 | 1.44E-02 | 1 | 1.44E-02 | 8.38E-06 | 1.29E-03 |
| 260 | 10 | 101912064 | -4.68239 | 3.490878 | 6.36E-03 | 1 | 6.36E-03 | 2.84E-06 | 4.81E-04 |
| 261 | 10 | 103373593 | -3.58205 | -3.08105 | 2.09E-02 | 1 | 2.09E-02 | 3.41E-04 | 2.06E-03 |
| 261 | 10 | 103950714 | -3.46581 | -3.342 | 2.02E-02 | 1 | 2.02E-02 | 5.29E-04 | 8.32E-04 |
| 262 | 10 | 114379753 | -3.60522 | -4.43101 | 1.37E-02 | 1 | 1.37E-02 | 3.12E-04 | 9.38E-06 |
| 263 | 10 | 114593006 | -4.14054 | -3.57713 | 4.84E-03 | 1 | 4.84E-03 | 3.46E-05 | 3.47E-04 |
| 264 | 10 | 114598202 | 9.028079 | 4.02685 | 9.93E-04 | 1 | 9.93E-04 | 1.75E-19 | 5.65E-05 |
| 264 | 10 | 114668045 | -3.57303 | -3.89752 | 1.51E-02 | 1 | 1.51E-02 | 3.53E-04 | 9.72E-05 |
| 264 | 10 | 114711883 | -7.7199 | -4.22125 | 4.70E-04 | 1 | 4.70E-04 | 1.16E-14 | 2.43E-05 |
| 264 | 10 | 114723845 | -3.21343 | -3.96387 | 3.84E-02 | 1 | 3.84E-02 | 1.31E-03 | 7.37E-05 |
| 264 | 10 | 114725079 | -5.73424 | -3.44468 | 7.35E-03 | 1 | 7.35E-03 | 9.80E-09 | 5.72E-04 |
| 264 | 10 | 114727067 | 4.672985 | -7.97107 | 3.02E-04 | 1 | 3.02E-04 | 2.97E-06 | 1.57E-15 |
| 264 | 10 | 114733456 | 20.16346 | 6.38296 | 8.44E-09 | 1 | 8.44E-09 | 2.05E-90 | 1.74E-10 |
| 264 | 10 | 114793572 | 15.30869 | 3.295899 | 1.14E-02 | 1 | 1.14E-02 | 6.69E-53 | 9.81E-04 |
| 264 | 10 | 114821527 | 23.0339 | 4.012386 | 1.05E-03 | 1 | 1.05E-03 | 2.13E-117 | 6.01E-05 |
| 265 | 10 | 114834411 | 7.828076 | 2.760255 | 4.70E-02 | 1 | 4.70E-02 | 4.95E-15 | 5.78E-03 |
| 266 | 10 | 114837575 | 5.076061 | 3.413218 | 8.07E-03 | 1 | 8.07E-03 | 3.85E-07 | 6.42E-04 |
| 266 | 10 | 114854383 | -11.7997 | -2.9024 | 3.33E-02 | 1 | 3.33E-02 | 3.92E-32 | 3.70E-03 |
| 267 | 10 | 114886511 | 3.227398 | -3.19205 | 3.86E-02 | 1 | 3.86E-02 | 1.25E-03 | 1.41E-03 |
| 268 | 10 | 115781367 | -4.3861 | -3.33286 | 1.03E-02 | 1 | 1.03E-02 | 1.15E-05 | 8.60E-04 |
| 269 | 10 | 122859270 | 3.177483 | -3.15892 | 4.43E-02 | 1 | 4.43E-02 | 1.49E-03 | 1.58E-03 |
| 270 | 10 | 124096306 | 4.936741 | 4.371772 | 2.59E-04 | 1 | 2.59E-04 | 7.94E-07 | 1.23E-05 |
| 270 | 10 | 124154626 | 4.271218 | -3.36152 | 9.44E-03 | 1 | 9.44E-03 | 1.94E-05 | 7.75E-04 |
| 271 | 10 | 134011002 | 3.428936 | 3.584407 | 2.22E-02 | 1 | 2.22E-02 | 6.06E-04 | 3.38E-04 |
| 272 | 10 | 134376691 | 3.315849 | 8.061456 | 2.98E-02 | 1 | 2.98E-02 | 9.14E-04 | 7.54E-16 |
| 273 | 11 | 2077271 | 5.105618 | 2.878865 | 3.52E-02 | 1 | 3.52E-02 | 3.30E-07 | 3.99E-03 |
| 274 | 11 | 8390036 | -3.54283 | -2.88537 | 3.47E-02 | 1 | 3.47E-02 | 3.96E-04 | 3.91E-03 |
| 275 | 11 | 9834403 | -3.69683 | 3.053307 | 2.26E-02 | 1 | 2.26E-02 | 2.18E-04 | 2.26E-03 |
| 276 | 11 | 16238644 | 3.176669 | -3.27777 | 4.22E-02 | 1 | 4.22E-02 | 1.49E-03 | 1.05E-03 |
| 277 | 11 | 16440714 | 3.547384 | 4.768616 | 1.61E-02 | 1 | 1.61E-02 | 3.89E-04 | 1.85E-06 |
| 277 | 11 | 16798672 | -4.47973 | -4.7982 | 6.75E-04 | 1 | 6.75E-04 | 7.47E-06 | 1.60E-06 |
| 278 | 11 | 27261193 | 3.126472 | 6.628139 | 4.74E-02 | 1 | 4.74E-02 | 1.77E-03 | 3.40E-11 |
| 278 | 11 | 27332151 | 3.420085 | 6.520057 | 2.27E-02 | 1 | 2.27E-02 | 6.26E-04 | 7.03E-11 |
| 278 | 11 | 27393813 | 3.664661 | 7.478783 | 1.16E-02 | 1 | 1.16E-02 | 2.48E-04 | 7.50E-14 |
| 278 | 11 | 27686196 | 4.632974 | 4.815481 | 3.59E-04 | 1 | 3.59E-04 | 3.60E-06 | 1.47E-06 |
| 279 | 11 | 28342947 | 3.375909 | -3.15892 | 2.71E-02 | 1 | 2.71E-02 | 7.36E-04 | 1.58E-03 |
| 280 | 11 | 34811180 | -3.4147 | -3.99265 | 2.30E-02 | 1 | 2.30E-02 | 6.39E-04 | 6.53E-05 |
| 281 | 11 | 46100914 | 3.874786 | -3.261 | 1.27E-02 | 1 | 1.27E-02 | 1.07E-04 | 1.11E-03 |
| 281 | 11 | 47065072 | -3.65947 | -3.18047 | 1.59E-02 | 1 | 1.59E-02 | 2.53E-04 | 1.47E-03 |
| 281 | 11 | 47899030 | -4.48792 | -3.40682 | 8.24E-03 | 1 | 8.24E-03 | 7.19E-06 | 6.57E-04 |
| 281 | 11 | 48260401 | 4.562729 | -4.67209 | 4.79E-04 | 1 | 4.79E-04 | 5.05E-06 | 2.98E-06 |
| 281 | 11 | 54893730 | 4.278565 | -3.83757 | 1.98E-03 | 1 | 1.98E-03 | 1.88E-05 | 1.24E-04 |
| 281 | 11 | 57268252 | 4.317021 | -4.17216 | 1.28E-03 | 1 | 1.28E-03 | 1.58E-05 | 3.02E-05 |
| 282 | 11 | 58274155 | -3.40935 | 3.053307 | 2.61E-02 | 1 | 2.61E-02 | 6.51E-04 | 2.26E-03 |
| 283 | 11 | 61126858 | 3.620458 | 3.92556 | 1.32E-02 | 1 | 1.32E-02 | 2.94E-04 | 8.65E-05 |
| 284 | 11 | 61565908 | -4.37735 | 5.890041 | 1.01E-03 | 1 | 1.01E-03 | 1.20E-05 | 3.86E-09 |
| 285 | 11 | 62199457 | 3.429386 | 4.28355 | 2.22E-02 | 1 | 2.22E-02 | 6.05E-04 | 1.84E-05 |
| 286 | 11 | 62248152 | -3.38543 | 3.028465 | 2.81E-02 | 1 | 2.81E-02 | 7.11E-04 | 2.46E-03 |
| 287 | 11 | 64096725 | 3.759289 | 3.14886 | 1.74E-02 | 1 | 1.74E-02 | 1.70E-04 | 1.64E-03 |
| 288 | 11 | 65294799 | 7.205528 | 8.49121 | 2.16E-10 | 1 | 2.16E-10 | 5.78E-13 | 2.04E-17 |
| 289 | 11 | 65390803 | -4.67374 | -9.21743 | 3.01E-04 | 1 | 3.01E-04 | 2.96E-06 | 3.04E-20 |
| 289 | 11 | 65604862 | -3.32673 | -3.1125 | 3.14E-02 | 1 | 3.14E-02 | 8.79E-04 | 1.86E-03 |
| 290 | 11 | 66158466 | -3.31467 | -3.46861 | 2.99E-02 | 1 | 2.99E-02 | 9.18E-04 | 5.23E-04 |
| 291 | 11 | 68100519 | 3.295666 | 3.897519 | 3.13E-02 | 1 | 3.13E-02 | 9.82E-04 | 9.72E-05 |
| 292 | 11 | 68602753 | -3.76424 | 5.223953 | 8.66E-03 | 1 | 8.66E-03 | 1.67E-04 | 1.75E-07 |
| 293 | 11 | 68811777 | -3.22729 | -3.5998 | 3.70E-02 | 1 | 3.70E-02 | 1.25E-03 | 3.18E-04 |
| 293 | 11 | 68839334 | 4.997975 | 11.63999 | 6.96E-05 | 1 | 6.96E-05 | 5.79E-07 | 2.58E-31 |
| 293 | 11 | 68932978 | 4.217125 | 6.257267 | 1.87E-03 | 1 | 1.87E-03 | 2.47E-05 | 3.92E-10 |
| 294 | 11 | 69451864 | -5.86699 | -3.58441 | 4.73E-03 | 1 | 4.73E-03 | 4.44E-09 | 3.38E-04 |
| 294 | 11 | 69452559 | 4.415513 | 3.515817 | 5.89E-03 | 1 | 5.89E-03 | 1.01E-05 | 4.38E-04 |
| 295 | 11 | 72462375 | -7.40316 | -3.31984 | 1.07E-02 | 1 | 1.07E-02 | 1.33E-13 | 9.01E-04 |
| 295 | 11 | 72774409 | -3.73447 | -2.80427 | 4.23E-02 | 1 | 4.23E-02 | 1.88E-04 | 5.04E-03 |
| 296 | 11 | 77507420 | 3.210808 | 3.081049 | 4.24E-02 | 1 | 4.24E-02 | 1.32E-03 | 2.06E-03 |
| 297 | 11 | 84778852 | 3.607745 | 3.016948 | 2.48E-02 | 1 | 2.48E-02 | 3.09E-04 | 2.55E-03 |
| 298 | 11 | 92681013 | 11.71324 | 3.440947 | 7.44E-03 | 1 | 7.44E-03 | 1.09E-31 | 5.80E-04 |
| 299 | 11 | 100572709 | -3.18024 | -3.04685 | 4.64E-02 | 1 | 4.64E-02 | 1.47E-03 | 2.31E-03 |
| 300 | 11 | 103020971 | -4.36718 | -3.261 | 1.27E-02 | 1 | 1.27E-02 | 1.26E-05 | 1.11E-03 |
| 301 | 11 | 112913053 | -3.3694 | -6.19236 | 2.59E-02 | 1 | 2.59E-02 | 7.53E-04 | 5.93E-10 |
| 302 | 11 | 113253899 | 3.294201 | 3.261004 | 3.18E-02 | 1 | 3.18E-02 | 9.87E-04 | 1.11E-03 |
| 303 | 11 | 115009670 | 4.024133 | -12.4567 | 3.75E-03 | 1 | 3.75E-03 | 5.72E-05 | 1.29E-35 |
| 304 | 11 | 116707338 | 3.591963 | 3.245416 | 1.46E-02 | 1 | 1.46E-02 | 3.28E-04 | 1.17E-03 |
| 305 | 11 | 121932484 | 3.852976 | -4.20604 | 6.59E-03 | 1 | 6.59E-03 | 1.17E-04 | 2.60E-05 |
| 306 | 11 | 125110079 | -3.29206 | 3.19205 | 3.29E-02 | 1 | 3.29E-02 | 9.95E-04 | 1.41E-03 |
| 307 | 12 | 367902 | 3.776902 | -3.08105 | 2.09E-02 | 1 | 2.09E-02 | 1.59E-04 | 2.06E-03 |
| 308 | 12 | 4384844 | -16.745 | -8.75783 | 2.35E-16 | 1 | 2.35E-16 | 6.16E-63 | 1.99E-18 |
| 308 | 12 | 4399050 | 6.88906 | 3.330636 | 1.04E-02 | 1 | 1.04E-02 | 5.62E-12 | 8.66E-04 |
| 309 | 12 | 6712980 | 4.353808 | 3.676883 | 3.48E-03 | 1 | 3.48E-03 | 1.34E-05 | 2.36E-04 |
| 310 | 12 | 12479837 | -3.49331 | -3.24542 | 1.91E-02 | 1 | 1.91E-02 | 4.77E-04 | 1.17E-03 |
| 311 | 12 | 13224027 | 3.199273 | -3.01695 | 4.49E-02 | 1 | 4.49E-02 | 1.38E-03 | 2.55E-03 |
| 312 | 12 | 13269946 | 3.162054 | 5.23352 | 4.34E-02 | 1 | 4.34E-02 | 1.57E-03 | 1.66E-07 |
| 313 | 12 | 14751319 | -3.28262 | -2.9024 | 3.88E-02 | 1 | 3.88E-02 | 1.03E-03 | 3.70E-03 |
| 314 | 12 | 19702802 | -3.12876 | 3.556733 | 4.71E-02 | 1 | 4.71E-02 | 1.76E-03 | 3.75E-04 |
| 315 | 12 | 26358170 | -3.47906 | -3.261 | 1.97E-02 | 1 | 1.97E-02 | 5.03E-04 | 1.11E-03 |
| 315 | 12 | 26398707 | -5.20598 | -5.80067 | 2.60E-05 | 1 | 2.60E-05 | 1.93E-07 | 6.60E-09 |
| 315 | 12 | 26457190 | 6.043966 | 9.356936 | 3.54E-07 | 1 | 3.54E-07 | 1.50E-09 | 8.21E-21 |
| 316 | 12 | 27945518 | 4.155514 | 2.740738 | 4.91E-02 | 1 | 4.91E-02 | 3.25E-05 | 6.13E-03 |
| 317 | 12 | 33585465 | 4.005033 | 2.95306 | 2.92E-02 | 1 | 2.92E-02 | 6.20E-05 | 3.15E-03 |
| 318 | 12 | 33593127 | -3.6501 | -4.36633 | 1.21E-02 | 1 | 1.21E-02 | 2.62E-04 | 1.26E-05 |
| 319 | 12 | 39426137 | 4.341935 | 3.746825 | 2.74E-03 | 1 | 2.74E-03 | 1.41E-05 | 1.79E-04 |
| 320 | 12 | 46213867 | 3.759674 | 2.878865 | 3.52E-02 | 1 | 3.52E-02 | 1.70E-04 | 3.99E-03 |
| 321 | 12 | 48512285 | -3.3928 | 3.998993 | 2.44E-02 | 1 | 2.44E-02 | 6.92E-04 | 6.36E-05 |
| 321 | 12 | 48582961 | 4.425199 | -5.30553 | 8.38E-04 | 1 | 8.38E-04 | 9.64E-06 | 1.12E-07 |
| 322 | 12 | 48815243 | 3.56144 | -4.40881 | 1.55E-02 | 1 | 1.55E-02 | 3.69E-04 | 1.04E-05 |
| 323 | 12 | 50263148 | 4.881396 | 4.691161 | 1.20E-04 | 1 | 1.20E-04 | 1.05E-06 | 2.72E-06 |
| 324 | 12 | 51159140 | -3.27442 | 2.740738 | 4.98E-02 | 1 | 4.98E-02 | 1.06E-03 | 6.13E-03 |
| 325 | 12 | 66254930 | 4.790853 | 3.104232 | 1.97E-02 | 1 | 1.97E-02 | 1.66E-06 | 1.91E-03 |
| 326 | 12 | 66371880 | -8.04833 | -5.62504 | 6.94E-07 | 1 | 6.94E-07 | 8.39E-16 | 1.85E-08 |
| 327 | 12 | 69653146 | 3.50981 | 2.909556 | 3.28E-02 | 1 | 3.28E-02 | 4.48E-04 | 3.62E-03 |
| 328 | 12 | 71656547 | 4.634046 | 2.843004 | 3.86E-02 | 1 | 3.86E-02 | 3.59E-06 | 4.47E-03 |
| 329 | 12 | 89909623 | 3.538803 | 3.591958 | 1.65E-02 | 1 | 1.65E-02 | 4.02E-04 | 3.28E-04 |
| 330 | 12 | 90413738 | -3.25016 | -10.2974 | 3.51E-02 | 1 | 3.51E-02 | 1.15E-03 | 7.24E-25 |
| 330 | 12 | 90440798 | -3.47952 | -2.95306 | 2.92E-02 | 1 | 2.92E-02 | 5.02E-04 | 3.15E-03 |
| 331 | 12 | 90955246 | 3.144216 | 6.475478 | 4.54E-02 | 1 | 4.54E-02 | 1.67E-03 | 9.45E-11 |
| 332 | 12 | 93968256 | 3.485393 | 3.104232 | 2.08E-02 | 1 | 2.08E-02 | 4.91E-04 | 1.91E-03 |
| 333 | 12 | 95842603 | 4.471439 | -2.85734 | 3.73E-02 | 1 | 3.73E-02 | 7.77E-06 | 4.27E-03 |
| 334 | 12 | 102824921 | -3.1212 | -3.34911 | 4.79E-02 | 1 | 4.79E-02 | 1.80E-03 | 8.11E-04 |
| 335 | 12 | 111910219 | 3.279441 | 2.83209 | 4.07E-02 | 1 | 4.07E-02 | 1.04E-03 | 4.62E-03 |
| 336 | 12 | 121675307 | 3.303016 | 3.104232 | 3.34E-02 | 1 | 3.34E-02 | 9.57E-04 | 1.91E-03 |
| 337 | 12 | 123593382 | -5.55714 | -5.37245 | 4.88E-06 | 1 | 4.88E-06 | 2.74E-08 | 7.77E-08 |
| 338 | 12 | 124306729 | 3.782625 | -3.44468 | 8.18E-03 | 1 | 8.18E-03 | 1.55E-04 | 5.72E-04 |
| 338 | 12 | 124458002 | -4.45038 | 5.546546 | 7.60E-04 | 1 | 7.60E-04 | 8.57E-06 | 2.91E-08 |
| 338 | 12 | 124510391 | -5.20909 | 6.13076 | 2.56E-05 | 1 | 2.56E-05 | 1.90E-07 | 8.75E-10 |
| 339 | 12 | 133702615 | -4.43235 | -3.68902 | 3.34E-03 | 1 | 3.34E-03 | 9.32E-06 | 2.25E-04 |
| 340 | 13 | 27987643 | -3.25342 | -2.87568 | 4.24E-02 | 1 | 4.24E-02 | 1.14E-03 | 4.03E-03 |
| 341 | 13 | 36939146 | 3.234168 | 2.878865 | 4.43E-02 | 1 | 4.43E-02 | 1.22E-03 | 3.99E-03 |
| 342 | 13 | 41584690 | 3.372424 | -2.94456 | 3.03E-02 | 1 | 3.03E-02 | 7.45E-04 | 3.23E-03 |
| 343 | 13 | 42102597 | 3.261013 | 4.267939 | 3.42E-02 | 1 | 3.42E-02 | 1.11E-03 | 1.97E-05 |
| 344 | 13 | 42454184 | 3.264397 | 4.180931 | 3.39E-02 | 1 | 3.39E-02 | 1.10E-03 | 2.90E-05 |
| 345 | 13 | 46517909 | -3.5396 | 3.129968 | 1.83E-02 | 1 | 1.83E-02 | 4.01E-04 | 1.75E-03 |
| 346 | 13 | 50745971 | 3.788424 | 5.251095 | 8.04E-03 | 1 | 8.04E-03 | 1.52E-04 | 1.51E-07 |
| 347 | 13 | 50953245 | 3.47573 | 7.653247 | 1.96E-02 | 1 | 1.96E-02 | 5.09E-04 | 1.96E-14 |
| 347 | 13 | 50968501 | -3.71246 | -3.33738 | 1.02E-02 | 1 | 1.02E-02 | 2.05E-04 | 8.46E-04 |
| 347 | 13 | 51094114 | -5.27096 | -5.77827 | 1.92E-05 | 1 | 1.92E-05 | 1.36E-07 | 7.55E-09 |
| 347 | 13 | 51172615 | -3.43755 | -4.51125 | 2.17E-02 | 1 | 2.17E-02 | 5.87E-04 | 6.44E-06 |
| 348 | 13 | 59090885 | -4.34884 | -3.12107 | 1.88E-02 | 1 | 1.88E-02 | 1.37E-05 | 1.80E-03 |
| 349 | 13 | 76309023 | -3.38524 | 2.94456 | 2.99E-02 | 1 | 2.99E-02 | 7.11E-04 | 3.23E-03 |
| 350 | 13 | 80699166 | 3.623779 | 3.472879 | 1.31E-02 | 1 | 1.31E-02 | 2.90E-04 | 5.15E-04 |
| 350 | 13 | 80720142 | -6.69668 | -3.73069 | 2.89E-03 | 1 | 2.89E-03 | 2.13E-11 | 1.91E-04 |
| 351 | 13 | 81289362 | 3.691342 | -3.1125 | 1.92E-02 | 1 | 1.92E-02 | 2.23E-04 | 1.86E-03 |
| 351 | 13 | 81419502 | -3.51873 | 2.845806 | 3.84E-02 | 1 | 3.84E-02 | 4.34E-04 | 4.43E-03 |
| 352 | 13 | 91945377 | -3.51768 | -3.12107 | 1.89E-02 | 1 | 1.89E-02 | 4.35E-04 | 1.80E-03 |
| 353 | 13 | 94025367 | 3.231309 | -3.50548 | 3.67E-02 | 1 | 3.67E-02 | 1.23E-03 | 4.56E-04 |
| 354 | 13 | 109860174 | -3.79978 | -2.8544 | 3.75E-02 | 1 | 3.75E-02 | 1.45E-04 | 4.31E-03 |
| 355 | 13 | 110431626 | -4.49249 | -8.20366 | 6.42E-04 | 1 | 6.42E-04 | 7.04E-06 | 2.33E-16 |
| 356 | 14 | 38756561 | -4.01817 | -3.15892 | 1.69E-02 | 1 | 1.69E-02 | 5.87E-05 | 1.58E-03 |
| 357 | 14 | 58846731 | 3.754463 | 2.804269 | 4.23E-02 | 1 | 4.23E-02 | 1.74E-04 | 5.04E-03 |
| 358 | 14 | 61353995 | -3.50758 | 3.73069 | 1.80E-02 | 1 | 1.80E-02 | 4.52E-04 | 1.91E-04 |
| 359 | 14 | 68032235 | 3.406094 | 2.845806 | 3.84E-02 | 1 | 3.84E-02 | 6.59E-04 | 4.43E-03 |
| 360 | 14 | 90055468 | 3.548803 | 3.12107 | 1.88E-02 | 1 | 1.88E-02 | 3.87E-04 | 1.80E-03 |
| 361 | 14 | 94838142 | -3.82005 | -4.33695 | 7.30E-03 | 1 | 7.30E-03 | 1.33E-04 | 1.44E-05 |
| 362 | 14 | 103858673 | -5.41823 | -16.1153 | 9.55E-06 | 1 | 9.55E-06 | 6.02E-08 | 1.99E-58 |
| 362 | 14 | 103914842 | -3.95609 | -2.74074 | 4.91E-02 | 1 | 4.91E-02 | 7.62E-05 | 6.13E-03 |
| 362 | 14 | 104310174 | 3.709748 | 3.423196 | 1.02E-02 | 1 | 1.02E-02 | 2.07E-04 | 6.19E-04 |
| 363 | 15 | 41968226 | 5.375416 | -3.41982 | 7.91E-03 | 1 | 7.91E-03 | 7.64E-08 | 6.27E-04 |
| 364 | 15 | 57587111 | 3.999006 | -3.32843 | 1.04E-02 | 1 | 1.04E-02 | 6.36E-05 | 8.73E-04 |
| 365 | 15 | 57734014 | -3.37242 | -3.2959 | 2.58E-02 | 1 | 2.58E-02 | 7.45E-04 | 9.81E-04 |
| 366 | 15 | 62411514 | -3.42499 | 2.740738 | 4.91E-02 | 1 | 4.91E-02 | 6.15E-04 | 6.13E-03 |
| 367 | 15 | 62716409 | -3.76267 | 3.654645 | 8.70E-03 | 1 | 8.70E-03 | 1.68E-04 | 2.58E-04 |
| 368 | 15 | 63313581 | 3.584085 | -3.09625 | 2.01E-02 | 1 | 2.01E-02 | 3.38E-04 | 1.96E-03 |
| 369 | 15 | 63878336 | 5.535084 | 4.623201 | 9.21E-05 | 1 | 9.21E-05 | 3.11E-08 | 3.78E-06 |
| 370 | 15 | 67292740 | 3.73848 | -3.95352 | 9.35E-03 | 1 | 9.35E-03 | 1.85E-04 | 7.70E-05 |
| 370 | 15 | 67337827 | 4.085165 | -3.5005 | 6.18E-03 | 1 | 6.18E-03 | 4.40E-05 | 4.64E-04 |
| 371 | 15 | 67441750 | -3.48763 | 3.515817 | 1.90E-02 | 1 | 1.90E-02 | 4.87E-04 | 4.38E-04 |
| 372 | 15 | 67704983 | -3.47068 | 3.21716 | 2.05E-02 | 1 | 2.05E-02 | 5.19E-04 | 1.29E-03 |
| 373 | 15 | 70399191 | 3.650102 | 3.397562 | 1.21E-02 | 1 | 1.21E-02 | 2.62E-04 | 6.80E-04 |
| 374 | 15 | 70821137 | -3.3592 | -3.20426 | 2.76E-02 | 1 | 2.76E-02 | 7.82E-04 | 1.35E-03 |
| 375 | 15 | 73649395 | -3.48493 | -4.17216 | 1.91E-02 | 1 | 1.91E-02 | 4.92E-04 | 3.02E-05 |
| 376 | 15 | 76606591 | -3.78482 | 3.423196 | 8.13E-03 | 1 | 8.13E-03 | 1.54E-04 | 6.19E-04 |
| 376 | 15 | 77326746 | -3.58409 | 3.000644 | 2.59E-02 | 1 | 2.59E-02 | 3.38E-04 | 2.69E-03 |
| 376 | 15 | 77826969 | 3.45883 | -3.13922 | 2.20E-02 | 1 | 2.20E-02 | 5.43E-04 | 1.69E-03 |
| 377 | 15 | 84343382 | 3.174898 | 3.129968 | 4.52E-02 | 1 | 4.52E-02 | 1.50E-03 | 1.75E-03 |
| 378 | 15 | 90365422 | -4.44497 | -2.99035 | 2.66E-02 | 1 | 2.66E-02 | 8.79E-06 | 2.79E-03 |
| 379 | 15 | 93614127 | 3.399028 | 4.138096 | 2.40E-02 | 1 | 2.40E-02 | 6.76E-04 | 3.50E-05 |
| 380 | 15 | 98536642 | 3.519789 | 2.975694 | 2.77E-02 | 1 | 2.77E-02 | 4.32E-04 | 2.92E-03 |
| 381 | 16 | 112376 | 3.193884 | 3.495632 | 4.03E-02 | 1 | 4.03E-02 | 1.40E-03 | 4.73E-04 |
| 382 | 16 | 319858 | -3.33908 | 3.204257 | 2.91E-02 | 1 | 2.91E-02 | 8.41E-04 | 1.35E-03 |
| 383 | 16 | 977944 | 3.650102 | -3.09625 | 2.01E-02 | 1 | 2.01E-02 | 2.62E-04 | 1.96E-03 |
| 384 | 16 | 1110581 | 3.933567 | 5.942663 | 5.08E-03 | 1 | 5.08E-03 | 8.37E-05 | 2.80E-09 |
| 385 | 16 | 1146126 | 3.412081 | 2.892044 | 3.42E-02 | 1 | 3.42E-02 | 6.45E-04 | 3.83E-03 |
| 386 | 16 | 3821047 | -3.71613 | -3.31774 | 1.08E-02 | 1 | 1.08E-02 | 2.02E-04 | 9.07E-04 |
| 387 | 16 | 4439131 | 4.400187 | 4.402034 | 9.27E-04 | 1 | 9.27E-04 | 1.08E-05 | 1.07E-05 |
| 388 | 16 | 19941557 | -4.11157 | -4.18457 | 2.75E-03 | 1 | 2.75E-03 | 3.93E-05 | 2.86E-05 |
| 389 | 16 | 24698903 | -3.34172 | -3.12997 | 2.99E-02 | 1 | 2.99E-02 | 8.33E-04 | 1.75E-03 |
| 390 | 16 | 29925445 | -4.06226 | 4.315885 | 3.28E-03 | 1 | 3.28E-03 | 4.86E-05 | 1.59E-05 |
| 391 | 16 | 31131174 | -3.58504 | 3.929936 | 1.46E-02 | 1 | 1.46E-02 | 3.37E-04 | 8.50E-05 |
| 392 | 16 | 50626092 | 3.73848 | 3.277765 | 1.21E-02 | 1 | 1.21E-02 | 1.85E-04 | 1.05E-03 |
| 393 | 16 | 53814363 | 17.4855 | 7.965917 | 1.38E-13 | 1 | 1.38E-13 | 1.85E-68 | 1.64E-15 |
| 393 | 16 | 53843848 | 4.798384 | 2.878865 | 3.52E-02 | 1 | 3.52E-02 | 1.60E-06 | 3.99E-03 |
| 393 | 16 | 53865975 | 5.206853 | 3.426629 | 7.76E-03 | 1 | 7.76E-03 | 1.92E-07 | 6.11E-04 |
| 393 | 16 | 53911770 | -3.20137 | -3.04685 | 4.41E-02 | 1 | 4.41E-02 | 1.37E-03 | 2.31E-03 |
| 394 | 16 | 56111959 | 3.27617 | 2.760255 | 4.76E-02 | 1 | 4.76E-02 | 1.05E-03 | 5.78E-03 |
| 395 | 16 | 64610269 | -3.16539 | 3.204257 | 4.47E-02 | 1 | 4.47E-02 | 1.55E-03 | 1.35E-03 |
| 396 | 16 | 69549749 | 4.968261 | -4.60119 | 1.01E-04 | 1 | 1.01E-04 | 6.76E-07 | 4.20E-06 |
| 396 | 16 | 69555247 | 3.728378 | 3.358978 | 9.63E-03 | 1 | 9.63E-03 | 1.93E-04 | 7.82E-04 |
| 397 | 16 | 71850632 | 3.140207 | -3.19205 | 4.78E-02 | 1 | 4.78E-02 | 1.69E-03 | 1.41E-03 |
| 398 | 16 | 72470883 | -3.18006 | -3.12997 | 4.47E-02 | 1 | 4.47E-02 | 1.47E-03 | 1.75E-03 |
| 399 | 16 | 75501567 | 3.574078 | 3.14886 | 1.74E-02 | 1 | 1.74E-02 | 3.51E-04 | 1.64E-03 |
| 400 | 16 | 81463967 | 3.723765 | -3.79001 | 9.75E-03 | 1 | 9.75E-03 | 1.96E-04 | 1.51E-04 |
| 401 | 16 | 81533789 | 7.068771 | -2.80427 | 4.23E-02 | 1 | 4.23E-02 | 1.56E-12 | 5.04E-03 |
| 402 | 16 | 81610480 | -4.72461 | 8.51302 | 2.42E-04 | 1 | 2.42E-04 | 2.31E-06 | 1.69E-17 |
| 403 | 16 | 81635763 | 3.356325 | -4.12456 | 2.68E-02 | 1 | 2.68E-02 | 7.90E-04 | 3.71E-05 |
| 404 | 16 | 88064938 | -4.01268 | 3.261004 | 1.27E-02 | 1 | 1.27E-02 | 6.00E-05 | 1.11E-03 |
| 405 | 16 | 88790147 | -3.48345 | -3.74683 | 1.92E-02 | 1 | 1.92E-02 | 4.95E-04 | 1.79E-04 |
| 406 | 16 | 89084927 | 3.548803 | 3.059944 | 2.22E-02 | 1 | 2.22E-02 | 3.87E-04 | 2.21E-03 |
| 406 | 16 | 89628621 | 4.250841 | 2.975694 | 2.77E-02 | 1 | 2.77E-02 | 2.13E-05 | 2.92E-03 |
| 407 | 16 | 89899940 | 3.36594 | 3.341999 | 2.62E-02 | 1 | 2.62E-02 | 7.63E-04 | 8.32E-04 |
| 408 | 17 | 2243581 | 3.467621 | 5.075039 | 2.01E-02 | 1 | 2.01E-02 | 5.25E-04 | 3.87E-07 |
| 408 | 17 | 2243628 | -3.16574 | 3.746825 | 4.30E-02 | 1 | 4.30E-02 | 1.55E-03 | 1.79E-04 |
| 409 | 17 | 3978531 | 5.798661 | 5.004452 | 1.67E-05 | 1 | 1.67E-05 | 6.68E-09 | 5.60E-07 |
| 409 | 17 | 3980551 | 3.806746 | 3.486231 | 7.61E-03 | 1 | 7.61E-03 | 1.41E-04 | 4.90E-04 |
| 409 | 17 | 4308647 | 3.562189 | 3.034438 | 2.37E-02 | 1 | 2.37E-02 | 3.68E-04 | 2.41E-03 |
| 410 | 17 | 4725920 | 3.212646 | -3.6444 | 3.84E-02 | 1 | 3.84E-02 | 1.32E-03 | 2.68E-04 |
| 411 | 17 | 7452078 | 3.212646 | 3.230844 | 3.94E-02 | 1 | 3.94E-02 | 1.32E-03 | 1.23E-03 |
| 412 | 17 | 9780387 | 4.95578 | 3.715791 | 3.04E-03 | 1 | 3.04E-03 | 7.20E-07 | 2.03E-04 |
| 413 | 17 | 17615528 | -3.59126 | 3.073803 | 2.14E-02 | 1 | 2.14E-02 | 3.29E-04 | 2.11E-03 |
| 414 | 17 | 27595775 | 4.062257 | 3.19205 | 1.54E-02 | 1 | 1.54E-02 | 4.86E-05 | 1.41E-03 |
| 415 | 17 | 27945339 | -3.70997 | 7.27385 | 1.02E-02 | 1 | 1.02E-02 | 2.07E-04 | 3.49E-13 |
| 416 | 17 | 29250853 | 3.569153 | 2.975694 | 2.77E-02 | 1 | 2.77E-02 | 3.58E-04 | 2.92E-03 |
| 417 | 17 | 37628760 | 4.145616 | 3.066772 | 2.18E-02 | 1 | 2.18E-02 | 3.39E-05 | 2.16E-03 |
| 418 | 17 | 40698075 | 6.284981 | 5.363643 | 2.76E-06 | 1 | 2.76E-06 | 3.28E-10 | 8.16E-08 |
| 419 | 17 | 41798545 | 3.116762 | 15.38897 | 4.84E-02 | 1 | 4.84E-02 | 1.83E-03 | 1.94E-53 |
| 420 | 17 | 42175073 | -3.12021 | 10.21161 | 4.81E-02 | 1 | 4.81E-02 | 1.81E-03 | 1.76E-24 |
| 420 | 17 | 42268274 | -4.29908 | -6.06561 | 1.37E-03 | 1 | 1.37E-03 | 1.72E-05 | 1.31E-09 |
| 421 | 17 | 42673671 | 3.445888 | 3.654645 | 2.13E-02 | 1 | 2.13E-02 | 5.69E-04 | 2.58E-04 |
| 421 | 17 | 42933626 | -4.0401 | -3.66545 | 3.61E-03 | 1 | 3.61E-03 | 5.34E-05 | 2.47E-04 |
| 422 | 17 | 46123642 | -4.30878 | -5.74274 | 1.32E-03 | 1 | 1.32E-03 | 1.64E-05 | 9.32E-09 |
| 423 | 17 | 48591209 | -3.11996 | -4.2456 | 4.81E-02 | 1 | 4.81E-02 | 1.81E-03 | 2.18E-05 |
| 423 | 17 | 48634564 | -4.31272 | -4.33252 | 1.30E-03 | 1 | 1.30E-03 | 1.61E-05 | 1.47E-05 |
| 424 | 17 | 60442703 | -3.26518 | 4.439125 | 3.38E-02 | 1 | 3.38E-02 | 1.09E-03 | 9.03E-06 |
| 424 | 17 | 60523911 | 3.805142 | -3.23084 | 1.38E-02 | 1 | 1.38E-02 | 1.42E-04 | 1.23E-03 |
| 424 | 17 | 60720058 | -4.38022 | 4.798198 | 1.00E-03 | 1 | 1.00E-03 | 1.19E-05 | 1.60E-06 |
| 425 | 17 | 61675576 | -4.35112 | -3.93442 | 1.40E-03 | 1 | 1.40E-03 | 1.35E-05 | 8.34E-05 |
| 426 | 17 | 65390645 | -3.49084 | -2.85734 | 3.73E-02 | 1 | 3.73E-02 | 4.82E-04 | 4.27E-03 |
| 427 | 17 | 66078536 | 5.122402 | -3.99899 | 1.10E-03 | 1 | 1.10E-03 | 3.02E-07 | 6.36E-05 |
| 428 | 17 | 66395726 | 3.244601 | -2.84864 | 4.40E-02 | 1 | 4.40E-02 | 1.18E-03 | 4.39E-03 |
| 429 | 17 | 70645446 | -3.53183 | 2.834775 | 3.94E-02 | 1 | 3.94E-02 | 4.13E-04 | 4.59E-03 |
| 430 | 17 | 72762646 | 3.906819 | 3.526701 | 5.69E-03 | 1 | 5.69E-03 | 9.35E-05 | 4.21E-04 |
| 431 | 17 | 76404893 | 3.345988 | 5.215002 | 2.75E-02 | 1 | 2.75E-02 | 8.20E-04 | 1.84E-07 |
| 432 | 17 | 76680261 | 3.988718 | -3.27777 | 1.21E-02 | 1 | 1.21E-02 | 6.64E-05 | 1.05E-03 |
| 433 | 17 | 79473743 | -3.49572 | 3.812309 | 1.86E-02 | 1 | 1.86E-02 | 4.73E-04 | 1.38E-04 |
| 434 | 17 | 79972901 | 3.442789 | 4.962016 | 2.14E-02 | 1 | 2.14E-02 | 5.76E-04 | 6.98E-07 |
| 435 | 18 | 42659922 | -3.38176 | 2.89545 | 3.39E-02 | 1 | 3.39E-02 | 7.20E-04 | 3.79E-03 |
| 436 | 18 | 53154167 | -4.90338 | 4.51125 | 1.47E-04 | 1 | 1.47E-04 | 9.42E-07 | 6.44E-06 |
| 436 | 18 | 53178061 | 4.245855 | -3.48168 | 6.53E-03 | 1 | 6.53E-03 | 2.18E-05 | 4.98E-04 |
| 436 | 18 | 53337912 | 3.274421 | -3.5998 | 3.30E-02 | 1 | 3.30E-02 | 1.06E-03 | 3.18E-04 |
| 436 | 18 | 53443943 | 3.875636 | -4.58392 | 6.14E-03 | 1 | 6.14E-03 | 1.06E-04 | 4.56E-06 |
| 436 | 18 | 53489366 | 3.70463 | -3.19205 | 1.54E-02 | 1 | 1.54E-02 | 2.12E-04 | 1.41E-03 |
| 437 | 18 | 55135865 | 3.114809 | -3.46861 | 4.87E-02 | 1 | 4.87E-02 | 1.84E-03 | 5.23E-04 |
| 438 | 18 | 57876227 | 6.969718 | 5.216455 | 5.86E-06 | 1 | 5.86E-06 | 3.18E-12 | 1.82E-07 |
| 438 | 18 | 57971625 | 5.273957 | 4.620599 | 9.30E-05 | 1 | 9.30E-05 | 1.34E-07 | 3.83E-06 |
| 439 | 18 | 60068549 | -4.06226 | -7.87327 | 3.28E-03 | 1 | 3.28E-03 | 4.86E-05 | 3.45E-15 |
| 441 | 18 | 60846738 | -3.82761 | -4.18828 | 7.13E-03 | 1 | 7.13E-03 | 1.29E-04 | 2.81E-05 |
| 442 | 18 | 73497644 | 3.377762 | 3.112499 | 2.75E-02 | 1 | 2.75E-02 | 7.31E-04 | 1.86E-03 |
| 443 | 19 | 829824 | -3.26411 | -3.57713 | 3.39E-02 | 1 | 3.39E-02 | 1.10E-03 | 3.47E-04 |
| 444 | 19 | 1170445 | -3.66242 | -12.7556 | 1.17E-02 | 1 | 1.17E-02 | 2.50E-04 | 2.90E-37 |
| 445 | 19 | 1225547 | 4.111948 | 3.382974 | 8.87E-03 | 1 | 8.87E-03 | 3.92E-05 | 7.17E-04 |
| 446 | 19 | 1649363 | 3.328687 | -3.01695 | 3.27E-02 | 1 | 3.27E-02 | 8.73E-04 | 2.55E-03 |
| 447 | 19 | 3098612 | 3.174779 | 4.034542 | 4.22E-02 | 1 | 4.22E-02 | 1.50E-03 | 5.47E-05 |
| 448 | 19 | 5174882 | -3.3315 | -4.05981 | 2.86E-02 | 1 | 2.86E-02 | 8.64E-04 | 4.91E-05 |
| 449 | 19 | 8429323 | -4.00054 | -3.24542 | 1.33E-02 | 1 | 1.33E-02 | 6.32E-05 | 1.17E-03 |
| 450 | 19 | 13061983 | 5.344882 | 2.980483 | 2.73E-02 | 1 | 2.73E-02 | 9.05E-08 | 2.88E-03 |
| 450 | 19 | 13160556 | 3.814443 | 3.112499 | 1.92E-02 | 1 | 1.92E-02 | 1.36E-04 | 1.86E-03 |
| 451 | 19 | 18814330 | 3.753198 | -3.261 | 1.27E-02 | 1 | 1.27E-02 | 1.75E-04 | 1.11E-03 |
| 452 | 19 | 19397789 | 5.126009 | 3.158919 | 1.69E-02 | 1 | 1.69E-02 | 2.96E-07 | 1.58E-03 |
| 453 | 19 | 33786208 | 3.7304 | 3.245416 | 1.33E-02 | 1 | 1.33E-02 | 1.91E-04 | 1.17E-03 |
| 454 | 19 | 33953595 | -3.41005 | 2.95306 | 2.92E-02 | 1 | 2.92E-02 | 6.50E-04 | 3.15E-03 |
| 455 | 19 | 41887919 | -3.25907 | -3.00064 | 3.92E-02 | 1 | 3.92E-02 | 1.12E-03 | 2.69E-03 |
| 456 | 19 | 46160458 | 7.944491 | -2.76026 | 4.70E-02 | 1 | 4.70E-02 | 1.95E-15 | 5.78E-03 |
| 456 | 19 | 46210692 | -5.3208 | 3.73069 | 2.89E-03 | 1 | 2.89E-03 | 1.03E-07 | 1.91E-04 |
| 458 | 20 | 6759706 | 3.235387 | 7.939936 | 3.63E-02 | 1 | 3.63E-02 | 1.21E-03 | 2.02E-15 |
| 459 | 20 | 10616382 | -3.57592 | -9.50403 | 1.49E-02 | 1 | 1.49E-02 | 3.49E-04 | 2.02E-21 |
| 460 | 20 | 10991790 | -3.25016 | -7.98601 | 3.51E-02 | 1 | 3.51E-02 | 1.15E-03 | 1.39E-15 |
| 461 | 20 | 32308275 | 5.008318 | 6.440931 | 6.64E-05 | 1 | 6.64E-05 | 5.49E-07 | 1.19E-10 |
| 461 | 20 | 32426842 | -3.72532 | 2.975694 | 2.77E-02 | 1 | 2.77E-02 | 1.95E-04 | 2.92E-03 |
| 461 | 20 | 32469951 | 3.593795 | 2.781336 | 4.48E-02 | 1 | 4.48E-02 | 3.26E-04 | 5.41E-03 |
| 462 | 20 | 32668244 | -5.71074 | -4.34151 | 2.93E-04 | 1 | 2.93E-04 | 1.12E-08 | 1.42E-05 |
| 463 | 20 | 39938122 | -3.81207 | 4.341505 | 7.49E-03 | 1 | 7.49E-03 | 1.38E-04 | 1.42E-05 |
| 464 | 20 | 43057480 | -3.35535 | 2.913211 | 3.25E-02 | 1 | 3.25E-02 | 7.93E-04 | 3.58E-03 |
| 465 | 20 | 44638781 | -3.30255 | 3.770035 | 3.07E-02 | 1 | 3.07E-02 | 9.58E-04 | 1.63E-04 |
| 466 | 20 | 45578923 | -3.38088 | 6.791357 | 2.52E-02 | 1 | 2.52E-02 | 7.23E-04 | 1.11E-11 |
| 466 | 20 | 45582472 | 5.857937 | -8.22075 | 1.00E-06 | 1 | 1.00E-06 | 4.69E-09 | 2.02E-16 |
| 467 | 20 | 48836248 | -5.41731 | 3.245416 | 1.33E-02 | 1 | 1.33E-02 | 6.05E-08 | 1.17E-03 |
| 468 | 20 | 57566637 | 3.447412 | 3.245416 | 2.16E-02 | 1 | 2.16E-02 | 5.66E-04 | 1.17E-03 |
| 469 | 20 | 61279980 | 3.84084 | -3.46861 | 6.84E-03 | 1 | 6.84E-03 | 1.23E-04 | 5.23E-04 |
| 470 | 20 | 62337406 | -3.76248 | 2.860302 | 3.70E-02 | 1 | 3.70E-02 | 1.68E-04 | 4.23E-03 |
| 471 | 21 | 40343918 | 3.445329 | 5.70317 | 2.13E-02 | 1 | 2.13E-02 | 5.70E-04 | 1.18E-08 |
| 472 | 21 | 46931727 | -3.3315 | -2.8821 | 3.50E-02 | 1 | 3.50E-02 | 8.64E-04 | 3.95E-03 |
| 473 | 21 | 47608580 | 3.28806 | 3.654645 | 3.18E-02 | 1 | 3.18E-02 | 1.01E-03 | 2.58E-04 |
| 474 | 22 | 18911333 | 3.286546 | 4.11199 | 3.20E-02 | 1 | 3.20E-02 | 1.01E-03 | 3.92E-05 |
| 475 | 22 | 19675740 | -3.29311 | 11.06564 | 3.14E-02 | 1 | 3.14E-02 | 9.91E-04 | 1.84E-28 |
| 476 | 22 | 28620907 | 3.328119 | 3.344345 | 2.89E-02 | 1 | 2.89E-02 | 8.74E-04 | 8.25E-04 |
| 476 | 22 | 29430459 | 4.798204 | 6.274821 | 1.74E-04 | 1 | 1.74E-04 | 1.60E-06 | 3.50E-10 |
| 477 | 22 | 29518796 | -3.9151 | 10.46299 | 5.41E-03 | 1 | 5.41E-03 | 9.04E-05 | 1.28E-25 |
| 478 | 22 | 29805444 | -4.38632 | 3.158919 | 1.69E-02 | 1 | 1.69E-02 | 1.15E-05 | 1.58E-03 |
| 479 | 22 | 29831277 | 3.501382 | -3.261 | 1.86E-02 | 1 | 1.86E-02 | 4.63E-04 | 1.11E-03 |
| 480 | 22 | 30135928 | -6.45539 | 5.300077 | 3.83E-06 | 1 | 3.83E-06 | 1.08E-10 | 1.16E-07 |
| 480 | 22 | 30597810 | 4.675018 | -3.54419 | 5.36E-03 | 1 | 5.36E-03 | 2.94E-06 | 3.94E-04 |
| 481 | 22 | 30611088 | 3.276855 | 4.539567 | 3.28E-02 | 1 | 3.28E-02 | 1.05E-03 | 5.64E-06 |
| 481 | 22 | 30736921 | -3.13287 | 3.893837 | 4.66E-02 | 1 | 4.66E-02 | 1.73E-03 | 9.87E-05 |
| 482 | 22 | 41637119 | 3.13862 | 5.735229 | 4.60E-02 | 1 | 4.60E-02 | 1.70E-03 | 9.74E-09 |
| 483 | 22 | 46621994 | 4.787874 | 4.645743 | 1.83E-04 | 1 | 1.83E-04 | 1.69E-06 | 3.39E-06 |
| 484 | 22 | 50178288 | 3.416268 | 2.913211 | 3.25E-02 | 1 | 3.25E-02 | 6.35E-04 | 3.58E-03 |
| 485 | 22 | 50303530 | -3.13391 | -4.51125 | 4.65E-02 | 1 | 4.65E-02 | 1.72E-03 | 6.44E-06 |
| 486 | 22 | 50584820 | -3.2265 | -2.92839 | 4.39E-02 | 1 | 4.39E-02 | 1.25E-03 | 3.41E-03 |
| 487 | 22 | 50634552 | 3.731282 | 3.532366 | 9.55E-03 | 1 | 9.55E-03 | 1.91E-04 | 4.12E-04 |
| 487 | 22 | 50700723 | -4.27235 | -4.32821 | 1.52E-03 | 1 | 1.52E-03 | 1.93E-05 | 1.50E-05 |

**Supplementary Table 10.** The characteristics of participants and comparison between individuals with and without type 2 diabetes.

| **Baseline characteristic** |  | **Whether participants with type 2 diabetes or not** | | |
| --- | --- | --- | --- | --- |
|  | **Overall (N=352,879)** | **No (N=339,062)** | **Yes (N=13,817)** | **P** |
| Male (%) | 159347 (45.2) | 150646 (44.4) | 8701 (63.0) | < 2.2e-16 |
| Reference age (mean (SD)) | 60.65 (7.80) | 60.55 (7.80) | 63.20 (7.24) | < 2.2e-16 |
| Smoking status (%) |  |  |  | < 2.2e-16 |
| Prefer not to answer | 1211 (0.3) | 1134 (0.3) | 77 (0.6) |  |
| Never | 191127 (54.2) | 185518 (54.7) | 5609 (40.6) |  |
| Previous | 126496 (35.8) | 120078 (35.4) | 6418 (46.5) |  |
| Current | 34045 (9.6) | 32332(9.5) | 1713 (12.4) |  |
| Alcohol intake frequency (%) |  |  |  | < 2.2e-16 |
| Prefer not to answer | 223 (0.1) | 208 (0.1) | 15 (0.1) |  |
| Daily or almost daily | 77332 (21.9) | 75027 (22.1) | 2305(16.7) |  |
| Three or four times a week | 85635 (24.3) | 83220 (24.5) | 2415 (17.5) |  |
| Once or twice a week | 91960 (26.1) | 88540 (26.1) | 3420 (24.8) |  |
| One to three times a month | 38059 (10.8) | 36350 (10.7) | 1709 (12.4) |  |
| Special occasions only | 36925 (10.5) | 34585 (10.2) | 2340 (16.9) |  |
| Never | 22745 (6.4) | 21132 (6.2) | 1613 (11.7) |  |
| physical activity (%) | 244885 (69.4) | 237041 (69.9) | 7844 (56.8) | < 2.2e-16 |
| Medication treatments (%) | 6644 (1.9) | 1363 (0.4) | 5281 (38.2) | < 2.2e-16 |
| BMI (mean (SD)) | 27.28 (4.63) | 27.08 (4.47) | 32.07 (5.80) | < 2.2e-16 |
| BMD (mean (SD)) | 0.54 (0.14) | 0.54 (0.13) | 0.57 (0.16) | < 2.2e-16 |
| Falls in the last year (%) | 66969 (19.0) | 63404 (18.7) | 3565 (25.9) | < 2.2e-16 |
| Fracture (%) | 16147 (4.6) | 15406 (4.5) | 741 (5.4) | 1.30E-06 |
| HbA1c (mean (SD)) | 35.62 (5.39) | 35.12 (4.20) | 47.98 (12.25) | < 2.2e-16 |
| Follow-up time (mean (SD)),years | 9.03 (3.06) | 9.05 (3.03) | 8.34 (3.56) | < 2.2e-16 |

| **Supplementary Table 11.** The regression between type 2 diabetes, fracture and BMD. | | | | | |
| --- | --- | --- | --- | --- | --- |
| Traits | HR | BETA | 95% CI Lower | 95% CI Upper | P value |
|  | Pooled (n=352,879) |  |  |  |  |
| Fracture ^a^ | 1.527 | - | 1.385 | 1.685 | < 2e-16 |
| Fracture ^b^ | 1.574 | - | 1.425 | 1.739 | < 2e-16 |
| BMD ^a^ | - | 0.00957 | 0.00665 | 0.01249 | 1.35E-10 |
|  | Male (n=159,347) | |  |  |  |
| Fracture ^a^ | 1.587 | - | 1.379 | 1.828 | 1.26E-10 |
| Fracture ^b^ | 1.607 | - | 1.393 | 1.853 | 7.21E-11 |
| BMD ^a^ | - | 0.00293 | -0.00129 | 0.00715 | 0.173 |
|  | Female (n=193,532) | |  |  |  |
| Fracture ^a^ | 1.530 | - | 1.334 | 1.756 | 1.27E-09 |
| Fracture ^b^ | 1.601 | - | 1.393 | 1.841 | 3.59E-11 |
| BMD ^a^ | - | 0.0126 | 0.00846 | 0.01668 | 2.02E-09 |
| ^a^ Model 0 adjusted for reference age, sex, BMI, physical activity, fall history, HbA1c and medication treatments  ^b^ Model 1 adjusted for a + BMD | | | | | |

**Supplementary Table** **12.** Assessment of the mediators (BMI) for the association between type 2 diabetes and fracture.

| Mediator | Effect | Estimate | 95% CI Lower | 95% CI Upper | P value |
| --- | --- | --- | --- | --- | --- |
| BMI | Total Effect | 0.011 | 0.0066 | 0.0165 | <2e-16 |
|  | ACME (average) | -0.003 | -0.0034 | -0.0025 | <2e-16 |
|  | ADE (average) | 0.014 | 0.0094 | 0.0198 | <2e-16 |
|  | Prop. Mediated (average) | -0.302 | -0.4522 | -0.1903 | <2e-16 |
| Total effect: direct (ADE)+indirect (ACME). | | | | | |
| Prop. mediated: conceptually ACME/total effect. | | | | | |
| ACME, average causal mediation effect; ADE, average direct effect.  adjusted for the reference age, sex, physical activity, fall history, HbA1c and medication treatments | | | | | |

| **Supplementary Table 13.** Baseline characteristics of participants with different Numbers of Risk Factors. | | | |
| --- | --- | --- | --- |
| Baseline characteristics | Numbers of Risk Factors | | |
|  | 0 | 1 | ≥2 |
| Number of T2D Patients at risk | 2,303 | 4,128 | 4,252 |
| Risk factors(N,%) |  |  |  |
| BMI≤25kg/m^2^ | - | 293(7.1) | 669(15.7) |
| no physical activity | - | 1,582(38.3) | 2,189(51.5) |
| falls in the last year | - | 743(18.0) | 1,911(44.9) |
| HbA1c≥47.5mmol/mol | - | 834(20.2) | 2,831(66.6) |
| antidiabetic medication treatment | - | 676(16.4) | 2,376(55.9) |
| Fracture(N,%) | 97(4.2) | 202(4.9) | 260(6.1) |
| Bone mineral density(mean,SD) | 0.572(0.157) | 0.567(0.163) | 0.563(0.166) |

| **Supplementary Table 14.** Detailed information on the field ID and codes for participants included in UK Biobank. | | |
| --- | --- | --- |
| Description | Field IDs | Codes |
| Type 2 diabetes * | 20002 | 1223 |
| Type 2 diabetes † | 41270 | E110, E11, E111, E112, E113, E114, E115, E116, E117, E118, E119, O241 |
| Type 2 diabetes ‡ | 41203 | 25000, 25010 |
| Fracture* | 20002 | 1626, 1627, 1628, 1629,1630, 1631, 1632, 1633, 1634, 1635, 1636, 1637,  1638, 1639, 1640, 1644, 1645, 1646, 1647, 1648, 1649,1650, 1651, 1652,  1653, 1654, 1655, 1656 |
| Fracture† | 41270 | M484, M840, M841, M842, M843, S02, S12, S22, S32, S42, S52, S62, S72, S82, S92, T02, T08, T10, T10X0, T12, T142, T1420; T902, T911, T912, T921, T922, T931, T932, X5909, Z544 |
| Fracture‡ | 41203 | 7338, 800, 801, 802, 803, 805, 806, 807, 808, 809, 810, 811, 812, 813, 814,  815, 816, 817, 820, 821, 822, 823, 824, 825, 826, 828, 829, 905, 9050, 9052, 9053, 9054 |
| HbA1c | 30750 | \ |
| Age when attended assessment center | 21003 | \ |
| Sex | 31 | \ |
| Body mass index | 21001 | \ |
| Ethnic background | 21000 | 1001, 1002, 1003, 1 |
| Kinship | 22021 | 0 |
| Heel bone mineral density | 4105, 3148, 3084 | \ |
| Falls in the last year | 2296 | \ |
| Physical activity | 884, 894, 904, 914 | \ |
| Medication treatments | 20003 | 1140883066, 1140884600, 1141153254, 1141171646, 1141177600,  1140874674, 1140874718, 1140874744, 1140874646, 1140874658,  1141152590, 1140874706, 1140874674, 1141168660, 1141173882 |
| Date of attending assessment center | 53 | \ |
| Date lost to follow-up | 191 | \ |
| Date of death | 40000 | \ |
| Year when the non-cancer illness first diagnosed | 20008 | \ |
| Date of first in-patient diagnosis based on ICD-9 | 41263 | \ |
| Date of first in-patient diagnosis based on ICD-10 | 41280 | \ |
| *self-report; †ICD-10, the International Classification of Diseases, the 10th Revisions, ‡ICD-9, the International Classification of Diseases, 9^th^ Revision. | | |

| **Supplementary Table 15.** The International Classification of Diseases (ICD) codes and self-reported codes for excluded diseases. (Diseases \| Field IDs \| Codes; Type 1 diabetes \| 20002 \| 1222; \| 41270 \| E10, E100, E101, E102, E103, E104, E105, E106, E107, E108, E109, O240; \| 41203 \| 25001, 25011; Fracture of bone in neoplastic disease \| 41270 \| M907, M9070, M9071, M9072, M9073, M9074, M9075, M9076, M9077, M9078, M9079; Follow-up care involving removal of fracture plate and other internal fixation devices \| 41270 \| Z470; Pathological fracture \| 41203 \| 7331, 73313, 73315, 73316; Rheumatoid Arthritis \| 20002 \| 1464; \| 41270 \| M0530, M0531, M0532, M0533, M0534, M0535, M0536, M0537, M0538, M0539, M0580, M0581, M0582, M0583, M0584, M0585, M0586, M058, M0588, M0589, M0590, M0591, M0592, M0593, M0594, M0595, M0596, M0597, M0598, M0599, M0600, M0601, M0602, M0603, M0604, M0605, M0606, M0607, M0608, M0609, M0680, M0681, M0682, M0683, M0684, M0685, M0686, M0687, M0688, M0689, M0690, M0691, M0692, M0693, M0694, M0695, M0696, M0697, M0698, M0699, M0800, M0801, M0802, M0804, M080, M0806, M0807, M0808, M0809; \| 41203 \| 71400, 71401, 71402, 71403, 71404, 71405, 71406, 71407, 71408, 71409, 71423, 71424; Ulcerative Colitis \| 20002 \| 1463; \| 41270 \| K518, K519, M0750, M0751, M0752, M0753, M0754, M0755, M0756, M0757, M0758, M0759, M0920, M0921, M0922, M0923, M0924, M0925, M0926, M0927, M0928, M0929; Multiple sclerosis \| 20002 \| 1261; \| 41270 \| G35; \| 41203 \| 3409; Crohn's disease \| 20002 \| 1462; \| 41270 \| K50, K500, K501, K508, K509, M074, M0740, M0741, M0742, M0743, M0744, M0745, M0746, M0747, M0748, M0749, M091, M0910, M0911, M0912, M0913, M0914, M0915, M0916, M0917, M0918, M0919; Hyperthyroidism \| 20002 \| 1225; \| 41270 \| E050, E051, E052, E053,,E058, E059, E062; \| 41203 \| 2424, 2428, 2429, 7753; Lupus erythematosus \| 41270 \| L930, L931, L932, M320, M321, M328, M329, M3290; \| 41203 \| 6954, 7100) **Supplementary Table 16.** Detailed information on the field ID and codes for specific fracture sites included in UK Biobank. \| \|; Description \| Field IDs \| Codes; weight-bearing bones fracture \| 20002 \| 1630, 1646, 1647, 1648, 1649, 1652; \| 41270 \| M484, M4840, M4842, M4844, M4845, M4846, M4847, M4848, M4849, M8405, M8415, M8425, M8435, S12, S120, S1200, S121, S1210, S122, S1220, S127, S1270, S128, S1280, S1281, S129, S1290, S1291, S221, S2210, S32, S320, S3200, S321, S3210, S3211, S322, S3220, S323, S3230, S3231, S324, S3240, S3241, S325, S3250, S3251, S327, S3270, S3271, S328, S3280, S3281, S72, S720, S7200, S7201, S721, S7210, S7211, S722, S7220, S7221, S723, S72230, S7231, S724, S7240, S7241, S727, S7270, S728, S7280, S729, S7290, S7291, S821, S8210, S8211, S822, S8220, S8221, S823, S8231, S827, S8270, S8271, S828, S8280, S8281, S8286, S829, S8290, S8291, T02, T0200, T021, T0210, T027, T0270, T08, T08X0, T911, T931; \| 41203 \| 73385, 73386, 805, 8050, 8052, 8054, 8056, 8058, 808, 8080, 8082, 8084, 8088, 8089, 820, 8200, 8202, 8208, 8210, 8211, 8212, 9053; other bones fracture \| 20002 \| 1626, 1627, 1628, 1629, 1631, 1632, 1633, 1634, 1635, 1636, 1637, 1638, 1639, 1640, 1644, 1645, 1650, 1651, 1653, 1654, 1655, 1656; \| 41270 \| M84, M840, M8400, M8401, M8402, M8403, M8404, M8406, M8407, M8408, M841, M8410, M8411, M8412, M8413, M8414, M8416, M8417, M8418, M8419, M842, M8421, M8422, M8423, M8424, M8426, M8427, M8428, M843, M8430, M8436, M8437, M8438, M8439, S02, S020, S0200, S0201, S021, S0210, S0211, S022, S0220, S0221, S023, S0230, S0231, S024, S0240, S0241, S025, S0250, S0251, S026, S0260, S0261, S027, S0270, S0271, S028, S0280, S0281, S029, S0290, S0291, S22, S220, S2200, S222, S2220, S223, S2230, S2231, S224, S2241, S225, S2250, S2251, S228, S2280, S229, S2290, S42, S420, S4200, S4201, S421, S4210, S422, S4220, S4221, S423, S4230, S4231, S424, S4240, S4241, S427, S4270, S4271, S428, S4280, S429, S4290, S52, S520, S5200, S5201, S521, S5210, S5211, S522, S5220, S5221, S523, S5230, S5231, S524, S5240, S5241, S525, S5250, S5251, S526, S5260, S5261, S527, S5270, S5271, S528, S5280, S5281, S529, S5290, S62, S620, S6200, S621, S6210, S6211, S622, S6220, S6221, S623, S6230, S6231, S624, S6240, S6241, S625, S6250, S6251, S626, S6261, S627, S6270, S6271, S628, S6280, S6281, S82, S820, S8200, S8201, S824, S8240, S8241, S825, S8250, S8251, S826, S8260, S8261, S92, S920, S9200, S9201, S921, S9210, S9211, S922, S9220, S9221, S923, S9230, S9231, S924, S9240, S9241, S925, S9250, S9251, S927, S9270, S9271, S929, S9290, S9291, T020, T022, T0220, T023, T0230, T0231, T024, T0240, T0241, T025, T0250, T0251, T026, T0260, T028, T0280, T029, T0290, T10, T10X0, T12, T142, T1420, T902, T912, T921, T922, T932, X5909, Z544; \| 41203 \| 7338, 73381, 73382, 73383, 73384, 73387, 73388, 73389, 800, 8000, 8001, 8002, 8003, 801, 8010, 8011, 802, 8020, 8022, 8023, 8024, 8026, 8028, 803, 8030, 8031, 806, 8064, 807, 8070, 8072, 8074, 808, 8080, 8082, 8084, 8088, 8089, 809, 8090, 8091, 810, 8100, 811, 8110, 812, 8120, 8121, 8122, 8123, 8124, 8125, 813, 8130, 8131, 8132, 8134, 8135, 814,8140, 8141, 815, 8150, 8151, 816, 8160, 8161, 817, 8170, 820, 822, 8220, 8221, 823, 8230, 8231, 8232, 8233, 8240, 8241, 8242, 8244, 8245, 8246, 8247, 8248, 8249, 825, 8250, 8252, 8253, 826, 8260, 8261, 828, 8280, 829, 8290, 905, 9050, 9052, 9054; vertebral fracture \| 20002 \| 1646; \| 41270 \| M484, M4840, M4842, M4844, M4845, M4846, M4847, M4848, M4849, S12, S120, S1200, S121, S1210, S122, S1220, S127, S1270, S128, S1280, S1281, S129, S1290, S1291, S22, S220, S2200, S221, S2210, S222, S2220, S320, S3200; \| 41203 \| 805, 8050, 8052, 8054, 8056, 806, 8064; hip fracture \| 20002 \| 1647, 1648, 1649; \| 41270 \| S324, S3240, S3241, S72, S720, S7200, S7201, S723, S7230, S7231, S724, S7240, S7241, S727, S7270, S728, S7280, S729, S7290, S7291, T931; \| 41203 \| 808, 8080, 8082, 8084, 8088, 8089, 820, 8200, 8202, 8208, 8210, 8211, 8212, 9053 | | | | | | | | | |
| --- | --- | --- | --- | --- | --- | --- | --- | --- | --- |
| **Supplementary Table 17.** Detailed information for instrumental variables of T2D and BMI on fracture and BMD. | | | | | | | | | |
| *Instrumental variables of T2D and BMI on fracture (N=390)* | | | | |  |  |  |  |  |
| SNP | beta_T2D | beta_BMI | se_T2D | se_BMI | beta_fracture | se_fracture | P_T2D | P_BMI | P_fracture |
| rs16951275 | 0.0340 | 0.0311 | 0.0077 | 0.0037 | 0.0023 | 0.0097 | 1.14E-05 | 1.91E-17 | 0.8169 |
| rs1962412 | -0.0521 | -0.0041 | 0.0070 | 0.0047 | 0.0036 | 0.0089 | 1.11E-13 | 0.383 | 0.6828 |
| rs1999536 | -0.0404 | 0.0056 | 0.0065 | 0.0038 | -0.0010 | 0.0083 | 4.86E-10 | 0.1406 | 0.9044 |
| rs11063069 | -0.0560 | 0.0100 | 0.0079 | 0.0041 | 0.0035 | 0.0107 | 9.94E-13 | 0.0155 | 0.7407 |
| rs2107133 | 0.0643 | -0.0016 | 0.0097 | 0.0059 | -0.0191 | 0.0121 | 4.02E-11 | 0.7862 | 0.1137 |
| rs16889471 | 0.0448 | -0.0028 | 0.0077 | 0.0038 | -0.0036 | 0.0103 | 7.38E-09 | 0.4615 | 0.727 |
| rs13188193 | 0.1106 | -0.0104 | 0.0147 | 0.0080 | -0.0221 | 0.0193 | 4.52E-14 | 0.1949 | 0.2514 |
| rs7678054 | -0.0394 | -0.0115 | 0.0064 | 0.0037 | 0.0008 | 0.0082 | 6.88E-10 | 0.001883 | 0.9213 |
| rs8032939 | -0.0418 | -0.0064 | 0.0074 | 0.0044 | 0.0005 | 0.0095 | 1.88E-08 | 0.1458 | 0.956 |
| rs4977218 | -0.0505 | 0.0073 | 0.0069 | 0.0040 | 0.0073 | 0.0085 | 2.71E-13 | 0.068 | 0.3881 |
| rs7178762 | -0.0387 | -0.0058 | 0.0064 | 0.0037 | 0.0042 | 0.0081 | 1.37E-09 | 0.117 | 0.6079 |
| rs12940622 | -0.0163 | -0.0182 | 0.0065 | 0.0031 | 0.0058 | 0.0082 | 0.01204 | 2.49E-09 | 0.4786 |
| rs693232 | -0.0542 | -0.0480 | 0.0080 | 0.0042 | 0.0124 | 0.0102 | 9.67E-12 | 7.78E-31 | 0.2226 |
| rs7124681 | 0.0362 | 0.0259 | 0.0065 | 0.0031 | -0.0088 | 0.0082 | 2.45E-08 | 1.16E-16 | 0.2858 |
| rs2596429 | -0.0640 | 0.0062 | 0.0088 | 0.0052 | 0.0241 | 0.0113 | 3.42E-13 | 0.2331 | 0.0329 |
| rs7642311 | 0.0545 | 0.0030 | 0.0096 | 0.0057 | 0.0149 | 0.0123 | 1.53E-08 | 0.5987 | 0.2242 |
| rs3812547 | 0.0495 | 0.0020 | 0.0070 | 0.0052 | -0.0044 | 0.0090 | 1.71E-12 | 0.7005 | 0.622 |
| rs1431841 | 0.0429 | 0.0132 | 0.0077 | 0.0046 | -0.0001 | 0.0101 | 3.08E-08 | 0.00411 | 0.9936 |
| rs1000940 | -0.0218 | -0.0192 | 0.0069 | 0.0034 | 0.0072 | 0.0089 | 0.001607 | 1.28E-08 | 0.4207 |
| rs10938397 | -0.0414 | -0.0402 | 0.0065 | 0.0031 | 0.0139 | 0.0084 | 1.80E-10 | 3.21E-38 | 0.09776 |
| rs9957320 | -0.0487 | -0.0226 | 0.0085 | 0.0051 | 0.0134 | 0.0109 | 9.34E-09 | 9.36E-06 | 0.2172 |
| rs12266632 | -0.2255 | 0.0185 | 0.0128 | 0.0068 | -0.0335 | 0.0164 | 9.57E-70 | 0.006341 | 0.0411 |
| rs6446298 | -0.0373 | -0.0151 | 0.0067 | 0.0040 | 0.0139 | 0.0086 | 2.60E-08 | 0.00016 | 0.1063 |
| rs12910361 | -0.0814 | -0.0083 | 0.0070 | 0.0041 | 0.0217 | 0.0089 | 3.95E-31 | 0.04293 | 0.01516 |
| rs1879523 | -0.0190 | -0.0200 | 0.0068 | 0.0033 | 0.0024 | 0.0087 | 0.005242 | 1.34E-09 | 0.7855 |
| rs3131012 | -0.0471 | 0.0023 | 0.0064 | 0.0037 | 0.0071 | 0.0082 | 1.65E-13 | 0.5342 | 0.3853 |
| rs10733682 | 0.0219 | 0.0174 | 0.0065 | 0.0031 | -0.0090 | 0.0083 | 0.000742 | 1.83E-08 | 0.281 |
| rs3862948 | 0.0414 | 0.0133 | 0.0075 | 0.0044 | -0.0223 | 0.0096 | 3.98E-08 | 0.002505 | 0.02068 |
| rs7243357 | 0.0432 | 0.0217 | 0.0084 | 0.0040 | -0.0126 | 0.0107 | 2.50E-07 | 3.86E-08 | 0.2419 |
| rs12484907 | 0.0866 | -0.0021 | 0.0152 | 0.0074 | -0.0025 | 0.0201 | 1.17E-08 | 0.776 | 0.9027 |
| rs11074422 | -0.0041 | -0.0188 | 0.0067 | 0.0034 | 0.0055 | 0.0089 | 0.5406 | 3.34E-08 | 0.5387 |
| rs738409 | -0.0436 | 0.0060 | 0.0076 | 0.0046 | -0.0213 | 0.0098 | 1.17E-08 | 0.1921 | 0.03037 |
| rs6477694 | -0.0196 | -0.0174 | 0.0066 | 0.0031 | 0.0155 | 0.0085 | 0.002965 | 2.67E-08 | 0.06846 |
| rs2252867 | 0.0539 | 0.0023 | 0.0066 | 0.0033 | -0.0003 | 0.0085 | 3.05E-16 | 0.4852 | 0.9702 |
| rs2066827 | -0.0473 | 0.0020 | 0.0081 | 0.0063 | 0.0062 | 0.0107 | 4.44E-09 | 0.7509 | 0.5602 |
| rs2112347 | 0.0467 | 0.0261 | 0.0067 | 0.0031 | 0.0022 | 0.0085 | 3.19E-12 | 6.19E-17 | 0.7958 |
| rs10516495 | -0.0418 | -0.0133 | 0.0069 | 0.0040 | -0.0146 | 0.0088 | 1.46E-09 | 0.000884 | 0.09738 |
| rs10974438 | -0.0514 | -0.0005 | 0.0067 | 0.0033 | 0.0037 | 0.0086 | 1.71E-14 | 0.8826 | 0.6673 |
| rs508419 | -0.0811 | 0.0028 | 0.0075 | 0.0044 | 0.0125 | 0.0098 | 5.43E-27 | 0.5245 | 0.2035 |
| rs1528435 | 0.0275 | 0.0178 | 0.0066 | 0.0031 | -0.0153 | 0.0084 | 3.06E-05 | 1.20E-08 | 0.06818 |
| rs12962523 | -0.0147 | -0.0342 | 0.0080 | 0.0051 | 0.0043 | 0.0104 | 0.0647 | 2.00E-11 | 0.6754 |
| rs9971402 | 0.0528 | 0.0027 | 0.0091 | 0.0054 | -0.0004 | 0.0118 | 6.78E-09 | 0.6171 | 0.9704 |
| rs9687846 | 0.0701 | -0.0050 | 0.0081 | 0.0040 | -0.0129 | 0.0103 | 3.47E-18 | 0.2125 | 0.2085 |
| rs1558902 | 0.1218 | 0.0818 | 0.0065 | 0.0031 | -0.0257 | 0.0083 | 1.52E-78 | 7.51E-153 | 0.002059 |
| rs5015480 | -0.1091 | 0.0035 | 0.0065 | 0.0037 | -0.0017 | 0.0082 | 2.19E-63 | 0.3442 | 0.8397 |
| rs1412234 | -0.0393 | -0.0235 | 0.0068 | 0.0041 | 0.0049 | 0.0088 | 7.71E-09 | 9.94E-09 | 0.5787 |
| rs11625769 | 0.0303 | 0.0190 | 0.0068 | 0.0034 | 0.0040 | 0.0087 | 8.50E-06 | 1.70E-08 | 0.6477 |
| rs3802120 | -0.0582 | -0.0124 | 0.0067 | 0.0040 | -0.0250 | 0.0086 | 3.78E-18 | 0.001935 | 0.003571 |
| rs1796330 | -0.0487 | 0.0008 | 0.0065 | 0.0037 | -0.0007 | 0.0082 | 6.28E-14 | 0.8288 | 0.9342 |
| rs4671328 | 0.0328 | 0.0215 | 0.0067 | 0.0037 | 0.0049 | 0.0083 | 9.84E-07 | 6.22E-09 | 0.55 |
| rs10838524 | 0.0350 | 0.0012 | 0.0064 | 0.0034 | -0.0100 | 0.0082 | 4.25E-08 | 0.7303 | 0.2227 |
| rs205262 | 0.0037 | -0.0221 | 0.0072 | 0.0035 | -0.0120 | 0.0091 | 0.6085 | 1.75E-10 | 0.1893 |
| rs9271608 | -0.0847 | 0.0013 | 0.0087 | 0.0048 | -0.0080 | 0.0103 | 1.91E-22 | 0.7865 | 0.4365 |
| rs29941 | -0.0229 | -0.0182 | 0.0068 | 0.0033 | 0.0272 | 0.0087 | 0.000766 | 2.41E-08 | 0.001704 |
| rs10509406 | 0.0564 | 0.0115 | 0.0086 | 0.0052 | -0.0182 | 0.0108 | 5.06E-11 | 0.027 | 0.09193 |
| rs1063192 | 0.0575 | 0.0002 | 0.0065 | 0.0031 | 0.0000 | 0.0082 | 8.17E-19 | 0.9574 | 0.9979 |
| rs9945063 | 0.0221 | 0.0217 | 0.0074 | 0.0038 | 0.0089 | 0.0100 | 0.00295 | 1.35E-08 | 0.3717 |
| rs7969720 | -0.0383 | 0.0019 | 0.0069 | 0.0042 | 0.0013 | 0.0088 | 2.98E-08 | 0.651 | 0.8795 |
| rs13130484 | 0.0435 | 0.0401 | 0.0067 | 0.0031 | -0.0156 | 0.0084 | 8.49E-11 | 4.24E-38 | 0.06247 |
| rs11074446 | 0.0363 | 0.0256 | 0.0094 | 0.0045 | -0.0121 | 0.0120 | 0.000117 | 1.31E-08 | 0.3141 |
| rs2616055 | 0.0799 | -0.0003 | 0.0137 | 0.0084 | -0.0081 | 0.0180 | 5.70E-09 | 0.9715 | 0.6513 |
| rs2862874 | 0.0360 | 0.0208 | 0.0066 | 0.0038 | 0.0015 | 0.0084 | 4.83E-08 | 4.41E-08 | 0.8536 |
| rs719727 | 0.0592 | -0.0060 | 0.0074 | 0.0036 | 0.0185 | 0.0095 | 1.67E-15 | 0.0979 | 0.05213 |
| rs17791513 | 0.1016 | -0.0024 | 0.0132 | 0.0060 | -0.0356 | 0.0167 | 1.35E-14 | 0.695 | 0.03327 |
| rs7957197 | -0.0672 | -0.0011 | 0.0080 | 0.0039 | 0.0019 | 0.0102 | 3.04E-17 | 0.7684 | 0.8494 |
| rs2796441 | -0.0674 | 0.0067 | 0.0065 | 0.0033 | 0.0018 | 0.0084 | 2.97E-25 | 0.04045 | 0.8293 |
| rs7467618 | -0.0393 | 0.0038 | 0.0065 | 0.0038 | 0.0020 | 0.0083 | 1.41E-09 | 0.3173 | 0.8054 |
| rs10811658 | -0.0522 | 0.0088 | 0.0071 | 0.0040 | 0.0030 | 0.0100 | 2.27E-13 | 0.02994 | 0.7602 |
| rs7227255 | -0.1365 | -0.0856 | 0.0224 | 0.0116 | 0.0284 | 0.0287 | 1.12E-09 | 1.70E-13 | 0.3213 |
| rs17724992 | 0.0148 | 0.0194 | 0.0075 | 0.0035 | 0.0043 | 0.0096 | 0.04962 | 3.42E-08 | 0.6489 |
| rs2272662 | -0.0402 | 0.0075 | 0.0072 | 0.0061 | -0.0198 | 0.0094 | 2.63E-08 | 0.2189 | 0.03412 |
| rs9368222 | 0.1379 | -0.0095 | 0.0071 | 0.0034 | 0.0110 | 0.0092 | 1.42E-83 | 0.00546 | 0.2312 |
| rs2071479 | 0.1160 | -0.0095 | 0.0180 | 0.0101 | 0.0108 | 0.0236 | 1.19E-10 | 0.3469 | 0.6465 |
| rs805262 | 0.0379 | 0.0025 | 0.0064 | 0.0037 | -0.0177 | 0.0082 | 2.96E-09 | 0.4992 | 0.03024 |
| rs17603828 | 0.0800 | 0.0040 | 0.0133 | 0.0082 | -0.0212 | 0.0168 | 1.78E-09 | 0.6257 | 0.2077 |
| rs1552224 | 0.0981 | -0.0184 | 0.0087 | 0.0042 | -0.0022 | 0.0111 | 1.50E-29 | 9.40E-06 | 0.8456 |
| rs4686698 | 0.0953 | -0.0098 | 0.0172 | 0.0091 | 0.0229 | 0.0226 | 2.86E-08 | 0.2793 | 0.3105 |
| rs3101336 | -0.0248 | -0.0334 | 0.0066 | 0.0031 | -0.0103 | 0.0083 | 0.00017 | 2.66E-26 | 0.2138 |
| rs1477199 | -0.0226 | -0.0242 | 0.0091 | 0.0044 | -0.0016 | 0.0117 | 0.0131 | 4.56E-08 | 0.8907 |
| rs9956279 | 0.0368 | 0.0348 | 0.0069 | 0.0033 | -0.0078 | 0.0089 | 1.01E-07 | 2.62E-25 | 0.3818 |
| rs1800961 | 0.1602 | -0.0031 | 0.0175 | 0.0092 | -0.0203 | 0.0234 | 5.10E-20 | 0.7349 | 0.3866 |
| rs13191362 | 0.0235 | 0.0277 | 0.0098 | 0.0048 | -0.0019 | 0.0124 | 0.01695 | 7.34E-09 | 0.8767 |
| rs2242517 | 0.0430 | 0.0034 | 0.0066 | 0.0041 | 0.0002 | 0.0086 | 7.09E-11 | 0.407 | 0.9843 |
| rs4580892 | 0.0404 | -0.0029 | 0.0070 | 0.0034 | -0.0575 | 0.0092 | 8.46E-09 | 0.3972 | 3.68E-10 |
| rs7572970 | -0.0447 | -0.0066 | 0.0072 | 0.0043 | -0.0061 | 0.0091 | 6.12E-10 | 0.1248 | 0.5038 |
| rs1808579 | -0.0251 | -0.0167 | 0.0064 | 0.0031 | -0.0095 | 0.0082 | 8.50E-05 | 4.17E-08 | 0.2462 |
| rs2023681 | -0.0826 | 0.0017 | 0.0115 | 0.0068 | -0.0036 | 0.0144 | 7.40E-13 | 0.8026 | 0.8005 |
| rs13257021 | 0.0485 | 0.0045 | 0.0064 | 0.0037 | 0.0048 | 0.0081 | 3.11E-14 | 0.2239 | 0.5573 |
| rs17218700 | -0.0615 | -0.0218 | 0.0104 | 0.0048 | 0.0070 | 0.0126 | 2.97E-09 | 6.33E-06 | 0.5789 |
| rs2239802 | -0.0634 | 0.0046 | 0.0077 | 0.0044 | 0.0085 | 0.0100 | 2.77E-16 | 0.2958 | 0.3926 |
| rs17001654 | -0.0188 | -0.0306 | 0.0089 | 0.0053 | -0.0057 | 0.0113 | 0.03465 | 7.76E-09 | 0.6161 |
| rs6804842 | -0.0175 | -0.0185 | 0.0065 | 0.0031 | -0.0075 | 0.0083 | 0.007022 | 2.48E-09 | 0.3683 |
| rs5215 | -0.0706 | 0.0138 | 0.0066 | 0.0031 | -0.0042 | 0.0084 | 9.85E-27 | 1.13E-05 | 0.6195 |
| rs657452 | 0.0037 | 0.0227 | 0.0066 | 0.0031 | 0.0122 | 0.0083 | 0.5748 | 5.48E-13 | 0.1427 |
| rs2052673 | -0.0557 | -0.0011 | 0.0089 | 0.0051 | 0.0156 | 0.0108 | 3.88E-10 | 0.8292 | 0.1495 |
| rs1359939 | -0.0399 | -0.0107 | 0.0069 | 0.0041 | 0.0040 | 0.0089 | 7.74E-09 | 0.009061 | 0.6559 |
| rs2033732 | -0.0136 | -0.0192 | 0.0073 | 0.0035 | 0.0029 | 0.0094 | 0.06351 | 4.89E-08 | 0.76 |
| rs3789587 | -0.0674 | 0.0184 | 0.0112 | 0.0064 | 0.0321 | 0.0141 | 1.79E-09 | 0.00404 | 0.02319 |
| rs17772814 | -0.0746 | -0.0210 | 0.0125 | 0.0210 | -0.0160 | 0.0190 | 2.13E-09 | 0.3173 | 0.3998 |
| rs3802177 | -0.1077 | 0.0097 | 0.0069 | 0.0034 | 0.0197 | 0.0089 | 9.16E-55 | 0.004228 | 0.02647 |
| rs7633675 | -0.1085 | 0.0100 | 0.0068 | 0.0033 | 0.0210 | 0.0087 | 3.21E-57 | 0.002355 | 0.01644 |
| rs1561927 | -0.0420 | -0.0035 | 0.0072 | 0.0034 | -0.0102 | 0.0093 | 6.12E-09 | 0.3147 | 0.2688 |
| rs10171620 | 0.0399 | -0.0032 | 0.0065 | 0.0039 | 0.0089 | 0.0084 | 7.92E-10 | 0.4034 | 0.2864 |
| rs4925109 | 0.0476 | -0.0127 | 0.0069 | 0.0040 | -0.0030 | 0.0088 | 5.65E-12 | 0.001498 | 0.7298 |
| rs6937438 | -0.0514 | 0.0036 | 0.0070 | 0.0034 | 0.0106 | 0.0090 | 2.35E-13 | 0.2879 | 0.2396 |
| rs6905288 | 0.0374 | -0.0110 | 0.0065 | 0.0034 | -0.0103 | 0.0086 | 8.35E-09 | 0.001298 | 0.2346 |
| rs4135247 | -0.0438 | 0.0042 | 0.0065 | 0.0038 | -0.0003 | 0.0083 | 1.51E-11 | 0.269 | 0.9734 |
| rs9379084 | -0.0994 | 0.0165 | 0.0106 | 0.0056 | 0.0245 | 0.0132 | 5.48E-21 | 0.00303 | 0.06309 |
| rs10078815 | -0.0423 | -0.0018 | 0.0066 | 0.0044 | -0.0172 | 0.0085 | 1.43E-10 | 0.6825 | 0.0438 |
| rs17694555 | -0.0614 | 0.0108 | 0.0110 | 0.0053 | -0.0102 | 0.0144 | 2.34E-08 | 0.04213 | 0.477 |
| rs4804833 | 0.0480 | 0.0048 | 0.0066 | 0.0033 | 0.0044 | 0.0083 | 3.42E-13 | 0.1467 | 0.5949 |
| rs849135 | -0.0906 | 0.0121 | 0.0064 | 0.0031 | 0.0023 | 0.0082 | 1.13E-45 | 7.12E-05 | 0.7811 |
| rs2215383 | -0.0641 | 0.0042 | 0.0064 | 0.0031 | -0.0139 | 0.0081 | 1.06E-23 | 0.177 | 0.08832 |
| rs2237897 | -0.1924 | 0.0053 | 0.0166 | 0.0075 | 0.0364 | 0.0209 | 6.79E-31 | 0.4732 | 0.08167 |
| rs1885283 | 0.0727 | -0.0035 | 0.0102 | 0.0050 | 0.0042 | 0.0124 | 8.17E-13 | 0.4766 | 0.7343 |
| rs9860730 | 0.0539 | -0.0149 | 0.0070 | 0.0033 | -0.0064 | 0.0090 | 1.55E-14 | 8.18E-06 | 0.4729 |
| rs6600191 | 0.0587 | 0.0027 | 0.0085 | 0.0051 | 0.0145 | 0.0106 | 4.47E-12 | 0.5965 | 0.1713 |
| rs1903002 | -0.0362 | 0.0058 | 0.0064 | 0.0040 | -0.0086 | 0.0084 | 1.45E-08 | 0.1471 | 0.3066 |
| rs16952522 | -0.0884 | -0.0547 | 0.0161 | 0.0108 | -0.0025 | 0.0269 | 4.20E-08 | 4.53E-07 | 0.9274 |
| rs6990912 | 0.0426 | 0.0076 | 0.0067 | 0.0040 | -0.0113 | 0.0086 | 2.05E-10 | 0.05743 | 0.1867 |
| rs7325671 | 0.0545 | 0.0030 | 0.0096 | 0.0058 | -0.0119 | 0.0126 | 1.53E-08 | 0.605 | 0.3465 |
| rs3860374 | -0.0385 | 0.0059 | 0.0065 | 0.0038 | 0.0005 | 0.0083 | 3.02E-09 | 0.1205 | 0.9548 |
| rs12910825 | -0.0525 | 0.0010 | 0.0067 | 0.0032 | -0.0283 | 0.0085 | 4.70E-15 | 0.761 | 0.000848 |
| rs17712208 | 0.1854 | -0.0225 | 0.0185 | 0.0116 | -0.0177 | 0.0270 | 1.46E-23 | 0.05282 | 0.5109 |
| rs7219033 | 0.0387 | 0.0111 | 0.0068 | 0.0040 | -0.0018 | 0.0087 | 1.30E-08 | 0.00552 | 0.8389 |
| rs891368 | -0.0378 | -0.0026 | 0.0064 | 0.0038 | 0.0152 | 0.0082 | 3.25E-09 | 0.4938 | 0.0631 |
| rs2289739 | 0.0469 | 0.0052 | 0.0067 | 0.0042 | 0.0086 | 0.0087 | 2.58E-12 | 0.2157 | 0.3222 |
| rs459193 | -0.0722 | 0.0071 | 0.0073 | 0.0035 | 0.0147 | 0.0094 | 6.78E-23 | 0.04289 | 0.1172 |
| rs7903146 | 0.3142 | -0.0234 | 0.0070 | 0.0034 | 0.0182 | 0.0089 | 4.31E-438 | 1.11E-11 | 0.04123 |
| rs3217795 | 0.1065 | -0.0003 | 0.0119 | 0.0079 | 0.0020 | 0.0147 | 4.56E-19 | 0.9697 | 0.8939 |
| rs2287019 | 0.0219 | -0.0360 | 0.0082 | 0.0042 | -0.0005 | 0.0105 | 0.007327 | 4.59E-18 | 0.9649 |
| rs10068434 | -0.0393 | -0.0098 | 0.0071 | 0.0042 | 0.0215 | 0.0093 | 3.39E-08 | 0.01963 | 0.02002 |
| rs11165643 | 0.0216 | 0.0218 | 0.0065 | 0.0031 | -0.0052 | 0.0083 | 0.000877 | 2.07E-12 | 0.5266 |
| rs17522122 | 0.0356 | 0.0152 | 0.0064 | 0.0033 | 0.0001 | 0.0082 | 2.49E-08 | 3.15E-06 | 0.9939 |
| rs10406431 | 0.0603 | 0.0079 | 0.0065 | 0.0038 | 0.0014 | 0.0088 | 1.56E-20 | 0.03761 | 0.8755 |
| rs17405819 | 0.0126 | 0.0224 | 0.0069 | 0.0033 | -0.0048 | 0.0089 | 0.06825 | 2.07E-11 | 0.5898 |
| rs9925964 | 0.0219 | 0.0192 | 0.0067 | 0.0031 | 0.0021 | 0.0081 | 0.001082 | 8.11E-10 | 0.795 |
| rs17066842 | -0.0895 | -0.0626 | 0.0165 | 0.0083 | -0.0004 | 0.0222 | 6.30E-08 | 6.40E-14 | 0.9853 |
| rs6499653 | 0.0430 | 0.0269 | 0.0073 | 0.0037 | -0.0101 | 0.0096 | 4.44E-09 | 2.32E-13 | 0.2922 |
| rs7216064 | 0.0507 | 0.0109 | 0.0080 | 0.0046 | 0.0161 | 0.0102 | 1.87E-10 | 0.01781 | 0.1148 |
| rs4932265 | 0.0653 | -0.0030 | 0.0072 | 0.0043 | -0.0247 | 0.0093 | 1.59E-19 | 0.4854 | 0.007663 |
| rs11819995 | 0.0498 | 0.0037 | 0.0077 | 0.0046 | -0.0089 | 0.0098 | 1.30E-10 | 0.4212 | 0.3626 |
| rs7755852 | -0.0417 | -0.0069 | 0.0066 | 0.0046 | 0.0088 | 0.0089 | 2.59E-10 | 0.1363 | 0.3192 |
| rs2347101 | 0.0600 | -0.0005 | 0.0098 | 0.0050 | -0.0172 | 0.0126 | 1.09E-09 | 0.919 | 0.1733 |
| rs7896811 | -0.1465 | 0.0046 | 0.0091 | 0.0043 | -0.0133 | 0.0117 | 3.37E-58 | 0.282 | 0.255 |
| rs12325539 | -0.0410 | -0.0173 | 0.0065 | 0.0038 | -0.0093 | 0.0083 | 2.69E-10 | 5.30E-06 | 0.262 |
| rs7138803 | 0.0342 | 0.0315 | 0.0066 | 0.0031 | -0.0180 | 0.0084 | 2.16E-07 | 8.15E-24 | 0.03191 |
| rs2245368 | -0.0319 | -0.0317 | 0.0086 | 0.0057 | -0.0209 | 0.0123 | 0.000203 | 3.19E-08 | 0.08756 |
| rs12299509 | -0.0496 | 0.0058 | 0.0068 | 0.0034 | -0.0008 | 0.0087 | 3.15E-13 | 0.08817 | 0.9263 |
| rs7968682 | 0.0538 | -0.0087 | 0.0064 | 0.0031 | -0.0080 | 0.0083 | 3.65E-17 | 0.005209 | 0.3357 |
| rs3751239 | 0.0749 | -0.0031 | 0.0081 | 0.0047 | 0.0007 | 0.0102 | 1.54E-20 | 0.5095 | 0.945 |
| rs10176391 | 0.0749 | 0.0793 | 0.0310 | 0.0133 | -0.0886 | 0.0325 | 0.01566 | 2.64E-09 | 0.006409 |
| rs17013433 | 0.1134 | 0.0155 | 0.0175 | 0.0090 | -0.0030 | 0.0237 | 8.85E-11 | 0.08402 | 0.8979 |
| rs2251468 | -0.0474 | 0.0001 | 0.0067 | 0.0032 | 0.0029 | 0.0086 | 1.51E-12 | 0.9733 | 0.7374 |
| rs2820446 | 0.0593 | -0.0083 | 0.0070 | 0.0034 | 0.0020 | 0.0089 | 2.83E-17 | 0.01371 | 0.8248 |
| rs11030104 | 0.0193 | 0.0414 | 0.0080 | 0.0038 | -0.0030 | 0.0101 | 0.01529 | 5.56E-28 | 0.764 |
| rs4077404 | -0.0570 | -0.0069 | 0.0077 | 0.0037 | -0.0058 | 0.0097 | 1.88E-13 | 0.05918 | 0.549 |
| rs11847697 | 0.0464 | 0.0492 | 0.0160 | 0.0084 | -0.0062 | 0.0206 | 0.003774 | 3.99E-09 | 0.7618 |
| rs2365389 | -0.0097 | -0.0200 | 0.0065 | 0.0031 | 0.0061 | 0.0086 | 0.1351 | 1.63E-10 | 0.4736 |
| rs2121279 | 0.0287 | 0.0245 | 0.0093 | 0.0044 | -0.0092 | 0.0121 | 0.002071 | 2.31E-08 | 0.4437 |
| rs7733501 | 0.0446 | 0.0080 | 0.0071 | 0.0042 | -0.0166 | 0.0091 | 3.75E-10 | 0.05681 | 0.06691 |
| rs1007090 | -0.0440 | -0.0105 | 0.0068 | 0.0040 | -0.0020 | 0.0086 | 1.01E-10 | 0.008665 | 0.8149 |
| rs9947301 | -0.0260 | -0.0377 | 0.0121 | 0.0057 | 0.0005 | 0.0156 | 0.0323 | 3.70E-11 | 0.9743 |
| rs4256980 | -0.0268 | -0.0209 | 0.0067 | 0.0031 | 0.0198 | 0.0085 | 6.35E-05 | 2.90E-11 | 0.02006 |
| rs3764002 | -0.0511 | -0.0049 | 0.0073 | 0.0038 | 0.0038 | 0.0093 | 3.12E-12 | 0.1949 | 0.6795 |
| rs11709077 | -0.1040 | 0.0241 | 0.0097 | 0.0046 | 0.0103 | 0.0125 | 1.26E-26 | 1.81E-07 | 0.4072 |
| rs13389219 | -0.0605 | 0.0122 | 0.0065 | 0.0031 | 0.0002 | 0.0083 | 1.17E-20 | 8.96E-05 | 0.9837 |
| rs3887925 | 0.0539 | -0.0009 | 0.0065 | 0.0038 | -0.0035 | 0.0082 | 1.01E-16 | 0.8128 | 0.6673 |
| rs2972144 | -0.0911 | 0.0140 | 0.0066 | 0.0031 | -0.0044 | 0.0085 | 2.19E-43 | 7.01E-06 | 0.6006 |
| rs4740619 | 0.0146 | 0.0179 | 0.0064 | 0.0031 | -0.0047 | 0.0082 | 0.02226 | 4.56E-09 | 0.569 |
| rs340874 | -0.0678 | -0.0014 | 0.0065 | 0.0031 | 0.0041 | 0.0082 | 1.56E-25 | 0.6621 | 0.6199 |
| rs7899106 | -0.0113 | -0.0395 | 0.0144 | 0.0071 | -0.0107 | 0.0184 | 0.4342 | 2.96E-08 | 0.5617 |
| rs10750397 | 0.0394 | -0.0034 | 0.0071 | 0.0041 | 0.0013 | 0.0091 | 3.13E-08 | 0.407 | 0.8819 |
| rs4813428 | 0.0599 | 0.0274 | 0.0109 | 0.0065 | -0.0182 | 0.0141 | 3.78E-08 | 2.49E-05 | 0.1952 |
| rs11118352 | -0.0456 | 0.0054 | 0.0081 | 0.0048 | 0.0283 | 0.0104 | 1.55E-08 | 0.2606 | 0.006434 |
| rs2642588 | -0.0510 | 0.0001 | 0.0070 | 0.0042 | -0.0025 | 0.0089 | 3.59E-13 | 0.981 | 0.78 |
| rs6567160 | -0.0545 | -0.0556 | 0.0075 | 0.0036 | -0.0155 | 0.0096 | 4.85E-13 | 3.93E-53 | 0.1081 |
| rs6499646 | 0.0656 | 0.0410 | 0.0119 | 0.0077 | -0.0071 | 0.0154 | 3.89E-08 | 1.01E-07 | 0.6468 |
| rs2855812 | 0.0430 | 0.0024 | 0.0074 | 0.0036 | -0.0173 | 0.0095 | 7.28E-09 | 0.5098 | 0.06911 |
| rs947791 | 0.0606 | 0.0135 | 0.0081 | 0.0048 | -0.0166 | 0.0101 | 5.62E-14 | 0.004916 | 0.09873 |
| rs10225433 | -0.0767 | 0.0040 | 0.0138 | 0.0107 | -0.0432 | 0.0224 | 2.86E-08 | 0.7089 | 0.0537 |
| rs1377807 | 0.0524 | 0.0045 | 0.0069 | 0.0041 | -0.0133 | 0.0088 | 3.38E-14 | 0.2724 | 0.1326 |
| rs987237 | -0.0595 | -0.0440 | 0.0082 | 0.0040 | 0.0121 | 0.0105 | 3.20E-13 | 1.96E-28 | 0.2523 |
| rs11196236 | -0.0739 | -0.0004 | 0.0079 | 0.0048 | 0.0016 | 0.0100 | 4.92E-21 | 0.9336 | 0.8712 |
| rs12004367 | -0.0374 | 0.0028 | 0.0068 | 0.0040 | 0.0174 | 0.0088 | 3.90E-08 | 0.4839 | 0.04866 |
| rs2028150 | 0.0537 | 0.0060 | 0.0065 | 0.0038 | -0.0110 | 0.0083 | 1.31E-16 | 0.1143 | 0.1826 |
| rs1872992 | -0.0457 | 0.0093 | 0.0072 | 0.0035 | 0.0084 | 0.0094 | 2.52E-10 | 0.008036 | 0.371 |
| rs7240767 | -0.0372 | 0.0033 | 0.0066 | 0.0040 | 0.0034 | 0.0084 | 1.71E-08 | 0.4094 | 0.6895 |
| rs7669833 | -0.0572 | 0.0026 | 0.0070 | 0.0041 | 0.0100 | 0.0090 | 3.52E-16 | 0.526 | 0.268 |
| rs10513432 | 0.0920 | 0.0102 | 0.0163 | 0.0091 | 0.0052 | 0.0208 | 1.78E-08 | 0.2623 | 0.8032 |
| rs4788115 | -0.0237 | -0.0285 | 0.0089 | 0.0048 | 0.0077 | 0.0118 | 0.007745 | 3.28E-09 | 0.5161 |
| rs745805 | 0.0632 | -0.0079 | 0.0084 | 0.0051 | 0.0028 | 0.0108 | 4.52E-14 | 0.1214 | 0.7979 |
| rs879620 | 0.0038 | 0.0244 | 0.0067 | 0.0040 | -0.0006 | 0.0085 | 0.5707 | 1.06E-09 | 0.9429 |
| rs11075986 | 0.0590 | 0.0423 | 0.0120 | 0.0060 | -0.0015 | 0.0156 | 9.58E-07 | 1.23E-12 | 0.925 |
| rs2307111 | 0.0502 | 0.0235 | 0.0065 | 0.0031 | 0.0007 | 0.0083 | 1.05E-14 | 3.77E-14 | 0.9363 |
| rs1260326 | -0.0644 | -0.0124 | 0.0066 | 0.0032 | -0.0022 | 0.0083 | 1.62E-22 | 9.20E-05 | 0.793 |
| rs3888190 | 0.0291 | 0.0309 | 0.0065 | 0.0031 | 0.0138 | 0.0083 | 7.37E-06 | 3.14E-23 | 0.09577 |
| rs11030107 | -0.0316 | -0.0297 | 0.0073 | 0.0035 | 0.0019 | 0.0093 | 1.62E-05 | 3.15E-17 | 0.8415 |
| rs11688816 | -0.0193 | -0.0172 | 0.0064 | 0.0031 | 0.0082 | 0.0082 | 0.002512 | 1.89E-08 | 0.3173 |
| rs10749128 | -0.0460 | 0.0007 | 0.0074 | 0.0046 | -0.0088 | 0.0094 | 6.10E-10 | 0.879 | 0.3523 |
| rs12429545 | 0.0384 | 0.0334 | 0.0101 | 0.0047 | 0.0550 | 0.0122 | 0.000133 | 1.09E-12 | 6.09E-06 |
| rs9828639 | 0.0383 | 0.0001 | 0.0068 | 0.0040 | -0.0086 | 0.0086 | 1.83E-08 | 0.9801 | 0.3196 |
| rs8089364 | -0.0535 | -0.0510 | 0.0072 | 0.0035 | -0.0110 | 0.0092 | 1.31E-13 | 3.05E-49 | 0.2338 |
| rs11671664 | 0.0612 | -0.0277 | 0.0099 | 0.0053 | -0.0075 | 0.0133 | 7.61E-10 | 1.45E-07 | 0.5717 |
| rs10885410 | -0.0858 | 0.0062 | 0.0074 | 0.0052 | -0.0103 | 0.0100 | 8.12E-31 | 0.2331 | 0.3035 |
| rs543874 | -0.0547 | -0.0482 | 0.0081 | 0.0039 | 0.0104 | 0.0102 | 1.16E-11 | 2.62E-35 | 0.3093 |
| rs11752908 | -0.0456 | 0.0043 | 0.0065 | 0.0037 | -0.0077 | 0.0082 | 2.15E-12 | 0.2452 | 0.3478 |
| rs9312873 | 0.0684 | 0.0068 | 0.0111 | 0.0065 | -0.0222 | 0.0138 | 7.14E-10 | 0.2955 | 0.1059 |
| rs576674 | -0.0538 | 0.0082 | 0.0086 | 0.0044 | 0.0025 | 0.0110 | 3.70E-10 | 0.06097 | 0.8213 |
| rs2280141 | 0.0489 | 0.0008 | 0.0064 | 0.0038 | -0.0203 | 0.0081 | 1.91E-14 | 0.8333 | 0.01238 |
| rs1016287 | 0.0360 | 0.0229 | 0.0074 | 0.0034 | -0.0033 | 0.0089 | 1.28E-06 | 2.25E-11 | 0.7086 |
| rs13737 | -0.0481 | 0.0019 | 0.0075 | 0.0046 | 0.0150 | 0.0097 | 1.77E-10 | 0.6796 | 0.1238 |
| rs17630640 | 0.0537 | -0.0139 | 0.0096 | 0.0058 | 0.0005 | 0.0121 | 2.48E-08 | 0.01655 | 0.9655 |
| rs11126052 | 0.0401 | 0.0017 | 0.0072 | 0.0042 | 0.0281 | 0.0093 | 2.85E-08 | 0.6857 | 0.002576 |
| rs17689007 | -0.0468 | -0.0132 | 0.0065 | 0.0038 | 0.0077 | 0.0082 | 5.62E-13 | 0.000513 | 0.3498 |
| rs9316500 | 0.0387 | -0.0027 | 0.0070 | 0.0041 | 0.0062 | 0.0089 | 3.45E-08 | 0.5102 | 0.4826 |
| rs1061810 | 0.0502 | 0.0153 | 0.0070 | 0.0042 | -0.0058 | 0.0091 | 8.31E-13 | 0.00027 | 0.5252 |
| rs6963 | 0.0468 | 0.0040 | 0.0071 | 0.0042 | -0.0357 | 0.0090 | 4.92E-11 | 0.3409 | 7.31E-05 |
| rs16851483 | 0.0280 | 0.0483 | 0.0130 | 0.0077 | -0.0013 | 0.0164 | 0.03103 | 3.55E-10 | 0.9355 |
| rs1531583 | 0.1001 | -0.0126 | 0.0157 | 0.0101 | -0.0220 | 0.0207 | 1.85E-10 | 0.2122 | 0.2871 |
| rs13396935 | -0.0513 | -0.0596 | 0.0085 | 0.0042 | -0.0015 | 0.0108 | 1.46E-09 | 1.63E-46 | 0.8909 |
| rs9563615 | 0.0408 | 0.0103 | 0.0072 | 0.0042 | 0.0075 | 0.0091 | 1.63E-08 | 0.01419 | 0.4073 |
| rs7903767 | -0.0550 | -0.0022 | 0.0064 | 0.0037 | -0.0087 | 0.0082 | 7.21E-18 | 0.5521 | 0.2852 |
| rs17109221 | 0.0556 | 0.0252 | 0.0077 | 0.0046 | -0.0019 | 0.0099 | 7.18E-13 | 4.30E-08 | 0.8489 |
| rs1493694 | 0.0800 | -0.0014 | 0.0102 | 0.0050 | -0.0109 | 0.0132 | 3.35E-15 | 0.7791 | 0.4088 |
| rs1634746 | 0.0469 | 0.0047 | 0.0067 | 0.0047 | -0.0039 | 0.0094 | 2.58E-12 | 0.3173 | 0.681 |
| rs11583200 | -0.0050 | -0.0177 | 0.0065 | 0.0031 | 0.0034 | 0.0083 | 0.4412 | 1.48E-08 | 0.6872 |
| rs11187152 | -0.0695 | 0.0179 | 0.0124 | 0.0089 | -0.0003 | 0.0164 | 1.85E-08 | 0.0443 | 0.9868 |
| rs4776970 | 0.0378 | 0.0244 | 0.0067 | 0.0031 | 0.0075 | 0.0085 | 1.69E-08 | 8.87E-15 | 0.3744 |
| rs231907 | -0.0389 | 0.0070 | 0.0070 | 0.0036 | -0.0072 | 0.0093 | 2.94E-08 | 0.05539 | 0.4406 |
| rs569255 | -0.0396 | 0.0000 | 0.0064 | 0.0031 | 0.0017 | 0.0082 | 5.64E-10 | 0.9905 | 0.8368 |
| rs4273712 | -0.0591 | 0.0066 | 0.0071 | 0.0035 | 0.0302 | 0.0092 | 1.03E-16 | 0.06027 | 0.000978 |
| rs3094515 | -0.0464 | -0.0018 | 0.0072 | 0.0051 | -0.0129 | 0.0099 | 1.34E-10 | 0.7241 | 0.1937 |
| rs10182181 | -0.0102 | -0.0307 | 0.0064 | 0.0031 | -0.0092 | 0.0081 | 0.1103 | 8.78E-24 | 0.2575 |
| rs16841827 | -0.0822 | 0.0050 | 0.0133 | 0.0079 | 0.0206 | 0.0168 | 6.34E-10 | 0.5268 | 0.2208 |
| rs13021737 | -0.0493 | -0.0601 | 0.0085 | 0.0040 | -0.0018 | 0.0112 | 6.13E-09 | 1.11E-50 | 0.8707 |
| rs255761 | 0.0541 | -0.0135 | 0.0080 | 0.0047 | 0.0018 | 0.0102 | 1.06E-11 | 0.004074 | 0.8628 |
| rs4585612 | -0.0414 | 0.0088 | 0.0065 | 0.0038 | 0.0092 | 0.0083 | 1.80E-10 | 0.02057 | 0.2673 |
| rs17094222 | -0.0263 | -0.0249 | 0.0079 | 0.0038 | 0.0122 | 0.0102 | 0.000811 | 5.94E-11 | 0.2287 |
| rs10848958 | -0.0451 | 0.0044 | 0.0083 | 0.0047 | 0.0143 | 0.0102 | 4.97E-08 | 0.3492 | 0.1628 |
| rs474513 | 0.0399 | -0.0116 | 0.0064 | 0.0038 | -0.0157 | 0.0085 | 4.18E-10 | 0.002268 | 0.06372 |
| rs5771069 | -0.0409 | 0.0008 | 0.0065 | 0.0043 | 0.0004 | 0.0082 | 2.97E-10 | 0.8524 | 0.9586 |
| rs1574190 | 0.0476 | -0.0024 | 0.0067 | 0.0038 | 0.0088 | 0.0083 | 1.22E-12 | 0.5277 | 0.2926 |
| rs7143394 | -0.0364 | -0.0068 | 0.0064 | 0.0037 | 0.0088 | 0.0082 | 1.20E-08 | 0.06609 | 0.2829 |
| rs17168486 | 0.0672 | -0.0047 | 0.0083 | 0.0041 | 0.0011 | 0.0107 | 4.50E-16 | 0.2567 | 0.9176 |
| rs6857 | -0.0660 | -0.0213 | 0.0086 | 0.0056 | 0.0007 | 0.0108 | 1.50E-14 | 0.00013 | 0.9519 |
| rs12401738 | 0.0032 | 0.0211 | 0.0067 | 0.0033 | 0.0169 | 0.0084 | 0.633 | 1.15E-10 | 0.04527 |
| rs11496066 | 0.0508 | 0.0002 | 0.0083 | 0.0048 | 0.0004 | 0.0105 | 8.17E-10 | 0.9668 | 0.9661 |
| rs6901126 | -0.0356 | -0.0093 | 0.0065 | 0.0031 | 0.0072 | 0.0082 | 4.16E-08 | 0.002358 | 0.3837 |
| rs12446632 | -0.0392 | -0.0403 | 0.0093 | 0.0046 | 0.0096 | 0.0117 | 2.59E-05 | 1.48E-18 | 0.4117 |
| rs7933438 | -0.0580 | 0.0050 | 0.0093 | 0.0052 | -0.0084 | 0.0115 | 4.84E-10 | 0.3363 | 0.4624 |
| rs329122 | 0.0366 | -0.0086 | 0.0065 | 0.0031 | -0.0059 | 0.0082 | 1.72E-08 | 0.005809 | 0.4726 |
| rs12187196 | -0.0399 | -0.0005 | 0.0065 | 0.0038 | -0.0142 | 0.0083 | 7.92E-10 | 0.8953 | 0.08843 |
| rs17685538 | 0.0954 | -0.0140 | 0.0107 | 0.0055 | -0.0046 | 0.0142 | 4.15E-19 | 0.01133 | 0.7485 |
| rs9267658 | -0.0716 | 0.0062 | 0.0091 | 0.0044 | 0.0317 | 0.0117 | 3.83E-15 | 0.162 | 0.006838 |
| rs2075650 | 0.0620 | 0.0258 | 0.0091 | 0.0045 | -0.0039 | 0.0115 | 1.00E-11 | 1.25E-08 | 0.735 |
| rs2456530 | 0.0540 | 0.0114 | 0.0096 | 0.0058 | 0.0048 | 0.0122 | 2.07E-08 | 0.04935 | 0.6963 |
| rs1375561 | 0.0099 | 0.0179 | 0.0067 | 0.0031 | -0.0095 | 0.0085 | 0.1396 | 9.93E-09 | 0.2671 |
| rs2292662 | -0.0645 | 0.0038 | 0.0088 | 0.0051 | 0.0140 | 0.0113 | 2.24E-13 | 0.4562 | 0.2142 |
| rs17024393 | -0.0483 | -0.0658 | 0.0181 | 0.0088 | 0.0184 | 0.0249 | 0.007664 | 7.03E-14 | 0.4592 |
| rs2001433 | -0.0442 | -0.0082 | 0.0065 | 0.0038 | 0.0206 | 0.0083 | 9.84E-12 | 0.03094 | 0.01311 |
| rs889512 | 0.1111 | -0.0037 | 0.0107 | 0.0064 | 0.0092 | 0.0136 | 2.40E-25 | 0.5632 | 0.4961 |
| rs10946394 | -0.0566 | 0.0008 | 0.0074 | 0.0037 | -0.0112 | 0.0099 | 2.66E-14 | 0.8285 | 0.2578 |
| rs6878122 | -0.0605 | 0.0071 | 0.0070 | 0.0035 | 0.0027 | 0.0089 | 6.45E-18 | 0.04305 | 0.7633 |
| rs11108094 | 0.0714 | 0.0026 | 0.0128 | 0.0078 | 0.0343 | 0.0161 | 2.28E-08 | 0.7389 | 0.03346 |
| rs17746916 | 0.1491 | -0.0210 | 0.0133 | 0.0095 | 0.0106 | 0.0177 | 3.53E-29 | 0.02707 | 0.5495 |
| rs3826482 | 0.0367 | -0.0044 | 0.0066 | 0.0035 | -0.0141 | 0.0083 | 2.64E-08 | 0.2149 | 0.09081 |
| rs6955948 | 0.0395 | 0.0064 | 0.0070 | 0.0043 | -0.0205 | 0.0092 | 1.79E-08 | 0.1367 | 0.02563 |
| rs1496653 | 0.0665 | -0.0069 | 0.0079 | 0.0036 | -0.0194 | 0.0100 | 2.49E-17 | 0.05717 | 0.05263 |
| rs7203521 | 0.0424 | 0.0326 | 0.0066 | 0.0032 | -0.0074 | 0.0084 | 1.29E-10 | 3.46E-24 | 0.3767 |
| rs703972 | -0.0698 | -0.0045 | 0.0065 | 0.0037 | 0.0013 | 0.0081 | 5.77E-27 | 0.2239 | 0.87 |
| rs3808415 | -0.0349 | -0.0107 | 0.0064 | 0.0037 | 0.0114 | 0.0082 | 4.65E-08 | 0.003829 | 0.1632 |
| rs2244510 | 0.0354 | -0.0010 | 0.0065 | 0.0040 | 0.0066 | 0.0084 | 4.95E-08 | 0.8026 | 0.4306 |
| rs11727676 | -0.0532 | 0.0358 | 0.0109 | 0.0064 | -0.0046 | 0.0152 | 1.03E-06 | 2.55E-08 | 0.7633 |
| rs4650985 | -0.0383 | -0.0103 | 0.0070 | 0.0034 | -0.0054 | 0.0090 | 4.77E-08 | 0.002492 | 0.5485 |
| rs31911 | 0.0369 | 0.0056 | 0.0064 | 0.0037 | -0.0088 | 0.0082 | 7.58E-09 | 0.1301 | 0.2818 |
| rs243019 | -0.0588 | 0.0006 | 0.0064 | 0.0031 | 0.0013 | 0.0082 | 3.37E-20 | 0.8496 | 0.8698 |
| rs6808574 | -0.0610 | -0.0018 | 0.0066 | 0.0032 | -0.0139 | 0.0083 | 2.29E-20 | 0.5823 | 0.09471 |
| rs10811661 | 0.1597 | -0.0043 | 0.0086 | 0.0042 | 0.0074 | 0.0108 | 3.16E-77 | 0.3024 | 0.4942 |
| rs11057405 | 0.0306 | -0.0307 | 0.0109 | 0.0055 | 0.0027 | 0.0138 | 0.004952 | 2.02E-08 | 0.8462 |
| rs10770141 | 0.0658 | 0.0105 | 0.0068 | 0.0044 | -0.0160 | 0.0085 | 4.11E-22 | 0.01702 | 0.06104 |
| rs6031580 | -0.0429 | 0.0033 | 0.0074 | 0.0043 | 0.0072 | 0.0094 | 7.89E-09 | 0.4428 | 0.4451 |
| rs9400239 | -0.0079 | -0.0188 | 0.0070 | 0.0033 | -0.0139 | 0.0089 | 0.2601 | 1.61E-08 | 0.1198 |
| rs8028830 | -0.0562 | -0.0079 | 0.0091 | 0.0052 | 0.0170 | 0.0115 | 6.84E-10 | 0.1287 | 0.1396 |
| rs2925979 | 0.0546 | -0.0012 | 0.0070 | 0.0033 | -0.0163 | 0.0089 | 7.07E-15 | 0.721 | 0.0654 |
| rs1421085 | -0.1218 | -0.0813 | 0.0065 | 0.0031 | 0.0238 | 0.0087 | 1.52E-78 | 8.83E-151 | 0.006019 |
| rs13431750 | -0.1117 | -0.0196 | 0.0204 | 0.0104 | 0.0478 | 0.0271 | 4.48E-08 | 0.05999 | 0.07743 |
| rs3783394 | -0.0380 | -0.0157 | 0.0067 | 0.0038 | 0.0249 | 0.0085 | 1.42E-08 | 3.60E-05 | 0.003517 |
| rs545572 | -0.0762 | -0.0004 | 0.0068 | 0.0048 | -0.0047 | 0.0086 | 4.24E-29 | 0.9336 | 0.584 |
| rs10444213 | 0.0419 | 0.0090 | 0.0065 | 0.0042 | 0.0160 | 0.0086 | 1.09E-10 | 0.03212 | 0.06263 |
| rs3820981 | 0.0402 | -0.0102 | 0.0065 | 0.0031 | 0.0052 | 0.0083 | 5.92E-10 | 0.001063 | 0.5335 |
| rs3754346 | 0.0795 | 0.0084 | 0.0076 | 0.0046 | -0.0264 | 0.0099 | 2.45E-25 | 0.06784 | 0.007661 |
| rs11708067 | 0.0882 | -0.0152 | 0.0077 | 0.0038 | -0.0239 | 0.0096 | 5.05E-30 | 6.23E-05 | 0.01313 |
| rs7132277 | -0.0475 | 0.0032 | 0.0082 | 0.0041 | 0.0121 | 0.0106 | 6.02E-09 | 0.4374 | 0.2532 |
| rs2817419 | 0.0313 | 0.0275 | 0.0073 | 0.0035 | 0.0133 | 0.0094 | 1.95E-05 | 3.66E-15 | 0.1551 |
| rs3817334 | 0.0344 | 0.0262 | 0.0065 | 0.0031 | -0.0087 | 0.0083 | 1.16E-07 | 5.15E-17 | 0.2905 |
| rs9579083 | 0.0273 | 0.0295 | 0.0081 | 0.0047 | 0.0181 | 0.0105 | 0.000709 | 3.46E-10 | 0.08324 |
| rs13107325 | 0.0460 | 0.0477 | 0.0135 | 0.0068 | -0.0005 | 0.0163 | 0.00066 | 1.83E-12 | 0.9744 |
| rs2767036 | -0.0389 | -0.0091 | 0.0070 | 0.0047 | 0.0031 | 0.0090 | 2.94E-08 | 0.05285 | 0.7344 |
| rs13078960 | -0.0118 | -0.0297 | 0.0081 | 0.0039 | -0.0005 | 0.0101 | 0.1433 | 1.74E-14 | 0.9627 |
| rs10830963 | -0.1010 | -0.0062 | 0.0071 | 0.0036 | -0.0139 | 0.0095 | 1.12E-45 | 0.08769 | 0.1409 |
| rs6903706 | -0.0620 | -0.0046 | 0.0084 | 0.0039 | -0.0080 | 0.0105 | 1.34E-13 | 0.2468 | 0.4437 |
| rs11677557 | 0.0590 | -0.0010 | 0.0090 | 0.0043 | -0.0012 | 0.0115 | 5.66E-11 | 0.8171 | 0.9182 |
| rs6909558 | 0.0848 | -0.0057 | 0.0115 | 0.0058 | 0.0184 | 0.0151 | 1.80E-13 | 0.3242 | 0.2221 |
| rs17662402 | 0.0835 | 0.0023 | 0.0142 | 0.0072 | 0.0046 | 0.0181 | 4.52E-09 | 0.7531 | 0.8001 |
| rs320369 | 0.0372 | 0.0050 | 0.0068 | 0.0040 | 0.0012 | 0.0088 | 4.60E-08 | 0.2113 | 0.8878 |
| rs12865499 | -0.0423 | 0.0105 | 0.0075 | 0.0044 | -0.0049 | 0.0096 | 2.01E-08 | 0.01702 | 0.6093 |
| rs6457684 | -0.0369 | -0.0031 | 0.0065 | 0.0038 | 0.0143 | 0.0085 | 1.31E-08 | 0.4146 | 0.09261 |
| rs3849570 | 0.0113 | 0.0188 | 0.0067 | 0.0034 | -0.0031 | 0.0085 | 0.09173 | 2.60E-08 | 0.7149 |
| rs2921077 | 0.0373 | 0.0094 | 0.0065 | 0.0038 | -0.0242 | 0.0083 | 9.14E-09 | 0.01337 | 0.003571 |
| rs12583517 | -0.0423 | -0.0141 | 0.0077 | 0.0044 | 0.0065 | 0.0096 | 4.78E-08 | 0.001353 | 0.4948 |
| rs6012876 | 0.0428 | -0.0074 | 0.0065 | 0.0040 | 0.0248 | 0.0083 | 4.31E-11 | 0.06431 | 0.002869 |
| rs3810291 | 0.0434 | 0.0283 | 0.0069 | 0.0036 | 0.0006 | 0.0089 | 3.38E-10 | 4.81E-15 | 0.9454 |
| rs3736485 | 0.0213 | 0.0176 | 0.0065 | 0.0031 | 0.0052 | 0.0082 | 0.001034 | 7.41E-09 | 0.5262 |
| rs1848068 | 0.0388 | -0.0014 | 0.0067 | 0.0040 | -0.0120 | 0.0086 | 7.03E-09 | 0.7263 | 0.1605 |
| rs12347779 | -0.0792 | 0.0107 | 0.0144 | 0.0092 | 0.0238 | 0.0220 | 4.22E-08 | 0.2448 | 0.2797 |
| rs755162 | 0.0402 | -0.0028 | 0.0070 | 0.0042 | 0.0000 | 0.0090 | 1.00E-08 | 0.505 | 0.9988 |
| rs7819706 | 0.0669 | 0.0002 | 0.0101 | 0.0048 | 0.0034 | 0.0126 | 2.82E-11 | 0.959 | 0.7854 |
| rs7729395 | 0.1472 | 0.0008 | 0.0149 | 0.0082 | -0.0232 | 0.0198 | 4.14E-23 | 0.9262 | 0.2401 |
| rs163184 | -0.0780 | 0.0036 | 0.0066 | 0.0031 | 0.0180 | 0.0082 | 2.90E-32 | 0.2497 | 0.02807 |
| rs10401969 | -0.0876 | 0.0188 | 0.0120 | 0.0063 | -0.0332 | 0.0160 | 3.46E-13 | 0.002761 | 0.03812 |
| rs9269081 | -0.0566 | 0.0063 | 0.0071 | 0.0042 | 0.0039 | 0.0091 | 1.87E-15 | 0.1336 | 0.669 |
| rs2820292 | -0.0257 | -0.0195 | 0.0065 | 0.0031 | -0.0023 | 0.0082 | 7.53E-05 | 1.83E-10 | 0.7818 |
| rs11243150 | 0.0512 | 0.0002 | 0.0065 | 0.0038 | -0.0014 | 0.0084 | 3.09E-15 | 0.958 | 0.8719 |
| rs11023461 | -0.0751 | 0.0025 | 0.0129 | 0.0073 | -0.0058 | 0.0178 | 5.49E-09 | 0.7293 | 0.7459 |
| rs244418 | -0.0429 | -0.0149 | 0.0065 | 0.0037 | -0.0088 | 0.0082 | 3.88E-11 | 5.65E-05 | 0.2863 |
| rs290483 | 0.0701 | -0.0038 | 0.0067 | 0.0033 | -0.0065 | 0.0084 | 1.30E-25 | 0.246 | 0.4391 |
| rs1516725 | -0.0267 | -0.0451 | 0.0094 | 0.0046 | -0.0118 | 0.0119 | 0.004605 | 1.89E-22 | 0.3231 |
| rs12286929 | -0.0234 | -0.0217 | 0.0064 | 0.0031 | -0.0215 | 0.0081 | 0.000249 | 1.31E-12 | 0.008416 |
| rs154021 | 0.0417 | -0.0006 | 0.0074 | 0.0044 | 0.0019 | 0.0096 | 2.03E-08 | 0.8915 | 0.8451 |
| rs177045 | -0.0500 | -0.0046 | 0.0069 | 0.0042 | -0.0103 | 0.0089 | 4.64E-13 | 0.2734 | 0.247 |
| rs8037894 | -0.0467 | -0.0041 | 0.0065 | 0.0038 | -0.0039 | 0.0083 | 6.29E-13 | 0.2806 | 0.6339 |
| rs2244648 | -0.0363 | -0.0071 | 0.0065 | 0.0040 | 0.0160 | 0.0083 | 2.25E-08 | 0.0759 | 0.0547 |
| rs4812034 | 0.0416 | 0.0023 | 0.0064 | 0.0038 | -0.0142 | 0.0082 | 7.35E-11 | 0.545 | 0.08111 |
| rs7599312 | -0.0049 | -0.0220 | 0.0072 | 0.0034 | 0.0031 | 0.0092 | 0.4976 | 1.17E-10 | 0.7374 |
| rs12523853 | 0.0973 | -0.0110 | 0.0169 | 0.0106 | 0.0342 | 0.0250 | 7.83E-09 | 0.2987 | 0.1707 |
| rs231361 | 0.0607 | 0.0013 | 0.0074 | 0.0036 | -0.0049 | 0.0095 | 3.21E-16 | 0.7102 | 0.6083 |
| rs3918298 | -0.1349 | -0.0239 | 0.0209 | 0.0209 | 0.0454 | 0.0307 | 1.18E-10 | 0.2528 | 0.139 |
| rs2624847 | -0.0410 | -0.0016 | 0.0074 | 0.0037 | 0.0066 | 0.0094 | 3.48E-08 | 0.6657 | 0.4815 |
| rs7141420 | 0.0317 | 0.0235 | 0.0064 | 0.0031 | -0.0070 | 0.0082 | 6.93E-07 | 1.23E-14 | 0.3925 |
| rs7901695 | -0.2847 | 0.0213 | 0.0069 | 0.0034 | -0.0193 | 0.0089 | 5.13E-371 | 2.66E-10 | 0.03007 |
| rs12354626 | -0.1090 | -0.0005 | 0.0192 | 0.0116 | 0.0146 | 0.0269 | 1.28E-08 | 0.9654 | 0.5866 |
| rs702634 | 0.0503 | -0.0122 | 0.0069 | 0.0033 | 0.0034 | 0.0088 | 3.37E-13 | 0.000258 | 0.6973 |
| rs11257655 | 0.0859 | -0.0077 | 0.0077 | 0.0038 | -0.0180 | 0.0100 | 1.46E-28 | 0.04285 | 0.07173 |
| rs208302 | 0.0414 | 0.0062 | 0.0075 | 0.0046 | 0.0104 | 0.0097 | 3.98E-08 | 0.1777 | 0.2835 |
| rs10132280 | -0.0324 | -0.0230 | 0.0070 | 0.0034 | -0.0137 | 0.0090 | 3.86E-06 | 1.14E-11 | 0.1281 |
| rs5758223 | 0.0401 | 0.0089 | 0.0071 | 0.0041 | 0.0104 | 0.0090 | 1.78E-08 | 0.02995 | 0.2477 |
| rs6891076 | 0.0657 | -0.0007 | 0.0109 | 0.0053 | -0.0214 | 0.0143 | 1.60E-09 | 0.895 | 0.1349 |
| rs2041547 | -0.0408 | -0.0008 | 0.0064 | 0.0031 | -0.0104 | 0.0082 | 1.68E-10 | 0.8048 | 0.2084 |
| rs2971669 | 0.0602 | 0.0032 | 0.0080 | 0.0040 | 0.0111 | 0.0100 | 3.87E-14 | 0.4225 | 0.2689 |
| rs8192675 | 0.0659 | -0.0124 | 0.0070 | 0.0034 | 0.0066 | 0.0090 | 5.77E-21 | 0.000218 | 0.4618 |
| rs4689381 | 0.0380 | 0.0035 | 0.0066 | 0.0031 | 0.0005 | 0.0082 | 8.37E-09 | 0.2612 | 0.9483 |
| rs11126666 | 0.0005 | 0.0207 | 0.0072 | 0.0034 | 0.0126 | 0.0092 | 0.9448 | 1.33E-09 | 0.1734 |
| rs1928295 | 0.0121 | 0.0188 | 0.0064 | 0.0031 | 0.0019 | 0.0082 | 0.05816 | 7.91E-10 | 0.8201 |
| rs2650492 | 0.0060 | 0.0207 | 0.0070 | 0.0035 | 0.0063 | 0.0089 | 0.3924 | 1.92E-09 | 0.4786 |
| rs10885414 | 0.0949 | -0.0087 | 0.0070 | 0.0037 | -0.0009 | 0.0093 | 1.07E-41 | 0.0192 | 0.9267 |
| rs12454712 | 0.0477 | -0.0169 | 0.0068 | 0.0039 | -0.0018 | 0.0092 | 2.40E-12 | 1.46E-05 | 0.8476 |
| rs1359790 | -0.0817 | 0.0030 | 0.0071 | 0.0034 | -0.0137 | 0.0090 | 1.76E-30 | 0.3814 | 0.1279 |
| rs10937721 | 0.0850 | 0.0031 | 0.0066 | 0.0033 | -0.0017 | 0.0084 | 5.39E-38 | 0.3438 | 0.8427 |
| rs12885454 | -0.0215 | -0.0207 | 0.0067 | 0.0033 | 0.0055 | 0.0085 | 0.001334 | 1.94E-10 | 0.5164 |
| rs3134996 | -0.0417 | -0.0003 | 0.0069 | 0.0044 | -0.0108 | 0.0085 | 1.60E-09 | 0.9456 | 0.2074 |
| rs11591689 | 0.0486 | -0.0047 | 0.0077 | 0.0044 | 0.0084 | 0.0097 | 3.55E-10 | 0.2854 | 0.3839 |
| rs17296280 | -0.0456 | -0.0104 | 0.0070 | 0.0041 | 0.0169 | 0.0085 | 8.02E-11 | 0.01119 | 0.04574 |
| rs2497313 | 0.0614 | 0.0056 | 0.0107 | 0.0053 | 0.0097 | 0.0135 | 8.96E-09 | 0.2944 | 0.4719 |
| rs8097783 | -0.0259 | -0.0398 | 0.0125 | 0.0060 | 0.0291 | 0.0156 | 0.03764 | 4.20E-11 | 0.06202 |
| rs1167827 | -0.0100 | -0.0202 | 0.0065 | 0.0033 | -0.0056 | 0.0083 | 0.1234 | 6.33E-10 | 0.5008 |
| rs2065501 | 0.0443 | -0.0049 | 0.0073 | 0.0037 | -0.0029 | 0.0097 | 1.50E-09 | 0.1797 | 0.7652 |
| rs16860219 | 0.1222 | -0.0021 | 0.0208 | 0.0114 | 0.0731 | 0.0275 | 4.49E-09 | 0.8522 | 0.007895 |
| rs758747 | 0.0297 | 0.0225 | 0.0072 | 0.0037 | -0.0319 | 0.0093 | 3.94E-05 | 7.47E-10 | 0.000593 |
| rs2207139 | -0.0573 | -0.0447 | 0.0083 | 0.0040 | 0.0175 | 0.0108 | 4.29E-12 | 4.13E-29 | 0.105 |
| rs1317006 | -0.0147 | -0.0214 | 0.0069 | 0.0034 | 0.0034 | 0.0089 | 0.0334 | 4.97E-10 | 0.705 |
| rs6948511 | 0.0641 | 0.0007 | 0.0112 | 0.0055 | 0.0040 | 0.0145 | 1.06E-08 | 0.8965 | 0.7839 |
| rs2176598 | 0.0457 | 0.0198 | 0.0073 | 0.0036 | 0.0000 | 0.0094 | 4.51E-10 | 2.97E-08 | 0.9993 |
| rs12566985 | 0.0033 | -0.0242 | 0.0064 | 0.0031 | -0.0012 | 0.0082 | 0.6054 | 3.28E-15 | 0.8808 |
| rs11191560 | 0.0018 | -0.0308 | 0.0115 | 0.0053 | 0.0139 | 0.0149 | 0.8758 | 8.45E-09 | 0.3511 |
| rs1665901 | 0.0398 | 0.0059 | 0.0068 | 0.0048 | 0.0073 | 0.0094 | 4.97E-09 | 0.219 | 0.4391 |
| rs997258 | 0.0376 | 0.0026 | 0.0065 | 0.0038 | 0.0068 | 0.0082 | 6.95E-09 | 0.4938 | 0.4041 |
| rs1882297 | 0.0474 | 0.0041 | 0.0070 | 0.0035 | -0.0075 | 0.0087 | 1.41E-11 | 0.234 | 0.3934 |
| rs2009075 | 0.1707 | -0.0009 | 0.0216 | 0.0131 | 0.0070 | 0.0281 | 2.49E-15 | 0.9452 | 0.8027 |
| rs17030845 | -0.1191 | 0.0109 | 0.0108 | 0.0052 | -0.0194 | 0.0133 | 2.35E-28 | 0.03634 | 0.144 |
| rs2612069 | -0.1034 | 0.0021 | 0.0109 | 0.0064 | -0.0153 | 0.0139 | 2.19E-21 | 0.7428 | 0.2714 |
| rs2241896 | 0.0451 | -0.0005 | 0.0070 | 0.0041 | 0.0037 | 0.0087 | 1.28E-10 | 0.9029 | 0.6677 |
| rs10968576 | -0.0370 | -0.0249 | 0.0068 | 0.0033 | 0.0060 | 0.0087 | 5.43E-08 | 6.61E-14 | 0.4951 |
| rs1447059 | -0.0522 | 0.0085 | 0.0066 | 0.0032 | -0.0067 | 0.0085 | 2.50E-15 | 0.008232 | 0.4319 |
| rs4239217 | -0.0677 | 0.0088 | 0.0066 | 0.0035 | 0.0052 | 0.0086 | 1.03E-24 | 0.0126 | 0.5451 |
| rs6545714 | -0.0363 | -0.0180 | 0.0065 | 0.0031 | 0.0115 | 0.0083 | 2.25E-08 | 8.87E-09 | 0.1677 |
| rs7667864 | -0.0402 | -0.0044 | 0.0071 | 0.0042 | 0.0114 | 0.0090 | 1.64E-08 | 0.2948 | 0.2054 |
| rs7748720 | 0.0421 | -0.0027 | 0.0076 | 0.0037 | -0.0078 | 0.0099 | 3.63E-08 | 0.4658 | 0.4297 |
| rs505922 | -0.0473 | -0.0009 | 0.0068 | 0.0033 | -0.0016 | 0.0090 | 3.65E-12 | 0.7921 | 0.8611 |
| rs1127215 | -0.0491 | 0.0024 | 0.0065 | 0.0044 | 0.0095 | 0.0082 | 3.92E-14 | 0.5854 | 0.2483 |
| *Instrumental variables of T2D and BMI on BMD (N=379)* | | | | |  |  |  |  |  |
| SNP | beta_T2D | beta_BMI | SE_T2D | se_BMI | beta_BMD | se_BMD | P_T2D | P_BMI | P_BMD |
| rs3754346 | 0.0795 | 0.0084 | 0.0076 | 0.0046 | 0.0140 | 0.0022 | 2.45E-25 | 0.06784 | 7.80E-06 |
| rs657452 | 0.0037 | 0.0227 | 0.0066 | 0.0031 | -0.0016 | 0.0019 | 0.5748 | 5.48E-13 | 1 |
| rs11583200 | -0.0050 | -0.0177 | 0.0065 | 0.0031 | -0.0011 | 0.0019 | 0.4412 | 1.48E-08 | 0.34 |
| rs3789587 | -0.0674 | 0.0184 | 0.0112 | 0.0064 | -0.0174 | 0.0032 | 1.79E-09 | 0.00404 | 5.10E-05 |
| rs3101336 | -0.0248 | -0.0334 | 0.0066 | 0.0031 | -0.0098 | 0.0019 | 0.00017 | 2.66E-26 | 7.10E-05 |
| rs12566985 | 0.0033 | -0.0242 | 0.0064 | 0.0031 | -0.0094 | 0.0018 | 0.6054 | 3.28E-15 | 3.80E-06 |
| rs12401738 | 0.0032 | 0.0211 | 0.0067 | 0.0033 | -0.0029 | 0.0019 | 0.633 | 1.15E-10 | 0.12 |
| rs11165643 | 0.0216 | 0.0218 | 0.0065 | 0.0031 | 0.0001 | 0.0019 | 0.000877 | 2.07E-12 | 0.97 |
| rs17024393 | -0.0483 | -0.0658 | 0.0181 | 0.0088 | -0.0042 | 0.0058 | 0.007664 | 7.03E-14 | 0.73 |
| rs1127215 | -0.0491 | 0.0024 | 0.0065 | 0.0044 | -0.0034 | 0.0019 | 3.92E-14 | 0.5854 | 0.14 |
| rs320369 | 0.0372 | 0.0050 | 0.0068 | 0.0040 | 0.0021 | 0.0020 | 4.60E-08 | 0.2113 | 0.61 |
| rs1493694 | 0.0800 | -0.0014 | 0.0102 | 0.0050 | -0.0066 | 0.0029 | 3.35E-15 | 0.7791 | 0.074 |
| rs1359939 | -0.0399 | -0.0107 | 0.0069 | 0.0041 | -0.0027 | 0.0020 | 7.74E-09 | 0.009061 | 0.084 |
| rs693232 | -0.0542 | -0.0480 | 0.0080 | 0.0042 | -0.0115 | 0.0022 | 9.67E-12 | 7.78E-31 | 4.90E-06 |
| rs543874 | -0.0547 | -0.0482 | 0.0081 | 0.0039 | -0.0118 | 0.0023 | 1.16E-11 | 2.62E-35 | 3.60E-06 |
| rs4650985 | -0.0383 | -0.0103 | 0.0070 | 0.0034 | -0.0054 | 0.0020 | 4.77E-08 | 0.002492 | 0.0032 |
| rs2820292 | -0.0257 | -0.0195 | 0.0065 | 0.0031 | -0.0058 | 0.0018 | 7.53E-05 | 1.83E-10 | 0.027 |
| rs3862948 | 0.0414 | 0.0133 | 0.0075 | 0.0044 | 0.0039 | 0.0022 | 3.98E-08 | 0.002505 | 0.26 |
| rs17712208 | 0.1854 | -0.0225 | 0.0185 | 0.0116 | -0.0014 | 0.0050 | 1.46E-23 | 0.05282 | 0.65 |
| rs340874 | -0.0678 | -0.0014 | 0.0065 | 0.0031 | -0.0042 | 0.0018 | 1.56E-25 | 0.6621 | 0.088 |
| rs2820446 | 0.0593 | -0.0083 | 0.0070 | 0.0034 | -0.0023 | 0.0020 | 2.83E-17 | 0.01371 | 0.34 |
| rs11118352 | -0.0456 | 0.0054 | 0.0081 | 0.0048 | -0.0157 | 0.0023 | 1.55E-08 | 0.2606 | 7.80E-09 |
| rs10176391 | 0.0749 | 0.0793 | 0.0310 | 0.0133 | 0.0180 | 0.0089 | 0.01566 | 2.64E-09 | 0.048 |
| rs13021737 | -0.0493 | -0.0601 | 0.0085 | 0.0040 | -0.0150 | 0.0024 | 6.13E-09 | 1.11E-50 | 3.60E-08 |
| rs1879523 | -0.0190 | -0.0200 | 0.0068 | 0.0033 | -0.0021 | 0.0019 | 0.005242 | 1.34E-09 | 0.063 |
| rs13396935 | -0.0513 | -0.0596 | 0.0085 | 0.0042 | -0.0149 | 0.0024 | 1.46E-09 | 1.63E-46 | 5.90E-08 |
| rs1317006 | -0.0147 | -0.0214 | 0.0069 | 0.0034 | -0.0022 | 0.0020 | 0.0334 | 4.97E-10 | 0.58 |
| rs10182181 | -0.0102 | -0.0307 | 0.0064 | 0.0031 | -0.0037 | 0.0018 | 0.1103 | 8.78E-24 | 0.076 |
| rs11126052 | 0.0401 | 0.0017 | 0.0072 | 0.0042 | -0.0023 | 0.0021 | 2.85E-08 | 0.6857 | 0.63 |
| rs11126666 | 0.0005 | 0.0207 | 0.0072 | 0.0034 | 0.0003 | 0.0021 | 0.9448 | 1.33E-09 | 0.91 |
| rs1260326 | -0.0644 | -0.0124 | 0.0066 | 0.0032 | -0.0048 | 0.0019 | 1.62E-22 | 9.20E-05 | 0.016 |
| rs10171620 | 0.0399 | -0.0032 | 0.0065 | 0.0039 | -0.0104 | 0.0018 | 7.92E-10 | 0.4034 | 2.30E-08 |
| rs17030845 | -0.1191 | 0.0109 | 0.0108 | 0.0052 | 0.0271 | 0.0030 | 2.35E-28 | 0.03634 | 3.30E-17 |
| rs4671328 | 0.0328 | 0.0215 | 0.0067 | 0.0037 | -0.0018 | 0.0019 | 9.84E-07 | 6.22E-09 | 0.28 |
| rs2862874 | 0.0360 | 0.0208 | 0.0066 | 0.0038 | -0.0015 | 0.0019 | 4.83E-08 | 4.41E-08 | 0.37 |
| rs1016287 | 0.0360 | 0.0229 | 0.0074 | 0.0034 | -0.0027 | 0.0020 | 1.28E-06 | 2.25E-11 | 0.18 |
| rs6545714 | -0.0363 | -0.0180 | 0.0065 | 0.0031 | 0.0033 | 0.0019 | 2.25E-08 | 8.87E-09 | 0.14 |
| rs243019 | -0.0588 | 0.0006 | 0.0064 | 0.0031 | 0.0021 | 0.0018 | 3.37E-20 | 0.8496 | 0.56 |
| rs11688816 | -0.0193 | -0.0172 | 0.0064 | 0.0031 | -0.0028 | 0.0019 | 0.002512 | 1.89E-08 | 0.27 |
| rs2252867 | 0.0539 | 0.0023 | 0.0066 | 0.0033 | 0.0027 | 0.0019 | 3.05E-16 | 0.4852 | 0.3 |
| rs13431750 | -0.1117 | -0.0196 | 0.0204 | 0.0104 | -0.0121 | 0.0060 | 4.48E-08 | 0.05999 | 0.11 |
| rs2028150 | 0.0537 | 0.0060 | 0.0065 | 0.0038 | 0.0097 | 0.0019 | 1.31E-16 | 0.1143 | 0.00029 |
| rs11677557 | 0.0590 | -0.0010 | 0.0090 | 0.0043 | -0.0005 | 0.0025 | 5.66E-11 | 0.8171 | 0.99 |
| rs3860374 | -0.0385 | 0.0059 | 0.0065 | 0.0038 | 0.0030 | 0.0019 | 3.02E-09 | 0.1205 | 0.11 |
| rs2121279 | 0.0287 | 0.0245 | 0.0093 | 0.0044 | -0.0016 | 0.0028 | 0.002071 | 2.31E-08 | 0.43 |
| rs16841827 | -0.0822 | 0.0050 | 0.0133 | 0.0079 | -0.0040 | 0.0037 | 6.34E-10 | 0.5268 | 0.76 |
| rs7572970 | -0.0447 | -0.0066 | 0.0072 | 0.0043 | 0.0035 | 0.0020 | 6.12E-10 | 0.1248 | 0.15 |
| rs13389219 | -0.0605 | 0.0122 | 0.0065 | 0.0031 | 0.0025 | 0.0019 | 1.17E-20 | 8.96E-05 | 0.34 |
| rs3820981 | 0.0402 | -0.0102 | 0.0065 | 0.0031 | -0.0024 | 0.0019 | 5.92E-10 | 0.001063 | 0.57 |
| rs1528435 | 0.0275 | 0.0178 | 0.0066 | 0.0031 | 0.0035 | 0.0019 | 3.06E-05 | 1.20E-08 | 0.052 |
| rs7599312 | -0.0049 | -0.0220 | 0.0072 | 0.0034 | -0.0048 | 0.0021 | 0.4976 | 1.17E-10 | 0.15 |
| rs4675045 | -0.0543 | -0.0066 | 0.0081 | 0.0088 | 0.0006 | 0.0022 | 1.64E-11 | 0.4533 | 0.96 |
| rs2972144 | -0.0911 | 0.0140 | 0.0066 | 0.0031 | -0.0056 | 0.0019 | 2.19E-43 | 7.01E-06 | 0.018 |
| rs41504645 | -0.0689 | 0.0005 | 0.0097 | 0.0073 | -0.0030 | 0.0028 | 1.49E-12 | 0.9404 | 0.34 |
| rs2347101 | 0.0600 | -0.0005 | 0.0098 | 0.0050 | 0.0058 | 0.0028 | 1.09E-09 | 0.919 | 0.29 |
| rs11709077 | -0.1040 | 0.0241 | 0.0097 | 0.0046 | -0.0075 | 0.0028 | 1.26E-26 | 1.81E-07 | 0.0085 |
| rs4135247 | -0.0438 | 0.0042 | 0.0065 | 0.0038 | 0.0086 | 0.0019 | 1.51E-11 | 0.269 | 9.20E-06 |
| rs1496653 | 0.0665 | -0.0069 | 0.0079 | 0.0036 | 0.0027 | 0.0023 | 2.49E-17 | 0.05717 | 0.45 |
| rs17013433 | 0.1134 | 0.0155 | 0.0175 | 0.0090 | 0.0079 | 0.0055 | 8.85E-11 | 0.08402 | 0.37 |
| rs6804842 | -0.0175 | -0.0185 | 0.0065 | 0.0031 | -0.0032 | 0.0019 | 0.007022 | 2.48E-09 | 0.31 |
| rs6446298 | -0.0373 | -0.0151 | 0.0067 | 0.0040 | -0.0109 | 0.0020 | 2.60E-08 | 0.00016 | 3.30E-07 |
| rs2624847 | -0.0410 | -0.0016 | 0.0074 | 0.0037 | -0.0260 | 0.0022 | 3.48E-08 | 0.6657 | 6.10E-28 |
| rs891368 | -0.0378 | -0.0026 | 0.0064 | 0.0038 | -0.0119 | 0.0019 | 3.25E-09 | 0.4938 | 2.90E-08 |
| rs2365389 | -0.0097 | -0.0200 | 0.0065 | 0.0031 | -0.0074 | 0.0019 | 0.1351 | 1.63E-10 | 0.013 |
| rs2292662 | -0.0645 | 0.0038 | 0.0088 | 0.0051 | -0.0015 | 0.0026 | 2.24E-13 | 0.4562 | 0.5 |
| rs9860730 | 0.0539 | -0.0149 | 0.0070 | 0.0033 | 0.0024 | 0.0020 | 1.55E-14 | 8.18E-06 | 0.4 |
| rs3849570 | 0.0113 | 0.0188 | 0.0067 | 0.0034 | -0.0015 | 0.0020 | 0.09173 | 2.60E-08 | 0.32 |
| rs1375561 | 0.0099 | 0.0179 | 0.0067 | 0.0031 | 0.0058 | 0.0020 | 0.1396 | 9.93E-09 | 0.0075 |
| rs13078960 | -0.0118 | -0.0297 | 0.0081 | 0.0039 | -0.0038 | 0.0023 | 0.1433 | 1.74E-14 | 0.31 |
| rs11708067 | 0.0882 | -0.0152 | 0.0077 | 0.0038 | 0.0130 | 0.0021 | 5.05E-30 | 6.23E-05 | 2.50E-07 |
| rs569255 | -0.0396 | 0.0000 | 0.0064 | 0.0031 | 0.0004 | 0.0019 | 5.64E-10 | 0.9905 | 0.63 |
| rs16851483 | 0.0280 | 0.0483 | 0.0130 | 0.0077 | 0.0097 | 0.0037 | 0.03103 | 3.55E-10 | 0.004 |
| rs9828639 | 0.0383 | 0.0001 | 0.0068 | 0.0040 | 0.0008 | 0.0020 | 1.83E-08 | 0.9801 | 0.53 |
| rs10513432 | 0.0920 | 0.0102 | 0.0163 | 0.0091 | -0.0053 | 0.0048 | 1.78E-08 | 0.2623 | 0.031 |
| rs7642311 | 0.0545 | 0.0030 | 0.0096 | 0.0057 | -0.0031 | 0.0028 | 1.53E-08 | 0.5987 | 0.23 |
| rs8192675 | 0.0659 | -0.0124 | 0.0070 | 0.0034 | -0.0028 | 0.0020 | 5.77E-21 | 0.000218 | 0.11 |
| rs16860219 | 0.1222 | -0.0021 | 0.0208 | 0.0114 | 0.0249 | 0.0060 | 4.49E-09 | 0.8522 | 0.0027 |
| rs7633675 | -0.1085 | 0.0100 | 0.0068 | 0.0033 | -0.0146 | 0.0020 | 3.21E-57 | 0.002355 | 4.90E-08 |
| rs4686698 | 0.0953 | -0.0098 | 0.0172 | 0.0091 | 0.0105 | 0.0051 | 2.86E-08 | 0.2793 | 0.12 |
| rs1516725 | -0.0267 | -0.0451 | 0.0094 | 0.0046 | 0.0029 | 0.0027 | 0.004605 | 1.89E-22 | 0.38 |
| rs3887925 | 0.0539 | -0.0009 | 0.0065 | 0.0038 | 0.0023 | 0.0019 | 1.01E-16 | 0.8128 | 0.37 |
| rs6808574 | -0.0610 | -0.0018 | 0.0066 | 0.0032 | 0.0019 | 0.0019 | 2.29E-20 | 0.5823 | 0.11 |
| rs1531583 | 0.1001 | -0.0126 | 0.0157 | 0.0101 | 0.0076 | 0.0048 | 1.85E-10 | 0.2122 | 0.13 |
| rs4689381 | 0.0380 | 0.0035 | 0.0066 | 0.0031 | 0.0021 | 0.0019 | 8.37E-09 | 0.2612 | 0.42 |
| rs10937721 | 0.0850 | 0.0031 | 0.0066 | 0.0033 | 0.0000 | 0.0019 | 5.39E-38 | 0.3438 | 0.82 |
| rs7667864 | -0.0402 | -0.0044 | 0.0071 | 0.0042 | -0.0140 | 0.0020 | 1.64E-08 | 0.2948 | 2.10E-08 |
| rs13130484 | 0.0435 | 0.0401 | 0.0067 | 0.0031 | 0.0082 | 0.0019 | 8.49E-11 | 4.24E-38 | 4.90E-05 |
| rs10938397 | -0.0414 | -0.0402 | 0.0065 | 0.0031 | -0.0083 | 0.0019 | 1.80E-10 | 3.21E-38 | 5.10E-05 |
| rs17001654 | -0.0188 | -0.0306 | 0.0089 | 0.0053 | 0.0005 | 0.0026 | 0.03465 | 7.76E-09 | 0.86 |
| rs1848068 | 0.0388 | -0.0014 | 0.0067 | 0.0040 | 0.0026 | 0.0020 | 7.03E-09 | 0.7263 | 0.075 |
| rs1903002 | -0.0362 | 0.0058 | 0.0064 | 0.0040 | 0.0021 | 0.0018 | 1.45E-08 | 0.1471 | 0.44 |
| rs7678054 | -0.0394 | -0.0115 | 0.0064 | 0.0037 | 0.0183 | 0.0018 | 6.88E-10 | 0.001883 | 4.20E-17 |
| rs13107325 | 0.0460 | 0.0477 | 0.0135 | 0.0068 | -0.0162 | 0.0035 | 0.00066 | 1.83E-12 | 2.50E-05 |
| rs10516495 | -0.0418 | -0.0133 | 0.0069 | 0.0040 | 0.0079 | 0.0020 | 1.46E-09 | 0.000884 | 0.00098 |
| rs11727676 | -0.0532 | 0.0358 | 0.0109 | 0.0064 | -0.0179 | 0.0031 | 1.03E-06 | 2.55E-08 | 1.40E-07 |
| rs7669833 | -0.0572 | 0.0026 | 0.0070 | 0.0041 | -0.0027 | 0.0020 | 3.52E-16 | 0.526 | 0.32 |
| rs745805 | 0.0632 | -0.0079 | 0.0084 | 0.0051 | -0.0091 | 0.0024 | 4.52E-14 | 0.1214 | 0.00029 |
| rs9312873 | 0.0684 | 0.0068 | 0.0111 | 0.0065 | -0.0010 | 0.0031 | 7.14E-10 | 0.2955 | 0.78 |
| rs31911 | 0.0369 | 0.0056 | 0.0064 | 0.0037 | 0.0094 | 0.0019 | 7.58E-09 | 0.1301 | 4.10E-05 |
| rs12187196 | -0.0399 | -0.0005 | 0.0065 | 0.0038 | 0.0019 | 0.0019 | 7.92E-10 | 0.8953 | 0.61 |
| rs702634 | 0.0503 | -0.0122 | 0.0069 | 0.0033 | 0.0057 | 0.0020 | 3.37E-13 | 0.000258 | 0.00042 |
| rs255761 | 0.0541 | -0.0135 | 0.0080 | 0.0047 | -0.0016 | 0.0023 | 1.06E-11 | 0.004074 | 0.81 |
| rs154021 | 0.0417 | -0.0006 | 0.0074 | 0.0044 | 0.0027 | 0.0022 | 2.03E-08 | 0.8915 | 0.17 |
| rs459193 | -0.0722 | 0.0071 | 0.0073 | 0.0035 | 0.0076 | 0.0021 | 6.78E-23 | 0.04289 | 8.00E-04 |
| rs9687846 | 0.0701 | -0.0050 | 0.0081 | 0.0040 | 0.0088 | 0.0023 | 3.47E-18 | 0.2125 | 5.80E-05 |
| rs2307111 | 0.0502 | 0.0235 | 0.0065 | 0.0031 | 0.0075 | 0.0019 | 1.05E-14 | 3.77E-14 | 0.00017 |
| rs2112347 | 0.0467 | 0.0261 | 0.0067 | 0.0031 | 0.0067 | 0.0019 | 3.19E-12 | 6.19E-17 | 0.00074 |
| rs6878122 | -0.0605 | 0.0071 | 0.0070 | 0.0035 | -0.0035 | 0.0020 | 6.45E-18 | 0.04305 | 0.12 |
| rs10078815 | -0.0423 | -0.0018 | 0.0066 | 0.0044 | -0.0097 | 0.0019 | 1.43E-10 | 0.6825 | 1.10E-05 |
| rs6891076 | 0.0657 | -0.0007 | 0.0109 | 0.0053 | 0.0073 | 0.0032 | 1.60E-09 | 0.895 | 0.12 |
| rs7733501 | 0.0446 | 0.0080 | 0.0071 | 0.0042 | 0.0058 | 0.0020 | 3.75E-10 | 0.05681 | 0.01 |
| rs7729395 | 0.1472 | 0.0008 | 0.0149 | 0.0082 | 0.0079 | 0.0041 | 4.14E-23 | 0.9262 | 0.15 |
| rs17154889 | 0.0448 | 0.0117 | 0.0069 | 0.0041 | 0.0070 | 0.0020 | 8.99E-11 | 0.004322 | 0.00011 |
| rs13188193 | 0.1106 | -0.0104 | 0.0147 | 0.0080 | 0.0097 | 0.0042 | 4.52E-14 | 0.1949 | 0.051 |
| rs329122 | 0.0366 | -0.0086 | 0.0065 | 0.0031 | -0.0029 | 0.0019 | 1.72E-08 | 0.005809 | 0.22 |
| rs4585612 | -0.0414 | 0.0088 | 0.0065 | 0.0038 | -0.0002 | 0.0019 | 1.80E-10 | 0.02057 | 0.84 |
| rs9379084 | -0.0994 | 0.0165 | 0.0106 | 0.0056 | -0.0355 | 0.0029 | 5.48E-21 | 0.00303 | 5.10E-28 |
| rs11243150 | 0.0512 | 0.0002 | 0.0065 | 0.0038 | -0.0070 | 0.0019 | 3.09E-15 | 0.958 | 0.0052 |
| rs12523853 | 0.0973 | -0.0110 | 0.0169 | 0.0106 | -0.0042 | 0.0049 | 7.83E-09 | 0.2987 | 0.65 |
| rs6903706 | -0.0620 | -0.0046 | 0.0084 | 0.0039 | 0.0048 | 0.0024 | 1.34E-13 | 0.2468 | 0.23 |
| rs10946394 | -0.0566 | 0.0008 | 0.0074 | 0.0037 | 0.0027 | 0.0022 | 2.66E-14 | 0.8285 | 0.27 |
| rs9368222 | 0.1379 | -0.0095 | 0.0071 | 0.0034 | -0.0020 | 0.0021 | 1.42E-83 | 0.00546 | 0.48 |
| rs7748720 | 0.0421 | -0.0027 | 0.0076 | 0.0037 | 0.0021 | 0.0022 | 3.63E-08 | 0.4658 | 0.23 |
| rs6909558 | 0.0848 | -0.0057 | 0.0115 | 0.0058 | 0.0002 | 0.0034 | 1.80E-13 | 0.3242 | 0.88 |
| rs4077404 | -0.0570 | -0.0069 | 0.0077 | 0.0037 | 0.0044 | 0.0022 | 1.88E-13 | 0.05918 | 0.031 |
| rs3130531 | -0.0368 | -0.0037 | 0.0066 | 0.0032 | -0.0052 | 0.0019 | 2.42E-08 | 0.2488 | 0.0014 |
| rs3094682 | -0.0619 | 0.0080 | 0.0082 | 0.0054 | 0.0022 | 0.0024 | 3.47E-14 | 0.1373 | 0.53 |
| rs2855812 | 0.0430 | 0.0024 | 0.0074 | 0.0036 | 0.0019 | 0.0021 | 7.28E-09 | 0.5098 | 0.052 |
| rs805262 | 0.0379 | 0.0025 | 0.0064 | 0.0037 | 0.0058 | 0.0018 | 2.96E-09 | 0.4992 | 0.00092 |
| rs9267658 | -0.0716 | 0.0062 | 0.0091 | 0.0044 | 0.0005 | 0.0026 | 3.83E-15 | 0.162 | 0.92 |
| rs2239802 | -0.0634 | 0.0046 | 0.0077 | 0.0044 | -0.0014 | 0.0022 | 2.77E-16 | 0.2958 | 0.83 |
| rs9269081 | -0.0566 | 0.0063 | 0.0071 | 0.0042 | 0.0025 | 0.0021 | 1.87E-15 | 0.1336 | 0.12 |
| rs9271608 | -0.0847 | 0.0013 | 0.0087 | 0.0048 | 0.0094 | 0.0023 | 1.91E-22 | 0.7865 | 0.00027 |
| rs3134996 | -0.0417 | -0.0003 | 0.0069 | 0.0044 | -0.0005 | 0.0019 | 1.60E-09 | 0.9456 | 0.58 |
| rs2071479 | 0.1160 | -0.0095 | 0.0180 | 0.0101 | -0.0019 | 0.0056 | 1.19E-10 | 0.3469 | 0.25 |
| rs6457684 | -0.0369 | -0.0031 | 0.0065 | 0.0038 | -0.0004 | 0.0018 | 1.31E-08 | 0.4146 | 0.66 |
| rs205262 | 0.0037 | -0.0221 | 0.0072 | 0.0035 | -0.0006 | 0.0021 | 0.6085 | 1.75E-10 | 0.79 |
| rs6905288 | 0.0374 | -0.0110 | 0.0065 | 0.0034 | 0.0103 | 0.0018 | 8.35E-09 | 0.001298 | 1.00E-06 |
| rs6937438 | -0.0514 | 0.0036 | 0.0070 | 0.0034 | -0.0011 | 0.0020 | 2.35E-13 | 0.2879 | 0.37 |
| rs987237 | -0.0595 | -0.0440 | 0.0082 | 0.0040 | -0.0001 | 0.0024 | 3.20E-13 | 1.96E-28 | 0.54 |
| rs2817419 | 0.0313 | 0.0275 | 0.0073 | 0.0035 | 0.0030 | 0.0021 | 1.95E-05 | 3.66E-15 | 0.085 |
| rs2207139 | -0.0573 | -0.0447 | 0.0083 | 0.0040 | -0.0002 | 0.0024 | 4.29E-12 | 4.13E-29 | 0.49 |
| rs1665901 | 0.0398 | 0.0059 | 0.0068 | 0.0048 | 0.0002 | 0.0019 | 4.97E-09 | 0.219 | 0.99 |
| rs9400239 | -0.0079 | -0.0188 | 0.0070 | 0.0033 | 0.0063 | 0.0020 | 0.2601 | 1.61E-08 | 0.031 |
| rs4273712 | -0.0591 | 0.0066 | 0.0071 | 0.0035 | -0.0123 | 0.0021 | 1.03E-16 | 0.06027 | 2.30E-07 |
| rs4580892 | 0.0404 | -0.0029 | 0.0070 | 0.0034 | 0.0484 | 0.0021 | 8.46E-09 | 0.3972 | 1.50E-96 |
| rs719727 | 0.0592 | -0.0060 | 0.0074 | 0.0036 | -0.0345 | 0.0021 | 1.67E-15 | 0.0979 | 1.90E-53 |
| rs6901126 | -0.0356 | -0.0093 | 0.0065 | 0.0031 | -0.0026 | 0.0019 | 4.16E-08 | 0.002358 | 0.14 |
| rs474513 | 0.0399 | -0.0116 | 0.0064 | 0.0038 | 0.0009 | 0.0018 | 4.18E-10 | 0.002268 | 0.59 |
| rs13191362 | 0.0235 | 0.0277 | 0.0098 | 0.0048 | 0.0013 | 0.0028 | 0.01695 | 7.34E-09 | 0.48 |
| rs17630640 | 0.0537 | -0.0139 | 0.0096 | 0.0058 | 0.0312 | 0.0027 | 2.48E-08 | 0.01655 | 5.20E-25 |
| rs17168486 | 0.0672 | -0.0047 | 0.0083 | 0.0041 | -0.0009 | 0.0024 | 4.50E-16 | 0.2567 | 0.64 |
| rs2215383 | -0.0641 | 0.0042 | 0.0064 | 0.0031 | 0.0085 | 0.0018 | 1.06E-23 | 0.177 | 2.00E-04 |
| rs6948511 | 0.0641 | 0.0007 | 0.0112 | 0.0055 | 0.0098 | 0.0033 | 1.06E-08 | 0.8965 | 0.0091 |
| rs10225433 | -0.0767 | 0.0040 | 0.0138 | 0.0107 | 0.0018 | 0.0038 | 2.86E-08 | 0.7089 | 0.63 |
| rs849135 | -0.0906 | 0.0121 | 0.0064 | 0.0031 | 0.0063 | 0.0018 | 1.13E-45 | 7.12E-05 | 0.0057 |
| rs2041547 | -0.0408 | -0.0008 | 0.0064 | 0.0031 | -0.0006 | 0.0018 | 1.68E-10 | 0.8048 | 0.9 |
| rs2971669 | 0.0602 | 0.0032 | 0.0080 | 0.0040 | -0.0037 | 0.0022 | 3.87E-14 | 0.4225 | 0.15 |
| rs1167827 | -0.0100 | -0.0202 | 0.0065 | 0.0033 | -0.0034 | 0.0018 | 0.1234 | 6.33E-10 | 0.082 |
| rs2245368 | -0.0319 | -0.0317 | 0.0086 | 0.0057 | -0.0084 | 0.0024 | 0.000203 | 3.19E-08 | 0.0018 |
| rs11496066 | 0.0508 | 0.0002 | 0.0083 | 0.0048 | -0.0007 | 0.0023 | 8.17E-10 | 0.9668 | 0.88 |
| rs1447059 | -0.0522 | 0.0085 | 0.0066 | 0.0032 | 0.0054 | 0.0019 | 2.50E-15 | 0.008232 | 0.004 |
| rs6955948 | 0.0395 | 0.0064 | 0.0070 | 0.0043 | 0.0049 | 0.0020 | 1.79E-08 | 0.1367 | 0.015 |
| rs3802120 | -0.0582 | -0.0124 | 0.0067 | 0.0040 | 0.0036 | 0.0019 | 3.78E-18 | 0.001935 | 0.36 |
| rs2921077 | 0.0373 | 0.0094 | 0.0065 | 0.0038 | 0.0363 | 0.0019 | 9.14E-09 | 0.01337 | 1.30E-64 |
| rs6990912 | 0.0426 | 0.0076 | 0.0067 | 0.0040 | 0.0268 | 0.0019 | 2.05E-10 | 0.05743 | 8.50E-30 |
| rs17662402 | 0.0835 | 0.0023 | 0.0142 | 0.0072 | 0.0123 | 0.0039 | 4.52E-09 | 0.7531 | 0.012 |
| rs17689007 | -0.0468 | -0.0132 | 0.0065 | 0.0038 | -0.0242 | 0.0019 | 5.62E-13 | 0.000513 | 2.00E-31 |
| rs2001433 | -0.0442 | -0.0082 | 0.0065 | 0.0038 | -0.0362 | 0.0019 | 9.84E-12 | 0.03094 | 2.40E-60 |
| rs2244648 | -0.0363 | -0.0071 | 0.0065 | 0.0040 | -0.0376 | 0.0019 | 2.25E-08 | 0.0759 | 1.30E-66 |
| rs7819706 | 0.0669 | 0.0002 | 0.0101 | 0.0048 | 0.0065 | 0.0029 | 2.82E-11 | 0.959 | 0.07 |
| rs755162 | 0.0402 | -0.0028 | 0.0070 | 0.0042 | -0.0022 | 0.0020 | 1.00E-08 | 0.505 | 0.39 |
| rs508419 | -0.0811 | 0.0028 | 0.0075 | 0.0044 | -0.0008 | 0.0022 | 5.43E-27 | 0.5245 | 0.49 |
| rs2241896 | 0.0451 | -0.0005 | 0.0070 | 0.0041 | 0.0012 | 0.0019 | 1.28E-10 | 0.9029 | 0.66 |
| rs17603828 | 0.0800 | 0.0040 | 0.0133 | 0.0082 | 0.0064 | 0.0037 | 1.78E-09 | 0.6257 | 0.14 |
| rs17405819 | 0.0126 | 0.0224 | 0.0069 | 0.0033 | 0.0032 | 0.0020 | 0.06825 | 2.07E-11 | 0.29 |
| rs2033732 | -0.0136 | -0.0192 | 0.0073 | 0.0035 | -0.0008 | 0.0021 | 0.06351 | 4.89E-08 | 0.41 |
| rs13257021 | 0.0485 | 0.0045 | 0.0064 | 0.0037 | -0.0017 | 0.0019 | 3.11E-14 | 0.2239 | 0.5 |
| rs3808415 | -0.0349 | -0.0107 | 0.0064 | 0.0037 | -0.0098 | 0.0019 | 4.65E-08 | 0.003829 | 5.20E-06 |
| rs3802177 | -0.1077 | 0.0097 | 0.0069 | 0.0034 | -0.0036 | 0.0020 | 9.16E-55 | 0.004228 | 0.27 |
| rs16889471 | 0.0448 | -0.0028 | 0.0077 | 0.0038 | 0.0020 | 0.0022 | 7.38E-09 | 0.4615 | 0.35 |
| rs1561927 | -0.0420 | -0.0035 | 0.0072 | 0.0034 | -0.0006 | 0.0021 | 6.12E-09 | 0.3147 | 0.87 |
| rs4977218 | -0.0505 | 0.0073 | 0.0069 | 0.0040 | -0.0084 | 0.0019 | 2.71E-13 | 0.068 | 1.80E-06 |
| rs2272662 | -0.0402 | 0.0075 | 0.0072 | 0.0061 | -0.0047 | 0.0019 | 2.63E-08 | 0.2189 | 0.035 |
| rs997258 | 0.0376 | 0.0026 | 0.0065 | 0.0038 | 0.0000 | 0.0019 | 6.95E-09 | 0.4938 | 0.84 |
| rs10974438 | -0.0514 | -0.0005 | 0.0067 | 0.0033 | 0.0019 | 0.0019 | 1.71E-14 | 0.8826 | 0.12 |
| rs4740619 | 0.0146 | 0.0179 | 0.0064 | 0.0031 | 0.0036 | 0.0019 | 0.02226 | 4.56E-09 | 0.055 |
| rs7467618 | -0.0393 | 0.0038 | 0.0065 | 0.0038 | 0.0030 | 0.0019 | 1.41E-09 | 0.3173 | 0.15 |
| rs1063192 | 0.0575 | 0.0002 | 0.0065 | 0.0031 | 0.0011 | 0.0019 | 8.17E-19 | 0.9574 | 0.44 |
| rs17694555 | -0.0614 | 0.0108 | 0.0110 | 0.0053 | 0.0007 | 0.0033 | 2.34E-08 | 0.04213 | 0.77 |
| rs10811661 | 0.1597 | -0.0043 | 0.0086 | 0.0042 | -0.0040 | 0.0024 | 3.16E-77 | 0.3024 | 0.062 |
| rs7018475 | -0.1043 | 0.0131 | 0.0072 | 0.0054 | -0.0052 | 0.0021 | 3.03E-47 | 0.01514 | 0.012 |
| rs1412234 | -0.0393 | -0.0235 | 0.0068 | 0.0041 | 0.0004 | 0.0020 | 7.71E-09 | 9.94E-09 | 0.77 |
| rs10968576 | -0.0370 | -0.0249 | 0.0068 | 0.0033 | 0.0000 | 0.0020 | 5.43E-08 | 6.61E-14 | 0.96 |
| rs17791513 | 0.1016 | -0.0024 | 0.0132 | 0.0060 | 0.0108 | 0.0039 | 1.35E-14 | 0.695 | 0.023 |
| rs2796441 | -0.0674 | 0.0067 | 0.0065 | 0.0033 | 0.0025 | 0.0019 | 2.97E-25 | 0.04045 | 0.02 |
| rs12004367 | -0.0374 | 0.0028 | 0.0068 | 0.0040 | -0.0187 | 0.0020 | 3.90E-08 | 0.4839 | 8.40E-19 |
| rs6477694 | -0.0196 | -0.0174 | 0.0066 | 0.0031 | -0.0033 | 0.0019 | 0.002965 | 2.67E-08 | 0.15 |
| rs1928295 | 0.0121 | 0.0188 | 0.0064 | 0.0031 | 0.0014 | 0.0019 | 0.05816 | 7.91E-10 | 0.3 |
| rs505922 | -0.0473 | -0.0009 | 0.0068 | 0.0033 | 0.0175 | 0.0020 | 3.65E-12 | 0.7921 | 4.40E-13 |
| rs3812547 | 0.0495 | 0.0020 | 0.0070 | 0.0052 | 0.0005 | 0.0020 | 1.71E-12 | 0.7005 | 0.47 |
| rs11257655 | 0.0859 | -0.0077 | 0.0077 | 0.0038 | -0.0038 | 0.0023 | 1.46E-28 | 0.04285 | 0.077 |
| rs2616055 | 0.0799 | -0.0003 | 0.0137 | 0.0084 | -0.0050 | 0.0041 | 5.70E-09 | 0.9715 | 0.17 |
| rs177045 | -0.0500 | -0.0046 | 0.0069 | 0.0042 | 0.0043 | 0.0020 | 4.64E-13 | 0.2734 | 0.34 |
| rs2642588 | -0.0510 | 0.0001 | 0.0070 | 0.0042 | 0.0012 | 0.0020 | 3.59E-13 | 0.981 | 0.32 |
| rs11591689 | 0.0486 | -0.0047 | 0.0077 | 0.0044 | 0.0019 | 0.0022 | 3.55E-10 | 0.2854 | 0.56 |
| rs703972 | -0.0698 | -0.0045 | 0.0065 | 0.0037 | -0.0025 | 0.0018 | 5.77E-27 | 0.2239 | 0.14 |
| rs10509406 | 0.0564 | 0.0115 | 0.0086 | 0.0052 | 0.0033 | 0.0025 | 5.06E-11 | 0.027 | 0.22 |
| rs1574190 | 0.0476 | -0.0024 | 0.0067 | 0.0038 | -0.0008 | 0.0019 | 1.22E-12 | 0.5277 | 0.75 |
| rs7899106 | -0.0113 | -0.0395 | 0.0144 | 0.0071 | -0.0049 | 0.0042 | 0.4342 | 2.96E-08 | 0.25 |
| rs7903767 | -0.0550 | -0.0022 | 0.0064 | 0.0037 | -0.0061 | 0.0018 | 7.21E-18 | 0.5521 | 0.0037 |
| rs10882099 | 0.1095 | -0.0107 | 0.0065 | 0.0051 | 0.0015 | 0.0019 | 7.73E-64 | 0.03682 | 0.76 |
| rs11187150 | 0.0633 | -0.0034 | 0.0107 | 0.0104 | 0.0004 | 0.0031 | 3.08E-09 | 0.7455 | 0.85 |
| rs11187152 | -0.0695 | 0.0179 | 0.0124 | 0.0089 | 0.0013 | 0.0035 | 1.85E-08 | 0.0443 | 0.59 |
| rs17094222 | -0.0263 | -0.0249 | 0.0079 | 0.0038 | -0.0019 | 0.0023 | 0.000811 | 5.94E-11 | 0.95 |
| rs11191560 | 0.0018 | -0.0308 | 0.0115 | 0.0053 | -0.0165 | 0.0035 | 0.8758 | 8.45E-09 | 0.00012 |
| rs545572 | -0.0762 | -0.0004 | 0.0068 | 0.0048 | -0.0054 | 0.0019 | 4.24E-29 | 0.9336 | 0.011 |
| rs1885283 | 0.0727 | -0.0035 | 0.0102 | 0.0050 | -0.0057 | 0.0028 | 8.17E-13 | 0.4766 | 0.23 |
| rs2009075 | 0.1707 | -0.0009 | 0.0216 | 0.0131 | -0.0120 | 0.0064 | 2.49E-15 | 0.9452 | 0.32 |
| rs17746916 | 0.1491 | -0.0210 | 0.0133 | 0.0095 | 0.0129 | 0.0038 | 3.53E-29 | 0.02707 | 0.00044 |
| rs7901695 | -0.2847 | 0.0213 | 0.0069 | 0.0034 | -0.0105 | 0.0020 | 5.13E-371 | 2.66E-10 | 9.90E-09 |
| rs7903146 | 0.3142 | -0.0234 | 0.0070 | 0.0034 | 0.0113 | 0.0020 | 4.31E-438 | 1.11E-11 | 8.10E-09 |
| rs12266632 | -0.2255 | 0.0185 | 0.0128 | 0.0068 | -0.0022 | 0.0036 | 9.57E-70 | 0.006341 | 0.32 |
| rs7896811 | -0.1465 | 0.0046 | 0.0091 | 0.0043 | -0.0078 | 0.0026 | 3.37E-58 | 0.282 | 0.0053 |
| rs12354626 | -0.1090 | -0.0005 | 0.0192 | 0.0116 | -0.0154 | 0.0052 | 1.28E-08 | 0.9654 | 0.0075 |
| rs17685538 | 0.0954 | -0.0140 | 0.0107 | 0.0055 | 0.0012 | 0.0030 | 4.15E-19 | 0.01133 | 0.24 |
| rs10885414 | 0.0949 | -0.0087 | 0.0070 | 0.0037 | 0.0067 | 0.0020 | 1.07E-41 | 0.0192 | 0.005 |
| rs10749128 | -0.0460 | 0.0007 | 0.0074 | 0.0046 | -0.0049 | 0.0021 | 6.10E-10 | 0.879 | 0.0025 |
| rs11196236 | -0.0739 | -0.0004 | 0.0079 | 0.0048 | 0.0038 | 0.0023 | 4.92E-21 | 0.9336 | 0.11 |
| rs290483 | 0.0701 | -0.0038 | 0.0067 | 0.0033 | 0.0062 | 0.0019 | 1.30E-25 | 0.246 | 0.01 |
| rs2280141 | 0.0489 | 0.0008 | 0.0064 | 0.0038 | 0.0040 | 0.0018 | 1.91E-14 | 0.8333 | 0.1 |
| rs10770141 | 0.0658 | 0.0105 | 0.0068 | 0.0044 | 0.0040 | 0.0019 | 4.11E-22 | 0.01702 | 0.12 |
| rs11023461 | -0.0751 | 0.0025 | 0.0129 | 0.0073 | 0.0010 | 0.0036 | 5.49E-09 | 0.7293 | 0.76 |
| rs231361 | 0.0607 | 0.0013 | 0.0074 | 0.0036 | -0.0040 | 0.0021 | 3.21E-16 | 0.7102 | 0.13 |
| rs7480855 | 0.1007 | -0.0002 | 0.0133 | 0.0102 | -0.0025 | 0.0037 | 3.65E-14 | 0.9861 | 0.57 |
| rs2237895 | -0.0892 | 0.0070 | 0.0066 | 0.0052 | -0.0005 | 0.0019 | 1.15E-41 | 0.1765 | 0.8 |
| rs4256980 | -0.0268 | -0.0209 | 0.0067 | 0.0031 | -0.0053 | 0.0019 | 6.35E-05 | 2.90E-11 | 0.027 |
| rs5215 | -0.0706 | 0.0138 | 0.0066 | 0.0031 | -0.0014 | 0.0019 | 9.85E-27 | 1.13E-05 | 0.55 |
| rs11030104 | 0.0193 | 0.0414 | 0.0080 | 0.0038 | -0.0097 | 0.0023 | 0.01529 | 5.56E-28 | 0.00072 |
| rs11030107 | -0.0316 | -0.0297 | 0.0073 | 0.0035 | -0.0113 | 0.0021 | 1.62E-05 | 3.15E-17 | 1.80E-09 |
| rs2767036 | -0.0389 | -0.0091 | 0.0070 | 0.0047 | 0.0016 | 0.0020 | 2.94E-08 | 0.05285 | 0.31 |
| rs2176598 | 0.0457 | 0.0198 | 0.0073 | 0.0036 | -0.0025 | 0.0021 | 4.51E-10 | 2.97E-08 | 0.21 |
| rs1061810 | 0.0502 | 0.0153 | 0.0070 | 0.0042 | -0.0001 | 0.0020 | 8.31E-13 | 0.00027 | 0.79 |
| rs10838524 | 0.0350 | 0.0012 | 0.0064 | 0.0034 | -0.0026 | 0.0019 | 4.25E-08 | 0.7303 | 0.16 |
| rs7124681 | 0.0362 | 0.0259 | 0.0065 | 0.0031 | -0.0005 | 0.0019 | 2.45E-08 | 1.16E-16 | 0.93 |
| rs3817334 | 0.0344 | 0.0262 | 0.0065 | 0.0031 | -0.0006 | 0.0019 | 1.16E-07 | 5.15E-17 | 0.99 |
| rs947791 | 0.0606 | 0.0135 | 0.0081 | 0.0048 | 0.0258 | 0.0022 | 5.62E-14 | 0.004916 | 3.20E-23 |
| rs3918298 | -0.1349 | -0.0239 | 0.0209 | 0.0209 | -0.0257 | 0.0058 | 1.18E-10 | 0.2528 | 0.00011 |
| rs1552224 | 0.0981 | -0.0184 | 0.0087 | 0.0042 | 0.0082 | 0.0025 | 1.50E-29 | 9.40E-06 | 0.013 |
| rs10830963 | -0.1010 | -0.0062 | 0.0071 | 0.0036 | -0.0086 | 0.0021 | 1.12E-45 | 0.08769 | 0.00027 |
| rs9971402 | 0.0528 | 0.0027 | 0.0091 | 0.0054 | 0.0018 | 0.0027 | 6.78E-09 | 0.6171 | 0.29 |
| rs10444213 | 0.0419 | 0.0090 | 0.0065 | 0.0042 | 0.0027 | 0.0019 | 1.09E-10 | 0.03212 | 0.22 |
| rs12286929 | -0.0234 | -0.0217 | 0.0064 | 0.0031 | 0.0287 | 0.0018 | 0.000249 | 1.31E-12 | 1.00E-49 |
| rs7933438 | -0.0580 | 0.0050 | 0.0093 | 0.0052 | 0.0012 | 0.0026 | 4.84E-10 | 0.3363 | 0.93 |
| rs10750397 | 0.0394 | -0.0034 | 0.0071 | 0.0041 | -0.0037 | 0.0021 | 3.13E-08 | 0.407 | 0.023 |
| rs11819995 | 0.0498 | 0.0037 | 0.0077 | 0.0046 | 0.0038 | 0.0022 | 1.30E-10 | 0.4212 | 0.15 |
| rs10848958 | -0.0451 | 0.0044 | 0.0083 | 0.0047 | 0.0013 | 0.0023 | 4.97E-08 | 0.3492 | 0.27 |
| rs11063069 | -0.0560 | 0.0100 | 0.0079 | 0.0041 | -0.0026 | 0.0023 | 9.94E-13 | 0.0155 | 0.53 |
| rs3217795 | 0.1065 | -0.0003 | 0.0119 | 0.0079 | -0.0008 | 0.0033 | 4.56E-19 | 0.9697 | 0.95 |
| rs12299509 | -0.0496 | 0.0058 | 0.0068 | 0.0034 | -0.0066 | 0.0018 | 3.15E-13 | 0.08817 | 0.0019 |
| rs2066827 | -0.0473 | 0.0020 | 0.0081 | 0.0063 | -0.0040 | 0.0022 | 4.44E-09 | 0.7509 | 0.065 |
| rs1872992 | -0.0457 | 0.0093 | 0.0072 | 0.0035 | -0.0276 | 0.0022 | 2.52E-10 | 0.008036 | 4.80E-28 |
| rs2052673 | -0.0557 | -0.0011 | 0.0089 | 0.0051 | -0.0053 | 0.0025 | 3.88E-10 | 0.8292 | 0.013 |
| rs7969720 | -0.0383 | 0.0019 | 0.0069 | 0.0042 | 0.0022 | 0.0020 | 2.98E-08 | 0.651 | 0.29 |
| rs3751239 | 0.0749 | -0.0031 | 0.0081 | 0.0047 | 0.0041 | 0.0023 | 1.54E-20 | 0.5095 | 0.11 |
| rs7138803 | 0.0342 | 0.0315 | 0.0066 | 0.0031 | 0.0116 | 0.0019 | 2.16E-07 | 8.15E-24 | 9.20E-07 |
| rs2612069 | -0.1034 | 0.0021 | 0.0109 | 0.0064 | -0.0025 | 0.0031 | 2.19E-21 | 0.7428 | 0.35 |
| rs7968682 | 0.0538 | -0.0087 | 0.0064 | 0.0031 | 0.0139 | 0.0019 | 3.65E-17 | 0.005209 | 4.10E-11 |
| rs1796330 | -0.0487 | 0.0008 | 0.0065 | 0.0037 | -0.0035 | 0.0019 | 6.28E-14 | 0.8288 | 0.22 |
| rs11108094 | 0.0714 | 0.0026 | 0.0128 | 0.0078 | -0.0084 | 0.0036 | 2.28E-08 | 0.7389 | 0.0023 |
| rs3764002 | -0.0511 | -0.0049 | 0.0073 | 0.0038 | -0.0056 | 0.0021 | 3.12E-12 | 0.1949 | 0.28 |
| rs2251468 | -0.0474 | 0.0001 | 0.0067 | 0.0032 | -0.0001 | 0.0020 | 1.51E-12 | 0.9733 | 0.6 |
| rs7957197 | -0.0672 | -0.0011 | 0.0080 | 0.0039 | -0.0032 | 0.0023 | 3.04E-17 | 0.7684 | 0.12 |
| rs208302 | 0.0414 | 0.0062 | 0.0075 | 0.0046 | -0.0035 | 0.0022 | 3.98E-08 | 0.1777 | 0.18 |
| rs11057405 | 0.0306 | -0.0307 | 0.0109 | 0.0055 | 0.0009 | 0.0030 | 0.004952 | 2.02E-08 | 0.85 |
| rs7132277 | -0.0475 | 0.0032 | 0.0082 | 0.0041 | -0.0177 | 0.0024 | 6.02E-09 | 0.4374 | 2.90E-10 |
| rs1882297 | 0.0474 | 0.0041 | 0.0070 | 0.0035 | 0.0019 | 0.0020 | 1.41E-11 | 0.234 | 0.51 |
| rs12865499 | -0.0423 | 0.0105 | 0.0075 | 0.0044 | 0.0017 | 0.0022 | 2.01E-08 | 0.01702 | 0.7 |
| rs9579083 | 0.0273 | 0.0295 | 0.0081 | 0.0047 | -0.0010 | 0.0024 | 0.000709 | 3.46E-10 | 0.33 |
| rs576674 | -0.0538 | 0.0082 | 0.0086 | 0.0044 | 0.0011 | 0.0025 | 3.70E-10 | 0.06097 | 0.22 |
| rs9316500 | 0.0387 | -0.0027 | 0.0070 | 0.0041 | 0.0137 | 0.0020 | 3.45E-08 | 0.5102 | 1.20E-11 |
| rs12429545 | 0.0384 | 0.0334 | 0.0101 | 0.0047 | 0.0079 | 0.0028 | 0.000133 | 1.09E-12 | 0.0016 |
| rs12583517 | -0.0423 | -0.0141 | 0.0077 | 0.0044 | 0.0006 | 0.0022 | 4.78E-08 | 0.001353 | 0.78 |
| rs9563615 | 0.0408 | 0.0103 | 0.0072 | 0.0042 | 0.0061 | 0.0020 | 1.63E-08 | 0.01419 | 0.0028 |
| rs1359790 | -0.0817 | 0.0030 | 0.0071 | 0.0034 | -0.0093 | 0.0020 | 1.76E-30 | 0.3814 | 2.00E-04 |
| rs7325671 | 0.0545 | 0.0030 | 0.0096 | 0.0058 | -0.0038 | 0.0028 | 1.53E-08 | 0.605 | 0.039 |
| rs10132280 | -0.0324 | -0.0230 | 0.0070 | 0.0034 | 0.0001 | 0.0020 | 3.86E-06 | 1.14E-11 | 0.55 |
| rs12885454 | -0.0215 | -0.0207 | 0.0067 | 0.0033 | -0.0054 | 0.0019 | 0.001334 | 1.94E-10 | 0.0096 |
| rs11847697 | 0.0464 | 0.0492 | 0.0160 | 0.0084 | 0.0076 | 0.0045 | 0.003774 | 3.99E-09 | 0.0086 |
| rs17522122 | 0.0356 | 0.0152 | 0.0064 | 0.0033 | -0.0020 | 0.0019 | 2.49E-08 | 3.15E-06 | 0.17 |
| rs7141420 | 0.0317 | 0.0235 | 0.0064 | 0.0031 | 0.0058 | 0.0019 | 6.93E-07 | 1.23E-14 | 0.0091 |
| rs11625769 | 0.0303 | 0.0190 | 0.0068 | 0.0034 | 0.0030 | 0.0020 | 8.50E-06 | 1.70E-08 | 0.66 |
| rs17109221 | 0.0556 | 0.0252 | 0.0077 | 0.0046 | 0.0068 | 0.0022 | 7.18E-13 | 4.30E-08 | 0.12 |
| rs7143394 | -0.0364 | -0.0068 | 0.0064 | 0.0037 | 0.0034 | 0.0019 | 1.20E-08 | 0.06609 | 0.25 |
| rs3783394 | -0.0380 | -0.0157 | 0.0067 | 0.0038 | -0.0423 | 0.0020 | 1.42E-08 | 3.60E-05 | 9.50E-80 |
| rs8032939 | -0.0418 | -0.0064 | 0.0074 | 0.0044 | 0.0044 | 0.0022 | 1.88E-08 | 0.1458 | 0.052 |
| rs2289739 | 0.0469 | 0.0052 | 0.0067 | 0.0042 | -0.0035 | 0.0020 | 2.58E-12 | 0.2157 | 0.031 |
| rs3736485 | 0.0213 | 0.0176 | 0.0065 | 0.0031 | 0.0033 | 0.0019 | 0.001034 | 7.41E-09 | 0.2 |
| rs2456530 | 0.0540 | 0.0114 | 0.0096 | 0.0058 | -0.0024 | 0.0027 | 2.07E-08 | 0.04935 | 0.5 |
| rs8037894 | -0.0467 | -0.0041 | 0.0065 | 0.0038 | 0.0035 | 0.0019 | 6.29E-13 | 0.2806 | 0.027 |
| rs7178762 | -0.0387 | -0.0058 | 0.0064 | 0.0037 | -0.0093 | 0.0019 | 1.37E-09 | 0.117 | 1.00E-05 |
| rs16951275 | 0.0340 | 0.0311 | 0.0077 | 0.0037 | 0.0003 | 0.0022 | 1.14E-05 | 1.91E-17 | 0.98 |
| rs4776970 | 0.0378 | 0.0244 | 0.0067 | 0.0031 | -0.0004 | 0.0019 | 1.69E-08 | 8.87E-15 | 0.37 |
| rs13737 | -0.0481 | 0.0019 | 0.0075 | 0.0046 | 0.0049 | 0.0022 | 1.77E-10 | 0.6796 | 0.12 |
| rs11630770 | 0.0538 | 0.0086 | 0.0090 | 0.0052 | 0.0015 | 0.0026 | 2.30E-09 | 0.09816 | 0.67 |
| rs12910361 | -0.0814 | -0.0083 | 0.0070 | 0.0041 | 0.0038 | 0.0021 | 3.95E-31 | 0.04293 | 0.054 |
| rs4932265 | 0.0653 | -0.0030 | 0.0072 | 0.0043 | 0.0030 | 0.0021 | 1.59E-19 | 0.4854 | 0.2 |
| rs12910825 | -0.0525 | 0.0010 | 0.0067 | 0.0032 | -0.0034 | 0.0019 | 4.70E-15 | 0.761 | 0.19 |
| rs6600191 | 0.0587 | 0.0027 | 0.0085 | 0.0051 | -0.0020 | 0.0024 | 4.47E-12 | 0.5965 | 0.41 |
| rs758747 | 0.0297 | 0.0225 | 0.0072 | 0.0037 | 0.0037 | 0.0021 | 3.94E-05 | 7.47E-10 | 0.071 |
| rs879620 | 0.0038 | 0.0244 | 0.0067 | 0.0040 | 0.0147 | 0.0019 | 0.5707 | 1.06E-09 | 2.00E-11 |
| rs12446632 | -0.0392 | -0.0403 | 0.0093 | 0.0046 | -0.0148 | 0.0026 | 2.59E-05 | 1.48E-18 | 7.50E-07 |
| rs11074422 | -0.0041 | -0.0188 | 0.0067 | 0.0034 | 0.0002 | 0.0020 | 0.5406 | 3.34E-08 | 0.74 |
| rs11074446 | 0.0363 | 0.0256 | 0.0094 | 0.0045 | 0.0051 | 0.0028 | 0.000117 | 1.31E-08 | 0.11 |
| rs2650492 | 0.0060 | 0.0207 | 0.0070 | 0.0035 | -0.0042 | 0.0020 | 0.3924 | 1.92E-09 | 0.037 |
| rs3888190 | 0.0291 | 0.0309 | 0.0065 | 0.0031 | -0.0028 | 0.0019 | 7.37E-06 | 3.14E-23 | 0.19 |
| rs12325539 | -0.0410 | -0.0173 | 0.0065 | 0.0038 | 0.0057 | 0.0019 | 2.69E-10 | 5.30E-06 | 2.00E-04 |
| rs9925964 | 0.0219 | 0.0192 | 0.0067 | 0.0031 | -0.0094 | 0.0019 | 0.001082 | 8.11E-10 | 4.00E-06 |
| rs1477199 | -0.0226 | -0.0242 | 0.0091 | 0.0044 | -0.0013 | 0.0027 | 0.0131 | 4.56E-08 | 0.78 |
| rs7203521 | 0.0424 | 0.0326 | 0.0066 | 0.0032 | 0.0051 | 0.0019 | 1.29E-10 | 3.46E-24 | 0.025 |
| rs1421085 | -0.1218 | -0.0813 | 0.0065 | 0.0031 | -0.0195 | 0.0019 | 1.52E-78 | 8.83E-151 | 1.00E-19 |
| rs1558902 | 0.1218 | 0.0818 | 0.0065 | 0.0031 | 0.0195 | 0.0019 | 1.52E-78 | 7.51E-153 | 1.20E-19 |
| rs11075986 | 0.0590 | 0.0423 | 0.0120 | 0.0060 | 0.0060 | 0.0034 | 9.58E-07 | 1.23E-12 | 0.23 |
| rs16952522 | -0.0884 | -0.0547 | 0.0161 | 0.0108 | -0.0174 | 0.0049 | 4.20E-08 | 4.53E-07 | 0.011 |
| rs6499646 | 0.0656 | 0.0410 | 0.0119 | 0.0077 | 0.0063 | 0.0034 | 3.89E-08 | 1.01E-07 | 0.19 |
| rs17218700 | -0.0615 | -0.0218 | 0.0104 | 0.0048 | -0.0033 | 0.0028 | 2.97E-09 | 6.33E-06 | 0.087 |
| rs6499653 | 0.0430 | 0.0269 | 0.0073 | 0.0037 | 0.0103 | 0.0022 | 4.44E-09 | 2.32E-13 | 2.00E-05 |
| rs244418 | -0.0429 | -0.0149 | 0.0065 | 0.0037 | 0.0026 | 0.0019 | 3.88E-11 | 5.65E-05 | 0.054 |
| rs889512 | 0.1111 | -0.0037 | 0.0107 | 0.0064 | 0.0071 | 0.0031 | 2.40E-25 | 0.5632 | 0.032 |
| rs2925979 | 0.0546 | -0.0012 | 0.0070 | 0.0033 | -0.0044 | 0.0020 | 7.07E-15 | 0.721 | 0.028 |
| rs3826482 | 0.0367 | -0.0044 | 0.0066 | 0.0035 | 0.0023 | 0.0019 | 2.64E-08 | 0.2149 | 0.12 |
| rs1377807 | 0.0524 | 0.0045 | 0.0069 | 0.0041 | 0.0111 | 0.0020 | 3.38E-14 | 0.2724 | 1.60E-07 |
| rs1000940 | -0.0218 | -0.0192 | 0.0069 | 0.0034 | -0.0066 | 0.0020 | 0.001607 | 1.28E-08 | 0.0015 |
| rs7219033 | 0.0387 | 0.0111 | 0.0068 | 0.0040 | 0.0093 | 0.0020 | 1.30E-08 | 0.00552 | 2.90E-05 |
| rs4925109 | 0.0476 | -0.0127 | 0.0069 | 0.0040 | -0.0088 | 0.0020 | 5.65E-12 | 0.001498 | 0.00049 |
| rs2107133 | 0.0643 | -0.0016 | 0.0097 | 0.0059 | 0.0013 | 0.0027 | 4.02E-11 | 0.7862 | 0.62 |
| rs11651755 | -0.0706 | 0.0102 | 0.0065 | 0.0047 | -0.0033 | 0.0019 | 1.51E-27 | 0.02832 | 0.17 |
| rs6963 | 0.0468 | 0.0040 | 0.0071 | 0.0042 | 0.0134 | 0.0020 | 4.92E-11 | 0.3409 | 1.10E-09 |
| rs1962412 | -0.0521 | -0.0041 | 0.0070 | 0.0047 | 0.0000 | 0.0020 | 1.11E-13 | 0.383 | 0.96 |
| rs7216064 | 0.0507 | 0.0109 | 0.0080 | 0.0046 | -0.0091 | 0.0023 | 1.87E-10 | 0.01781 | 0.00047 |
| rs12940622 | -0.0163 | -0.0182 | 0.0065 | 0.0031 | -0.0033 | 0.0019 | 0.01204 | 2.49E-09 | 0.18 |
| rs7240767 | -0.0372 | 0.0033 | 0.0066 | 0.0040 | 0.0019 | 0.0019 | 1.71E-08 | 0.4094 | 0.42 |
| rs1808579 | -0.0251 | -0.0167 | 0.0064 | 0.0031 | 0.0057 | 0.0019 | 8.50E-05 | 4.17E-08 | 0.01 |
| rs1431841 | 0.0429 | 0.0132 | 0.0077 | 0.0046 | 0.0066 | 0.0023 | 3.08E-08 | 0.00411 | 0.052 |
| rs9957320 | -0.0487 | -0.0226 | 0.0085 | 0.0051 | -0.0076 | 0.0025 | 9.34E-09 | 9.36E-06 | 0.025 |
| rs7243357 | 0.0432 | 0.0217 | 0.0084 | 0.0040 | 0.0073 | 0.0024 | 2.50E-07 | 3.86E-08 | 0.021 |
| rs9945063 | 0.0221 | 0.0217 | 0.0074 | 0.0038 | 0.0034 | 0.0022 | 0.00295 | 1.35E-08 | 0.15 |
| rs6567160 | -0.0545 | -0.0556 | 0.0075 | 0.0036 | -0.0147 | 0.0022 | 4.85E-13 | 3.93E-53 | 2.90E-07 |
| rs8089364 | -0.0535 | -0.0510 | 0.0072 | 0.0035 | -0.0163 | 0.0021 | 1.31E-13 | 3.05E-49 | 1.30E-09 |
| rs9947301 | -0.0260 | -0.0377 | 0.0121 | 0.0057 | -0.0128 | 0.0037 | 0.0323 | 3.70E-11 | 0.0029 |
| rs9956279 | 0.0368 | 0.0348 | 0.0069 | 0.0033 | 0.0128 | 0.0020 | 1.01E-07 | 2.62E-25 | 2.00E-08 |
| rs12962523 | -0.0147 | -0.0342 | 0.0080 | 0.0051 | -0.0105 | 0.0023 | 0.0647 | 2.00E-11 | 9.30E-05 |
| rs17066842 | -0.0895 | -0.0626 | 0.0165 | 0.0083 | -0.0004 | 0.0048 | 6.30E-08 | 6.40E-14 | 0.81 |
| rs8097783 | -0.0259 | -0.0398 | 0.0125 | 0.0060 | -0.0092 | 0.0036 | 0.03764 | 4.20E-11 | 0.19 |
| rs7227255 | -0.1365 | -0.0856 | 0.0224 | 0.0116 | -0.0062 | 0.0065 | 1.12E-09 | 1.70E-13 | 0.94 |
| rs12454712 | 0.0477 | -0.0169 | 0.0068 | 0.0039 | 0.0044 | 0.0019 | 2.40E-12 | 1.46E-05 | 0.23 |
| rs4804833 | 0.0480 | 0.0048 | 0.0066 | 0.0033 | -0.0023 | 0.0019 | 3.42E-13 | 0.1467 | 0.21 |
| rs2242517 | 0.0430 | 0.0034 | 0.0066 | 0.0041 | 0.0060 | 0.0019 | 7.09E-11 | 0.407 | 0.0043 |
| rs17724992 | 0.0148 | 0.0194 | 0.0075 | 0.0035 | 0.0005 | 0.0021 | 0.04962 | 3.42E-08 | 0.67 |
| rs10401969 | -0.0876 | 0.0188 | 0.0120 | 0.0063 | -0.0079 | 0.0035 | 3.46E-13 | 0.002761 | 0.056 |
| rs29941 | -0.0229 | -0.0182 | 0.0068 | 0.0033 | -0.0018 | 0.0020 | 0.000766 | 2.41E-08 | 0.76 |
| rs6857 | -0.0660 | -0.0213 | 0.0086 | 0.0056 | -0.0005 | 0.0025 | 1.50E-14 | 0.00013 | 0.85 |
| rs2075650 | 0.0620 | 0.0258 | 0.0091 | 0.0045 | 0.0012 | 0.0026 | 1.00E-11 | 1.25E-08 | 0.92 |
| rs10406431 | 0.0603 | 0.0079 | 0.0065 | 0.0038 | -0.0037 | 0.0019 | 1.56E-20 | 0.03761 | 0.14 |
| rs11671664 | 0.0612 | -0.0277 | 0.0099 | 0.0053 | -0.0025 | 0.0031 | 7.61E-10 | 1.45E-07 | 0.064 |
| rs2287019 | 0.0219 | -0.0360 | 0.0082 | 0.0042 | -0.0058 | 0.0024 | 0.007327 | 4.59E-18 | 0.0039 |
| rs3810291 | 0.0434 | 0.0283 | 0.0069 | 0.0036 | -0.0002 | 0.0020 | 3.38E-10 | 4.81E-15 | 0.62 |
| rs4813428 | 0.0599 | 0.0274 | 0.0109 | 0.0065 | 0.0054 | 0.0032 | 3.78E-08 | 2.49E-05 | 0.26 |
| rs1007090 | -0.0440 | -0.0105 | 0.0068 | 0.0040 | -0.0104 | 0.0019 | 1.01E-10 | 0.008665 | 3.60E-05 |
| rs6103716 | -0.0394 | 0.0063 | 0.0069 | 0.0041 | 0.0015 | 0.0020 | 1.19E-08 | 0.1244 | 0.69 |
| rs1800961 | 0.1602 | -0.0031 | 0.0175 | 0.0092 | 0.0102 | 0.0053 | 5.10E-20 | 0.7349 | 0.077 |
| rs1999536 | -0.0404 | 0.0056 | 0.0065 | 0.0038 | 0.0210 | 0.0019 | 4.86E-10 | 0.1406 | 6.30E-22 |
| rs6012876 | 0.0428 | -0.0074 | 0.0065 | 0.0040 | -0.0076 | 0.0019 | 4.31E-11 | 0.06431 | 0.0013 |
| rs4812034 | 0.0416 | 0.0023 | 0.0064 | 0.0038 | 0.0033 | 0.0019 | 7.35E-11 | 0.545 | 0.023 |
| rs2023681 | -0.0826 | 0.0017 | 0.0115 | 0.0068 | 0.0196 | 0.0032 | 7.40E-13 | 0.8026 | 1.60E-08 |
| rs5758223 | 0.0401 | 0.0089 | 0.0071 | 0.0041 | -0.0076 | 0.0021 | 1.78E-08 | 0.02995 | 0.011 |
| rs738409 | -0.0436 | 0.0060 | 0.0076 | 0.0046 | 0.0003 | 0.0023 | 1.17E-08 | 0.1921 | 0.57 |
| rs5771069 | -0.0409 | 0.0008 | 0.0065 | 0.0043 | -0.0036 | 0.0019 | 2.97E-10 | 0.8524 | 0.2 |
| rs12484907 | 0.0866 | -0.0021 | 0.0152 | 0.0074 | 0.0143 | 0.0046 | 1.17E-08 | 0.776 | 0.00051 |
